# Gene-environment interaction analysis in atopic eczema: evidence from large population datasets and modelling *in vitro*

**DOI:** 10.1101/2025.01.24.25321071

**Authors:** Marie Standl, Ashley Budu-Aggrey, Luke J Johnston, Martina S Elias, S Hasan Arshad, Peter Bager, Veronique Bataille, Helena Blakeway, Klaus Bonnelykke, Dorret Boomsma, Ben M Brumpton, Mariona Bustamante Pineda, Archie Campbell, John A Curtin, Anders Eliasen, João PS Fadista, Bjarke Feenstra, Trine Gerner, Carolina Medina Gomez, Sarah Grosche, Kristine B. Gutzkow, Anne-Sofie Halling, Caroline Hayward, John Henderson, Esther Herrera-Luis, John W Holloway, Joukejan Hottenga, Jonathan O’B Hourihane, Chen Hu, Kristian Hveem, Amaia Irizar, Benedicte Jacquemin, Leon Jessen, Sara Kress, Ramesh J Kurukulaaratchy, Susanne Lau, Sabrina Llop, Mari Løset, Ingo Marenholtz, Dan Mason, Daniel L McCartney, Mads Melbye, Erik Melén, Camelia Minica, Clare S Murray, Tamar Nijsten, Luba M Pardo, Suzanne Pasmans, Craig E Pennell, Maria R Rinnov, Gillian Santorelli, Tamara Schikowski, Darina Sheehan, Angela Simpson, Cilla Söderhäll, Laurent F Thomas, Jacob P Thyssen, Maties Torrent, Toos van Beijsterveldt, Alessia Visconti, Judith M. Vonk, Carol A Wang, Cheng-Jian Xu, Ali H Ziyab, UK Translational Research Network in Dermatology, BIOMAP consortium, Adnan Custovic, Paola Di Meglio, Liesbeth Duijts, Carsten Flohr, Alan D Irvine, Gerard H Koppelman, Young-Ae Lee, Nick J Reynolds, Catherine Smith, Sinéad M Langan, Lavinia Paternoster, Sara J Brown

## Abstract

**Background:** Environmental factors play a role in the pathogenesis of complex traits including atopic eczema (AE) and a greater understanding of gene-environment interactions (G*E) is needed to define pathomechanisms for disease prevention. We analysed data from 16 European studies to test for interaction between the 24 most significant AE-associated loci identified from genome-wide association studies and 18 early-life environmental factors. We tested for replication using a further 10 studies and *in vitro* modelling to independently assess findings.

**Results:** The discovery analysis showed suggestive evidence for interaction (p<0.05) between 7 environmental factors (antibiotic use, cat ownership, dog ownership, breastfeeding, elder sibling, smoking and washing practices) and at least one established variant for AE, 14 interactions in total (maxN=25,339). In replication analysis (maxN=252,040) dog exposure*rs10214237 (on chromosome 5p13.2 near *IL7R*) was nominally significant (OR_interaction_=0.91 [0.83-0.99] P=0.025), with a risk effect of the T allele observed only in those not exposed to dogs. A similar interaction with rs10214237 was observed for siblings in the discovery analysis (OR_interaction_=0.84[0.75-0.94] P=0.003), but replication analysis was under-powered OR_interaction_=1.09[0.82-1.46]). Rs10214237 homozygous risk genotype is associated with lower IL-7R expression in human keratinocytes, and dog exposure modelled *in vitro* showed a differential response according to rs10214237 genotype.

**Conclusions:** Interaction analysis and functional assessment provide evidence that early-life dog exposure may modify the genetic effect of rs10214237 on AE via *IL7R*, supporting observational epidemiology showing a protective effect for dog ownership. The lack of evidence for other G*E studied here implies that only weak effects are likely to occur.

## Background

Atopic eczema (AE, synonymous with atopic dermatitis or eczema [1]) is a chronic inflammatory skin and systemic condition affecting approximately 20% of children and 10% of adults in high-income countries. Eczema is the dermatosis which contributes the greatest number of disability-adjusted life years worldwide [2] and co-morbid conditions, including asthma and allergies, obesity, cardiovascular disease, anxiety and depression add substantially to the social, academic, occupational, and financial impact [3]. Atopic eczema is a heritable trait [4] but the rapid rise in prevalence in industrialised areas over the past 30 years [3, 5] illustrates the importance of environmental factors in aetiology. A greater understanding of environmental effects in driving pathology could facilitate disease prevention.

The European Academy of Allergy and Clinical Immunology published an umbrella review of systematic reviews in allergy epidemiology and identified a relative lack of research in eczema genetic epidemiology and environmental effects [6]. The investigation of environmental factors using observational epidemiology is inherently challenging in the context of AE because there are multiple confounding factors and possible reverse causation [7]. Genetic studies, however, have made substantial progress in defining mechanisms in eczema predisposition and pathogenesis, including skin barrier dysfunction and aberrant immune response [8]. The evidence of individual variation in susceptibility to environmental allergens and irritants supports the concept of gene-environment interaction (G*E) [9] playing a role in AE and loss-of-function variants in *FLG* encoding the skin barrier protein filaggrin have been implicated [10]. Knowledge of genetic risk may therefore provide an opportunity to identify key environmental effects and clarify important disease biology.

We aimed to investigate evidence of interaction between the most highly significant eczema risk loci defined by genome-wide association studies [11] and environmental risk factors selected based on previous literature [7, 10] and importance to patients and carers [12]. We used early-life environmental exposures (*in utero* and up to the first 12 months of life) to minimise reverse causation and focus on disease pathogenesis. G*E was tested in cohorts and data from European populations, in discovery and replication phases as a pragmatic approach to maximise sample size. Mechanistic assessment was carried out *in vitro* in a skin keratinocyte model to validate the observed interactions.

## Results

Analysis was conducted to assess observational association (of environmental effects) followed by interaction effect (of environmental and genetic risk factors) in the discovery cohorts; next the nominally significant findings and those with *a priori* evidence were tested for replication in available larger cohorts.

### Discovery analysis

In meta-analyses of between 1,084 and 22,263 participants (dependent on exposure, **Additional file 1**) we found strong evidence for antibiotic use increasing risk of AE (*in utero* p=0.004, at 6 months p=0.001 and at 12 months p=6×10^-4^); weaker evidence was found for a protective effect of dog ownership (p=0.03), protective effect of childhood smoke exposure (p=0.038) and risk effect of NO_2_ levels (p=0.035) (M1 models, **Additional file 2**). Little evidence (p>0.05) was found for main effects of caesarean delivery, cat ownership, breastfeeding, elder siblings, *in utero* smoke exposure, washing practices at 6 months and 2 years, PM10 exposure and house dust mite exposure at birth or 1 year (M1 models, **Additional file 2**).

Of the 432 interactions tested (between 24 genetic variants and 18 environmental exposures), we found no significant interactions that passed multiple testing correction, yet 14 nominally significant (p_int_<0.05) interactions (**Figure 1, Table 1**). Of these, 8 interactions indicated a higher genetic risk in the presence of the exposure (OR > 1) and 6 indicated a higher genetic risk in the unexposed stratum (OR < 1). Of the 18 environmental exposures tested, the two with the strongest evidence for interaction with *FLG* null variants were exposure to tobacco smoke between 0 and 2 years (p_int_=0.018) and washing practices during the same period (p_int_=0.045). There was little evidence (p>0.05) for interactions between *FLG* null variants and other tested exposures, though confidence intervals for some interaction estimates were wide (**Additional file 2**). Notably, there was little evidence for interaction between *FLG* null variants and cat exposure (p=0.36), with strong effects of *FLG* in both the unexposed and exposed strata.

**Figure 1.**
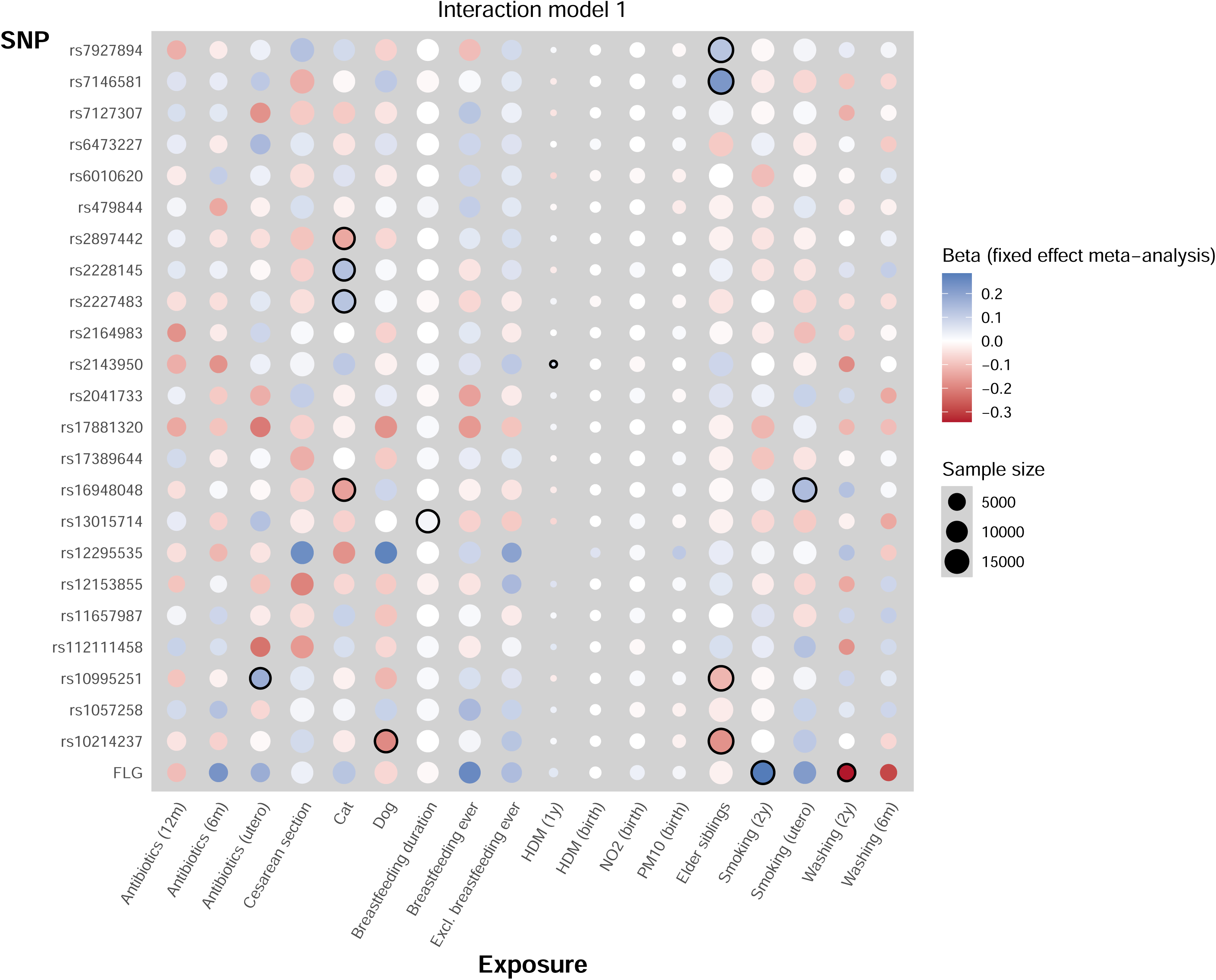
Heatmap to summarise results of interaction analyses. Strength of colour indicates beta in which blue is positive and red is a negative direction of effect; diameter of circle indicates sample size; 14 nominally significant interactions (p_int_<0.05) are highlighted with black outline; one association was reported in only one cohort, so was not pursued further.

**Table 1.**
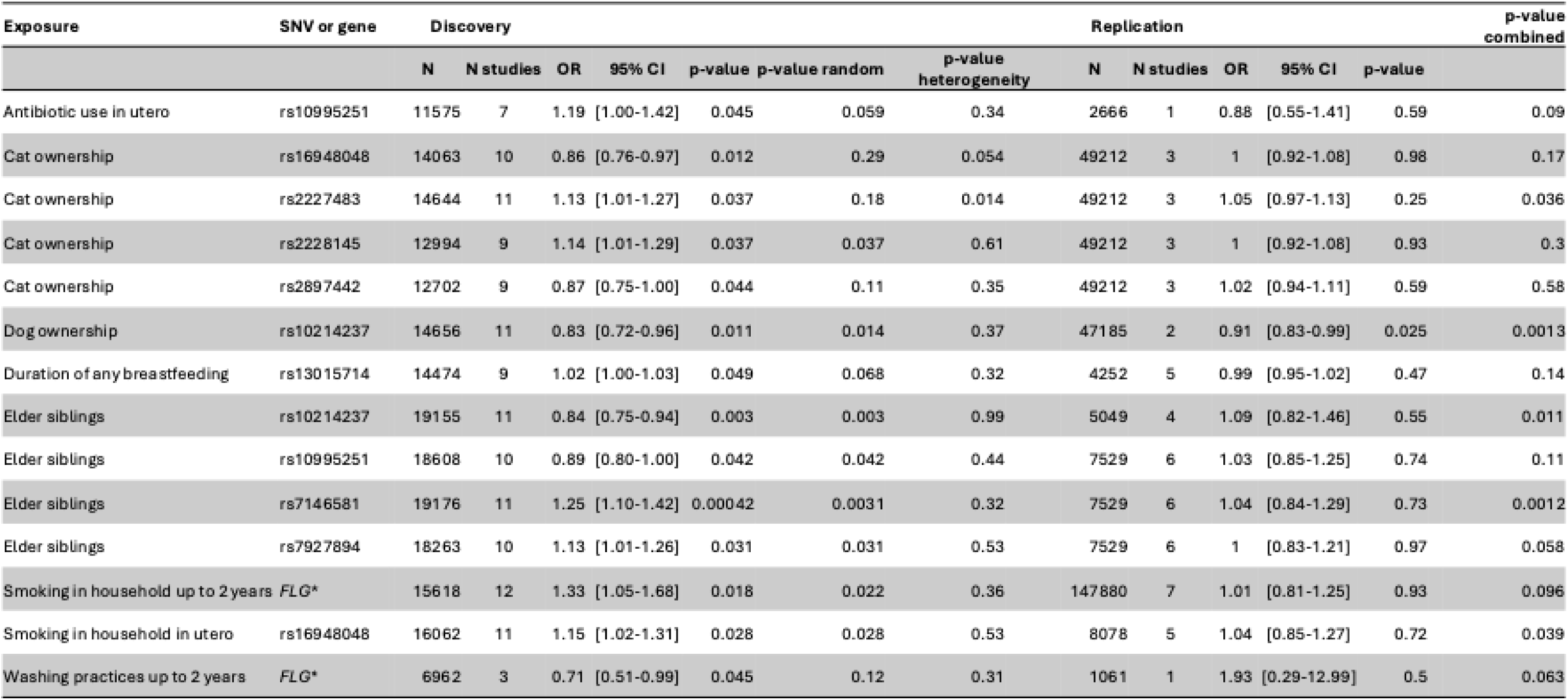
Nominally significant interaction results from discovery and replication analyses. Reults of testing for interaction between 24 genetic variants and 18 environmental exposures; *combined null genotype for 2 or more loss-of-function mutations in *FLG* as detailed in cohort descriptions (Additional file 6); nominal significance defined as unadjusted p_int_<0.05. N, number; OR, odds ratio; p-value indicates significance from fixed effects meta-analysis; p-value random indicates significance from random effects meta-analysis; p-value combined indicates significance from combined fixed effects meta-analysis of discovery and replication data.

Sensitivity analyses, additionally adjusting for family history of AE and socioeconomic status, broadly supported the results of the main analyses (**Additional file 2**), but many of the sensitivity analyses are based on much smaller sample sizes because of the requirement for data on additional covariates. There was little evidence of heterogeneity between cohorts (smallest p_het=_0.01) amongst the 14 reported interactions.

### Replication analysis

We took the 14 interactions with nominal evidence forward to replication, but also the exposures that had prior literature suggesting an interaction with *FLG* null variants (cat, siblings and breast-feeding [10]). In total, 19 interactions based on 8 different exposures and 10 genetic variants were included in the replication.

In replication analysis dog exposure and rs10214237 showed evidence for interaction (p=0.025), **Table 1**. In an analysis stratified by dog exposure the T allele increases risk of atopic eczema (OR=1.14, 95% CI 1.08 to 1.22), but only amongst those who are not exposed to a dog in the family home. In individuals who are exposed to dog in early life, this variant appears to have little effect (OR=0.98, 95% CI 0.68-1.11 **Figure 2**).

**Figure 2.**
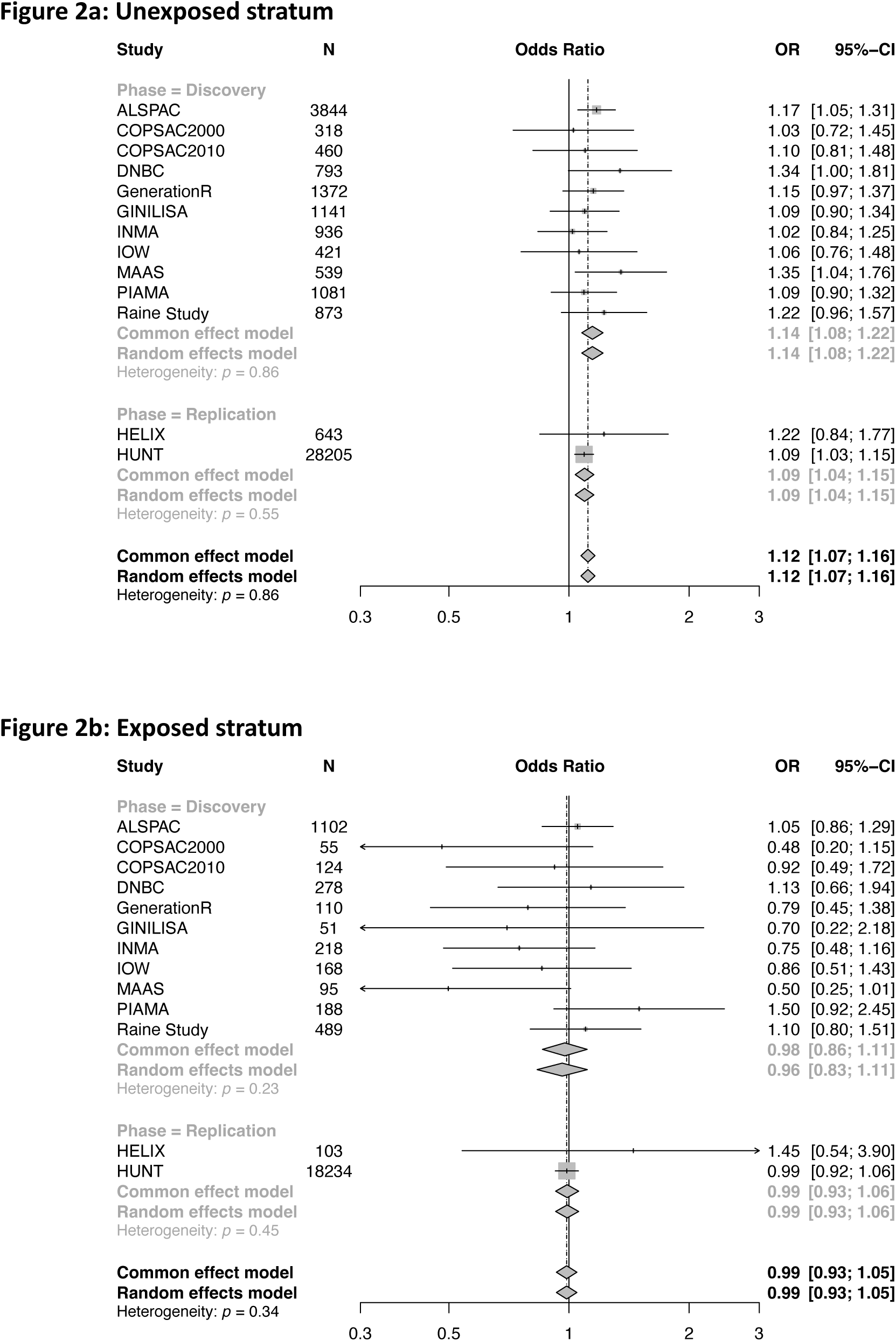
Forest plot showing interaction of dog exposure with rs10214237 in exposed and unexposed strata. Interaction analysis for discovery (N=18,045), replication (N=47,185) and combined meta-analysis (total N= 65,230) show the T allele of rs10214237 increases risk of atopic eczema only amongst those who are not exposed to a dog in the family home. Full names and study cohort descriptions are given in Additional file 6.

Availability of environmental data for replication varied, with many of our attempted replications of interactions being insufficiently powered to be conclusive. Washing practices (0-2y) and antibiotic use *in utero* interactions had only 3 and 4% power respectively (given the interaction effects observed in the discovery phase, **Additional file 3**). The tobacco exposure in utero interaction only reached 11% power and the four sibling interactions had between 8 and 37% power (dependent on variant). The breast-feeding duration interaction only had 4% power in the replication phase and so we extended the replication analysis to ‘ever breastfed’ to increase the power to 56%. The interactions with dog, cat and tobacco smoke exposure 0-2 years were all sufficiently powered (88%, 72-88% and 99%, respectively, **Additional file 3**). The previously reported interactions between *FLG* null mutations and cat, siblings and ever breastfed had 99% power given their reported interaction effects (**Additional file 3**).

### In silico follow-up of rs10214237*dog interaction

Rs10214237 is an intergenic variant (T>C) on chromosome 5p13.2; this was identified in association with eczema by a genome-wide association study (GWAS) [11] in which *IL7R* was prioritised as the likely causal gene based on evidence including eQTL colocalisation in macrophages and monocytes [13, 14]. The top single nucleotide variant (SNV) at this locus in more recent meta-analysis [13] is rs10214273, but this variant is in complete linkage disequilibrium with rs10214237 in European populations (R^2^=1, LDLink version 5.6.6, LDPair tool). Global population data from <u>gnomAD</u> shows ancestral difference in allele frequency, with rs10214237 being more frequent in European and South Asian populations (MAF 0.28 and 0.20 respectively) compared to African people (MAF 0.07) (1KG data accessed 10 Jan 2025).

Rs10214237 is within a region of open chromatin in keratinocytes and fibroblasts, but not the lymphoblastoid cell line GM12878 (UCSC Genome Browser 06 Feb and 27 Nov 2024). Open Targets V2G analyses confirm *IL7R* as most likely gene affected by this SNV based on pQTL, sQTL and eQTL (06 Feb and 27 Nov 2024). GTEx data show that expression of *IL7R* is higher with T:T genotype in whole blood and cultured fibroblasts and in newly generated data we show that individuals with the T:T genotype have slightly higher *IL7R* mRNA expression in primary human keratinocytes than those with the C:C genotype (**Additional file 4**). Single cell data from the Human Protein Atlas [15, 16] confirms that IL-7R is expressed at protein level in human keratinocytes, in addition to circulating immune cells.

### In vitro testing of the effects of dog allergen on human keratinocytes

Human keratinocytes comprise the outermost layer of skin and can therefore represent the first line of interaction in an allergen encounter *in utero* or early life. To further investigate the effect of dog exposure in early life, primary normal human keratinocytes were exposed to clinical-grade dog epithelial extract, a standardised reagent used for allergy testing in the clinic [17]. Dog allergen exposure stimulated an up-regulation in *CXCL8* (IL-8), *CSF2*, *CCL2* and *TNF* mRNA but the atopy-related cytokines *IL33* and *TSLP* mRNA were down-regulated (**Figure 3A**). Network analysis of the proteins encoded by the upregulated transcripts showed significant enrichment for IL-10 signalling (**Figure 3B**, Reactome pathway FDR 7.71e-08) which plays a suppressive role in contact dermatitis and atopic eczema [18]. To test the keratinocyte response more broadly, we used an ELISA panel of 64 cytokine, chemokines and receptors (**Additional file 5**). This confirmed the signature of increased IL-10 signalling (**Additional file 5**).

**Figure 3.**
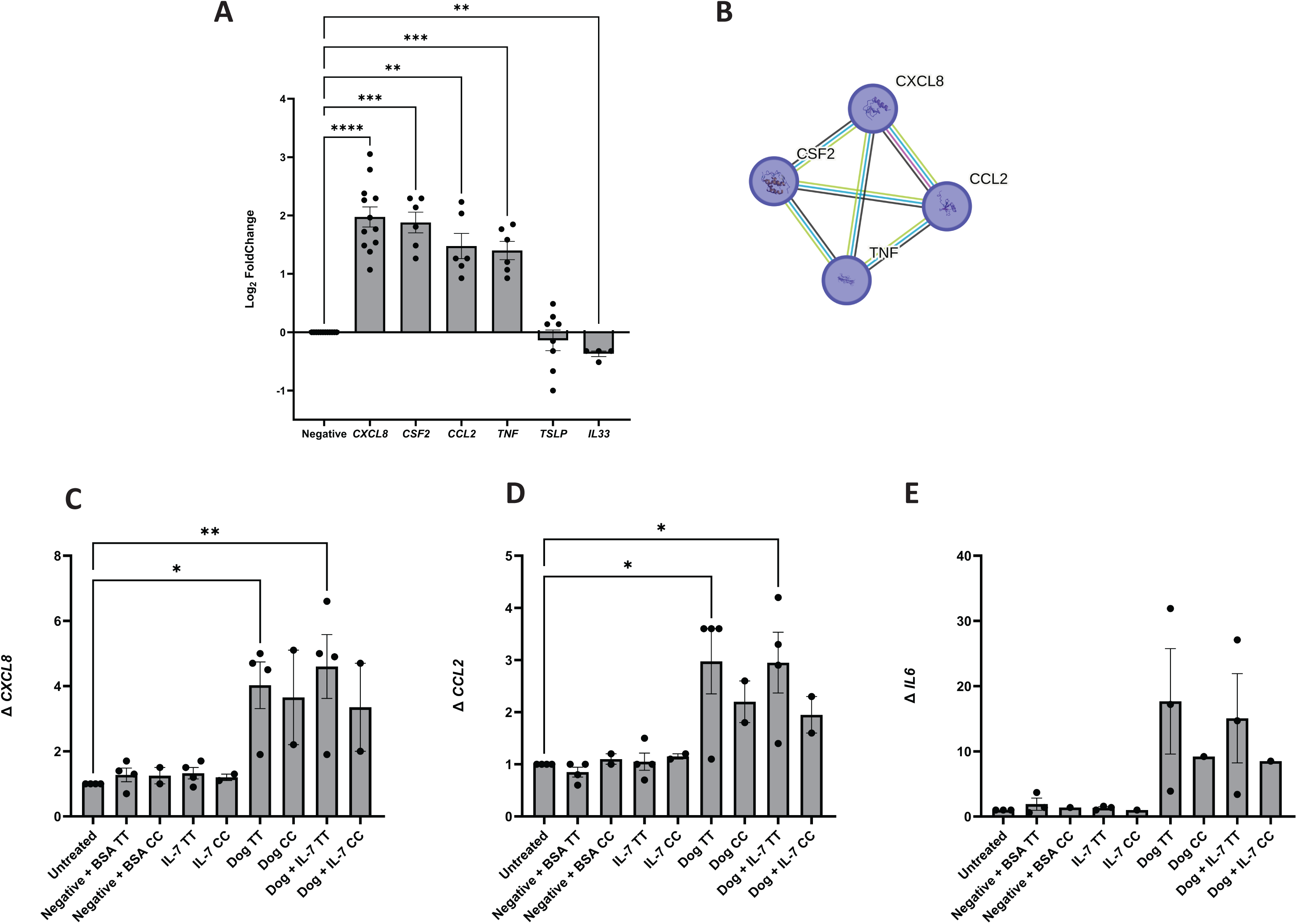
In vitro testing of the effects of dog allergen on primary human keratinocytes. (3A) Dog allergen exposure stimulated a reduction in IL33 and TSLP mRNA but upregulation of CXCL8 (IL-8), CSF2, CCL2 and TNF; negative indicates keratinocyte media with dog allergen carrier solution; 5-12 donor isolates shown, bars represent SEM one-way ANOVA, Dunnet post hoc test compared to negative control, **p<0.01, ***p<0.001 ****p<0.0001. (3B) IL-10 signalling was the most significantly enriched Reactome pathway (4 out of 45 genes/proteins, FDR 7.71e-08). (3C-3E) Effects of IL-7 and dog allergen stimulation on primary human keratinocytes with different rs10214237 genotypes in which T is eczema risk allele; graphs represent the mean fold change in cytokine mRNA expression relative to the housekeeping gene EF1A, from 4 keratinocyte isolates with T:T genotype and 2 keratinocyte isolates from donors of C:C genotype; untreated indicates keratinocyte media only and negative is keratinocyte media with dog allergen carrier solution; BSA as 0.0002% included for as carrier protein for recombinant Il-7; two-way ANOVA with Dunnett’s post-hoc test, compared to the negative control, bars represent SEM, *p<0.05, **p<0.01.

Next, using primary human keratinocytes of known rs10214237 genotype and focusing on CXCL8 (IL-8), CCL2 and IL-6 as molecules of relevance to IL-7R signalling in epithelial cells, we investigated the effect of dog allergen exposure, with and without IL-7 stimulation (**Figure 3C-E**). There was no difference in expression levels after IL-7 stimulation, but on stimulation with dog extract (or IL-7 plus dog extract), keratinocytes of T:T genotype (homozygous for the eczema-risk allele) showed a greater response than the C:C genotype.

Together these observations provide a possible mechanistic explanation for the finding that the T allele at rs10214237 increases risk for atopic eczema; the T:T genotype shows greater IL-7R mRNA expression, but in the context of dog exposure the risk effect is overshadowed by an increase in cytokines and chemokines in the IL-10 pathway which suppresses eczema to a greater extent in T:T than C:C individuals.

## Discussion

Our collaborative work represents the largest and most comprehensive analysis to date investigating G*E in atopic eczema, using a systematic approach focussed on the most significant genetic loci and selected environmental factors. We first meta-analysed data from available observational studies to test for association and then applied interaction analysis to investigate G*E. Statistical power remains a limiting factor and the nominal significance level (p<0.05 without correction for multiple testing) means cautious interpretation is needed. We have identified important negative results as well as one interaction with functional validation *in vitro* and others that warrant further follow up.

A variety of sources provide evidence that G*E plays a role in the aetiology of atopic eczema. These include rapidly rising prevalence [5], clinical observation [4], epidemiological studies [10], and *in vitro* analyses demonstrating molecular effects that include aryl hydrocarbon receptor signalling [19]. Some authors have even stated that ‘atopic eczema is an environmental disease’ [20]. Our meta-analysis of observational associations provides evidence that early-life exposure to antibiotics and NO_2_ levels associate with increased risk of AE, whilst early-life exposure to dog or tobacco smoke is associated with a lower risk of AE in the populations studied. However, these associations may be affected by bias through confounding and reverse causation.

Statistical interaction analysis indicates that early-life dog exposure may modify the genetic effect of rs10214237. Functional genetic analyses show an effect mediated via the gene *IL7R* which encodes the alpha-subunit of the IL-7 receptor. Rs10214237 T:T genotype was associated with an increased risk of atopic eczema in population as a whole and in the sub-population without dog exposure (**Figure 2**) consistent with the T:T genotype showing greater IL7R mRNA expression (**Additional file 4**). The IL-7 receptor is a heterodimer composed of IL7R-alpha and IL2R-gamma. It is expressed is multiple cell-types and tissues, including T-cells, NK-cells, glandular and stratified epithelial cells (data from Human Protein Atlas [15, 16]). IL7R-alpha also contributes to a heteromeric complex with the thymic stromal lymphopoietin (TSLP) receptor but our experimental work to test TSLP as an alternative ligand in keratinocytes was not informative (data not shown) likely, in part, because the TSLPR is only very lowly expressed in this cell type [21].

Our detailed *in vitro* work focussed on human epidermal keratinocytes as the earliest tissue to encounter dog allergen in the initiation of atopic disease, *in utero* or early infancy. We have shown that keratinocytes display a direct response to dog allergen exposure, with down-regulation of IL-33 and TSLP mRNA (both inducers of type 2 immune responses in atopy [22, 23]) and upregulation of a network of genes encoding chemokines and cytokines of IL-10 signalling (Reactome pathway HAS-6783783), contributing to the suppression of atopic inflammation [18]. This is consistent with observational epidemiology showing an apparent protective effect of dog exposure early in life [24] [25]. Gene ontology analysis of the same network indicates a role in cellular response to lipopolysaccharide (GO:0071222), likely to reflect a response to gram negative components of the canine microbiome.

The proposed interaction with genotype was investigated using keratinocytes of known rs10214237 status. Here the T:T genotype showed a greater increase in IL-10 signalling in response to dog allergen exposure than the C:C genotype, which is consistent with the suppression of atopic eczema risk on a population level in the dog-exposed T:T individuals, whilst non-dog-exposed T:T individuals remain at risk of disease. The interaction is analogous to a 17q21*dog interaction demonstrated in asthma [26] in which the risk of persistent wheeze is attenuated by dog ownership [26]. There is an interesting parallel in the interaction of rs10214237 with exposure to older siblings, in which the older sibling abrogates risk effect for rs10214237. We speculate that this may be related to the increased microbial exposure experienced by an infant with older siblings (or a dog) in the household, and there is evidence of shared skin and gut microbiome between humans and their pets[27], but it could also reflect lifestyle choices of dog-owning families and these hypotheses require further testing.

There are some limitations to this work. The discovery analysis used selected SNVs to represent known eczema risk loci, rather than conducting a genome-wide interaction analysis. This restricted approach has been shown to be effective in other traits [28]; it is needed because of power constraints, even in large population datasets. A post-hoc estimation of statistical power (**Additional file 3**) showed that our replication sample sizes were insufficient for some interactions. Therefore, where replication results do not meet our pre-specified significance threshold it is not possible to definitively *exclude* an interaction, but we report the interaction effect sizes for which we had good statistical power, to demonstrate the magnitude of interactions which are unlikely to exist, given our null results (**Additional file 3**). Furthermore, by focusing on selected SNVs within the known AE risk loci, we acknowledge that there may be loci in which an effect is only apparent in the context of interaction with an environmental exposure. These would not be detected by our analysis strategy and genome-wide interaction analysis should be considered in future work if far larger sample sizes than used here become available. An important limitation to this work is the use of European cohort data including people of predominantly white ancestry; this reflects the current sparsity of diverse ancestries in population genetic studies of sufficient size to carry out these analyses. The observed differences in allele frequency of rs10214237 in African compared to European and South Asian populations illustrates the limited transferability of this variant effect across population, although other population-specific variants in the same locus may contribute to similar mechanistic effects. International efforts are on-going to address this limitation [29], and future G*E studies are needed to investigate population-specific environmental effects. More detailed sub-phenotyping of AE may, in the future, reveal that more specific genetic and environmental drivers exist in distinct ancestral or sub-phenotype groups.

In our previous systematic review focusing on gene-environment interactions with *FLG* null mutations [10] we found some published evidence for *FLG*\*environment interactions with exposures including early-life cat ownership, older siblings, water hardness, phthalate exposure, and prolonged breastfeeding from the small number of previous studies. The lack of replication of *FLG*\*cat ownership interaction in the large well-powered study reported here, and another recent meta-analysis [30] represents an interesting null finding, contrasting with two small birth cohort studies ([31, 32] n=379 and n=503) which reported p values for interaction <0.01 with evidence for increased risks of atopic eczema in those with *FLG* null mutations exposed to cat in early life. Evidence for these G*E interactions came from small numbers of individuals with *FLG* mutation, cat exposure and development of atopic eczema (five people in one study [31]). We had very good power (99%) for the interaction magnitude previously reported (OR_int_=11[31]) and 80% for an interaction as small as OR_int_=1.26, suggesting very little evidence in our data for this interaction. We found little evidence for *FLG*\*breastfeeding, consistent with our systematic review [10], where studies reported no evidence for interactions with breastfeeding, although an *FLG*\*breastfeeding duration interaction was reported from the Isle of Wight cohort [33]. Here, our post-hoc power calculation (**Additional file 3**) showed adequate power (99%) for the *FLG*\*breastfed-ever interaction, but low power (<1%) for *FLG*\*breastfeeding duration analyses, which may explain the discrepancy.

## Conclusions

We report observational evidence for an association of atopic eczema with exposure to antibiotics, NO_2_, and tobacco smoke in early life, but the precise nature and mechanisms of action of these environmental factors on atopic skin inflammation remain unclear. We also detected an observational association between early life dog exposure and reduction in prevalence of atopic eczema. Further interaction analysis and functional assessments have provided evidence that dog exposure reduces the genetic risk effect of rs10214237 in a pathway via *IL7R* and possibly IL-10, to suppress skin inflammation. There may be an equivalent interaction effect with siblings, but this is not possible to model *in vitro*. The lack of statistical evidence for other G*E explored in this analysis suggests that only weak interactions are likely to exist, indicating that on a population level the interactions tested and found to be null are unlikely to have important contributions to AE pathogenesis. Therefore further, larger longitudinal studies should focus on alternative mechanistic questions.

## Methods

### Aim

This work aimed to investigate evidence of interaction between 24 genetic risk loci for atopic eczema and 18 early-life environmental effects.

### Study design and setting

Genetic risk loci were defined by the 24 top hits at each locus from genome-wide association analysis [11, 13] and coded for the risk-increasing allele as effect allele (**Additional file 8**). *FLG* null genotype was coded as presence/absence (0/1) of any of the loss-of-function variants prevalent in the white European population (R501X, 2282del4, R2447X, S3247X as previously reported [11, 34]).

Environmental exposures were selected on the basis of our recently published literature review [10], interest from representative of a national eczema support group [12] and refined for pragmatic reasons, based on data availability.

Genetic epidemiology and interaction analysis was used for discovery and replication. *In vitro* modelling was performed to independently assess the one G*E effect that showed a nominally significant interaction in the discovery and replication analyses.

### Characteristics of participants Cohort descriptions

The discovery analysis included 16 population-based cohorts from people of European ancestry (N = 25,339) and a further 10 European population-based cohorts were included in the replication stage (N = 254,532), giving a maximum total of 279,871 (maxN) in the final meta-analysis (**Additional file 1A and 1B**). Disease status was determined by either parental report or doctor diagnosis for those who had “ever had eczema”. Further details on the phenotype definitions for the included studies can be found in **Additional file 6**.

### Keratinocyte culture and gene expression

Primary human keratinocytes were isolated from normal human skin samples excised during routine surgical procedures, with patient consent, under governance of the Lothian Bioresource (reference SR1665). Samples were genotyped for rs10214237 using KASP^TM^ (LGC Genomics, Teddington, England). IL7R mRNA expression was quantified in 34 keratinocyte samples (3 of C:C genotype, 15 T:C and 16 T:T) using RT-qPCR. RNA was isolated with TRIzol (15596026, Invitrogen, Carlsbad, USA) and spin filtration columns using Direct-zol (R2072, Zymo, Irvine, USA). cDNA was prepared using 200ng/ml random primers (48190011, Invitrogen, Carlsbad, USA) with reverse transcriptase using SuperScript IV (18090050, Invitrogen, Carlsbad, USA). qPCR was carried out using exon-spanning probes (*IL7R*: HS00902334_m1, Thermo, Waltham, USA) and (*EF1A*: HS.PT.58.24345862, Integrated DNA Technologies, San Diego, USA) with TaqMan Universal Master Mix II (4440040, Thermo, Waltham, USA) and run on a CFX384 PCR Detection System (Bio-Rad, Hercules, USA) using cycling conditions: 95°C for 10 mins, 40 cycles of [95°C for 15 secs, 60°C for 1 min]. Fold changes in gene expression were derived via the 2(-Delta Delta C[T]) method, using *EF1A* as the reference gene.

### In vitro analysis for rs10214237*dog interaction

To investigate the effect of dog allergen on human keratinocytes, monolayers were treated for 8h with 10ug/ml dog allergen (Can f 1, catalogue E802, Immunotek, Madrid, Spain). RNA isolation and RT-qPCR were carried out as above (*CXCL8*: Hs.PT.58.38869678.g, *CSF2*: Hs.PT.58.20138984, *CCL2*: Hs.PT.58.45467977, *TNF*: Hs.PT.58.45380900, *IL33*: Hs.PT.58.21416460, *EF1A*: HS.PT.58.24345862, Integrated DNA Technologies, San Diego, USA) and (*TSLP*: Hs00263639_m1, Thermo, Waltham, USA). Experiments were replicated using keratinocytes from 5-12 independent donors.

To investigate a genotype-specific effect of IL-7 and/or dog allergen stimulation, keratinocytes were grown to confluency and treated with 100ng/ml recombinant human IL-7 (rhIL-7) (BioTechne, Minneapolis, USA, catalogue: 207-IL) and 500ng/ml Dog dander (Lofarma, Milan, Italy) for 8 hours.

The carrier solution for dog dander (Lofarma) or 0.002% BSA, used as a carrier protein for rhIL-7 were negative controls and experiments were conducted in duplicate for each condition.

One-way analysis of variance (ANOVA) with Dunnett’s post-hoc test for multiple comparisons was used to compare samples’ means and results displayed showing standard error of the mean (SEM). Gene ontology, network and pathway analyses were conducted using STRING v12.0.

### Statistical genetic analysis

The early-life environmental exposures for investigation included pet ownership for cat and dog separately, house dust mite exposure, washing practices (to represent environmental irritants), cigarette tobacco smoking within the household, antibiotic use, environmental pollution, breast feeding mode of delivery and presence of older siblings. These are listed in **Additional file 7**, with details of their definition and coding. The exposures were tested for interaction effects with 24 SNPs previously reported for eczema risk [11, 35] (**Additional file 8**). This involved fitting a statistical model to include the main effect of the SNP upon eczema (G) (extracted from Paternoster et al, 2015[11]), the main effect of the environmental factors upon eczema (E), and the product of the SNP effect and the environmental effect (G*E). Logistic regression models were applied to identify the main effect of each environmental factor (models M1 to M4, **Additional file 9**), and to test for interaction between the exposure and each SNP while adjusting for sex (models I1 to I3, **Additional file 9**). Sensitivity analyses were also performed while adjusting for family history of atopic disease (asthma, eczema or hay fever) and parental education as a proxy for socioeconomic status (models S1 to S3, **Additional file 9**).

Analyses were performed separately within each cohort and then combined by performing fixed-effects meta-analyses. Genetic data was imputed separately for each cohort. Further imputation details can be found in the **Additional file 6**.

### Power calculation

Posthoc estimates of statistical power were calculated in Quanto (version 1.2.4). These were informed by effect size estimates from the discovery analyses or previously published studies, assuming case-to-control ratio of 1:3, and alpha=0.004 in replication analyses (0.05/14 for multiple testing of 14 gene-environment pairs) (**Additional file 3**).

## Data Availability

All data produced in the present study are available upon reasonable request to the authors

## Abbreviations

ANOVA: Analysis of variance
FLG: Gene encoding filaggrin
G*E: Gene-environment interaction
GWAS: Genome-wide association study
maxN: Maximum number of individuals in the analysis
OR: Odds ratio
SEM: Standard error of mean
SNV: Single nucleotide variant
TSLP: Thymic stromal lymphopoietin

## Declarations

### Ethics approval and consent to participate

Each contributing cohort has ethical approval for the sharing of anonymised data from study participants, with their written informed consent.

### Consent for publication

The named authors provide consent for publication of this work.

### Availability of data and materials

All results supporting the conclusions of this article are included within manuscript and Additional files. Specifically, full results for all the models tested are given in **Additional files 2 and 10**. Each cohort contributing to the analysis has their own controlled-access procedures and should be contacted directly to obtain access to individual level data.

### Authors’ contributions

AB-A, PB, KB, DB, SJB, MBP, ACu, CF, JHe, JWH, JO’BH, ADI, GK, PdM, SML, Y-AL, DM, EM, CSM, DMM, SP, LP, CP, NJR, AS, CS, MS, JPT and CW made substantial contributions to the conception or design of the work; all authors contributed to the acquisition, analysis, or interpretation of data; AB-A, SJB, ACu, LD, CF, ADI, GK, SML, Y-AL, LP, NJR, CS and MS drafted the work or substantively revised it.

All authors have approved the submitted version and have agreed both to be personally accountable for the author’s own contributions and to ensure that questions related to the accuracy or integrity of any part of the work, even ones in which the author was not personally involved, are appropriately investigated, resolved, and the resolution documented in the literature.

## Acknowledgements

We are grateful to all patients and participants in the contributing cohort studies and to the skin donors and surgical team at the Western General Hospital, Edinburgh, for providing samples of normal skin tissue for this research; to our UK-TREND collaborators including Mike Cork for informative discussion; to Heather J Cordell for advice on statistical genetics approaches; to Lisa Maier for preparing **Figure 1**; and to Dr Lauren Kelly and Ms Lucy Glass for administrative support.

## Authors’ information

MS, AB-A, LP are experts in statistical genetic analysis of complex traits including AE; SML is a clinical academic dermatologist with epidemiology expertise; SJB is a clinical academic dermatologist with expertise in genetic epidemiology and functional genetics. NJR is a clinical academic dermatologist with expertise in immune mediated inflammatory skin disorders and precision medicine.

## List of additional Files

**Additional file 1. List of cohorts and available exposure data**

**1A.** Included cohorts and exposure availability at discovery stage.

**1B.** Included cohorts and exposure availability at replication stage.

**Additional file 2. Full results of the discovery analysis**

Exposure, environmental exposure; N, number of individuals; N_studies, number of studies; OR_fixed, odds ratio from fixed effect meta-analysis; CI_fixed: 95%-confidence interval from fixed effect meta-analysis; p_fixed: p-value from fixed effect meta-analysis; OR_random: odds ratio from random effects meta-analysis; CI_random: 95%-confidence interval from random effects meta-analysis; p_random: p-value from random effects meta-analysis; p_heterogeneity: p-value from Q-statistic.

**Additional file 3. Estimation of statistical power**

Posthoc power calculations performed to facilitate interpretation of negative findings.

**Additional file 4. IL7R mRNA expression in cells of different rs10214237 genotype**

Rs10214237 T:T genotype is associated with higher expression level of IL7R mRNA than C:C genotype; **4A** and **4B**, Screenshots from GTEx Portal (https://www.gtexportal.org/home/snp/rs10214237 accessed 14/04/2024) showing T:T genotype is associated with higher IL7R mRNA expression in cultured fibroblasts and whole blood; **4C**, shows higher mRNA expression levels in primary human keratinocytes of T:T than C:C genotype.

**Additional file 5. Results of cytokine, chemokine and receptor expression on human primary keratinocytes following dog allergen exposure**

**5A.** Protein detection by ELISA.

**5B.** STRING network analysis of upregulated genes.

**5C.** Cytokine expression from primary human keratinocytes.

**5D.** STRING network analysis of genes showing no significant change in expression.

**Additional file 6. Supplementary methods**

**Additional file 7. Definition and coding of environmental exposures**

**Additional file 8. Table of selected SNVs, risk alleles and risk allele frequencies**

**Additional file 9. Logistic regression models on ever having atopic eczema**

**Additional file 10. Full results of the replication analysis**

Exposure, environmental exposure; N, number of individuals; N_studies, number of studies; OR_fixed, odds ratio from fixed effect meta-analysis; CI_fixed, 95%-confidence interval from fixed effect meta-analysis; p_fixed, p-value from fixed effect meta-analysis; OR_random, odds ratio from random effects meta-analysis; CI_random, 95%-confidence interval from random effects meta-analysis; p_random, p-value from random effects meta-analysis; p_heterogeneity, p-value from Q-statistic.

