## Additional files for "Gene-environment interaction analysis in atopic eczema: evidence from large population datasets and modelling *in vitro*"

### Additional file 1A: Included cohorts and exposure availability at discovery stage

| | Antibiotic use | | | Mode of delivery | Pet ownership | | Breastfeeding | | | Siblings | Smoking in household | | Washing practices | | Air pollution | | House dust mite | | |
|---|---|---|---|---|---|---|---|---|---|---|---|---|---|---|---|---|---|---|---|
| | in utero | 6 months | 12 months | | Cat | Dog | Duration | Ever any | Ever excl. | | in utero | up to 2 years | 6 months | 2 years | NO2 | PM10 | Birth | 1 year | **Max. N** |
| **ALSPAC** | 5,854 | 5,883 | 5,825 | 5,856 | 6,028 | 6,028 | 3,886 | 5,998 | 5,892 | 5,859 | 5,912 | 5,800 | 5,874 | 5,731 | | | | | 6,028 |
| **COPSAC2000** | | 432 | 432 | 432 | 419 | 419 | | | | | 432 | 411 | | | | | | | 432 |
| **COPSAC2010** | 613 | 614 | 614 | 614 | 614 | 614 | | | 613 | 614 | 614 | 609 | | | | | | | 614 |
| **DNBC** | | | | 1,513 | 1,071 | 1,071 | 891 | | 633 | 1,513 | 1,513 | 893 | | | | | | | 1,513 |
| **GenerationR** | 1,539 | 1,488 | 934 | 1,512 | 1,544 | 1,538 | 1,316 | 1,584 | 1,489 | 1,625 | 1,581 | 1,526 | | | 1,536 | 1,536 | | | 1,625 |
| **GINILISA** | 552 | | | 1,432 | 1,438 | 1,448 | 1,463 | 1,465 | 1,462 | 1,470 | 546 | 1,446 | | | 1,447 | 1,447 | 531 | | 1,470 |
| **INMA** | 834 | | 878 | 1,321 | 1,338 | 1,337 | 1,344 | 1,344 | 1,009 | 1,344 | 1,326 | 1,328 | 996 | 996 | 986 | | | | 1,344 |
| **INMA_MNC** | | | | | | | | | | | | | | | | | 333 | | 333 |
| **INMA_SAB** | | 391 | | | | | | | | | | | | | | 382 | | | 382 |
| **IOW** | | | | 794 | 976 | 976 | 910 | 910 | | 847 | 977 | 977 | | | | | | | 977 |
| **MAAS** | | 564 | 564 | 561 | 637 | 637 | 629 | 629 | 629 | 634 | 647 | 623 | | | 519 | 519 | 592 | 309 | 647 |
| **MAS** | | 1,218 | 1,218 | 1,217 | 1,106 | 1,106 | 1,218 | 1,217 | 1,172 | 1,218 | 1,126 | 1,200 | 1,170 | 1,020 | | | 937 | 775 | 1,218 |
| **NTR** | 2,094 | | 2,012 | 1,438 | | | 2,179 | 2,179 | | 2,298 | 2,283 | 2,313 | | | | | | | 2,313 |
| **PIAMA** | | | | 1,261 | 1,268 | 1,269 | 1,262 | 1,262 | | 1,270 | 1,263 | 1,270 | | | 1,265 | | 484 | | 1,270 |
| **Raine Study** | 1,602 | | | 1,600 | 1,602 | 1,602 | 1,330 | 1,523 | 1,523 | | 1,602 | 1,526 | | | | | | | 1,602 |
| **TwinsUK** | | | | 2,609 | | | | | | 3,571 | | | | | | | | | 3,571 |
| **Total** | **13,088** | **10,590** | **12,477** | **22,160** | **18,041** | **18,045** | **16,428** | **18,111** | **14,422** | **22,263** | **19,822** | **19,922** | **8,040** | **7,747** | **5,753** | **3,884** | **2,877** | **1,084** | **25,339** |

### Additional file 1B: Included cohorts and exposure availability at replication stage

| | Breastfeeding | | Siblings | Environmental tobacco smoke | | Pets | | Washing practices up to 2 years | Antibiotics in utero | Max N |
|---|---|---|---|---|---|---|---|---|---|---|
| | Duration | Ever any | | in utero | 0-2y | Cat | Dog | | | |
| **BABYSKIN** | 210 | | 261 | | | | | | | 261 |
| **BAMSE1** | 448 | | 453 | 416 | | | | | | 453 |
| **BAMSE2** | 2,005 | | 2,027 | 1,756 | | 2,027 | | | | 2,027 |
| **BASELINE** | 1,051 | 1,051 | 879 | | 1,061 | 1,061 | | 1,061 | | 1,061 |
| **BiB** | 909 | 920 | 2,575 | 2,670 | 829 | | | | 2,666 | 2,670 |
| **GENSCOT** | | | 1,266 | 2,492 | 2,492 | | | | | 2,492 |
| **GINI_WESEL** | 456 | 459 | 460 | | 453 | | | | | 460 |
| **HELIX** | 434 | | 748 | 744 | 733 | 746 | 746 | | | 748 |
| **HUNT** | | | | | 46,319 | 46,439 | 46,439 | | | 46,439 |
| **UKBIOBANK** | | 197,921 | | | 98,326 | | | | | 197,921 |
| **Total** | **5,513** | **200,351** | **8,669** | **8,078** | **150,213** | **50,273** | **47,185** | **1,061** | **2,666** | **254,532** |

Additional file 2: Full results of the discovery analysis

| exposure | SNP | model | N | N_studies | OR_fixed | CI_fixed | p_fixed | OR_random | CI_random | p_random | p_heterogeneity |
|---|---|---|---|---|---|---|---|---|---|---|---|
| antibio_12m | flg | I1 | 9374 | 7 | 0.90 | [0.68-1.19] | 0.45 | 0.90 | [0.68-1.19] | 0.45 | 0.95 |
| antibio_12m | flg | I2 | 4803 | 7 | 2.09 | [1.72-2.53] | 5.80E-14 | 2.09 | [1.72-2.53] | 5.80E-14 | 0.83 |
| antibio_12m | flg | I3 | 4571 | 7 | 1.88 | [1.55-2.28] | 9.30E-11 | 1.88 | [1.55-2.28] | 9.30E-11 | 0.76 |
| antibio_12m | flg | S1 | 8794 | 7 | 0.84 | [0.63-1.14] | 0.26 | 0.84 | [0.63-1.14] | 0.26 | 0.91 |
| antibio_12m | flg | S2 | 7642 | 7 | 0.92 | [0.66-1.27] | 0.6 | 0.92 | [0.66-1.27] | 0.6 | 0.87 |
| antibio_12m | flg | S3 | 7088 | 7 | 0.87 | [0.62-1.22] | 0.41 | 0.87 | [0.62-1.22] | 0.41 | 0.81 |
| antibio_12m | rs10214237 | I1 | 9996 | 7 | 0.96 | [0.82-1.11] | 0.56 | 0.96 | [0.82-1.11] | 0.56 | 0.71 |
| antibio_12m | rs10214237 | I2 | 5293 | 7 | 1.14 | [1.03-1.27] | 0.01 | 1.14 | [1.03-1.27] | 1.30E-02 | 0.87 |
| antibio_12m | rs10214237 | I3 | 4703 | 7 | 1.11 | [1.00-1.23] | 0.05 | 1.10 | [0.98-1.24] | 0.12 | 0.38 |
| antibio_12m | rs10214237 | S1 | 9368 | 7 | 0.95 | [0.81-1.12] | 0.56 | 0.95 | [0.81-1.12] | 0.56 | 0.48 |
| antibio_12m | rs10214237 | S2 | 6441 | 7 | 1.00 | [0.83-1.19] | 0.96 | 0.94 | [0.72-1.30] | 0.68 | 0.22 |
| antibio_12m | rs10214237 | S3 | 5832 | 7 | 0.98 | [0.82-1.19] | 0.87 | 0.87 | [0.59-1.45] | 0.5 | 0.09 |
| antibio_12m | rs1057258 | I1 | 9978 | 7 | 1.08 | [0.90-1.29] | 0.4 | 1.08 | [0.90-1.29] | 0.4 | 0.84 |
| antibio_12m | rs1057258 | I2 | 5287 | 7 | 0.98 | [0.88-1.11] | 0.79 | 0.98 | [0.88-1.11] | 0.79 | 0.74 |
| antibio_12m | rs1057258 | I3 | 4691 | 7 | 1.08 | [0.95-1.22] | 0.22 | 1.08 | [0.95-1.22] | 0.22 | 0.91 |
| antibio_12m | rs1057258 | S1 | 9368 | 7 | 1.03 | [0.85-1.24] | 0.76 | 1.03 | [0.85-1.24] | 0.76 | 0.63 |
| antibio_12m | rs1057258 | S2 | 6423 | 7 | 1.09 | [0.88-1.33] | 0.44 | 1.09 | [0.88-1.33] | 0.44 | 0.80 |
| antibio_12m | rs1057258 | S3 | 5832 | 7 | 1.06 | [0.85-1.31] | 0.62 | 1.06 | [0.85-1.31] | 0.62 | 0.81 |
| antibio_12m | rs10995251 | I1 | 7980 | 6 | 0.91 | [0.79-1.05] | 0.21 | 0.91 | [0.79-1.05] | 0.21 | 0.64 |
| antibio_12m | rs10995251 | I2 | 3974 | 6 | 1.04 | [0.95-1.14] | 0.4 | 1.04 | [0.95-1.14] | 0.4 | 0.60 |
| antibio_12m | rs10995251 | I3 | 4006 | 6 | 0.95 | [0.87-1.05] | 0.34 | 0.95 | [0.87-1.05] | 0.34 | 0.72 |
| antibio_12m | rs10995251 | S1 | 7356 | 6 | 0.93 | [0.80-1.08] | 0.35 | 0.93 | [0.80-1.08] | 0.35 | 0.50 |
| antibio_12m | rs10995251 | S2 | 6437 | 6 | 0.98 | [0.83-1.15] | 0.8 | 0.98 | [0.83-1.15] | 0.8 | 0.68 |
| antibio_12m | rs10995251 | S3 | 5832 | 6 | 1.02 | [0.86-1.21] | 0.84 | 1.02 | [0.86-1.21] | 0.84 | 0.49 |
| antibio_12m | rs112111458 | I1 | 9015 | 5 | 1.10 | [0.89-1.34] | 0.38 | 1.09 | [0.87-1.38] | 0.45 | 0.36 |
| antibio_12m | rs112111458 | I2 | 4820 | 5 | 0.96 | [0.84-1.10] | 0.58 | 0.96 | [0.84-1.10] | 0.58 | 0.62 |
| antibio_12m | rs112111458 | I3 | 4195 | 5 | 1.09 | [0.95-1.26] | 0.2 | 1.18 | [0.86-1.50] | 0.31 | 0.03 |
| antibio_12m | rs112111458 | S1 | 8403 | 5 | 1.02 | [0.82-1.26] | 0.88 | 1.02 | [0.82-1.26] | 0.88 | 0.82 |
| antibio_12m | rs112111458 | S2 | 6425 | 5 | 1.06 | [0.85-1.32] | 0.61 | 1.06 | [0.82-1.37] | 0.65 | 0.33 |
| antibio_12m | rs112111458 | S3 | 5832 | 5 | 0.99 | [0.79-1.25] | 0.94 | 0.99 | [0.79-1.25] | 0.94 | 0.67 |
| antibio_12m | rs11657987 | I1 | 9967 | 7 | 1.02 | [0.89-1.17] | 0.78 | 1.02 | [0.89-1.17] | 0.78 | 0.64 |
| antibio_12m | rs11657987 | I2 | 5285 | 7 | 1.02 | [0.93-1.11] | 0.75 | 1.02 | [0.93-1.11] | 0.75 | 0.99 |
| antibio_12m | rs11657987 | I3 | 4682 | 7 | 1.05 | [0.96-1.15] | 0.26 | 1.02 | [0.91-1.18] | 0.71 | 0.32 |
| antibio_12m | rs11657987 | S1 | 9368 | 7 | 1.02 | [0.88-1.17] | 0.8 | 1.02 | [0.88-1.17] | 0.8 | 0.41 |
| antibio_12m | rs11657987 | S2 | 6412 | 7 | 1.07 | [0.92-1.25] | 0.39 | 1.07 | [0.92-1.25] | 0.39 | 0.79 |
| antibio_12m | rs11657987 | S3 | 5832 | 7 | 1.07 | [0.91-1.26] | 0.43 | 1.07 | [0.91-1.26] | 0.43 | 0.71 |
| antibio_12m | rs12153855 | I1 | 7020 | 4 | 0.91 | [0.71-1.17] | 0.48 | 0.96 | [0.67-1.32] | 0.84 | 0.21 |
| antibio_12m | rs12153855 | I2 | 3509 | 4 | 1.14 | [0.96-1.36] | 0.13 | 1.13 | [0.84-1.53] | 0.43 | 0.09 |
| antibio_12m | rs12153855 | I3 | 3511 | 4 | 1.13 | [0.96-1.33] | 0.15 | 1.13 | [0.96-1.33] | 0.15 | 0.48 |
| antibio_12m | rs12153855 | S1 | 6391 | 4 | 0.99 | [0.76-1.28] | 0.92 | 1.08 | [0.73-1.46] | 0.7 | 0.22 |
| antibio_12m | rs12153855 | S2 | 6442 | 4 | 0.94 | [0.72-1.22] | 0.64 | 0.98 | [0.67-1.38] | 0.9 | 0.20 |
| antibio_12m | rs12153855 | S3 | 5832 | 4 | 1.02 | [0.77-1.34] | 0.9 | 1.09 | [0.75-1.49] | 0.65 | 0.25 |
| antibio_12m | rs12295535 | I1 | 9997 | 7 | 0.95 | [0.62-1.44] | 0.8 | 0.95 | [0.62-1.45] | 0.81 | 0.42 |
| antibio_12m | rs12295535 | I2 | 5293 | 7 | 1.15 | [0.87-1.53] | 0.32 | 1.15 | [0.87-1.53] | 0.32 | 0.81 |
| antibio_12m | rs12295535 | I3 | 4704 | 7 | 1.07 | [0.81-1.42] | 0.64 | 1.07 | [0.77-1.49] | 0.69 | 0.38 |
| antibio_12m | rs12295535 | S1 | 9368 | 7 | 0.90 | [0.58-1.40] | 0.63 | 0.96 | [0.53-1.63] | 0.88 | 0.34 |
| antibio_12m | rs12295535 | S2 | 6442 | 7 | 0.84 | [0.52-1.36] | 0.47 | 0.84 | [0.52-1.36] | 0.47 | 0.89 |
| antibio_12m | rs12295535 | S3 | 5832 | 7 | 0.78 | [0.47-1.30] | 0.34 | 0.78 | [0.47-1.30] | 0.34 | 0.94 |
| antibio_12m | rs13015714 | I1 | 9997 | 7 | 1.04 | [0.89-1.22] | 0.63 | 1.04 | [0.83-1.31] | 0.74 | 0.21 |
| antibio_12m | rs13015714 | I2 | 5293 | 7 | 1.09 | [0.98-1.22] | 0.1 | 1.09 | [0.98-1.22] | 0.1 | 0.75 |
| antibio_12m | rs13015714 | I3 | 4704 | 7 | 1.15 | [1.03-1.28] | 1.40E-02 | 1.17 | [1.00-1.34] | 5.60E-02 | 0.23 |
| antibio_12m | rs13015714 | S1 | 9368 | 7 | 1.02 | [0.87-1.21] | 0.79 | 1.01 | [0.80-1.29] | 0.95 | 0.26 |
| antibio_12m | rs13015714 | S2 | 6442 | 7 | 1.13 | [0.94-1.36] | 0.18 | 1.16 | [0.90-1.47] | 0.26 | 0.23 |
| antibio_12m | rs13015714 | S3 | 5832 | 7 | 1.09 | [0.90-1.33] | 0.36 | 1.09 | [0.87-1.37] | 0.44 | 0.32 |
| antibio_12m | rs16948048 | I1 | 7983 | 6 | 0.95 | [0.82-1.09] | 0.46 | 0.95 | [0.82-1.09] | 0.46 | 0.88 |
| antibio_12m | rs16948048 | I2 | 3974 | 6 | 1.05 | [0.96-1.16] | 0.28 | 1.05 | [0.96-1.16] | 0.28 | 0.92 |
| antibio_12m | rs16948048 | I3 | 4009 | 6 | 1.00 | [0.91-1.10] | 0.95 | 1.00 | [0.91-1.10] | 0.95 | 0.94 |
| antibio_12m | rs16948048 | S1 | 7356 | 6 | 0.94 | [0.81-1.08] | 0.37 | 0.94 | [0.81-1.08] | 0.37 | 0.90 |
| antibio_12m | rs16948048 | S2 | 6440 | 6 | 0.96 | [0.82-1.13] | 0.62 | 0.96 | [0.82-1.13] | 0.62 | 0.88 |
| antibio_12m | rs16948048 | S3 | 5832 | 6 | 0.95 | [0.80-1.13] | 0.58 | 0.95 | [0.80-1.13] | 0.58 | 0.75 |
| antibio_12m | rs17389644 | I1 | 9985 | 7 | 1.08 | [0.92-1.27] | 0.37 | 1.10 | [0.91-1.30] | 0.31 | 0.36 |
| antibio_12m | rs17389644 | I2 | 5287 | 7 | 1.04 | [0.93-1.16] | 0.48 | 0.98 | [0.83-1.24] | 0.82 | 0.14 |
| antibio_12m | rs17389644 | I3 | 4698 | 7 | 1.13 | [1.01-1.26] | 0.03 | 1.13 | [1.01-1.26] | 3.10E-02 | 0.56 |
| antibio_12m | rs17389644 | S1 | 9368 | 7 | 1.06 | [0.89-1.26] | 0.49 | 1.13 | [0.88-1.37] | 0.34 | 0.22 |
| antibio_12m | rs17389644 | S2 | 6430 | 7 | 1.07 | [0.88-1.29] | 0.50 | 1.07 | [0.88-1.29] | 0.5 | 0.67 |
| antibio_12m | rs17389644 | S3 | 5832 | 7 | 1.04 | [0.85-1.27] | 0.70 | 1.04 | [0.85-1.27] | 0.7 | 0.71 |
| antibio_12m | rs17881320 | I1 | 9979 | 7 | 0.87 | [0.68-1.11] | 0.26 | 0.87 | [0.68-1.11] | 0.26 | 1.00 |
| antibio_12m | rs17881320 | I2 | 5285 | 7 | 1.18 | [1.00-1.39] | 0.05 | 1.18 | [1.00-1.39] | 5.20E-02 | 0.57 |
| antibio_12m | rs17881320 | I3 | 4694 | 7 | 1.10 | [0.93-1.29] | 0.27 | 1.10 | [0.93-1.29] | 0.27 | 0.43 |
| antibio_12m | rs17881320 | S1 | 9368 | 7 | 0.90 | [0.70-1.16] | 0.42 | 0.90 | [0.70-1.16] | 0.42 | 0.99 |
| antibio_12m | rs17881320 | S2 | 6424 | 7 | 0.85 | [0.64-1.13] | 0.27 | 0.85 | [0.64-1.13] | 0.27 | 1.00 |
| antibio_12m | rs17881320 | S3 | 5832 | 7 | 0.88 | [0.65-1.19] | 0.39 | 0.88 | [0.65-1.19] | 0.39 | 0.93 |
| antibio_12m | rs2041733 | I1 | 7985 | 6 | 1.03 | [0.89-1.18] | 0.72 | 1.00 | [0.84-1.22] | 0.98 | 0.29 |
| antibio_12m | rs2041733 | I2 | 3976 | 6 | 1.03 | [0.94-1.13] | 0.5 | 1.03 | [0.92-1.16] | 0.57 | 0.29 |
| antibio_12m | rs2041733 | I3 | 4009 | 6 | 1.04 | [0.95-1.14] | 0.41 | 1.02 | [0.89-1.19] | 0.82 | 0.18 |
| antibio_12m | rs2041733 | S1 | 7356 | 6 | 1.04 | [0.90-1.20] | 0.62 | 1.00 | [0.80-1.29] | 0.99 | 0.21 |
| antibio_12m | rs2041733 | S2 | 6442 | 6 | 1.02 | [0.87-1.19] | 0.79 | 0.97 | [0.75-1.33] | 0.84 | 0.14 |
| antibio_12m | rs2041733 | S3 | 5832 | 6 | 1.04 | [0.88-1.23] | 0.64 | 0.96 | [0.67-1.49] | 0.84 | 0.07 |
| antibio_12m | rs2143950 | I1 | 9993 | 7 | 0.88 | [0.74-1.05] | 0.17 | 0.93 | [0.72-1.14] | 0.56 | 0.21 |
| antibio_12m | rs2143950 | I2 | 5290 | 7 | 1.18 | [1.04-1.33] | 8.20E-03 | 1.15 | [0.99-1.37] | 6.70E-02 | 0.30 |
| antibio_12m | rs2143950 | I3 | 4703 | 7 | 1.00 | [0.89-1.13] | 0.94 | 1.00 | [0.89-1.13] | 0.94 | 0.44 |
| antibio_12m | rs2143950 | S1 | 9368 | 7 | 0.91 | [0.76-1.10] | 0.34 | 0.91 | [0.76-1.10] | 0.34 | 0.50 |
| antibio_12m | rs2143950 | S2 | 6438 | 7 | 0.82 | [0.67-1.01] | 6.10E-02 | 0.82 | [0.62-1.09] | 0.16 | 0.26 |
| antibio_12m | rs2143950 | S3 | 5832 | 7 | 0.87 | [0.70-1.08] | 0.2 | 0.87 | [0.70-1.08] | 0.2 | 0.68 |
| antibio_12m | rs2164983 | I1 | 9032 | 5 | 0.84 | [0.69-1.02] | 8.30E-02 | 0.84 | [0.69-1.02] | 8.30E-02 | 0.49 |
| antibio_12m | rs2164983 | I2 | 4826 | 5 | 1.23 | [1.08-1.41] | 1.80E-03 | 1.22 | [1.02-1.47] | 2.80E-02 | 0.27 |
| antibio_12m | rs2164983 | I3 | 4206 | 5 | 1.08 | [0.95-1.23] | 0.26 | 1.08 | [0.95-1.23] | 0.26 | 0.61 |
| antibio_12m | rs2164983 | S1 | 8403 | 5 | 0.88 | [0.71-1.07] | 0.2 | 0.88 | [0.71-1.07] | 0.2 | 0.56 |
| antibio_12m | rs2164983 | S2 | 6442 | 5 | 0.82 | [0.67-1.01] | 6.80E-02 | 0.82 | [0.67-1.01] | 6.80E-02 | 0.58 |
| antibio_12m | rs2164983 | S3 | 5832 | 5 | 0.86 | [0.69-1.07] | 0.18 | 0.86 | [0.69-1.07] | 0.18 | 0.73 |

| | | | | | | | | | | | |
|---|---|---|---|---|---|---|---|---|---|---|---|
| antibio_12m | rs2227483 | I1 | 9993 | 7 | 0.95 | [0.83-1.09] | 0.45 | 0.94 | [0.81-1.11] | 0.44 | 0.38 |
| antibio_12m | rs2227483 | I2 | 5291 | 7 | 1.09 | [0.99-1.20] | 6.70E-02 | 1.09 | [0.96-1.24] | 0.18 | 0.20 |
| antibio_12m | rs2227483 | I3 | 4702 | 7 | 1.06 | [0.97-1.16] | 0.2 | 1.05 | [0.94-1.19] | 0.38 | 0.33 |
| antibio_12m | rs2227483 | S1 | 9368 | 7 | 0.98 | [0.85-1.13] | 0.78 | 0.98 | [0.85-1.13] | 0.78 | 0.61 |
| antibio_12m | rs2227483 | S2 | 6438 | 7 | 0.95 | [0.81-1.11] | 0.52 | 0.93 | [0.75-1.17] | 0.48 | 0.26 |
| antibio_12m | rs2227483 | S3 | 5832 | 7 | 1.00 | [0.84-1.18] | 0.97 | 1.00 | [0.84-1.18] | 0.97 | 0.44 |
| antibio_12m | rs2228145 | I1 | 7985 | 6 | 1.05 | [0.91-1.21] | 0.5 | 1.05 | [0.91-1.21] | 0.5 | 0.58 |
| antibio_12m | rs2228145 | I2 | 3976 | 6 | 1.00 | [0.91-1.10] | 0.98 | 1.00 | [0.91-1.10] | 0.98 | 0.72 |
| antibio_12m | rs2228145 | I3 | 4009 | 6 | 1.04 | [0.95-1.14] | 0.42 | 1.04 | [0.95-1.14] | 0.42 | 0.64 |
| antibio_12m | rs2228145 | S1 | 7356 | 6 | 1.07 | [0.92-1.24] | 0.36 | 1.07 | [0.92-1.24] | 0.36 | 0.63 |
| antibio_12m | rs2228145 | S2 | 6442 | 6 | 1.05 | [0.89-1.23] | 0.58 | 1.03 | [0.83-1.29] | 0.8 | 0.28 |
| antibio_12m | rs2228145 | S3 | 5832 | 6 | 1.07 | [0.91-1.27] | 0.4 | 1.07 | [0.88-1.31] | 0.48 | 0.33 |
| antibio_12m | rs2897442 | I1 | 7984 | 6 | 1.03 | [0.88-1.20] | 0.75 | 0.94 | [0.71-1.36] | 0.68 | 0.06 |
| antibio_12m | rs2897442 | I2 | 3975 | 6 | 1.13 | [1.02-1.26] | 2.10E-02 | 1.16 | [0.98-1.34] | 7.50E-02 | 0.11 |
| antibio_12m | rs2897442 | I3 | 4009 | 6 | 1.16 | [1.05-1.29] | 4.20E-03 | 1.13 | [0.94-1.40] | 0.2 | 0.08 |
| antibio_12m | rs2897442 | S1 | 7356 | 6 | 1.04 | [0.89-1.23] | 0.6 | 0.93 | [0.67-1.46] | 0.69 | 0.04 |
| antibio_12m | rs2897442 | S2 | 6441 | 6 | 1.00 | [0.83-1.19] | 0.99 | 0.92 | [0.67-1.37] | 0.59 | 0.12 |
| antibio_12m | rs2897442 | S3 | 5832 | 6 | 1.01 | [0.84-1.22] | 0.93 | 0.87 | [0.57-1.54] | 0.51 | 0.06 |
| antibio_12m | rs479844 | I1 | 9997 | 7 | 1.02 | [0.89-1.17] | 0.77 | 0.87 | [0.67-1.33] | 0.32 | 0.04 |
| antibio_12m | rs479844 | I2 | 5293 | 7 | 1.14 | [1.03-1.25] | 7.40E-03 | 1.19 | [1.03-1.32] | 2.10E-02 | 0.10 |
| antibio_12m | rs479844 | I3 | 4704 | 7 | 1.18 | [1.08-1.30] | 3.80E-04 | 1.18 | [1.08-1.30] | 3.80E-04 | 0.64 |
| antibio_12m | rs479844 | S1 | 9368 | 7 | 1.06 | [0.92-1.22] | 0.44 | 0.97 | [0.77-1.33] | 0.8 | 0.17 |
| antibio_12m | rs479844 | S2 | 6442 | 7 | 1.05 | [0.90-1.24] | 0.54 | 0.90 | [0.63-1.51] | 0.59 | 0.02 |
| antibio_12m | rs479844 | S3 | 5832 | 7 | 1.12 | [0.94-1.32] | 0.2 | 1.12 | [0.94-1.32] | 0.2 | 0.46 |
| antibio_12m | rs6010620 | I1 | 9997 | 7 | 0.97 | [0.83-1.15] | 0.76 | 0.97 | [0.83-1.15] | 0.76 | 0.57 |
| antibio_12m | rs6010620 | I2 | 5293 | 7 | 1.13 | [1.02-1.27] | 2.60E-02 | 1.13 | [1.02-1.27] | 2.60E-02 | 0.60 |
| antibio_12m | rs6010620 | I3 | 4704 | 7 | 1.09 | [0.98-1.21] | 0.13 | 1.09 | [0.98-1.21] | 0.13 | 0.70 |
| antibio_12m | rs6010620 | S1 | 9368 | 7 | 0.96 | [0.81-1.13] | 0.61 | 0.96 | [0.81-1.13] | 0.61 | 0.51 |
| antibio_12m | rs6010620 | S2 | 6442 | 7 | 1.04 | [0.86-1.25] | 0.71 | 1.04 | [0.86-1.25] | 0.71 | 0.45 |
| antibio_12m | rs6010620 | S3 | 5832 | 7 | 1.00 | [0.83-1.22] | 0.97 | 1.00 | [0.83-1.22] | 0.97 | 0.58 |
| antibio_12m | rs6473227 | I1 | 9997 | 7 | 1.04 | [0.90-1.20] | 0.58 | 1.04 | [0.90-1.20] | 0.58 | 0.45 |
| antibio_12m | rs6473227 | I2 | 5293 | 7 | 1.05 | [0.95-1.15] | 0.32 | 1.05 | [0.95-1.15] | 0.32 | 0.79 |
| antibio_12m | rs6473227 | I3 | 4704 | 7 | 1.11 | [1.02-1.22] | 2.30E-02 | 1.11 | [1.02-1.22] | 2.30E-02 | 0.78 |
| antibio_12m | rs6473227 | S1 | 9368 | 7 | 1.05 | [0.91-1.22] | 0.5 | 1.05 | [0.91-1.22] | 0.5 | 0.47 |
| antibio_12m | rs6473227 | S2 | 6442 | 7 | 1.07 | [0.91-1.25] | 0.43 | 1.07 | [0.91-1.25] | 0.43 | 0.85 |
| antibio_12m | rs6473227 | S3 | 5832 | 7 | 1.07 | [0.90-1.27] | 0.44 | 1.07 | [0.90-1.27] | 0.44 | 0.75 |
| antibio_12m | rs7127307 | I1 | 7985 | 6 | 1.07 | [0.93-1.23] | 0.32 | 1.07 | [0.93-1.23] | 0.32 | 0.75 |
| antibio_12m | rs7127307 | I2 | 3976 | 6 | 1.05 | [0.96-1.15] | 0.31 | 1.05 | [0.96-1.15] | 0.31 | 0.70 |
| antibio_12m | rs7127307 | I3 | 4009 | 6 | 1.11 | [1.01-1.22] | 2.40E-02 | 1.11 | [1.01-1.22] | 2.40E-02 | 0.47 |
| antibio_12m | rs7127307 | S1 | 7356 | 6 | 1.05 | [0.91-1.22] | 0.47 | 1.05 | [0.91-1.22] | 0.47 | 0.75 |
| antibio_12m | rs7127307 | S2 | 6442 | 6 | 1.05 | [0.90-1.23] | 0.56 | 1.05 | [0.90-1.23] | 0.56 | 0.85 |
| antibio_12m | rs7127307 | S3 | 5832 | 6 | 1.03 | [0.87-1.22] | 0.73 | 1.03 | [0.87-1.22] | 0.73 | 0.96 |
| antibio_12m | rs7146581 | I1 | 9996 | 7 | 1.06 | [0.90-1.25] | 0.47 | 1.06 | [0.90-1.25] | 0.47 | 0.62 |
| antibio_12m | rs7146581 | I2 | 5292 | 7 | 1.01 | [0.90-1.13] | 0.86 | 1.01 | [0.90-1.14] | 0.86 | 0.39 |
| antibio_12m | rs7146581 | I3 | 4704 | 7 | 1.05 | [0.94-1.17] | 0.43 | 1.05 | [0.94-1.17] | 0.43 | 0.69 |
| antibio_12m | rs7146581 | S1 | 9368 | 7 | 1.04 | [0.88-1.23] | 0.67 | 1.04 | [0.88-1.23] | 0.67 | 0.59 |
| antibio_12m | rs7146581 | S2 | 6441 | 7 | 1.06 | [0.88-1.28] | 0.55 | 1.06 | [0.88-1.28] | 0.55 | 0.64 |
| antibio_12m | rs7146581 | S3 | 5832 | 7 | 1.02 | [0.84-1.24] | 0.86 | 1.02 | [0.84-1.24] | 0.86 | 0.66 |
| antibio_12m | rs7927894 | I1 | 9961 | 7 | 0.88 | [0.77-1.01] | 6.50E-02 | 0.88 | [0.77-1.01] | 6.50E-02 | 0.85 |
| antibio_12m | rs7927894 | I2 | 5277 | 7 | 1.08 | [0.98-1.19] | 0.11 | 1.07 | [0.94-1.23] | 0.31 | 0.22 |
| antibio_12m | rs7927894 | I3 | 4684 | 7 | 0.98 | [0.89-1.07] | 0.63 | 0.96 | [0.84-1.11] | 0.5 | 0.25 |
| antibio_12m | rs7927894 | S1 | 9368 | 7 | 0.87 | [0.76-1.01] | 6.20E-02 | 0.87 | [0.76-1.01] | 6.20E-02 | 0.77 |
| antibio_12m | rs7927894 | S2 | 6406 | 7 | 0.83 | [0.71-0.97] | 2.20E-02 | 0.83 | [0.71-0.97] | 2.20E-02 | 0.62 |
| antibio_12m | rs7927894 | S3 | 5832 | 7 | 0.82 | [0.70-0.97] | 2.40E-02 | 0.82 | [0.70-0.97] | 2.40E-02 | 0.52 |
| antibio_12m | | M1 | 11259 | 7 | 1.17 | [1.07-1.28] | 5.80E-04 | 1.17 | [1.07-1.28] | 5.80E-04 | 0.57 |
| antibio_12m | | M2 | 10606 | 7 | 1.16 | [1.05-1.27] | 2.00E-03 | 1.16 | [1.05-1.27] | 2.00E-03 | 0.61 |
| antibio_12m | | M3 | 8684 | 7 | 1.21 | [1.10-1.33] | 1.30E-04 | 1.21 | [1.10-1.33] | 1.30E-04 | 0.62 |
| antibio_12m | | M4 | 8057 | 7 | 1.19 | [1.08-1.32] | 4.70E-04 | 1.19 | [1.08-1.32] | 4.70E-04 | 0.60 |
| antibio_6m | flg | I1 | 9750 | 7 | 1.26 | [0.93-1.70] | 0.13 | 1.26 | [0.93-1.70] | 0.13 | 0.93 |
| antibio_6m | flg | I2 | 7008 | 7 | 1.81 | [1.54-2.12] | 3.20E-13 | 1.81 | [1.54-2.12] | 6.20E-13 | 0.42 |
| antibio_6m | flg | I3 | 2742 | 7 | 2.30 | [1.80-2.94] | 3.10E-11 | 2.30 | [1.80-2.94] | 3.10E-11 | 0.97 |
| antibio_6m | flg | S1 | 9172 | 7 | 1.21 | [0.89-1.67] | 0.23 | 1.21 | [0.89-1.67] | 0.23 | 0.84 |
| antibio_6m | flg | S2 | 8002 | 7 | 1.32 | [0.95-1.84] | 9.80E-02 | 1.32 | [0.95-1.84] | 9.80E-02 | 0.85 |
| antibio_6m | flg | S3 | 7448 | 7 | 1.27 | [0.90-1.80] | 0.18 | 1.27 | [0.90-1.80] | 0.18 | 0.63 |
| antibio_6m | rs10214237 | I1 | 8119 | 6 | 0.93 | [0.80-1.10] | 0.41 | 0.92 | [0.71-1.21] | 0.51 | 0.15 |
| antibio_6m | rs10214237 | I2 | 5729 | 6 | 1.15 | [1.05-1.25] | 2.30E-03 | 1.14 | [1.04-1.26] | 6.20E-03 | 0.37 |
| antibio_6m | rs10214237 | I3 | 2390 | 6 | 1.08 | [0.95-1.24] | 0.23 | 1.08 | [0.90-1.29] | 0.41 | 0.29 |
| antibio_6m | rs10214237 | S1 | 7491 | 6 | 0.92 | [0.78-1.09] | 0.35 | 0.89 | [0.64-1.27] | 0.46 | 0.09 |
| antibio_6m | rs10214237 | S2 | 6564 | 6 | 0.99 | [0.83-1.18] | 0.89 | 0.91 | [0.63-1.43] | 0.63 | 0.04 |
| antibio_6m | rs10214237 | S3 | 5952 | 6 | 0.96 | [0.80-1.16] | 0.68 | 0.83 | [0.51-1.55] | 0.44 | 0.02 |
| antibio_6m | rs1057258 | I1 | 8101 | 6 | 1.14 | [0.95-1.37] | 0.16 | 1.17 | [0.93-1.44] | 0.19 | 0.30 |
| antibio_6m | rs1057258 | I2 | 5718 | 6 | 1.01 | [0.91-1.12] | 0.88 | 1.01 | [0.91-1.12] | 0.88 | 0.67 |
| antibio_6m | rs1057258 | I3 | 2383 | 6 | 1.17 | [1.00-1.36] | 4.70E-02 | 1.17 | [1.00-1.36] | 4.70E-02 | 0.50 |
| antibio_6m | rs1057258 | S1 | 7491 | 6 | 1.10 | [0.90-1.33] | 0.36 | 1.10 | [0.90-1.33] | 0.36 | 0.70 |
| antibio_6m | rs1057258 | S2 | 6546 | 6 | 1.21 | [0.99-1.48] | 0.07 | 1.28 | [0.95-1.62] | 9.80E-02 | 0.19 |
| antibio_6m | rs1057258 | S3 | 5952 | 6 | 1.16 | [0.94-1.44] | 0.17 | 1.16 | [0.94-1.44] | 0.17 | 0.81 |
| antibio_6m | rs10995251 | I1 | 8115 | 6 | 0.98 | [0.84-1.13] | 0.76 | 0.91 | [0.72-1.25] | 0.47 | 0.15 |
| antibio_6m | rs10995251 | I2 | 5724 | 6 | 1.01 | [0.93-1.09] | 0.83 | 1.02 | [0.91-1.13] | 0.71 | 0.16 |
| antibio_6m | rs10995251 | I3 | 2391 | 6 | 0.98 | [0.86-1.11] | 0.73 | 0.98 | [0.86-1.11] | 0.73 | 0.42 |
| antibio_6m | rs10995251 | S1 | 7491 | 6 | 0.99 | [0.85-1.16] | 0.91 | 0.89 | [0.64-1.37] | 0.46 | 0.07 |
| antibio_6m | rs10995251 | S2 | 6560 | 6 | 1.00 | [0.85-1.18] | 0.95 | 0.92 | [0.66-1.39] | 0.6 | 0.05 |
| antibio_6m | rs10995251 | S3 | 5952 | 6 | 1.03 | [0.86-1.22] | 0.76 | 0.91 | [0.59-1.58] | 0.66 | 0.02 |
| antibio_6m | rs112111458 | I1 | 7138 | 4 | 1.07 | [0.87-1.31] | 0.53 | 1.07 | [0.87-1.31] | 0.53 | 0.42 |
| antibio_6m | rs112111458 | I2 | 4870 | 4 | 0.95 | [0.85-1.07] | 0.39 | 0.95 | [0.85-1.07] | 0.39 | 0.52 |
| antibio_6m | rs112111458 | I3 | 2268 | 4 | 1.02 | [0.86-1.21] | 0.83 | 1.17 | [0.82-1.45] | 0.38 | 0.04 |
| antibio_6m | rs112111458 | S1 | 6526 | 4 | 1.02 | [0.82-1.27] | 0.83 | 1.02 | [0.82-1.27] | 0.83 | 0.46 |
| antibio_6m | rs112111458 | S2 | 6548 | 4 | 1.07 | [0.86-1.32] | 0.57 | 1.07 | [0.85-1.33] | 0.56 | 0.38 |
| antibio_6m | rs112111458 | S3 | 5952 | 4 | 1.03 | [0.82-1.29] | 0.82 | 1.03 | [0.82-1.29] | 0.82 | 0.46 |
| antibio_6m | rs11657987 | I1 | 8090 | 6 | 1.09 | [0.94-1.25] | 0.26 | 0.99 | [0.77-1.40] | 0.95 | 0.10 |
| antibio_6m | rs11657987 | I2 | 5708 | 6 | 1.04 | [0.96-1.12] | 0.33 | 1.04 | [0.96-1.12] | 0.33 | 0.63 |
| antibio_6m | rs11657987 | I3 | 2382 | 6 | 1.12 | [0.99-1.26] | 6.20E-02 | 0.99 | [0.76-1.46] | 0.97 | 0.02 |

| | | | | | | | | | | | |
|---|---|---|---|---|---|---|---|---|---|---|---|
| antibio_6m | rs11657987 | S1 | 7491 | 6 | 1.12 | [0.96-1.30] | 0.16 | 1.03 | [0.76-1.51] | 0.87 | 0.09 |
| antibio_6m | rs11657987 | S2 | 6535 | 6 | 1.13 | [0.96-1.32] | 0.14 | 1.13 | [0.96-1.32] | 0.14 | 0.78 |
| antibio_6m | rs11657987 | S3 | 5952 | 6 | 1.15 | [0.97-1.35] | 0.11 | 1.15 | [0.97-1.35] | 0.11 | 0.73 |
| antibio_6m | rs12153855 | I1 | 7155 | 4 | 1.02 | [0.79-1.31] | 0.89 | 1.02 | [0.79-1.31] | 0.89 | 0.58 |
| antibio_6m | rs12153855 | I2 | 4882 | 4 | 1.15 | [1.00-1.33] | 5.50E-02 | 1.15 | [1.00-1.33] | 5.50E-02 | 0.80 |
| antibio_6m | rs12153855 | I3 | 2273 | 4 | 1.17 | [0.95-1.43] | 0.13 | 1.17 | [0.95-1.43] | 0.13 | 0.72 |
| antibio_6m | rs12153855 | S1 | 6526 | 4 | 0.98 | [0.75-1.28] | 0.9 | 0.98 | [0.75-1.28] | 0.9 | 0.62 |
| antibio_6m | rs12153855 | S2 | 6565 | 4 | 1.03 | [0.79-1.33] | 0.85 | 1.03 | [0.79-1.33] | 0.85 | 0.51 |
| antibio_6m | rs12153855 | S3 | 5952 | 4 | 0.97 | [0.74-1.29] | 0.85 | 0.97 | [0.74-1.29] | 0.85 | 0.52 |
| antibio_6m | rs12295535 | I1 | 8120 | 6 | 0.89 | [0.58-1.37] | 0.6 | 0.63 | [0.28-2.00] | 0.27 | 0.14 |
| antibio_6m | rs12295535 | I2 | 5729 | 6 | 1.21 | [0.96-1.54] | 0.11 | 1.21 | [0.96-1.54] | 0.11 | 0.69 |
| antibio_6m | rs12295535 | I3 | 2391 | 6 | 1.07 | [0.76-1.53] | 0.69 | 0.78 | [0.39-2.17] | 0.49 | 0.12 |
| antibio_6m | rs12295535 | S1 | 7491 | 6 | 0.97 | [0.61-1.53] | 0.89 | 0.83 | [0.39-2.06] | 0.62 | 0.25 |
| antibio_6m | rs12295535 | S2 | 6565 | 6 | 0.88 | [0.54-1.43] | 0.61 | 0.58 | [0.20-2.57] | 0.32 | 0.08 |
| antibio_6m | rs12295535 | S3 | 5952 | 6 | 0.94 | [0.57-1.56] | 0.82 | 0.78 | [0.30-2.47] | 0.62 | 0.16 |
| antibio_6m | rs13015714 | I1 | 8120 | 6 | 0.93 | [0.78-1.10] | 0.39 | 0.93 | [0.78-1.10] | 0.39 | 0.56 |
| antibio_6m | rs13015714 | I2 | 5729 | 6 | 1.15 | [1.05-1.26] | 2.30E-03 | 1.15 | [1.05-1.26] | 2.30E-03 | 0.85 |
| antibio_6m | rs13015714 | I3 | 2391 | 6 | 1.06 | [0.92-1.22] | 0.4 | 1.06 | [0.92-1.22] | 0.4 | 0.49 |
| antibio_6m | rs13015714 | S1 | 7491 | 6 | 0.92 | [0.77-1.10] | 0.37 | 0.92 | [0.77-1.10] | 0.37 | 0.42 |
| antibio_6m | rs13015714 | S2 | 6565 | 6 | 0.91 | [0.76-1.09] | 0.31 | 0.91 | [0.76-1.09] | 0.31 | 0.75 |
| antibio_6m | rs13015714 | S3 | 5952 | 6 | 0.90 | [0.74-1.09] | 0.29 | 0.90 | [0.74-1.09] | 0.29 | 0.70 |
| antibio_6m | rs16948048 | I1 | 8118 | 6 | 1.01 | [0.87-1.17] | 0.87 | 1.00 | [0.80-1.26] | 1 | 0.21 |
| antibio_6m | rs16948048 | I2 | 5727 | 6 | 1.04 | [0.96-1.12] | 0.36 | 1.04 | [0.95-1.13] | 0.36 | 0.36 |
| antibio_6m | rs16948048 | I3 | 2391 | 6 | 1.04 | [0.92-1.18] | 0.52 | 1.04 | [0.88-1.23] | 0.68 | 0.27 |
| antibio_6m | rs16948048 | S1 | 7491 | 6 | 0.98 | [0.84-1.14] | 0.8 | 0.95 | [0.77-1.21] | 0.64 | 0.28 |
| antibio_6m | rs16948048 | S2 | 6563 | 6 | 0.93 | [0.79-1.09] | 0.36 | 0.93 | [0.79-1.09] | 0.36 | 0.50 |
| antibio_6m | rs16948048 | S3 | 5952 | 6 | 0.89 | [0.75-1.06] | 0.2 | 0.89 | [0.75-1.06] | 0.2 | 0.89 |
| antibio_6m | rs17389644 | I1 | 8108 | 6 | 0.97 | [0.82-1.16] | 0.75 | 1.03 | [0.82-1.23] | 0.81 | 0.27 |
| antibio_6m | rs17389644 | I2 | 5719 | 6 | 1.13 | [1.03-1.24] | 0.01 | 1.07 | [0.92-1.30] | 0.39 | 0.13 |
| antibio_6m | rs17389644 | I3 | 2389 | 6 | 1.10 | [0.95-1.27] | 0.2 | 1.10 | [0.95-1.27] | 0.2 | 0.79 |
| antibio_6m | rs17389644 | S1 | 7491 | 6 | 1.00 | [0.83-1.19] | 0.96 | 1.09 | [0.81-1.34] | 0.57 | 0.19 |
| antibio_6m | rs17389644 | S2 | 6553 | 6 | 0.93 | [0.77-1.12] | 0.44 | 1.04 | [0.72-1.34] | 0.84 | 0.05 |
| antibio_6m | rs17389644 | S3 | 5952 | 6 | 0.94 | [0.77-1.15] | 0.58 | 1.09 | [0.68-1.51] | 0.71 | 0.03 |
| antibio_6m | rs17881320 | I1 | 8102 | 6 | 0.91 | [0.70-1.19] | 0.49 | 0.91 | [0.70-1.19] | 0.49 | 0.63 |
| antibio_6m | rs17881320 | I2 | 5716 | 6 | 1.12 | [0.97-1.28] | 0.12 | 1.12 | [0.90-1.39] | 0.3 | 0.13 |
| antibio_6m | rs17881320 | I3 | 2386 | 6 | 1.07 | [0.86-1.34] | 0.52 | 1.07 | [0.86-1.34] | 0.52 | 0.67 |
| antibio_6m | rs17881320 | S1 | 7491 | 6 | 0.96 | [0.73-1.27] | 0.78 | 0.96 | [0.73-1.27] | 0.78 | 0.77 |
| antibio_6m | rs17881320 | S2 | 6547 | 6 | 0.98 | [0.73-1.32] | 0.9 | 0.98 | [0.73-1.32] | 0.9 | 0.68 |
| antibio_6m | rs17881320 | S3 | 5952 | 6 | 1.05 | [0.77-1.43] | 0.77 | 1.05 | [0.77-1.43] | 0.77 | 0.75 |
| antibio_6m | rs2041733 | I1 | 8120 | 6 | 0.92 | [0.80-1.06] | 0.26 | 0.92 | [0.74-1.15] | 0.47 | 0.17 |
| antibio_6m | rs2041733 | I2 | 5729 | 6 | 1.06 | [0.98-1.14] | 0.13 | 1.06 | [0.96-1.16] | 0.23 | 0.29 |
| antibio_6m | rs2041733 | I3 | 2391 | 6 | 1.00 | [0.88-1.12] | 0.95 | 1.00 | [0.81-1.23] | 0.97 | 0.10 |
| antibio_6m | rs2041733 | S1 | 7491 | 6 | 0.94 | [0.80-1.09] | 0.39 | 0.96 | [0.73-1.23] | 0.75 | 0.12 |
| antibio_6m | rs2041733 | S2 | 6565 | 6 | 0.94 | [0.80-1.10] | 0.43 | 0.97 | [0.74-1.23] | 0.8 | 0.11 |
| antibio_6m | rs2041733 | S3 | 5952 | 6 | 0.97 | [0.82-1.14] | 0.71 | 1.06 | [0.76-1.35] | 0.74 | 0.08 |
| antibio_6m | rs2143950 | I1 | 8116 | 6 | 0.84 | [0.70-1.02] | 7.50E-02 | 0.84 | [0.70-1.02] | 7.50E-02 | 0.87 |
| antibio_6m | rs2143950 | I2 | 5726 | 6 | 1.14 | [1.03-1.26] | 1.10E-02 | 1.14 | [1.03-1.26] | 1.10E-02 | 0.52 |
| antibio_6m | rs2143950 | I3 | 2390 | 6 | 0.97 | [0.84-1.14] | 0.74 | 0.97 | [0.84-1.14] | 0.74 | 0.77 |
| antibio_6m | rs2143950 | S1 | 7491 | 6 | 0.85 | [0.70-1.03] | 0.1 | 0.85 | [0.70-1.03] | 0.1 | 0.76 |
| antibio_6m | rs2143950 | S2 | 6561 | 6 | 0.83 | [0.68-1.03] | 8.50E-02 | 0.83 | [0.68-1.03] | 8.50E-02 | 0.93 |
| antibio_6m | rs2143950 | S3 | 5952 | 6 | 0.84 | [0.67-1.04] | 0.11 | 0.84 | [0.67-1.04] | 0.11 | 0.73 |
| antibio_6m | rs2164983 | I1 | 7155 | 4 | 0.97 | [0.80-1.19] | 0.8 | 0.91 | [0.69-1.29] | 0.5 | 0.23 |
| antibio_6m | rs2164983 | I2 | 4882 | 4 | 1.17 | [1.05-1.31] | 6.20E-03 | 1.17 | [1.05-1.31] | 6.20E-03 | 0.46 |
| antibio_6m | rs2164983 | I3 | 2273 | 4 | 1.13 | [0.96-1.34] | 0.13 | 0.99 | [0.70-1.60] | 0.94 | 0.04 |
| antibio_6m | rs2164983 | S1 | 6526 | 4 | 1.01 | [0.82-1.25] | 0.91 | 0.90 | [0.60-1.51] | 0.6 | 0.11 |
| antibio_6m | rs2164983 | S2 | 6565 | 4 | 0.94 | [0.76-1.16] | 0.57 | 0.88 | [0.66-1.25] | 0.4 | 0.24 |
| antibio_6m | rs2164983 | S3 | 5952 | 4 | 0.98 | [0.79-1.23] | 0.88 | 0.89 | [0.60-1.46] | 0.55 | 0.13 |
| antibio_6m | rs2227483 | I1 | 8116 | 6 | 0.95 | [0.83-1.10] | 0.53 | 0.85 | [0.64-1.26] | 0.24 | 0.06 |
| antibio_6m | rs2227483 | I2 | 5726 | 6 | 1.07 | [0.99-1.15] | 0.11 | 1.08 | [0.95-1.21] | 0.24 | 0.11 |
| antibio_6m | rs2227483 | I3 | 2390 | 6 | 1.04 | [0.92-1.17] | 0.56 | 0.95 | [0.74-1.33] | 0.66 | 0.03 |
| antibio_6m | rs2227483 | S1 | 7491 | 6 | 0.96 | [0.83-1.12] | 0.63 | 0.87 | [0.65-1.30] | 0.36 | 0.08 |
| antibio_6m | rs2227483 | S2 | 6561 | 6 | 0.99 | [0.84-1.16] | 0.9 | 0.90 | [0.68-1.30] | 0.45 | 0.09 |
| antibio_6m | rs2227483 | S3 | 5952 | 6 | 1.02 | [0.86-1.21] | 0.81 | 0.96 | [0.72-1.35] | 0.77 | 0.14 |
| antibio_6m | rs2228145 | I1 | 8120 | 6 | 1.03 | [0.89-1.19] | 0.67 | 1.03 | [0.89-1.19] | 0.67 | 0.81 |
| antibio_6m | rs2228145 | I2 | 5729 | 6 | 1.01 | [0.93-1.09] | 0.82 | 1.01 | [0.93-1.09] | 0.82 | 0.74 |
| antibio_6m | rs2228145 | I3 | 2391 | 6 | 1.05 | [0.93-1.19] | 0.41 | 1.05 | [0.93-1.19] | 0.41 | 0.83 |
| antibio_6m | rs2228145 | S1 | 7491 | 6 | 1.06 | [0.91-1.23] | 0.47 | 1.06 | [0.91-1.23] | 0.47 | 0.75 |
| antibio_6m | rs2228145 | S2 | 6565 | 6 | 1.04 | [0.89-1.22] | 0.6 | 1.04 | [0.89-1.22] | 0.6 | 0.58 |
| antibio_6m | rs2228145 | S3 | 5952 | 6 | 1.07 | [0.91-1.27] | 0.41 | 1.07 | [0.91-1.27] | 0.41 | 0.47 |
| antibio_6m | rs2897442 | I1 | 8119 | 6 | 0.96 | [0.82-1.13] | 0.62 | 0.96 | [0.82-1.13] | 0.62 | 0.45 |
| antibio_6m | rs2897442 | I2 | 5728 | 6 | 1.17 | [1.07-1.28] | 3.50E-04 | 1.20 | [1.04-1.34] | 9.80E-03 | 0.12 |
| antibio_6m | rs2897442 | I3 | 2391 | 6 | 1.12 | [0.98-1.28] | 0.11 | 1.12 | [0.98-1.28] | 0.11 | 0.53 |
| antibio_6m | rs2897442 | S1 | 7491 | 6 | 0.94 | [0.80-1.12] | 0.51 | 0.94 | [0.80-1.12] | 0.51 | 0.41 |
| antibio_6m | rs2897442 | S2 | 6564 | 6 | 0.94 | [0.78-1.12] | 0.49 | 0.93 | [0.77-1.15] | 0.5 | 0.35 |
| antibio_6m | rs2897442 | S3 | 5952 | 6 | 0.92 | [0.76-1.11] | 0.37 | 0.90 | [0.69-1.18] | 0.4 | 0.25 |
| antibio_6m | rs479844 | I1 | 8120 | 6 | 0.87 | [0.76-1.01] | 6.90E-02 | 0.92 | [0.74-1.09] | 0.45 | 0.19 |
| antibio_6m | rs479844 | I2 | 5729 | 6 | 1.20 | [1.11-1.30] | 6.40E-06 | 1.20 | [1.11-1.30] | 6.40E-06 | 0.76 |
| antibio_6m | rs479844 | I3 | 2391 | 6 | 1.05 | [0.93-1.18] | 0.41 | 1.09 | [0.90-1.27] | 0.38 | 0.17 |
| antibio_6m | rs479844 | S1 | 7491 | 6 | 0.85 | [0.73-0.99] | 3.80E-02 | 0.93 | [0.69-1.14] | 0.61 | 0.09 |
| antibio_6m | rs479844 | S2 | 6565 | 6 | 0.84 | [0.72-0.98] | 2.90E-02 | 0.84 | [0.72-0.98] | 2.90E-02 | 0.99 |
| antibio_6m | rs479844 | S3 | 5952 | 6 | 0.81 | [0.69-0.96] | 1.60E-02 | 0.81 | [0.69-0.96] | 1.60E-02 | 0.98 |
| antibio_6m | rs6010620 | I1 | 8120 | 6 | 1.11 | [0.94-1.31] | 0.23 | 1.10 | [0.92-1.32] | 0.28 | 0.40 |
| antibio_6m | rs6010620 | I2 | 5729 | 6 | 1.05 | [0.96-1.15] | 0.26 | 1.05 | [0.96-1.15] | 0.26 | 0.86 |
| antibio_6m | rs6010620 | I3 | 2391 | 6 | 1.17 | [1.01-1.34] | 3.10E-02 | 1.16 | [0.99-1.36] | 6.20E-02 | 0.38 |
| antibio_6m | rs6010620 | S1 | 7491 | 6 | 1.12 | [0.94-1.33] | 0.22 | 1.11 | [0.91-1.36] | 0.31 | 0.37 |
| antibio_6m | rs6010620 | S2 | 6565 | 6 | 1.15 | [0.96-1.39] | 0.12 | 1.15 | [0.96-1.39] | 0.12 | 0.90 |
| antibio_6m | rs6010620 | S3 | 5952 | 6 | 1.17 | [0.96-1.42] | 0.12 | 1.17 | [0.96-1.42] | 0.12 | 0.88 |
| antibio_6m | rs6473227 | I1 | 8120 | 6 | 0.97 | [0.84-1.13] | 0.72 | 1.01 | [0.80-1.23] | 0.95 | 0.16 |
| antibio_6m | rs6473227 | I2 | 5729 | 6 | 1.09 | [1.01-1.18] | 3.50E-02 | 1.07 | [0.94-1.24] | 0.29 | 0.12 |
| antibio_6m | rs6473227 | I3 | 2391 | 6 | 1.06 | [0.94-1.20] | 0.34 | 1.06 | [0.94-1.20] | 0.34 | 0.63 |
| antibio_6m | rs6473227 | S1 | 7491 | 6 | 0.95 | [0.82-1.11] | 0.55 | 0.97 | [0.73-1.26] | 0.81 | 0.13 |

| | | | | | | | | | | | |
|---|---|---|---|---|---|---|---|---|---|---|---|
| antibio_6m | rs6473227 | S2 | 6565 | 6 | 1.00 | [0.85-1.17] | 0.97 | 1.02 | [0.84-1.21] | 0.87 | 0.30 |
| antibio_6m | rs6473227 | S3 | 5952 | 6 | 0.97 | [0.82-1.15] | 0.71 | 0.98 | [0.79-1.20] | 0.85 | 0.28 |
| antibio_6m | rs7127307 | I1 | 8120 | 6 | 1.05 | [0.91-1.22] | 0.48 | 1.16 | [0.88-1.38] | 0.29 | 0.06 |
| antibio_6m | rs7127307 | I2 | 5729 | 6 | 1.05 | [0.97-1.13] | 0.24 | 1.04 | [0.95-1.15] | 0.41 | 0.33 |
| antibio_6m | rs7127307 | I3 | 2391 | 6 | 1.09 | [0.96-1.23] | 0.17 | 1.12 | [0.93-1.31] | 0.25 | 0.19 |
| antibio_6m | rs7127307 | S1 | 7491 | 6 | 1.00 | [0.86-1.16] | 1 | 1.03 | [0.82-1.26] | 0.8 | 0.23 |
| antibio_6m | rs7127307 | S2 | 6565 | 6 | 1.03 | [0.88-1.21] | 0.68 | 1.08 | [0.80-1.40] | 0.61 | 0.06 |
| antibio_6m | rs7127307 | S3 | 5952 | 6 | 0.97 | [0.82-1.14] | 0.68 | 0.97 | [0.82-1.14] | 0.68 | 0.67 |
| antibio_6m | rs7146581 | I1 | 8119 | 6 | 1.04 | [0.87-1.24] | 0.67 | 1.04 | [0.87-1.24] | 0.67 | 0.64 |
| antibio_6m | rs7146581 | I2 | 5728 | 6 | 1.00 | [0.91-1.10] | 0.97 | 0.98 | [0.86-1.13] | 0.7 | 0.22 |
| antibio_6m | rs7146581 | I3 | 2391 | 6 | 1.03 | [0.89-1.19] | 0.69 | 1.03 | [0.89-1.19] | 0.69 | 0.48 |
| antibio_6m | rs7146581 | S1 | 7491 | 6 | 1.01 | [0.84-1.21] | 0.95 | 1.01 | [0.84-1.21] | 0.95 | 0.81 |
| antibio_6m | rs7146581 | S2 | 6564 | 6 | 1.00 | [0.82-1.21] | 0.97 | 1.00 | [0.82-1.21] | 0.97 | 0.45 |
| antibio_6m | rs7146581 | S3 | 5952 | 6 | 0.95 | [0.78-1.17] | 0.65 | 0.95 | [0.78-1.17] | 0.65 | 0.74 |
| antibio_6m | rs7927894 | I1 | 8084 | 6 | 0.97 | [0.84-1.12] | 0.69 | 0.97 | [0.84-1.12] | 0.69 | 0.65 |
| antibio_6m | rs7927894 | I2 | 5702 | 6 | 1.02 | [0.94-1.10] | 0.64 | 1.02 | [0.94-1.10] | 0.64 | 0.57 |
| antibio_6m | rs7927894 | I3 | 2382 | 6 | 0.99 | [0.87-1.11] | 0.81 | 0.98 | [0.86-1.12] | 0.76 | 0.40 |
| antibio_6m | rs7927894 | S1 | 7491 | 6 | 1.01 | [0.86-1.18] | 0.91 | 1.01 | [0.86-1.18] | 0.91 | 0.88 |
| antibio_6m | rs7927894 | S2 | 6529 | 6 | 0.94 | [0.80-1.11] | 0.47 | 0.93 | [0.78-1.12] | 0.44 | 0.35 |
| antibio_6m | rs7927894 | S3 | 5952 | 6 | 1.00 | [0.84-1.18] | 0.98 | 1.00 | [0.84-1.18] | 0.98 | 0.90 |
| antibio_6m | | M1 | 10590 | 7 | 1.17 | [1.06-1.28] | 9.60E-04 | 1.17 | [1.05-1.29] | 3.90E-03 | 0.36 |
| antibio_6m | | M2 | 9939 | 7 | 1.16 | [1.06-1.28] | 2.00E-03 | 1.18 | [1.03-1.33] | 1.70E-02 | 0.25 |
| antibio_6m | | M3 | 8795 | 7 | 1.14 | [1.03-1.25] | 1.20E-02 | 1.14 | [1.03-1.25] | 1.20E-02 | 0.93 |
| antibio_6m | | M4 | 8168 | 7 | 1.13 | [1.02-1.25] | 0.02 | 1.13 | [1.02-1.25] | 0.02 | 0.83 |
| antibio_utero | flg | I1 | 10062 | 6 | 1.19 | [0.79-1.78] | 0.41 | 1.19 | [0.79-1.78] | 0.41 | 0.42 |
| antibio_utero | flg | I2 | 8521 | 6 | 1.74 | [1.51-2.02] | 8.80E-14 | 1.70 | [1.40-2.12] | 1.00E-07 | 0.30 |
| antibio_utero | flg | I3 | 1541 | 6 | 1.86 | [1.30-2.66] | 6.60E-04 | 1.78 | [1.17-2.85] | 7.50E-03 | 0.30 |
| antibio_utero | flg | S1 | 10050 | 6 | 1.19 | [0.79-1.80] | 0.41 | 1.16 | [0.74-1.86] | 0.51 | 0.38 |
| antibio_utero | flg | S2 | 8698 | 6 | 1.45 | [0.91-2.31] | 0.12 | 1.45 | [0.91-2.31] | 0.12 | 0.75 |
| antibio_utero | flg | S3 | 8689 | 6 | 1.47 | [0.92-2.36] | 0.11 | 1.47 | [0.92-2.36] | 0.11 | 0.77 |
| antibio_utero | rs10214237 | I1 | 11575 | 7 | 0.99 | [0.82-1.20] | 0.92 | 1.00 | [0.73-1.36] | 0.99 | 0.04 |
| antibio_utero | rs10214237 | I2 | 9986 | 7 | 1.14 | [1.06-1.23] | 4.60E-04 | 1.14 | [1.06-1.23] | 4.60E-04 | 0.51 |
| antibio_utero | rs10214237 | I3 | 1589 | 7 | 1.09 | [0.92-1.30] | 0.31 | 1.08 | [0.83-1.42] | 0.56 | 0.06 |
| antibio_utero | rs10214237 | S1 | 11565 | 7 | 0.99 | [0.82-1.20] | 0.92 | 1.00 | [0.73-1.35] | 1 | 0.05 |
| antibio_utero | rs10214237 | S2 | 8282 | 7 | 1.06 | [0.86-1.32] | 0.57 | 1.07 | [0.74-1.53] | 0.72 | 0.04 |
| antibio_utero | rs10214237 | S3 | 8274 | 7 | 1.06 | [0.85-1.32] | 0.58 | 1.07 | [0.75-1.52] | 0.73 | 0.05 |
| antibio_utero | rs1057258 | I1 | 9481 | 6 | 0.94 | [0.74-1.18] | 0.57 | 0.93 | [0.71-1.22] | 0.58 | 0.28 |
| antibio_utero | rs1057258 | I2 | 7993 | 6 | 1.05 | [0.97-1.15] | 0.25 | 1.05 | [0.97-1.15] | 0.25 | 0.44 |
| antibio_utero | rs1057258 | I3 | 1488 | 6 | 0.96 | [0.78-1.18] | 0.7 | 0.96 | [0.78-1.18] | 0.7 | 0.65 |
| antibio_utero | rs1057258 | S1 | 9471 | 6 | 0.95 | [0.75-1.19] | 0.64 | 0.94 | [0.72-1.23] | 0.65 | 0.29 |
| antibio_utero | rs1057258 | S2 | 8282 | 6 | 0.83 | [0.65-1.07] | 0.16 | 0.83 | [0.65-1.07] | 0.16 | 0.85 |
| antibio_utero | rs1057258 | S3 | 8274 | 6 | 0.84 | [0.65-1.09] | 0.19 | 0.84 | [0.65-1.09] | 0.19 | 0.86 |
| antibio_utero | rs10995251 | I1 | 11575 | 7 | 1.19 | [1.00-1.42] | 4.50E-02 | 1.20 | [0.99-1.44] | 5.90E-02 | 0.34 |
| antibio_utero | rs10995251 | I2 | 9986 | 7 | 1.02 | [0.95-1.09] | 0.61 | 1.03 | [0.94-1.11] | 0.51 | 0.22 |
| antibio_utero | rs10995251 | I3 | 1589 | 7 | 1.25 | [1.07-1.46] | 4.50E-03 | 1.25 | [1.07-1.46] | 4.50E-03 | 0.43 |
| antibio_utero | rs10995251 | S1 | 11565 | 7 | 1.19 | [1.00-1.42] | 4.70E-02 | 1.21 | [0.98-1.47] | 7.60E-02 | 0.27 |
| antibio_utero | rs10995251 | S2 | 8282 | 7 | 1.23 | [1.02-1.50] | 3.40E-02 | 1.23 | [1.02-1.50] | 3.40E-02 | 0.62 |
| antibio_utero | rs10995251 | S3 | 8274 | 7 | 1.23 | [1.01-1.49] | 0.04 | 1.23 | [1.01-1.49] | 0.04 | 0.45 |
| antibio_utero | rs112111458 | I1 | 10992 | 6 | 0.80 | [0.62-1.03] | 7.80E-02 | 0.77 | [0.55-1.12] | 0.13 | 0.17 |
| antibio_utero | rs112111458 | I2 | 9618 | 6 | 1.02 | [0.93-1.12] | 0.73 | 1.10 | [0.91-1.22] | 0.33 | 0.02 |
| antibio_utero | rs112111458 | I3 | 1374 | 6 | 0.85 | [0.67-1.07] | 0.16 | 0.85 | [0.67-1.07] | 0.16 | 0.80 |
| antibio_utero | rs112111458 | S1 | 10982 | 6 | 0.79 | [0.61-1.02] | 6.70E-02 | 0.76 | [0.54-1.10] | 0.11 | 0.20 |
| antibio_utero | rs112111458 | S2 | 8282 | 6 | 0.72 | [0.55-0.94] | 1.40E-02 | 0.71 | [0.51-1.01] | 4.20E-02 | 0.20 |
| antibio_utero | rs112111458 | S3 | 8274 | 6 | 0.71 | [0.55-0.93] | 1.10E-02 | 0.70 | [0.49-1.00] | 0.04 | 0.18 |
| antibio_utero | rs11657987 | I1 | 11575 | 7 | 0.97 | [0.82-1.15] | 0.73 | 0.97 | [0.82-1.15] | 0.73 | 0.88 |
| antibio_utero | rs11657987 | I2 | 9986 | 7 | 1.06 | [0.99-1.13] | 0.1 | 1.06 | [0.99-1.13] | 0.1 | 0.45 |
| antibio_utero | rs11657987 | I3 | 1589 | 7 | 1.02 | [0.87-1.18] | 0.84 | 1.02 | [0.87-1.18] | 0.84 | 0.84 |
| antibio_utero | rs11657987 | S1 | 11565 | 7 | 0.97 | [0.82-1.15] | 0.7 | 0.97 | [0.82-1.15] | 0.7 | 0.87 |
| antibio_utero | rs11657987 | S2 | 8282 | 7 | 1.03 | [0.86-1.25] | 0.73 | 1.03 | [0.86-1.25] | 0.73 | 0.98 |
| antibio_utero | rs11657987 | S3 | 8274 | 7 | 1.02 | [0.85-1.24] | 0.8 | 1.02 | [0.85-1.24] | 0.8 | 0.99 |
| antibio_utero | rs12153855 | I1 | 10992 | 6 | 0.91 | [0.68-1.23] | 0.56 | 0.91 | [0.64-1.30] | 0.6 | 0.28 |
| antibio_utero | rs12153855 | I2 | 9618 | 6 | 1.12 | [1.00-1.26] | 4.20E-02 | 1.12 | [1.00-1.26] | 4.20E-02 | 0.44 |
| antibio_utero | rs12153855 | I3 | 1374 | 6 | 1.00 | [0.77-1.31] | 0.99 | 1.00 | [0.77-1.31] | 0.99 | 0.49 |
| antibio_utero | rs12153855 | S1 | 10982 | 6 | 0.90 | [0.66-1.21] | 0.48 | 0.89 | [0.63-1.27] | 0.53 | 0.31 |
| antibio_utero | rs12153855 | S2 | 8282 | 6 | 0.93 | [0.68-1.27] | 0.64 | 0.93 | [0.68-1.27] | 0.64 | 0.45 |
| antibio_utero | rs12153855 | S3 | 8274 | 6 | 0.93 | [0.67-1.27] | 0.64 | 0.93 | [0.67-1.27] | 0.64 | 0.48 |
| antibio_utero | rs12295535 | I1 | 11575 | 7 | 0.96 | [0.58-1.59] | 0.88 | 0.98 | [0.50-1.89] | 0.95 | 0.18 |
| antibio_utero | rs12295535 | I2 | 9986 | 7 | 1.20 | [0.99-1.46] | 5.90E-02 | 1.20 | [0.99-1.46] | 5.90E-02 | 0.89 |
| antibio_utero | rs12295535 | I3 | 1589 | 7 | 1.20 | [0.76-1.88] | 0.44 | 1.22 | [0.71-2.05] | 0.46 | 0.27 |
| antibio_utero | rs12295535 | S1 | 11565 | 7 | 0.96 | [0.58-1.60] | 0.88 | 0.97 | [0.48-1.94] | 0.94 | 0.18 |
| antibio_utero | rs12295535 | S2 | 8282 | 7 | 0.87 | [0.49-1.52] | 0.61 | 0.85 | [0.42-1.73] | 0.65 | 0.25 |
| antibio_utero | rs12295535 | S3 | 8274 | 7 | 0.84 | [0.48-1.48] | 0.55 | 0.83 | [0.43-1.63] | 0.57 | 0.30 |
| antibio_utero | rs13015714 | I1 | 11573 | 7 | 1.14 | [0.94-1.39] | 0.18 | 1.16 | [0.83-1.61] | 0.38 | 0.03 |
| antibio_utero | rs13015714 | I2 | 9985 | 7 | 1.10 | [1.02-1.19] | 1.70E-02 | 1.08 | [0.97-1.22] | 0.16 | 0.21 |
| antibio_utero | rs13015714 | I3 | 1588 | 7 | 1.24 | [1.04-1.48] | 1.70E-02 | 1.20 | [0.89-1.67] | 0.23 | 0.02 |
| antibio_utero | rs13015714 | S1 | 11563 | 7 | 1.15 | [0.94-1.40] | 0.16 | 1.17 | [0.84-1.61] | 0.35 | 0.03 |
| antibio_utero | rs13015714 | S2 | 8280 | 7 | 1.08 | [0.86-1.34] | 0.5 | 1.07 | [0.81-1.43] | 0.62 | 0.20 |
| antibio_utero | rs13015714 | S3 | 8272 | 7 | 1.08 | [0.86-1.34] | 0.52 | 1.07 | [0.81-1.42] | 0.62 | 0.21 |
| antibio_utero | rs16948048 | I1 | 11575 | 7 | 0.99 | [0.84-1.18] | 0.93 | 0.99 | [0.84-1.18] | 0.93 | 0.60 |
| antibio_utero | rs16948048 | I2 | 9986 | 7 | 1.04 | [0.98-1.12] | 0.21 | 1.04 | [0.98-1.12] | 0.21 | 0.67 |
| antibio_utero | rs16948048 | I3 | 1589 | 7 | 1.03 | [0.89-1.20] | 0.68 | 1.03 | [0.89-1.20] | 0.68 | 0.68 |
| antibio_utero | rs16948048 | S1 | 11565 | 7 | 1.00 | [0.84-1.18] | 0.96 | 1.00 | [0.84-1.18] | 0.96 | 0.55 |
| antibio_utero | rs16948048 | S2 | 8282 | 7 | 0.98 | [0.81-1.19] | 0.84 | 0.98 | [0.81-1.19] | 0.84 | 0.63 |
| antibio_utero | rs16948048 | S3 | 8274 | 7 | 0.98 | [0.81-1.19] | 0.85 | 0.98 | [0.81-1.19] | 0.85 | 0.64 |
| antibio_utero | rs17389644 | I1 | 11575 | 7 | 1.01 | [0.82-1.25] | 0.9 | 1.04 | [0.78-1.34] | 0.78 | 0.15 |
| antibio_utero | rs17389644 | I2 | 9986 | 7 | 1.08 | [1.00-1.17] | 4.80E-02 | 1.01 | [0.88-1.24] | 0.84 | 0.06 |
| antibio_utero | rs17389644 | I3 | 1589 | 7 | 1.09 | [0.91-1.31] | 0.35 | 1.09 | [0.87-1.37] | 0.46 | 0.22 |
| antibio_utero | rs17389644 | S1 | 11565 | 7 | 1.02 | [0.83-1.25] | 0.87 | 1.05 | [0.79-1.35] | 0.76 | 0.16 |
| antibio_utero | rs17389644 | S2 | 8282 | 7 | 0.98 | [0.78-1.24] | 0.86 | 1.01 | [0.71-1.39] | 0.96 | 0.08 |
| antibio_utero | rs17389644 | S3 | 8274 | 7 | 0.98 | [0.77-1.23] | 0.84 | 1.00 | [0.71-1.38] | 0.98 | 0.10 |
| antibio_utero | rs17881320 | I1 | 11575 | 7 | 0.81 | [0.58-1.13] | 0.22 | 0.81 | [0.58-1.13] | 0.22 | 0.44 |

| | | | | | | | | | | | |
|---|---|---|---|---|---|---|---|---|---|---|---|
| antibio_utero | rs17881320 | I2 | 9986 | 7 | 1.14 | [1.01-1.29] | 3.10E-02 | 1.14 | [0.99-1.31] | 6.90E-02 | 0.37 |
| antibio_utero | rs17881320 | I3 | 1589 | 7 | 0.91 | [0.68-1.23] | 0.55 | 0.91 | [0.68-1.23] | 0.55 | 0.51 |
| antibio_utero | rs17881320 | S1 | 11565 | 7 | 0.84 | [0.60-1.18] | 0.31 | 0.84 | [0.60-1.18] | 0.31 | 0.50 |
| antibio_utero | rs17881320 | S2 | 8282 | 7 | 0.92 | [0.63-1.36] | 0.69 | 0.92 | [0.63-1.36] | 0.69 | 0.83 |
| antibio_utero | rs17881320 | S3 | 8274 | 7 | 0.94 | [0.64-1.38] | 0.74 | 0.94 | [0.64-1.38] | 0.74 | 0.80 |
| antibio_utero | rs2041733 | I1 | 11574 | 7 | 0.88 | [0.75-1.05] | 0.15 | 0.88 | [0.74-1.06] | 0.17 | 0.37 |
| antibio_utero | rs2041733 | I2 | 9985 | 7 | 1.04 | [0.98-1.11] | 0.2 | 1.03 | [0.93-1.16] | 0.59 | 0.10 |
| antibio_utero | rs2041733 | I3 | 1589 | 7 | 0.89 | [0.77-1.04] | 0.15 | 0.89 | [0.77-1.04] | 0.15 | 0.91 |
| antibio_utero | rs2041733 | S1 | 11564 | 7 | 0.88 | [0.75-1.05] | 0.15 | 0.88 | [0.74-1.06] | 0.17 | 0.37 |
| antibio_utero | rs2041733 | S2 | 8281 | 7 | 0.78 | [0.64-0.94] | 8.90E-03 | 0.78 | [0.64-0.94] | 8.90E-03 | 0.68 |
| antibio_utero | rs2041733 | S3 | 8273 | 7 | 0.78 | [0.64-0.94] | 8.80E-03 | 0.78 | [0.64-0.94] | 8.80E-03 | 0.68 |
| antibio_utero | rs2143950 | I1 | 11575 | 7 | 1.03 | [0.82-1.29] | 0.78 | 1.03 | [0.82-1.29] | 0.78 | 0.52 |
| antibio_utero | rs2143950 | I2 | 9986 | 7 | 1.10 | [1.01-1.20] | 3.50E-02 | 1.09 | [0.97-1.24] | 0.15 | 0.21 |
| antibio_utero | rs2143950 | I3 | 1589 | 7 | 1.09 | [0.89-1.33] | 0.42 | 1.09 | [0.87-1.37] | 0.47 | 0.31 |
| antibio_utero | rs2143950 | S1 | 11565 | 7 | 1.03 | [0.82-1.30] | 0.78 | 1.03 | [0.80-1.33] | 0.8 | 0.34 |
| antibio_utero | rs2143950 | S2 | 8282 | 7 | 1.09 | [0.85-1.40] | 0.49 | 1.09 | [0.85-1.40] | 0.49 | 0.44 |
| antibio_utero | rs2143950 | S3 | 8274 | 7 | 1.10 | [0.85-1.41] | 0.48 | 1.11 | [0.82-1.48] | 0.5 | 0.26 |
| antibio_utero | rs2164983 | I1 | 10992 | 6 | 1.09 | [0.85-1.39] | 0.5 | 1.08 | [0.81-1.45] | 0.61 | 0.33 |
| antibio_utero | rs2164983 | I2 | 9618 | 6 | 1.14 | [1.04-1.25] | 6.00E-03 | 1.12 | [1.00-1.28] | 5.40E-02 | 0.29 |
| antibio_utero | rs2164983 | I3 | 1374 | 6 | 1.20 | [0.96-1.51] | 0.11 | 1.16 | [0.89-1.58] | 0.28 | 0.28 |
| antibio_utero | rs2164983 | S1 | 10982 | 6 | 1.08 | [0.84-1.38] | 0.57 | 1.06 | [0.80-1.43] | 0.67 | 0.33 |
| antibio_utero | rs2164983 | S2 | 8282 | 6 | 1.04 | [0.80-1.35] | 0.77 | 1.04 | [0.80-1.35] | 0.77 | 0.49 |
| antibio_utero | rs2164983 | S3 | 8274 | 6 | 1.04 | [0.80-1.36] | 0.75 | 1.04 | [0.80-1.36] | 0.75 | 0.53 |
| antibio_utero | rs2227483 | I1 | 11575 | 7 | 1.05 | [0.89-1.24] | 0.58 | 1.05 | [0.89-1.24] | 0.58 | 0.58 |
| antibio_utero | rs2227483 | I2 | 9986 | 7 | 1.04 | [0.97-1.11] | 0.3 | 1.03 | [0.96-1.12] | 0.42 | 0.33 |
| antibio_utero | rs2227483 | I3 | 1589 | 7 | 1.06 | [0.92-1.24] | 0.42 | 1.06 | [0.91-1.24] | 0.46 | 0.39 |
| antibio_utero | rs2227483 | S1 | 11565 | 7 | 1.05 | [0.89-1.25] | 0.55 | 1.05 | [0.89-1.25] | 0.55 | 0.57 |
| antibio_utero | rs2227483 | S2 | 8282 | 7 | 1.05 | [0.87-1.27] | 0.58 | 1.05 | [0.87-1.27] | 0.58 | 0.52 |
| antibio_utero | rs2227483 | S3 | 8274 | 7 | 1.06 | [0.88-1.28] | 0.55 | 1.06 | [0.88-1.28] | 0.55 | 0.54 |
| antibio_utero | rs2228145 | I1 | 11575 | 7 | 0.99 | [0.83-1.18] | 0.92 | 0.99 | [0.83-1.18] | 0.92 | 0.55 |
| antibio_utero | rs2228145 | I2 | 9986 | 7 | 1.03 | [0.96-1.10] | 0.41 | 1.03 | [0.93-1.13] | 0.57 | 0.19 |
| antibio_utero | rs2228145 | I3 | 1589 | 7 | 1.02 | [0.87-1.19] | 0.85 | 1.02 | [0.87-1.19] | 0.85 | 0.75 |
| antibio_utero | rs2228145 | S1 | 11565 | 7 | 0.98 | [0.82-1.17] | 0.84 | 0.98 | [0.82-1.17] | 0.84 | 0.55 |
| antibio_utero | rs2228145 | S2 | 8282 | 7 | 0.93 | [0.76-1.13] | 0.46 | 0.94 | [0.75-1.16] | 0.59 | 0.29 |
| antibio_utero | rs2228145 | S3 | 8274 | 7 | 0.92 | [0.76-1.12] | 0.42 | 0.93 | [0.75-1.15] | 0.52 | 0.32 |
| antibio_utero | rs2897442 | I1 | 10213 | 6 | 0.95 | [0.77-1.18] | 0.67 | 0.95 | [0.77-1.18] | 0.67 | 0.54 |
| antibio_utero | rs2897442 | I2 | 8885 | 6 | 1.11 | [1.02-1.20] | 1.20E-02 | 1.09 | [0.99-1.22] | 9.60E-02 | 0.31 |
| antibio_utero | rs2897442 | I3 | 1328 | 6 | 0.99 | [0.82-1.19] | 0.91 | 0.99 | [0.81-1.21] | 0.91 | 0.36 |
| antibio_utero | rs2897442 | S1 | 10203 | 6 | 0.95 | [0.77-1.17] | 0.61 | 0.95 | [0.77-1.17] | 0.61 | 0.47 |
| antibio_utero | rs2897442 | S2 | 6920 | 6 | 0.99 | [0.77-1.26] | 0.92 | 1.00 | [0.76-1.30] | 0.98 | 0.31 |
| antibio_utero | rs2897442 | S3 | 6912 | 6 | 0.98 | [0.77-1.25] | 0.87 | 1.00 | [0.74-1.32] | 0.98 | 0.25 |
| antibio_utero | rs479844 | I1 | 10213 | 6 | 0.98 | [0.81-1.18] | 0.82 | 0.98 | [0.81-1.18] | 0.82 | 0.86 |
| antibio_utero | rs479844 | I2 | 8885 | 6 | 1.14 | [1.06-1.22] | 3.60E-04 | 1.14 | [1.06-1.22] | 3.60E-04 | 0.48 |
| antibio_utero | rs479844 | I3 | 1328 | 6 | 1.11 | [0.94-1.31] | 0.21 | 1.11 | [0.94-1.31] | 0.21 | 0.94 |
| antibio_utero | rs479844 | S1 | 10203 | 6 | 0.99 | [0.82-1.20] | 0.93 | 0.99 | [0.82-1.20] | 0.93 | 0.89 |
| antibio_utero | rs479844 | S2 | 6920 | 6 | 1.00 | [0.81-1.25] | 0.98 | 1.00 | [0.81-1.25] | 0.98 | 0.72 |
| antibio_utero | rs479844 | S3 | 6912 | 6 | 1.01 | [0.82-1.26] | 0.91 | 1.01 | [0.82-1.26] | 0.91 | 0.73 |
| antibio_utero | rs6010620 | I1 | 11575 | 7 | 1.03 | [0.84-1.26] | 0.77 | 1.03 | [0.84-1.26] | 0.77 | 0.62 |
| antibio_utero | rs6010620 | I2 | 9986 | 7 | 1.07 | [0.99-1.16] | 7.10E-02 | 1.07 | [0.99-1.16] | 7.10E-02 | 0.74 |
| antibio_utero | rs6010620 | I3 | 1589 | 7 | 1.07 | [0.90-1.28] | 0.44 | 1.07 | [0.90-1.28] | 0.44 | 0.81 |
| antibio_utero | rs6010620 | S1 | 11565 | 7 | 1.02 | [0.84-1.25] | 0.82 | 1.02 | [0.84-1.25] | 0.82 | 0.57 |
| antibio_utero | rs6010620 | S2 | 8282 | 7 | 0.94 | [0.75-1.17] | 0.56 | 0.94 | [0.75-1.17] | 0.56 | 0.41 |
| antibio_utero | rs6010620 | S3 | 8274 | 7 | 0.93 | [0.74-1.16] | 0.52 | 0.93 | [0.74-1.16] | 0.52 | 0.41 |
| antibio_utero | rs6473227 | I1 | 11575 | 7 | 1.16 | [0.98-1.38] | 9.30E-02 | 1.16 | [0.98-1.38] | 9.30E-02 | 0.66 |
| antibio_utero | rs6473227 | I2 | 9986 | 7 | 1.05 | [0.98-1.13] | 0.13 | 1.04 | [0.96-1.14] | 0.3 | 0.31 |
| antibio_utero | rs6473227 | I3 | 1589 | 7 | 1.19 | [1.02-1.39] | 2.60E-02 | 1.19 | [1.02-1.39] | 2.60E-02 | 0.81 |
| antibio_utero | rs6473227 | S1 | 11565 | 7 | 1.16 | [0.98-1.38] | 8.80E-02 | 1.16 | [0.98-1.38] | 8.80E-02 | 0.66 |
| antibio_utero | rs6473227 | S2 | 8282 | 7 | 1.17 | [0.97-1.42] | 9.90E-02 | 1.17 | [0.97-1.42] | 9.90E-02 | 0.43 |
| antibio_utero | rs6473227 | S3 | 8274 | 7 | 1.17 | [0.97-1.42] | 0.1 | 1.17 | [0.97-1.42] | 0.1 | 0.45 |
| antibio_utero | rs7127307 | I1 | 11575 | 7 | 0.84 | [0.70-1.00] | 5.30E-02 | 0.84 | [0.70-1.00] | 5.30E-02 | 0.51 |
| antibio_utero | rs7127307 | I2 | 9986 | 7 | 1.07 | [1.00-1.15] | 4.70E-02 | 1.07 | [1.00-1.15] | 4.70E-02 | 0.95 |
| antibio_utero | rs7127307 | I3 | 1589 | 7 | 0.90 | [0.77-1.06] | 0.22 | 0.89 | [0.74-1.08] | 0.2 | 0.33 |
| antibio_utero | rs7127307 | S1 | 11565 | 7 | 0.83 | [0.69-0.99] | 3.80E-02 | 0.83 | [0.69-0.99] | 3.80E-02 | 0.49 |
| antibio_utero | rs7127307 | S2 | 8282 | 7 | 0.81 | [0.66-1.00] | 4.50E-02 | 0.81 | [0.66-1.00] | 4.50E-02 | 0.53 |
| antibio_utero | rs7127307 | S3 | 8274 | 7 | 0.80 | [0.65-0.99] | 3.60E-02 | 0.80 | [0.65-0.99] | 3.60E-02 | 0.55 |
| antibio_utero | rs7146581 | I1 | 9481 | 6 | 1.12 | [0.91-1.37] | 0.27 | 1.12 | [0.91-1.37] | 0.27 | 0.44 |
| antibio_utero | rs7146581 | I2 | 7993 | 6 | 1.00 | [0.92-1.08] | 0.97 | 1.00 | [0.92-1.08] | 0.97 | 0.79 |
| antibio_utero | rs7146581 | I3 | 1488 | 6 | 1.11 | [0.93-1.33] | 0.26 | 1.09 | [0.87-1.39] | 0.47 | 0.22 |
| antibio_utero | rs7146581 | S1 | 9471 | 6 | 1.12 | [0.91-1.37] | 0.27 | 1.12 | [0.91-1.38] | 0.3 | 0.39 |
| antibio_utero | rs7146581 | S2 | 8282 | 6 | 1.08 | [0.87-1.35] | 0.48 | 1.08 | [0.86-1.36] | 0.51 | 0.39 |
| antibio_utero | rs7146581 | S3 | 8274 | 6 | 1.09 | [0.87-1.36] | 0.47 | 1.08 | [0.85-1.38] | 0.54 | 0.36 |
| antibio_utero | rs7927894 | I1 | 11575 | 7 | 1.03 | [0.87-1.23] | 0.71 | 1.02 | [0.79-1.34] | 0.87 | 0.08 |
| antibio_utero | rs7927894 | I2 | 9986 | 7 | 1.02 | [0.95-1.09] | 0.6 | 1.02 | [0.89-1.16] | 0.76 | 0.01 |
| antibio_utero | rs7927894 | I3 | 1589 | 7 | 1.02 | [0.88-1.19] | 0.78 | 1.01 | [0.84-1.23] | 0.89 | 0.28 |
| antibio_utero | rs7927894 | S1 | 11565 | 7 | 1.02 | [0.86-1.22] | 0.79 | 1.01 | [0.77-1.35] | 0.96 | 0.06 |
| antibio_utero | rs7927894 | S2 | 8282 | 7 | 1.05 | [0.86-1.27] | 0.65 | 1.07 | [0.78-1.44] | 0.68 | 0.05 |
| antibio_utero | rs7927894 | S3 | 8274 | 7 | 1.04 | [0.86-1.27] | 0.68 | 1.06 | [0.77-1.45] | 0.71 | 0.04 |
| antibio_utero | | M1 | 13088 | 7 | 1.18 | [1.05-1.31] | 3.70E-03 | 1.18 | [1.05-1.31] | 3.70E-03 | 0.89 |
| antibio_utero | | M2 | 13076 | 7 | 1.18 | [1.05-1.31] | 3.70E-03 | 1.18 | [1.05-1.31] | 3.70E-03 | 0.88 |
| antibio_utero | | M3 | 9594 | 6 | 1.16 | [1.02-1.31] | 0.02 | 1.16 | [1.02-1.31] | 0.02 | 0.65 |
| antibio_utero | | M4 | 9585 | 6 | 1.16 | [1.02-1.31] | 1.90E-02 | 1.16 | [1.02-1.31] | 1.90E-02 | 0.65 |
| caes | flg | I1 | 14782 | 11 | 1.03 | [0.74-1.43] | 0.85 | 1.03 | [0.74-1.43] | 0.85 | 0.94 |
| caes | flg | I2 | 12749 | 11 | 1.84 | [1.63-2.08] | 5.60E-23 | 1.82 | [1.57-2.15] | 1.20E-14 | 0.26 |
| caes | flg | I3 | 2033 | 11 | 1.80 | [1.33-2.42] | 1.30E-04 | 1.80 | [1.33-2.42] | 1.30E-04 | 0.99 |
| caes | flg | S1 | 14205 | 11 | 0.99 | [0.70-1.41] | 0.97 | 0.99 | [0.70-1.41] | 0.97 | 0.87 |
| caes | flg | S2 | 13015 | 11 | 1.05 | [0.73-1.51] | 0.81 | 1.05 | [0.73-1.51] | 0.81 | 0.97 |
| caes | flg | S3 | 12461 | 11 | 1.02 | [0.69-1.51] | 0.91 | 1.02 | [0.69-1.51] | 0.91 | 0.91 |
| caes | rs10214237 | I1 | 17307 | 12 | 1.08 | [0.92-1.27] | 0.36 | 1.08 | [0.92-1.27] | 0.36 | 0.47 |
| caes | rs10214237 | I2 | 15133 | 12 | 1.10 | [1.04-1.17] | 1.10E-03 | 1.10 | [1.04-1.17] | 1.10E-03 | 0.69 |
| caes | rs10214237 | I3 | 2174 | 12 | 1.18 | [1.02-1.37] | 2.90E-02 | 1.18 | [1.02-1.37] | 2.90E-02 | 0.68 |
| caes | rs10214237 | S1 | 12590 | 12 | 1.14 | [0.96-1.37] | 0.13 | 1.15 | [0.95-1.39] | 0.16 | 0.33 |

| | | | | | | | | | | | |
|---|---|---|---|---|---|---|---|---|---|---|---|
| caes | rs10214237 | S2 | 15739 | 12 | 1.14 | [0.95-1.36] | 0.15 | 1.13 | [0.94-1.37] | 0.18 | 0.40 |
| caes | rs10214237 | S3 | 11037 | 12 | 1.23 | [1.01-1.50] | 3.60E-02 | 1.23 | [0.99-1.53] | 5.60E-02 | 0.33 |
| caes | rs1057258 | I1 | 18586 | 13 | 1.02 | [0.84-1.23] | 0.84 | 1.02 | [0.84-1.23] | 0.84 | 0.53 |
| caes | rs1057258 | I2 | 16128 | 13 | 1.03 | [0.96-1.11] | 0.37 | 1.03 | [0.96-1.11] | 0.37 | 0.97 |
| caes | rs1057258 | I3 | 2458 | 13 | 1.06 | [0.89-1.27] | 0.5 | 1.06 | [0.89-1.27] | 0.5 | 0.49 |
| caes | rs1057258 | S1 | 14029 | 13 | 0.96 | [0.78-1.18] | 0.69 | 0.96 | [0.78-1.18] | 0.69 | 0.66 |
| caes | rs1057258 | S2 | 15580 | 13 | 1.07 | [0.87-1.32] | 0.5 | 1.08 | [0.87-1.33] | 0.47 | 0.40 |
| caes | rs1057258 | S3 | 11038 | 13 | 1.00 | [0.80-1.25] | 0.99 | 1.00 | [0.80-1.25] | 0.99 | 0.48 |
| caes | rs10995251 | I1 | 18275 | 12 | 1.05 | [0.90-1.22] | 0.52 | 1.05 | [0.90-1.22] | 0.52 | 0.95 |
| caes | rs10995251 | I2 | 15894 | 12 | 1.03 | [0.97-1.08] | 0.33 | 1.03 | [0.97-1.10] | 0.37 | 0.23 |
| caes | rs10995251 | I3 | 2381 | 12 | 1.10 | [0.95-1.27] | 0.19 | 1.10 | [0.95-1.27] | 0.19 | 0.55 |
| caes | rs10995251 | S1 | 13549 | 12 | 1.03 | [0.87-1.22] | 0.73 | 1.03 | [0.87-1.22] | 0.73 | 0.95 |
| caes | rs10995251 | S2 | 15269 | 12 | 1.02 | [0.87-1.21] | 0.79 | 1.02 | [0.87-1.21] | 0.79 | 0.97 |
| caes | rs10995251 | S3 | 10558 | 12 | 0.99 | [0.82-1.20] | 0.93 | 0.99 | [0.82-1.20] | 0.93 | 0.99 |
| caes | rs112111458 | I1 | 16160 | 10 | 0.85 | [0.67-1.07] | 0.17 | 0.81 | [0.58-1.19] | 0.21 | 0.08 |
| caes | rs112111458 | I2 | 14178 | 10 | 1.04 | [0.96-1.12] | 0.31 | 1.08 | [0.96-1.17] | 0.18 | 0.06 |
| caes | rs112111458 | I3 | 1982 | 10 | 0.92 | [0.74-1.15] | 0.47 | 0.89 | [0.64-1.28] | 0.5 | 0.06 |
| caes | rs112111458 | S1 | 13062 | 10 | 0.93 | [0.73-1.19] | 0.56 | 0.93 | [0.73-1.19] | 0.56 | 0.44 |
| caes | rs112111458 | S2 | 14119 | 10 | 0.79 | [0.62-1.01] | 6.40E-02 | 0.75 | [0.52-1.13] | 0.11 | 0.06 |
| caes | rs112111458 | S3 | 11036 | 10 | 0.88 | [0.68-1.13] | 0.31 | 0.88 | [0.68-1.13] | 0.31 | 0.42 |
| caes | rs11657987 | I1 | 18632 | 13 | 0.95 | [0.82-1.10] | 0.51 | 0.95 | [0.82-1.10] | 0.51 | 0.85 |
| caes | rs11657987 | I2 | 16179 | 13 | 1.05 | [1.00-1.11] | 4.60E-02 | 1.05 | [1.00-1.11] | 5.90E-02 | 0.41 |
| caes | rs11657987 | I3 | 2453 | 13 | 0.99 | [0.87-1.13] | 0.91 | 0.99 | [0.87-1.13] | 0.91 | 0.62 |
| caes | rs11657987 | S1 | 14025 | 13 | 0.94 | [0.81-1.10] | 0.46 | 0.94 | [0.81-1.10] | 0.46 | 0.77 |
| caes | rs11657987 | S2 | 15626 | 13 | 0.98 | [0.84-1.15] | 0.8 | 0.98 | [0.84-1.15] | 0.8 | 0.93 |
| caes | rs11657987 | S3 | 11034 | 13 | 0.97 | [0.82-1.16] | 0.76 | 0.97 | [0.82-1.16] | 0.76 | 0.87 |
| caes | rs12153855 | I1 | 15812 | 9 | 0.82 | [0.63-1.08] | 0.17 | 0.82 | [0.63-1.08] | 0.17 | 0.73 |
| caes | rs12153855 | I2 | 13897 | 9 | 1.13 | [1.03-1.25] | 8.50E-03 | 1.13 | [1.03-1.25] | 8.50E-03 | 0.75 |
| caes | rs12153855 | I3 | 1915 | 9 | 0.90 | [0.70-1.16] | 0.4 | 0.90 | [0.70-1.16] | 0.4 | 0.70 |
| caes | rs12153855 | S1 | 12584 | 9 | 0.86 | [0.64-1.15] | 0.3 | 0.85 | [0.63-1.15] | 0.3 | 0.41 |
| caes | rs12153855 | S2 | 13771 | 9 | 0.88 | [0.66-1.18] | 0.4 | 0.87 | [0.64-1.20] | 0.38 | 0.38 |
| caes | rs12153855 | S3 | 10558 | 9 | 0.91 | [0.67-1.25] | 0.57 | 0.87 | [0.60-1.34] | 0.49 | 0.22 |
| caes | rs12295535 | I1 | 16161 | 12 | 1.28 | [0.83-1.96] | 0.26 | 1.28 | [0.83-1.96] | 0.26 | 1.00 |
| caes | rs12295535 | I2 | 13794 | 12 | 1.12 | [0.96-1.32] | 0.16 | 1.12 | [0.96-1.32] | 0.16 | 0.67 |
| caes | rs12295535 | I3 | 2367 | 12 | 1.34 | [0.91-1.97] | 0.14 | 1.34 | [0.91-1.97] | 0.14 | 1.00 |
| caes | rs12295535 | S1 | 14029 | 12 | 1.20 | [0.74-1.94] | 0.46 | 1.20 | [0.74-1.94] | 0.46 | 1.00 |
| caes | rs12295535 | S2 | 13155 | 12 | 1.31 | [0.82-2.10] | 0.26 | 1.31 | [0.82-2.10] | 0.26 | 0.97 |
| caes | rs12295535 | S3 | 11038 | 12 | 1.21 | [0.73-2.03] | 0.46 | 1.21 | [0.73-2.03] | 0.46 | 0.97 |
| caes | rs13015714 | I1 | 18288 | 12 | 0.97 | [0.81-1.15] | 0.69 | 0.97 | [0.81-1.15] | 0.69 | 0.69 |
| caes | rs13015714 | I2 | 15906 | 12 | 1.10 | [1.04-1.17] | 1.80E-03 | 1.10 | [1.04-1.17] | 1.80E-03 | 0.47 |
| caes | rs13015714 | I3 | 2382 | 12 | 1.11 | [0.95-1.30] | 0.2 | 1.11 | [0.94-1.30] | 0.21 | 0.41 |
| caes | rs13015714 | S1 | 13547 | 12 | 0.93 | [0.77-1.12] | 0.46 | 0.93 | [0.77-1.12] | 0.46 | 0.66 |
| caes | rs13015714 | S2 | 15282 | 12 | 0.96 | [0.80-1.16] | 0.71 | 0.96 | [0.80-1.16] | 0.71 | 0.64 |
| caes | rs13015714 | S3 | 10556 | 12 | 0.92 | [0.75-1.13] | 0.43 | 0.92 | [0.75-1.13] | 0.43 | 0.64 |
| caes | rs16948048 | I1 | 18264 | 12 | 0.94 | [0.81-1.10] | 0.46 | 0.94 | [0.80-1.11] | 0.45 | 0.39 |
| caes | rs16948048 | I2 | 15883 | 12 | 1.03 | [0.98-1.09] | 0.25 | 1.03 | [0.98-1.09] | 0.25 | 0.67 |
| caes | rs16948048 | I3 | 2381 | 12 | 0.98 | [0.85-1.13] | 0.8 | 0.98 | [0.85-1.13] | 0.8 | 0.52 |
| caes | rs16948048 | S1 | 13549 | 12 | 0.90 | [0.76-1.06] | 0.19 | 0.90 | [0.76-1.06] | 0.19 | 0.59 |
| caes | rs16948048 | S2 | 15258 | 12 | 0.96 | [0.81-1.14] | 0.65 | 0.96 | [0.80-1.15] | 0.64 | 0.36 |
| caes | rs16948048 | S3 | 10558 | 12 | 0.92 | [0.76-1.10] | 0.35 | 0.92 | [0.76-1.10] | 0.35 | 0.70 |
| caes | rs17389644 | I1 | 18225 | 12 | 0.88 | [0.73-1.05] | 0.15 | 0.87 | [0.72-1.06] | 0.14 | 0.38 |
| caes | rs17389644 | I2 | 15846 | 12 | 1.07 | [1.00-1.14] | 4.30E-02 | 1.05 | [0.97-1.15] | 0.2 | 0.20 |
| caes | rs17389644 | I3 | 2379 | 12 | 0.93 | [0.79-1.10] | 0.4 | 0.88 | [0.71-1.15] | 0.24 | 0.16 |
| caes | rs17389644 | S1 | 13549 | 12 | 0.91 | [0.75-1.11] | 0.35 | 0.88 | [0.70-1.15] | 0.29 | 0.26 |
| caes | rs17389644 | S2 | 15219 | 12 | 0.91 | [0.75-1.11] | 0.36 | 0.91 | [0.75-1.11] | 0.36 | 0.52 |
| caes | rs17389644 | S3 | 10558 | 12 | 0.98 | [0.79-1.22] | 0.84 | 0.98 | [0.79-1.22] | 0.84 | 0.41 |
| caes | rs17881320 | I1 | 18684 | 13 | 0.93 | [0.72-1.20] | 0.59 | 0.93 | [0.72-1.20] | 0.59 | 0.58 |
| caes | rs17881320 | I2 | 16224 | 13 | 1.12 | [1.02-1.24] | 1.40E-02 | 1.12 | [1.01-1.25] | 3.50E-02 | 0.29 |
| caes | rs17881320 | I3 | 2460 | 13 | 1.06 | [0.84-1.34] | 0.64 | 1.07 | [0.80-1.41] | 0.66 | 0.19 |
| caes | rs17881320 | S1 | 14029 | 13 | 0.81 | [0.61-1.08] | 0.16 | 0.81 | [0.61-1.08] | 0.16 | 0.51 |
| caes | rs17881320 | S2 | 15678 | 13 | 1.01 | [0.75-1.34] | 0.97 | 1.01 | [0.75-1.34] | 0.97 | 0.50 |
| caes | rs17881320 | S3 | 11038 | 13 | 0.87 | [0.63-1.21] | 0.41 | 0.87 | [0.63-1.21] | 0.41 | 0.46 |
| caes | rs2041733 | I1 | 18289 | 12 | 1.11 | [0.96-1.28] | 0.17 | 1.11 | [0.96-1.28] | 0.17 | 0.80 |
| caes | rs2041733 | I2 | 15907 | 12 | 1.06 | [1.00-1.12] | 3.30E-02 | 1.07 | [0.98-1.16] | 0.12 | 0.00 |
| caes | rs2041733 | I3 | 2382 | 12 | 1.15 | [1.00-1.31] | 4.50E-02 | 1.15 | [1.00-1.31] | 4.50E-02 | 0.99 |
| caes | rs2041733 | S1 | 13548 | 12 | 1.19 | [1.02-1.40] | 2.90E-02 | 1.19 | [1.02-1.40] | 2.90E-02 | 0.97 |
| caes | rs2041733 | S2 | 15283 | 12 | 1.05 | [0.89-1.23] | 0.57 | 1.05 | [0.89-1.23] | 0.57 | 0.78 |
| caes | rs2041733 | S3 | 10557 | 12 | 1.14 | [0.96-1.36] | 0.14 | 1.14 | [0.96-1.36] | 0.14 | 0.97 |
| caes | rs2143950 | I1 | 18721 | 13 | 1.02 | [0.84-1.24] | 0.84 | 1.01 | [0.80-1.28] | 0.92 | 0.21 |
| caes | rs2143950 | I2 | 16262 | 13 | 1.07 | [1.00-1.14] | 5.40E-02 | 1.07 | [1.00-1.14] | 5.40E-02 | 0.51 |
| caes | rs2143950 | I3 | 2459 | 13 | 1.05 | [0.88-1.26] | 0.57 | 1.04 | [0.84-1.30] | 0.71 | 0.25 |
| caes | rs2143950 | S1 | 14021 | 13 | 0.97 | [0.79-1.20] | 0.78 | 0.95 | [0.73-1.26] | 0.71 | 0.20 |
| caes | rs2143950 | S2 | 15715 | 13 | 1.05 | [0.85-1.29] | 0.67 | 1.06 | [0.80-1.39] | 0.7 | 0.10 |
| caes | rs2143950 | S3 | 11030 | 13 | 1.00 | [0.79-1.26] | 0.98 | 1.00 | [0.71-1.40] | 0.99 | 0.07 |
| caes | rs2164983 | I1 | 15812 | 9 | 1.01 | [0.81-1.26] | 0.92 | 1.01 | [0.81-1.26] | 0.92 | 0.77 |
| caes | rs2164983 | I2 | 13897 | 9 | 1.14 | [1.05-1.23] | 1.20E-03 | 1.14 | [1.05-1.23] | 1.20E-03 | 0.70 |
| caes | rs2164983 | I3 | 1915 | 9 | 1.17 | [0.95-1.45] | 0.13 | 1.17 | [0.95-1.45] | 0.13 | 0.44 |
| caes | rs2164983 | S1 | 12584 | 9 | 1.00 | [0.79-1.27] | 0.99 | 1.00 | [0.79-1.27] | 0.99 | 0.61 |
| caes | rs2164983 | S2 | 13771 | 9 | 0.95 | [0.75-1.19] | 0.64 | 0.95 | [0.75-1.19] | 0.64 | 0.86 |
| caes | rs2164983 | S3 | 10558 | 9 | 0.94 | [0.73-1.20] | 0.61 | 0.94 | [0.73-1.20] | 0.61 | 0.77 |
| caes | rs2227483 | I1 | 18725 | 13 | 0.95 | [0.82-1.09] | 0.46 | 0.95 | [0.82-1.09] | 0.46 | 0.73 |
| caes | rs2227483 | I2 | 16263 | 13 | 1.06 | [1.01-1.12] | 2.80E-02 | 1.06 | [1.00-1.12] | 0.05 | 0.32 |
| caes | rs2227483 | I3 | 2462 | 13 | 1.01 | [0.89-1.16] | 0.85 | 1.01 | [0.88-1.16] | 0.86 | 0.43 |
| caes | rs2227483 | S1 | 14025 | 13 | 0.94 | [0.80-1.10] | 0.42 | 0.94 | [0.80-1.10] | 0.42 | 0.64 |
| caes | rs2227483 | S2 | 15719 | 13 | 0.97 | [0.83-1.14] | 0.72 | 0.97 | [0.83-1.14] | 0.72 | 0.63 |
| caes | rs2227483 | S3 | 11034 | 13 | 0.96 | [0.80-1.14] | 0.61 | 0.96 | [0.80-1.14] | 0.61 | 0.53 |
| caes | rs2228145 | I1 | 16772 | 11 | 0.93 | [0.79-1.08] | 0.33 | 0.93 | [0.79-1.08] | 0.33 | 0.90 |
| caes | rs2228145 | I2 | 14657 | 11 | 1.04 | [0.98-1.10] | 0.16 | 1.04 | [0.98-1.10] | 0.16 | 0.68 |
| caes | rs2228145 | I3 | 2115 | 11 | 0.96 | [0.83-1.10] | 0.54 | 0.96 | [0.83-1.10] | 0.54 | 0.61 |
| caes | rs2228145 | S1 | 13549 | 11 | 0.90 | [0.76-1.05] | 0.18 | 0.90 | [0.76-1.05] | 0.18 | 0.86 |
| caes | rs2228145 | S2 | 13766 | 11 | 0.98 | [0.83-1.16] | 0.84 | 0.98 | [0.83-1.16] | 0.84 | 0.83 |

| | | | | | | | | | | | |
|---|---|---|---|---|---|---|---|---|---|---|---|
| caes | rs2228145 | S3 | 10558 | 11 | 0.96 | [0.80-1.15] | 0.68 | 0.96 | [0.80-1.15] | 0.68 | 0.60 |
| caes | rs2897442 | I1 | 16920 | 11 | 0.91 | [0.77-1.09] | 0.3 | 0.91 | [0.77-1.09] | 0.3 | 0.61 |
| caes | rs2897442 | I2 | 14822 | 11 | 1.12 | [1.05-1.19] | 5.10E-04 | 1.12 | [1.01-1.24] | 3.30E-02 | 0.01 |
| caes | rs2897442 | I3 | 2098 | 11 | 1.04 | [0.89-1.23] | 0.62 | 1.04 | [0.89-1.23] | 0.62 | 0.76 |
| caes | rs2897442 | S1 | 12188 | 11 | 0.99 | [0.81-1.20] | 0.91 | 0.99 | [0.81-1.20] | 0.91 | 0.59 |
| caes | rs2897442 | S2 | 13914 | 11 | 0.89 | [0.73-1.08] | 0.24 | 0.89 | [0.73-1.08] | 0.24 | 0.47 |
| caes | rs2897442 | S3 | 9197 | 11 | 0.97 | [0.78-1.21] | 0.76 | 0.97 | [0.77-1.21] | 0.77 | 0.39 |
| caes | rs479844 | I1 | 16929 | 11 | 1.07 | [0.92-1.26] | 0.39 | 1.07 | [0.92-1.26] | 0.39 | 0.91 |
| caes | rs479844 | I2 | 14830 | 11 | 1.14 | [1.08-1.20] | 3.10E-06 | 1.14 | [1.07-1.22] | 9.80E-05 | 0.24 |
| caes | rs479844 | I3 | 2099 | 11 | 1.24 | [1.07-1.43] | 5.10E-03 | 1.24 | [1.07-1.43] | 5.10E-03 | 0.92 |
| caes | rs479844 | S1 | 12188 | 11 | 1.14 | [0.95-1.35] | 0.16 | 1.14 | [0.95-1.35] | 0.16 | 0.98 |
| caes | rs479844 | S2 | 13923 | 11 | 1.09 | [0.92-1.30] | 0.33 | 1.09 | [0.92-1.30] | 0.33 | 0.69 |
| caes | rs479844 | S3 | 9197 | 11 | 1.17 | [0.96-1.43] | 0.12 | 1.17 | [0.96-1.43] | 0.12 | 0.86 |
| caes | rs6010620 | I1 | 18290 | 12 | 0.95 | [0.79-1.13] | 0.53 | 0.93 | [0.74-1.18] | 0.5 | 0.13 |
| caes | rs6010620 | I2 | 15908 | 12 | 1.11 | [1.04-1.18] | 1.10E-03 | 1.11 | [1.04-1.18] | 1.10E-03 | 0.88 |
| caes | rs6010620 | I3 | 2382 | 12 | 1.05 | [0.89-1.23] | 0.57 | 1.03 | [0.85-1.27] | 0.73 | 0.23 |
| caes | rs6010620 | S1 | 13549 | 12 | 0.95 | [0.78-1.15] | 0.58 | 0.91 | [0.68-1.25] | 0.49 | 0.06 |
| caes | rs6010620 | S2 | 15284 | 12 | 0.97 | [0.80-1.18] | 0.78 | 0.95 | [0.73-1.27] | 0.71 | 0.09 |
| caes | rs6010620 | S3 | 10558 | 12 | 0.98 | [0.79-1.22] | 0.85 | 0.94 | [0.67-1.36] | 0.69 | 0.06 |
| caes | rs6473227 | I1 | 17328 | 12 | 1.05 | [0.90-1.21] | 0.55 | 1.05 | [0.90-1.21] | 0.55 | 0.91 |
| caes | rs6473227 | I2 | 15154 | 12 | 1.07 | [1.01-1.12] | 1.60E-02 | 1.07 | [1.01-1.12] | 1.60E-02 | 0.69 |
| caes | rs6473227 | I3 | 2174 | 12 | 1.10 | [0.96-1.26] | 0.16 | 1.10 | [0.96-1.26] | 0.16 | 0.74 |
| caes | rs6473227 | S1 | 12591 | 12 | 1.09 | [0.93-1.28] | 0.27 | 1.09 | [0.93-1.28] | 0.27 | 0.95 |
| caes | rs6473227 | S2 | 15760 | 12 | 1.04 | [0.89-1.22] | 0.63 | 1.04 | [0.89-1.22] | 0.63 | 0.87 |
| caes | rs6473227 | S3 | 11038 | 12 | 1.10 | [0.92-1.31] | 0.3 | 1.10 | [0.92-1.31] | 0.3 | 0.95 |
| caes | rs7127307 | I1 | 18769 | 13 | 0.92 | [0.79-1.07] | 0.28 | 0.89 | [0.74-1.11] | 0.25 | 0.15 |
| caes | rs7127307 | I2 | 16306 | 13 | 1.07 | [1.02-1.13] | 7.60E-03 | 1.07 | [1.02-1.13] | 7.60E-03 | 0.75 |
| caes | rs7127307 | I3 | 2463 | 13 | 0.98 | [0.85-1.13] | 0.82 | 0.96 | [0.81-1.17] | 0.68 | 0.17 |
| caes | rs7127307 | S1 | 14028 | 13 | 0.97 | [0.82-1.15] | 0.76 | 0.94 | [0.76-1.21] | 0.6 | 0.15 |
| caes | rs7127307 | S2 | 15763 | 13 | 0.95 | [0.80-1.12] | 0.51 | 0.93 | [0.75-1.17] | 0.49 | 0.13 |
| caes | rs7127307 | S3 | 11037 | 13 | 1.03 | [0.85-1.24] | 0.75 | 1.02 | [0.80-1.32] | 0.89 | 0.15 |
| caes | rs7146581 | I1 | 18761 | 13 | 0.88 | [0.75-1.05] | 0.16 | 0.88 | [0.75-1.05] | 0.16 | 0.50 |
| caes | rs7146581 | I2 | 16298 | 13 | 1.05 | [0.99-1.12] | 0.12 | 1.05 | [0.97-1.14] | 0.21 | 0.11 |
| caes | rs7146581 | I3 | 2463 | 13 | 0.95 | [0.81-1.11] | 0.48 | 0.95 | [0.81-1.11] | 0.48 | 0.55 |
| caes | rs7146581 | S1 | 14024 | 13 | 0.88 | [0.73-1.06] | 0.17 | 0.88 | [0.73-1.06] | 0.17 | 0.50 |
| caes | rs7146581 | S2 | 15755 | 13 | 0.92 | [0.76-1.10] | 0.35 | 0.92 | [0.76-1.10] | 0.35 | 0.50 |
| caes | rs7146581 | S3 | 11033 | 13 | 0.91 | [0.74-1.11] | 0.35 | 0.91 | [0.74-1.11] | 0.35 | 0.49 |
| caes | rs7927894 | I1 | 18012 | 12 | 1.14 | [0.98-1.33] | 8.30E-02 | 1.12 | [0.88-1.46] | 0.37 | 0.02 |
| caes | rs7927894 | I2 | 15646 | 12 | 1.01 | [0.95-1.06] | 0.84 | 1.00 | [0.92-1.09] | 0.96 | 0.06 |
| caes | rs7927894 | I3 | 2366 | 12 | 1.14 | [0.99-1.31] | 7.80E-02 | 1.12 | [0.93-1.37] | 0.23 | 0.13 |
| caes | rs7927894 | S1 | 13549 | 12 | 1.17 | [0.99-1.39] | 6.20E-02 | 1.13 | [0.84-1.58] | 0.42 | 0.01 |
| caes | rs7927894 | S2 | 15006 | 12 | 1.18 | [1.00-1.39] | 4.90E-02 | 1.18 | [1.00-1.39] | 4.90E-02 | 0.86 |
| caes | rs7927894 | S3 | 10558 | 12 | 1.23 | [1.02-1.48] | 2.70E-02 | 1.23 | [1.02-1.48] | 2.70E-02 | 0.86 |
| caes | | M1 | 20943 | 13 | 1.04 | [0.94-1.14] | 0.45 | 1.04 | [0.94-1.14] | 0.45 | 0.98 |
| caes | | M2 | 16179 | 13 | 1.04 | [0.94-1.15] | 0.42 | 1.04 | [0.94-1.15] | 0.42 | 0.95 |
| caes | | M3 | 17690 | 12 | 1.02 | [0.92-1.13] | 0.65 | 1.02 | [0.92-1.13] | 0.65 | 0.93 |
| caes | | M4 | 14166 | 11 | 1.02 | [0.91-1.13] | 0.77 | 1.02 | [0.91-1.13] | 0.77 | 0.88 |
| cat | flg | I1 | 16043 | 12 | 1.13 | [0.87-1.45] | 0.36 | 1.15 | [0.81-1.59] | 0.43 | 0.22 |
| cat | flg | I2 | 11829 | 12 | 1.84 | [1.63-2.09] | 9.80E-22 | 1.84 | [1.52-2.23] | 2.00E-10 | 0.05 |
| cat | flg | I3 | 4214 | 12 | 2.01 | [1.62-2.50] | 2.40E-10 | 2.02 | [1.56-2.61] | 8.50E-08 | 0.34 |
| cat | flg | S1 | 14488 | 12 | 1.11 | [0.85-1.46] | 0.44 | 1.16 | [0.75-1.72] | 0.5 | 0.11 |
| cat | flg | S2 | 14219 | 12 | 1.11 | [0.84-1.46] | 0.45 | 1.09 | [0.78-1.55] | 0.6 | 0.31 |
| cat | flg | S3 | 12707 | 12 | 1.12 | [0.84-1.50] | 0.44 | 1.10 | [0.71-1.74] | 0.66 | 0.15 |
| cat | rs10214237 | I1 | 14651 | 11 | 0.97 | [0.85-1.10] | 0.62 | 0.97 | [0.85-1.10] | 0.62 | 0.75 |
| cat | rs10214237 | I2 | 10782 | 11 | 1.11 | [1.04-1.18] | 1.50E-03 | 1.11 | [1.04-1.18] | 1.50E-03 | 0.52 |
| cat | rs10214237 | I3 | 3869 | 11 | 1.11 | [1.00-1.24] | 5.40E-02 | 1.11 | [1.00-1.24] | 5.40E-02 | 0.91 |
| cat | rs10214237 | S1 | 12869 | 11 | 0.98 | [0.85-1.12] | 0.72 | 0.98 | [0.85-1.12] | 0.72 | 0.68 |
| cat | rs10214237 | S2 | 13042 | 11 | 0.96 | [0.84-1.11] | 0.6 | 0.96 | [0.84-1.11] | 0.6 | 0.54 |
| cat | rs10214237 | S3 | 11288 | 11 | 0.98 | [0.85-1.14] | 0.8 | 0.98 | [0.85-1.14] | 0.8 | 0.51 |
| cat | rs1057258 | I1 | 14635 | 11 | 1.02 | [0.88-1.19] | 0.8 | 1.02 | [0.88-1.19] | 0.8 | 0.65 |
| cat | rs1057258 | I2 | 10769 | 11 | 1.05 | [0.97-1.13] | 0.25 | 1.05 | [0.97-1.13] | 0.25 | 0.76 |
| cat | rs1057258 | I3 | 3866 | 11 | 1.06 | [0.94-1.21] | 0.34 | 1.06 | [0.94-1.21] | 0.34 | 0.83 |
| cat | rs1057258 | S1 | 12870 | 11 | 1.05 | [0.89-1.23] | 0.57 | 1.05 | [0.89-1.23] | 0.57 | 0.48 |
| cat | rs1057258 | S2 | 13026 | 11 | 0.98 | [0.84-1.16] | 0.86 | 0.98 | [0.84-1.16] | 0.86 | 0.55 |
| cat | rs1057258 | S3 | 11289 | 11 | 1.00 | [0.84-1.19] | 0.98 | 1.00 | [0.83-1.20] | 0.98 | 0.39 |
| cat | rs10995251 | I1 | 14060 | 10 | 0.98 | [0.87-1.10] | 0.72 | 1.00 | [0.82-1.19] | 0.99 | 0.04 |
| cat | rs10995251 | I2 | 10389 | 10 | 1.05 | [0.98-1.11] | 0.15 | 1.04 | [0.94-1.15] | 0.43 | 0.01 |
| cat | rs10995251 | I3 | 3671 | 10 | 1.02 | [0.92-1.13] | 0.76 | 1.04 | [0.89-1.20] | 0.62 | 0.07 |
| cat | rs10995251 | S1 | 12280 | 10 | 0.94 | [0.83-1.07] | 0.34 | 0.94 | [0.78-1.14] | 0.53 | 0.11 |
| cat | rs10995251 | S2 | 12451 | 10 | 1.02 | [0.89-1.16] | 0.77 | 1.06 | [0.87-1.24] | 0.54 | 0.08 |
| cat | rs10995251 | S3 | 10699 | 10 | 0.97 | [0.85-1.12] | 0.71 | 0.98 | [0.81-1.18] | 0.87 | 0.16 |
| cat | rs112111458 | I1 | 12606 | 8 | 1.07 | [0.90-1.26] | 0.46 | 1.03 | [0.84-1.31] | 0.77 | 0.29 |
| cat | rs112111458 | I2 | 9181 | 8 | 1.00 | [0.92-1.09] | 1 | 1.05 | [0.91-1.16] | 0.5 | 0.01 |
| cat | rs112111458 | I3 | 3425 | 8 | 1.02 | [0.88-1.17] | 0.79 | 1.02 | [0.88-1.17] | 0.79 | 0.63 |
| cat | rs112111458 | S1 | 11912 | 8 | 1.11 | [0.93-1.32] | 0.26 | 1.11 | [0.93-1.32] | 0.26 | 0.80 |
| cat | rs112111458 | S2 | 11953 | 8 | 1.10 | [0.92-1.31] | 0.3 | 1.06 | [0.87-1.35] | 0.55 | 0.30 |
| cat | rs112111458 | S3 | 11287 | 8 | 1.15 | [0.96-1.38] | 0.14 | 1.15 | [0.96-1.38] | 0.14 | 0.65 |
| cat | rs11657987 | I1 | 14618 | 11 | 1.10 | [0.98-1.24] | 9.40E-02 | 1.10 | [0.94-1.30] | 0.24 | 0.12 |
| cat | rs11657987 | I2 | 10758 | 11 | 1.02 | [0.96-1.08] | 0.49 | 1.02 | [0.94-1.10] | 0.68 | 0.12 |
| cat | rs11657987 | I3 | 3860 | 11 | 1.13 | [1.02-1.24] | 1.80E-02 | 1.12 | [0.96-1.31] | 0.14 | 0.06 |
| cat | rs11657987 | S1 | 12866 | 11 | 1.10 | [0.97-1.24] | 0.13 | 1.08 | [0.89-1.33] | 0.42 | 0.06 |
| cat | rs11657987 | S2 | 13009 | 11 | 1.15 | [1.02-1.30] | 2.50E-02 | 1.15 | [1.02-1.30] | 2.50E-02 | 0.73 |
| cat | rs11657987 | S3 | 11285 | 11 | 1.15 | [1.01-1.31] | 0.04 | 1.15 | [1.01-1.31] | 0.04 | 0.53 |
| cat | rs12153855 | I1 | 12038 | 7 | 0.94 | [0.76-1.16] | 0.57 | 0.94 | [0.76-1.16] | 0.57 | 0.73 |
| cat | rs12153855 | I2 | 8805 | 7 | 1.14 | [1.03-1.26] | 1.50E-02 | 1.14 | [1.03-1.26] | 1.50E-02 | 0.96 |
| cat | rs12153855 | I3 | 3233 | 7 | 1.09 | [0.91-1.30] | 0.37 | 1.09 | [0.91-1.30] | 0.37 | 0.72 |
| cat | rs12153855 | S1 | 11324 | 7 | 0.95 | [0.76-1.18] | 0.63 | 0.95 | [0.76-1.18] | 0.63 | 0.63 |
| cat | rs12153855 | S2 | 11385 | 7 | 0.95 | [0.76-1.18] | 0.62 | 0.95 | [0.76-1.18] | 0.62 | 0.67 |
| cat | rs12153855 | S3 | 10699 | 7 | 0.95 | [0.75-1.19] | 0.66 | 0.95 | [0.75-1.19] | 0.66 | 0.60 |
| cat | rs12295535 | I1 | 14655 | 11 | 0.84 | [0.59-1.20] | 0.34 | 0.84 | [0.59-1.20] | 0.34 | 0.70 |
| cat | rs12295535 | I2 | 10785 | 11 | 1.20 | [1.01-1.42] | 4.20E-02 | 1.20 | [1.01-1.42] | 4.20E-02 | 0.47 |

| | | | | | | | | | | | |
|---|---|---|---|---|---|---|---|---|---|---|---|
| cat | rs12295535 | I3 | 3870 | 11 | 1.02 | [0.76-1.38] | 0.89 | 1.02 | [0.76-1.38] | 0.89 | 0.63 |
| cat | rs12295535 | S1 | 12870 | 11 | 0.81 | [0.56-1.17] | 0.26 | 0.81 | [0.56-1.17] | 0.26 | 0.53 |
| cat | rs12295535 | S2 | 13046 | 11 | 0.82 | [0.56-1.20] | 0.31 | 0.82 | [0.56-1.20] | 0.31 | 0.55 |
| cat | rs12295535 | S3 | 11289 | 11 | 0.78 | [0.52-1.17] | 0.23 | 0.78 | [0.52-1.17] | 0.23 | 0.45 |
| cat | rs13015714 | I1 | 14063 | 10 | 0.93 | [0.81-1.07] | 0.3 | 0.93 | [0.81-1.07] | 0.3 | 0.64 |
| cat | rs13015714 | I2 | 10391 | 10 | 1.15 | [1.07-1.23] | 6.40E-05 | 1.15 | [1.07-1.23] | 6.40E-05 | 0.59 |
| cat | rs13015714 | I3 | 3672 | 10 | 1.08 | [0.96-1.21] | 0.22 | 1.08 | [0.96-1.21] | 0.22 | 0.55 |
| cat | rs13015714 | S1 | 12278 | 10 | 0.93 | [0.80-1.07] | 0.32 | 0.93 | [0.80-1.07] | 0.32 | 0.51 |
| cat | rs13015714 | S2 | 12454 | 10 | 0.90 | [0.78-1.05] | 0.17 | 0.90 | [0.78-1.05] | 0.17 | 0.48 |
| cat | rs13015714 | S3 | 10697 | 10 | 0.91 | [0.78-1.06] | 0.21 | 0.89 | [0.75-1.08] | 0.18 | 0.34 |
| cat | rs16948048 | I1 | 14063 | 10 | 0.86 | [0.76-0.97] | 1.20E-02 | 0.90 | [0.74-1.04] | 0.29 | 0.05 |
| cat | rs16948048 | I2 | 10390 | 10 | 1.07 | [1.01-1.13] | 3.30E-02 | 1.07 | [1.01-1.13] | 3.30E-02 | 0.98 |
| cat | rs16948048 | I3 | 3673 | 10 | 0.90 | [0.82-1.00] | 5.60E-02 | 0.94 | [0.79-1.08] | 0.47 | 0.04 |
| cat | rs16948048 | S1 | 12280 | 10 | 0.88 | [0.77-1.00] | 4.30E-02 | 0.95 | [0.76-1.09] | 0.67 | 0.04 |
| cat | rs16948048 | S2 | 12454 | 10 | 0.84 | [0.74-0.96] | 1.10E-02 | 0.84 | [0.74-0.96] | 1.10E-02 | 0.52 |
| cat | rs16948048 | S3 | 10699 | 10 | 0.86 | [0.75-0.98] | 2.90E-02 | 0.86 | [0.75-0.98] | 2.90E-02 | 0.46 |
| cat | rs17389644 | I1 | 14053 | 10 | 1.00 | [0.86-1.15] | 0.97 | 1.00 | [0.86-1.15] | 0.97 | 0.69 |
| cat | rs17389644 | I2 | 10385 | 10 | 1.05 | [0.98-1.12] | 0.21 | 1.03 | [0.95-1.13] | 0.5 | 0.25 |
| cat | rs17389644 | I3 | 3668 | 10 | 1.05 | [0.93-1.18] | 0.45 | 1.02 | [0.86-1.24] | 0.85 | 0.19 |
| cat | rs17389644 | S1 | 12280 | 10 | 1.01 | [0.87-1.18] | 0.85 | 1.01 | [0.87-1.18] | 0.85 | 0.52 |
| cat | rs17389644 | S2 | 12444 | 10 | 0.94 | [0.80-1.10] | 0.42 | 0.94 | [0.80-1.10] | 0.42 | 0.78 |
| cat | rs17389644 | S3 | 10699 | 10 | 0.96 | [0.81-1.13] | 0.6 | 0.96 | [0.81-1.13] | 0.6 | 0.58 |
| cat | rs17881320 | I1 | 14635 | 11 | 0.98 | [0.79-1.20] | 0.82 | 0.98 | [0.79-1.20] | 0.82 | 0.96 |
| cat | rs17881320 | I2 | 10769 | 11 | 1.13 | [1.02-1.26] | 1.80E-02 | 1.14 | [1.00-1.29] | 5.90E-02 | 0.17 |
| cat | rs17881320 | I3 | 3866 | 11 | 1.12 | [0.94-1.33] | 0.22 | 1.12 | [0.94-1.33] | 0.22 | 0.86 |
| cat | rs17881320 | S1 | 12868 | 11 | 0.95 | [0.76-1.20] | 0.69 | 0.95 | [0.76-1.20] | 0.69 | 0.94 |
| cat | rs17881320 | S2 | 13026 | 11 | 0.98 | [0.78-1.23] | 0.86 | 0.98 | [0.78-1.23] | 0.86 | 0.83 |
| cat | rs17881320 | S3 | 11287 | 11 | 0.95 | [0.74-1.20] | 0.65 | 0.95 | [0.74-1.20] | 0.65 | 0.94 |
| cat | rs2041733 | I1 | 14064 | 10 | 0.98 | [0.87-1.10] | 0.74 | 0.99 | [0.82-1.17] | 0.88 | 0.07 |
| cat | rs2041733 | I2 | 10391 | 10 | 1.04 | [0.98-1.10] | 0.18 | 1.04 | [0.98-1.10] | 0.18 | 0.53 |
| cat | rs2041733 | I3 | 3673 | 10 | 1.03 | [0.93-1.14] | 0.6 | 1.03 | [0.90-1.18] | 0.66 | 0.16 |
| cat | rs2041733 | S1 | 12279 | 10 | 1.03 | [0.91-1.17] | 0.64 | 1.03 | [0.91-1.17] | 0.64 | 0.44 |
| cat | rs2041733 | S2 | 12455 | 10 | 0.99 | [0.87-1.12] | 0.88 | 0.99 | [0.81-1.21] | 0.9 | 0.05 |
| cat | rs2041733 | S3 | 10698 | 10 | 1.04 | [0.91-1.19] | 0.54 | 1.05 | [0.91-1.20] | 0.51 | 0.38 |
| cat | rs2143950 | I1 | 14642 | 11 | 1.12 | [0.97-1.30] | 0.13 | 1.09 | [0.91-1.35] | 0.36 | 0.28 |
| cat | rs2143950 | I2 | 10775 | 11 | 1.02 | [0.94-1.10] | 0.68 | 1.02 | [0.94-1.10] | 0.68 | 0.61 |
| cat | rs2143950 | I3 | 3867 | 11 | 1.14 | [1.01-1.29] | 3.70E-02 | 1.12 | [0.97-1.31] | 0.13 | 0.37 |
| cat | rs2143950 | S1 | 12861 | 11 | 1.13 | [0.97-1.33] | 0.12 | 1.07 | [0.84-1.44] | 0.57 | 0.10 |
| cat | rs2143950 | S2 | 13033 | 11 | 1.12 | [0.95-1.31] | 0.18 | 1.10 | [0.89-1.37] | 0.38 | 0.22 |
| cat | rs2143950 | S3 | 11280 | 11 | 1.13 | [0.95-1.34] | 0.17 | 1.10 | [0.84-1.47] | 0.48 | 0.08 |
| cat | rs2164983 | I1 | 12038 | 7 | 1.00 | [0.84-1.18] | 0.97 | 1.00 | [0.84-1.18] | 0.97 | 0.74 |
| cat | rs2164983 | I2 | 8805 | 7 | 1.13 | [1.04-1.23] | 5.30E-03 | 1.11 | [1.00-1.26] | 5.60E-02 | 0.21 |
| cat | rs2164983 | I3 | 3233 | 7 | 1.15 | [1.00-1.32] | 4.50E-02 | 1.15 | [1.00-1.32] | 4.50E-02 | 0.67 |
| cat | rs2164983 | S1 | 11324 | 7 | 0.99 | [0.83-1.17] | 0.89 | 0.99 | [0.83-1.17] | 0.89 | 0.57 |
| cat | rs2164983 | S2 | 11385 | 7 | 1.00 | [0.84-1.19] | 1 | 1.00 | [0.84-1.19] | 1 | 0.75 |
| cat | rs2164983 | S3 | 10699 | 7 | 0.99 | [0.82-1.18] | 0.89 | 0.99 | [0.82-1.18] | 0.89 | 0.62 |
| cat | rs2227483 | I1 | 14644 | 11 | 1.13 | [1.01-1.27] | 3.70E-02 | 1.15 | [0.94-1.39] | 0.18 | 0.01 |
| cat | rs2227483 | I2 | 10775 | 11 | 1.03 | [0.97-1.09] | 0.4 | 1.02 | [0.94-1.11] | 0.67 | 0.05 |
| cat | rs2227483 | I3 | 3869 | 11 | 1.15 | [1.04-1.27] | 7.40E-03 | 1.15 | [0.97-1.36] | 9.90E-02 | 0.03 |
| cat | rs2227483 | S1 | 12863 | 11 | 1.11 | [0.98-1.25] | 0.11 | 1.08 | [0.86-1.38] | 0.51 | 0.01 |
| cat | rs2227483 | S2 | 13035 | 11 | 1.12 | [0.99-1.27] | 7.00E-02 | 1.07 | [0.87-1.38] | 0.51 | 0.03 |
| cat | rs2227483 | S3 | 11282 | 11 | 1.08 | [0.95-1.24] | 0.23 | 0.99 | [0.79-1.35] | 0.91 | 0.04 |
| cat | rs2228145 | I1 | 12994 | 9 | 1.14 | [1.01-1.29] | 3.70E-02 | 1.14 | [1.01-1.29] | 3.70E-02 | 0.61 |
| cat | rs2228145 | I2 | 9572 | 9 | 1.00 | [0.94-1.06] | 0.99 | 1.00 | [0.94-1.06] | 0.99 | 0.51 |
| cat | rs2228145 | I3 | 3422 | 9 | 1.15 | [1.04-1.27] | 8.80E-03 | 1.15 | [1.04-1.27] | 8.80E-03 | 0.62 |
| cat | rs2228145 | S1 | 12280 | 9 | 1.14 | [1.00-1.29] | 4.80E-02 | 1.14 | [1.00-1.29] | 4.80E-02 | 0.72 |
| cat | rs2228145 | S2 | 11385 | 9 | 1.10 | [0.96-1.25] | 0.16 | 1.10 | [0.96-1.25] | 0.16 | 0.89 |
| cat | rs2228145 | S3 | 10699 | 9 | 1.11 | [0.97-1.27] | 0.14 | 1.11 | [0.97-1.27] | 0.14 | 0.90 |
| cat | rs2897442 | I1 | 12702 | 9 | 0.87 | [0.75-1.00] | 4.40E-02 | 0.88 | [0.75-1.01] | 0.11 | 0.35 |
| cat | rs2897442 | I2 | 9501 | 9 | 1.13 | [1.06-1.21] | 3.90E-04 | 1.13 | [1.01-1.26] | 2.60E-02 | 0.02 |
| cat | rs2897442 | I3 | 3201 | 9 | 1.01 | [0.90-1.14] | 0.86 | 1.02 | [0.89-1.15] | 0.77 | 0.39 |
| cat | rs2897442 | S1 | 10918 | 9 | 0.84 | [0.72-0.97] | 0.02 | 0.84 | [0.71-0.99] | 4.40E-02 | 0.37 |
| cat | rs2897442 | S2 | 11093 | 9 | 0.91 | [0.78-1.06] | 0.23 | 0.91 | [0.78-1.06] | 0.23 | 0.53 |
| cat | rs2897442 | S3 | 9337 | 9 | 0.88 | [0.75-1.04] | 0.13 | 0.88 | [0.75-1.04] | 0.13 | 0.56 |
| cat | rs479844 | I1 | 12703 | 9 | 0.98 | [0.86-1.11] | 0.74 | 0.98 | [0.86-1.11] | 0.74 | 0.54 |
| cat | rs479844 | I2 | 9502 | 9 | 1.14 | [1.07-1.21] | 3.50E-05 | 1.14 | [1.06-1.22] | 2.60E-04 | 0.32 |
| cat | rs479844 | I3 | 3201 | 9 | 1.12 | [1.01-1.25] | 3.70E-02 | 1.12 | [1.01-1.25] | 3.70E-02 | 0.67 |
| cat | rs479844 | S1 | 10918 | 9 | 1.03 | [0.90-1.18] | 0.65 | 1.03 | [0.90-1.18] | 0.65 | 0.69 |
| cat | rs479844 | S2 | 11094 | 9 | 0.95 | [0.83-1.09] | 0.45 | 0.95 | [0.83-1.09] | 0.45 | 0.76 |
| cat | rs479844 | S3 | 9337 | 9 | 0.99 | [0.86-1.15] | 0.92 | 0.99 | [0.86-1.15] | 0.92 | 0.91 |
| cat | rs6010620 | I1 | 14065 | 10 | 1.06 | [0.92-1.21] | 0.43 | 1.05 | [0.87-1.28] | 0.63 | 0.16 |
| cat | rs6010620 | I2 | 10392 | 10 | 1.09 | [1.02-1.17] | 1.70E-02 | 1.09 | [1.02-1.17] | 1.70E-02 | 0.78 |
| cat | rs6010620 | I3 | 3673 | 10 | 1.13 | [1.01-1.27] | 3.20E-02 | 1.13 | [0.96-1.33] | 0.13 | 0.19 |
| cat | rs6010620 | S1 | 12280 | 10 | 1.09 | [0.94-1.26] | 0.24 | 1.10 | [0.89-1.34] | 0.39 | 0.15 |
| cat | rs6010620 | S2 | 12456 | 10 | 1.03 | [0.89-1.19] | 0.69 | 1.03 | [0.82-1.30] | 0.8 | 0.06 |
| cat | rs6010620 | S3 | 10699 | 10 | 1.07 | [0.91-1.25] | 0.41 | 1.10 | [0.84-1.39] | 0.49 | 0.05 |
| cat | rs6473227 | I1 | 14655 | 11 | 0.96 | [0.85-1.08] | 0.47 | 0.93 | [0.79-1.13] | 0.39 | 0.15 |
| cat | rs6473227 | I2 | 10785 | 11 | 1.08 | [1.02-1.14] | 0.01 | 1.08 | [1.02-1.14] | 0.01 | 0.52 |
| cat | rs6473227 | I3 | 3870 | 11 | 1.04 | [0.94-1.15] | 0.47 | 1.01 | [0.89-1.17] | 0.9 | 0.26 |
| cat | rs6473227 | S1 | 12870 | 11 | 0.95 | [0.84-1.08] | 0.46 | 0.92 | [0.76-1.15] | 0.37 | 0.09 |
| cat | rs6473227 | S2 | 13046 | 11 | 0.92 | [0.82-1.05] | 0.22 | 0.91 | [0.76-1.11] | 0.28 | 0.11 |
| cat | rs6473227 | S3 | 11289 | 11 | 0.92 | [0.81-1.05] | 0.22 | 0.90 | [0.73-1.13] | 0.3 | 0.08 |
| cat | rs7127307 | I1 | 14654 | 11 | 0.92 | [0.82-1.03] | 0.15 | 0.92 | [0.82-1.03] | 0.15 | 0.97 |
| cat | rs7127307 | I2 | 10785 | 11 | 1.08 | [1.01-1.14] | 1.70E-02 | 1.08 | [1.01-1.14] | 1.70E-02 | 0.62 |
| cat | rs7127307 | I3 | 3869 | 11 | 0.98 | [0.88-1.08] | 0.63 | 0.98 | [0.88-1.08] | 0.63 | 1.00 |
| cat | rs7127307 | S1 | 12869 | 11 | 0.91 | [0.80-1.03] | 0.13 | 0.91 | [0.80-1.03] | 0.13 | 0.93 |
| cat | rs7127307 | S2 | 13045 | 11 | 0.95 | [0.84-1.08] | 0.45 | 0.95 | [0.84-1.08] | 0.45 | 0.98 |
| cat | rs7127307 | S3 | 11288 | 11 | 0.94 | [0.83-1.08] | 0.4 | 0.94 | [0.83-1.08] | 0.4 | 0.90 |
| cat | rs7146581 | I1 | 14647 | 11 | 0.99 | [0.86-1.13] | 0.87 | 0.99 | [0.86-1.13] | 0.87 | 0.52 |
| cat | rs7146581 | I2 | 10780 | 11 | 1.03 | [0.97-1.11] | 0.35 | 1.03 | [0.97-1.11] | 0.35 | 0.72 |
| cat | rs7146581 | I3 | 3867 | 11 | 1.02 | [0.90-1.14] | 0.78 | 1.01 | [0.87-1.18] | 0.87 | 0.25 |

| | | | | | | | | | | | |
|---|---|---|---|---|---|---|---|---|---|---|---|
| cat | rs7146581 | S1 | 12864 | 11 | 0.98 | [0.84-1.13] | 0.75 | 0.98 | [0.84-1.13] | 0.75 | 0.66 |
| cat | rs7146581 | S2 | 13038 | 11 | 0.93 | [0.81-1.08] | 0.37 | 0.93 | [0.81-1.08] | 0.37 | 0.44 |
| cat | rs7146581 | S3 | 11283 | 11 | 0.93 | [0.79-1.08] | 0.33 | 0.93 | [0.79-1.08] | 0.33 | 0.47 |
| cat | rs7927894 | I1 | 14026 | 10 | 1.08 | [0.96-1.22] | 0.19 | 1.08 | [0.96-1.22] | 0.19 | 0.48 |
| cat | rs7927894 | I2 | 10355 | 10 | 0.99 | [0.93-1.05] | 0.76 | 0.98 | [0.92-1.06] | 0.62 | 0.33 |
| cat | rs7927894 | I3 | 3671 | 10 | 1.07 | [0.96-1.18] | 0.22 | 1.07 | [0.96-1.18] | 0.22 | 0.65 |
| cat | rs7927894 | S1 | 12280 | 10 | 1.04 | [0.91-1.18] | 0.56 | 1.04 | [0.91-1.18] | 0.56 | 0.84 |
| cat | rs7927894 | S2 | 12417 | 10 | 1.08 | [0.95-1.23] | 0.24 | 1.09 | [0.95-1.24] | 0.24 | 0.37 |
| cat | rs7927894 | S3 | 10699 | 10 | 1.03 | [0.90-1.18] | 0.7 | 1.03 | [0.90-1.18] | 0.7 | 0.85 |
| cat | | M1 | 18041 | 12 | 0.96 | [0.89-1.03] | 0.23 | 0.96 | [0.89-1.03] | 0.23 | 0.52 |
| cat | | M2 | 16222 | 12 | 0.96 | [0.89-1.04] | 0.33 | 0.95 | [0.87-1.05] | 0.26 | 0.33 |
| cat | | M3 | 16170 | 12 | 0.97 | [0.90-1.05] | 0.46 | 0.97 | [0.90-1.05] | 0.46 | 0.87 |
| cat | | M4 | 14394 | 12 | 0.98 | [0.90-1.06] | 0.57 | 0.98 | [0.90-1.06] | 0.57 | 0.84 |
| dog | flg | I1 | 16044 | 12 | 0.94 | [0.72-1.24] | 0.67 | 0.84 | [0.58-1.38] | 0.38 | 0.16 |
| dog | flg | I2 | 12889 | 12 | 1.93 | [1.71-2.19] | 3.10E-26 | 2.00 | [1.66-2.33] | 1.80E-13 | 0.05 |
| dog | flg | I3 | 3155 | 12 | 1.74 | [1.38-2.21] | 4.50E-06 | 1.66 | [1.24-2.32] | 5.60E-04 | 0.31 |
| dog | flg | S1 | 14489 | 12 | 1.03 | [0.77-1.38] | 0.83 | 0.94 | [0.61-1.59] | 0.76 | 0.14 |
| dog | flg | S2 | 14221 | 12 | 0.97 | [0.72-1.32] | 0.87 | 0.85 | [0.54-1.54] | 0.49 | 0.09 |
| dog | flg | S3 | 12709 | 12 | 1.12 | [0.81-1.55] | 0.49 | 1.00 | [0.58-1.92] | 1 | 0.07 |
| dog | rs10214237 | I1 | 14656 | 11 | 0.83 | [0.72-0.96] | 1.10E-02 | 0.82 | [0.70-0.97] | 1.40E-02 | 0.37 |
| dog | rs10214237 | I2 | 11778 | 11 | 1.14 | [1.08-1.22] | 1.50E-05 | 1.14 | [1.08-1.22] | 1.50E-05 | 0.86 |
| dog | rs10214237 | I3 | 2878 | 11 | 0.98 | [0.86-1.11] | 0.71 | 0.95 | [0.80-1.15] | 0.51 | 0.23 |
| dog | rs10214237 | S1 | 12874 | 11 | 0.83 | [0.71-0.97] | 2.10E-02 | 0.83 | [0.71-0.97] | 2.10E-02 | 0.64 |
| dog | rs10214237 | S2 | 13046 | 11 | 0.83 | [0.71-0.97] | 2.30E-02 | 0.83 | [0.70-0.98] | 2.60E-02 | 0.39 |
| dog | rs10214237 | S3 | 11292 | 11 | 0.85 | [0.71-1.00] | 5.20E-02 | 0.85 | [0.71-1.00] | 5.20E-02 | 0.63 |
| dog | rs1057258 | I1 | 14640 | 11 | 1.10 | [0.93-1.31] | 0.27 | 1.10 | [0.92-1.32] | 0.31 | 0.38 |
| dog | rs1057258 | I2 | 11762 | 11 | 1.03 | [0.96-1.11] | 0.38 | 1.03 | [0.96-1.11] | 0.38 | 0.95 |
| dog | rs1057258 | I3 | 2878 | 11 | 1.14 | [0.98-1.33] | 8.30E-02 | 1.14 | [0.98-1.33] | 8.30E-02 | 0.49 |
| dog | rs1057258 | S1 | 12875 | 11 | 1.09 | [0.91-1.31] | 0.34 | 1.09 | [0.91-1.31] | 0.34 | 0.46 |
| dog | rs1057258 | S2 | 13030 | 11 | 1.10 | [0.91-1.32] | 0.31 | 1.11 | [0.89-1.37] | 0.35 | 0.27 |
| dog | rs1057258 | S3 | 11293 | 11 | 1.09 | [0.89-1.32] | 0.41 | 1.10 | [0.87-1.38] | 0.44 | 0.27 |
| dog | rs10995251 | I1 | 14065 | 10 | 0.89 | [0.78-1.02] | 9.80E-02 | 0.89 | [0.78-1.02] | 9.80E-02 | 0.90 |
| dog | rs10995251 | I2 | 11354 | 10 | 1.05 | [0.99-1.11] | 7.90E-02 | 1.06 | [0.97-1.14] | 0.18 | 0.07 |
| dog | rs10995251 | I3 | 2711 | 10 | 0.96 | [0.85-1.09] | 0.54 | 0.97 | [0.85-1.10] | 0.62 | 0.37 |
| dog | rs10995251 | S1 | 12285 | 10 | 0.89 | [0.77-1.03] | 0.12 | 0.89 | [0.77-1.03] | 0.12 | 0.77 |
| dog | rs10995251 | S2 | 12455 | 10 | 0.88 | [0.76-1.02] | 8.70E-02 | 0.88 | [0.76-1.02] | 8.70E-02 | 0.93 |
| dog | rs10995251 | S3 | 10703 | 10 | 0.88 | [0.75-1.03] | 0.12 | 0.88 | [0.75-1.03] | 0.12 | 0.84 |
| dog | rs112111458 | I1 | 12610 | 8 | 0.94 | [0.77-1.14] | 0.51 | 0.94 | [0.77-1.14] | 0.51 | 0.51 |
| dog | rs112111458 | I2 | 10192 | 8 | 1.02 | [0.94-1.11] | 0.66 | 1.06 | [0.93-1.17] | 0.39 | 0.02 |
| dog | rs112111458 | I3 | 2418 | 8 | 0.96 | [0.81-1.14] | 0.61 | 0.96 | [0.81-1.14] | 0.61 | 0.78 |
| dog | rs112111458 | S1 | 11916 | 8 | 0.96 | [0.79-1.18] | 0.72 | 0.96 | [0.79-1.18] | 0.72 | 0.56 |
| dog | rs112111458 | S2 | 11957 | 8 | 0.93 | [0.76-1.14] | 0.51 | 0.93 | [0.76-1.14] | 0.51 | 0.56 |
| dog | rs112111458 | S3 | 11291 | 8 | 0.96 | [0.77-1.18] | 0.67 | 0.96 | [0.77-1.18] | 0.67 | 0.60 |
| dog | rs11657987 | I1 | 14623 | 11 | 0.91 | [0.80-1.04] | 0.15 | 0.90 | [0.76-1.07] | 0.21 | 0.22 |
| dog | rs11657987 | I2 | 11750 | 11 | 1.06 | [1.00-1.12] | 3.40E-02 | 1.06 | [0.99-1.14] | 0.12 | 0.15 |
| dog | rs11657987 | I3 | 2873 | 11 | 0.99 | [0.88-1.11] | 0.87 | 0.99 | [0.83-1.19] | 0.91 | 0.06 |
| dog | rs11657987 | S1 | 12871 | 11 | 0.94 | [0.81-1.08] | 0.36 | 0.94 | [0.78-1.12] | 0.5 | 0.22 |
| dog | rs11657987 | S2 | 13013 | 11 | 0.90 | [0.78-1.04] | 0.15 | 0.89 | [0.76-1.05] | 0.14 | 0.34 |
| dog | rs11657987 | S3 | 11289 | 11 | 0.93 | [0.80-1.09] | 0.37 | 0.93 | [0.79-1.10] | 0.37 | 0.37 |
| dog | rs12153855 | I1 | 12042 | 7 | 0.92 | [0.72-1.18] | 0.53 | 0.92 | [0.72-1.18] | 0.53 | 0.50 |
| dog | rs12153855 | I2 | 9788 | 7 | 1.13 | [1.03-1.25] | 1.30E-02 | 1.13 | [1.03-1.25] | 1.30E-02 | 0.59 |
| dog | rs12153855 | I3 | 2254 | 7 | 1.07 | [0.87-1.33] | 0.52 | 1.07 | [0.87-1.33] | 0.52 | 0.77 |
| dog | rs12153855 | S1 | 11328 | 7 | 0.92 | [0.71-1.19] | 0.52 | 0.93 | [0.69-1.25] | 0.66 | 0.31 |
| dog | rs12153855 | S2 | 11389 | 7 | 0.89 | [0.69-1.15] | 0.39 | 0.89 | [0.69-1.15] | 0.39 | 0.44 |
| dog | rs12153855 | S3 | 10703 | 7 | 0.89 | [0.68-1.17] | 0.4 | 0.90 | [0.65-1.23] | 0.54 | 0.29 |
| dog | rs12295535 | I1 | 14660 | 11 | 1.31 | [0.87-1.97] | 0.19 | 1.31 | [0.87-1.97] | 0.19 | 0.92 |
| dog | rs12295535 | I2 | 11780 | 11 | 1.09 | [0.93-1.29] | 0.29 | 1.09 | [0.90-1.32] | 0.4 | 0.29 |
| dog | rs12295535 | I3 | 2880 | 11 | 1.43 | [1.01-2.04] | 4.50E-02 | 1.43 | [1.01-2.04] | 4.50E-02 | 0.87 |
| dog | rs12295535 | S1 | 12875 | 11 | 1.16 | [0.73-1.82] | 0.53 | 1.16 | [0.73-1.82] | 0.53 | 0.91 |
| dog | rs12295535 | S2 | 13050 | 11 | 1.25 | [0.81-1.93] | 0.31 | 1.25 | [0.81-1.93] | 0.31 | 0.70 |
| dog | rs12295535 | S3 | 11293 | 11 | 1.06 | [0.66-1.71] | 0.8 | 1.06 | [0.66-1.71] | 0.8 | 0.82 |
| dog | rs13015714 | I1 | 14068 | 10 | 1.00 | [0.85-1.16] | 0.95 | 1.06 | [0.85-1.23] | 0.6 | 0.15 |
| dog | rs13015714 | I2 | 11357 | 10 | 1.12 | [1.05-1.20] | 5.40E-04 | 1.12 | [1.05-1.20] | 5.40E-04 | 0.91 |
| dog | rs13015714 | I3 | 2711 | 10 | 1.15 | [1.00-1.32] | 5.50E-02 | 1.22 | [0.98-1.43] | 7.50E-02 | 0.06 |
| dog | rs13015714 | S1 | 12283 | 10 | 0.96 | [0.81-1.13] | 0.61 | 0.96 | [0.81-1.13] | 0.61 | 0.45 |
| dog | rs13015714 | S2 | 12458 | 10 | 1.06 | [0.89-1.25] | 0.51 | 1.12 | [0.88-1.35] | 0.37 | 0.12 |
| dog | rs13015714 | S3 | 10701 | 10 | 1.01 | [0.84-1.22] | 0.88 | 1.02 | [0.83-1.25] | 0.83 | 0.32 |
| dog | rs16948048 | I1 | 14068 | 10 | 1.09 | [0.95-1.25] | 0.2 | 1.09 | [0.95-1.25] | 0.2 | 0.69 |
| dog | rs16948048 | I2 | 11358 | 10 | 1.01 | [0.96-1.07] | 0.64 | 1.01 | [0.96-1.07] | 0.64 | 0.60 |
| dog | rs16948048 | I3 | 2710 | 10 | 1.10 | [0.97-1.24] | 0.13 | 1.10 | [0.97-1.24] | 0.13 | 0.77 |
| dog | rs16948048 | S1 | 12285 | 10 | 1.08 | [0.93-1.25] | 0.3 | 1.08 | [0.93-1.25] | 0.3 | 0.53 |
| dog | rs16948048 | S2 | 12458 | 10 | 1.13 | [0.97-1.31] | 0.11 | 1.13 | [0.97-1.31] | 0.11 | 0.87 |
| dog | rs16948048 | S3 | 10703 | 10 | 1.12 | [0.95-1.31] | 0.18 | 1.12 | [0.95-1.31] | 0.18 | 0.75 |
| dog | rs17389644 | I1 | 14058 | 10 | 0.92 | [0.78-1.08] | 0.29 | 0.92 | [0.78-1.08] | 0.29 | 0.93 |
| dog | rs17389644 | I2 | 11350 | 10 | 1.06 | [0.99-1.13] | 7.90E-02 | 1.03 | [0.94-1.16] | 0.56 | 0.09 |
| dog | rs17389644 | I3 | 2708 | 10 | 0.97 | [0.84-1.12] | 0.7 | 0.97 | [0.84-1.12] | 0.7 | 0.78 |
| dog | rs17389644 | S1 | 12285 | 10 | 0.89 | [0.75-1.06] | 0.2 | 0.89 | [0.75-1.06] | 0.2 | 0.98 |
| dog | rs17389644 | S2 | 12448 | 10 | 0.91 | [0.77-1.08] | 0.28 | 0.91 | [0.77-1.08] | 0.28 | 0.95 |
| dog | rs17389644 | S3 | 10703 | 10 | 0.89 | [0.73-1.07] | 0.21 | 0.89 | [0.73-1.07] | 0.21 | 0.98 |
| dog | rs17881320 | I1 | 14640 | 11 | 0.84 | [0.66-1.08] | 0.17 | 0.83 | [0.62-1.13] | 0.19 | 0.30 |
| dog | rs17881320 | I2 | 11762 | 11 | 1.15 | [1.05-1.27] | 4.20E-03 | 1.15 | [1.01-1.32] | 3.90E-02 | 0.11 |
| dog | rs17881320 | I3 | 2878 | 11 | 1.02 | [0.82-1.26] | 0.87 | 1.00 | [0.78-1.31] | 0.98 | 0.32 |
| dog | rs17881320 | S1 | 12873 | 11 | 0.81 | [0.62-1.05] | 0.12 | 0.77 | [0.55-1.15] | 0.15 | 0.23 |
| dog | rs17881320 | S2 | 13030 | 11 | 0.76 | [0.58-1.00] | 0.05 | 0.72 | [0.49-1.11] | 8.90E-02 | 0.14 |
| dog | rs17881320 | S3 | 11291 | 11 | 0.69 | [0.51-0.94] | 1.70E-02 | 0.67 | [0.47-0.99] | 2.50E-02 | 0.31 |
| dog | rs2041733 | I1 | 14069 | 10 | 1.04 | [0.91-1.19] | 0.52 | 1.05 | [0.91-1.21] | 0.53 | 0.36 |
| dog | rs2041733 | I2 | 11359 | 10 | 1.03 | [0.98-1.09] | 0.28 | 1.03 | [0.97-1.09] | 0.31 | 0.37 |
| dog | rs2041733 | I3 | 2710 | 10 | 1.07 | [0.95-1.20] | 0.29 | 1.07 | [0.95-1.20] | 0.29 | 0.77 |
| dog | rs2041733 | S1 | 12284 | 10 | 1.07 | [0.93-1.24] | 0.33 | 1.09 | [0.91-1.28] | 0.34 | 0.26 |
| dog | rs2041733 | S2 | 12459 | 10 | 1.05 | [0.91-1.22] | 0.47 | 1.05 | [0.90-1.23] | 0.51 | 0.39 |
| dog | rs2041733 | S3 | 10702 | 10 | 1.09 | [0.93-1.27] | 0.3 | 1.08 | [0.90-1.30] | 0.4 | 0.32 |

| | | | | | | | | | | | |
|---|---|---|---|---|---|---|---|---|---|---|---|
| dog | rs2143950 | I1 | 14647 | 11 | 0.98 | [0.83-1.16] | 0.83 | 1.02 | [0.82-1.22] | 0.85 | 0.21 |
| dog | rs2143950 | I2 | 11769 | 11 | 1.06 | [0.98-1.14] | 0.13 | 1.06 | [0.98-1.14] | 0.13 | 0.47 |
| dog | rs2143950 | I3 | 2878 | 11 | 1.03 | [0.88-1.19] | 0.74 | 1.03 | [0.87-1.21] | 0.72 | 0.37 |
| dog | rs2143950 | S1 | 12866 | 11 | 0.91 | [0.76-1.10] | 0.33 | 0.95 | [0.75-1.16] | 0.67 | 0.21 |
| dog | rs2143950 | S2 | 13037 | 11 | 0.96 | [0.79-1.15] | 0.64 | 0.99 | [0.78-1.22] | 0.95 | 0.17 |
| dog | rs2143950 | S3 | 11284 | 11 | 0.87 | [0.71-1.06] | 0.17 | 0.88 | [0.70-1.10] | 0.28 | 0.30 |
| dog | rs2164983 | I1 | 12042 | 7 | 0.93 | [0.76-1.13] | 0.46 | 0.98 | [0.76-1.20] | 0.89 | 0.24 |
| dog | rs2164983 | I2 | 9788 | 7 | 1.15 | [1.06-1.24] | 9.50E-04 | 1.12 | [0.98-1.30] | 8.70E-02 | 0.05 |
| dog | rs2164983 | I3 | 2254 | 7 | 1.11 | [0.93-1.33] | 0.25 | 1.11 | [0.93-1.33] | 0.25 | 0.63 |
| dog | rs2164983 | S1 | 11328 | 7 | 0.95 | [0.77-1.17] | 0.65 | 1.03 | [0.77-1.28] | 0.85 | 0.19 |
| dog | rs2164983 | S2 | 11389 | 7 | 0.96 | [0.78-1.18] | 0.7 | 1.01 | [0.77-1.26] | 0.93 | 0.22 |
| dog | rs2164983 | S3 | 10703 | 7 | 0.99 | [0.79-1.23] | 0.91 | 1.05 | [0.78-1.34] | 0.73 | 0.18 |
| dog | rs2227483 | I1 | 14649 | 11 | 1.01 | [0.89-1.16] | 0.84 | 1.01 | [0.89-1.16] | 0.84 | 0.97 |
| dog | rs2227483 | I2 | 11772 | 11 | 1.05 | [1.00-1.11] | 6.90E-02 | 1.05 | [0.99-1.12] | 8.90E-02 | 0.39 |
| dog | rs2227483 | I3 | 2877 | 11 | 1.07 | [0.95-1.21] | 0.23 | 1.07 | [0.95-1.21] | 0.23 | 0.67 |
| dog | rs2227483 | S1 | 12868 | 11 | 1.00 | [0.86-1.15] | 0.96 | 1.00 | [0.86-1.15] | 0.96 | 0.92 |
| dog | rs2227483 | S2 | 13039 | 11 | 1.00 | [0.87-1.16] | 0.96 | 1.00 | [0.87-1.16] | 0.96 | 0.91 |
| dog | rs2227483 | S3 | 11286 | 11 | 0.99 | [0.85-1.16] | 0.94 | 0.99 | [0.85-1.16] | 0.94 | 0.79 |
| dog | rs2228145 | I1 | 12999 | 9 | 1.01 | [0.88-1.16] | 0.88 | 0.97 | [0.82-1.20] | 0.72 | 0.30 |
| dog | rs2228145 | I2 | 10566 | 9 | 1.03 | [0.97-1.09] | 0.29 | 1.04 | [0.97-1.11] | 0.26 | 0.16 |
| dog | rs2228145 | I3 | 2433 | 9 | 1.06 | [0.94-1.20] | 0.35 | 1.06 | [0.94-1.20] | 0.35 | 0.68 |
| dog | rs2228145 | S1 | 12285 | 9 | 1.00 | [0.86-1.16] | 1 | 0.91 | [0.73-1.25] | 0.42 | 0.10 |
| dog | rs2228145 | S2 | 11389 | 9 | 0.98 | [0.84-1.15] | 0.83 | 0.95 | [0.79-1.18] | 0.55 | 0.29 |
| dog | rs2228145 | S3 | 10703 | 9 | 0.98 | [0.83-1.15] | 0.76 | 0.90 | [0.71-1.24] | 0.38 | 0.13 |
| dog | rs2897442 | I1 | 12707 | 9 | 0.94 | [0.80-1.11] | 0.47 | 0.94 | [0.80-1.11] | 0.47 | 0.59 |
| dog | rs2897442 | I2 | 10485 | 9 | 1.11 | [1.04-1.18] | 2.10E-03 | 1.12 | [1.01-1.22] | 2.60E-02 | 0.05 |
| dog | rs2897442 | I3 | 2222 | 9 | 1.07 | [0.92-1.24] | 0.39 | 1.07 | [0.92-1.24] | 0.39 | 0.65 |
| dog | rs2897442 | S1 | 10923 | 9 | 0.95 | [0.79-1.13] | 0.55 | 0.94 | [0.76-1.16] | 0.52 | 0.36 |
| dog | rs2897442 | S2 | 11097 | 9 | 0.96 | [0.80-1.15] | 0.67 | 0.96 | [0.80-1.15] | 0.67 | 0.80 |
| dog | rs2897442 | S3 | 9341 | 9 | 0.97 | [0.80-1.19] | 0.8 | 0.97 | [0.80-1.19] | 0.8 | 0.56 |
| dog | rs479844 | I1 | 12708 | 9 | 1.01 | [0.87-1.17] | 0.86 | 1.01 | [0.87-1.17] | 0.86 | 0.94 |
| dog | rs479844 | I2 | 10486 | 9 | 1.13 | [1.07-1.20] | 2.80E-05 | 1.13 | [1.06-1.20] | 7.50E-05 | 0.39 |
| dog | rs479844 | I3 | 2222 | 9 | 1.15 | [1.01-1.32] | 3.60E-02 | 1.15 | [1.01-1.32] | 3.60E-02 | 0.91 |
| dog | rs479844 | S1 | 10923 | 9 | 1.04 | [0.89-1.22] | 0.64 | 1.04 | [0.89-1.22] | 0.64 | 0.81 |
| dog | rs479844 | S2 | 11098 | 9 | 1.02 | [0.87-1.20] | 0.82 | 1.02 | [0.87-1.20] | 0.82 | 0.91 |
| dog | rs479844 | S3 | 9341 | 9 | 1.05 | [0.88-1.26] | 0.61 | 1.05 | [0.88-1.26] | 0.61 | 0.79 |
| dog | rs6010620 | I1 | 14070 | 10 | 0.97 | [0.83-1.14] | 0.72 | 1.00 | [0.79-1.22] | 0.98 | 0.10 |
| dog | rs6010620 | I2 | 11359 | 10 | 1.11 | [1.04-1.19] | 1.90E-03 | 1.11 | [1.04-1.19] | 1.90E-03 | 0.52 |
| dog | rs6010620 | I3 | 2711 | 10 | 1.07 | [0.93-1.23] | 0.32 | 1.09 | [0.90-1.29] | 0.39 | 0.18 |
| dog | rs6010620 | S1 | 12285 | 10 | 1.02 | [0.86-1.21] | 0.82 | 1.10 | [0.85-1.32] | 0.48 | 0.11 |
| dog | rs6010620 | S2 | 12460 | 10 | 0.94 | [0.79-1.11] | 0.45 | 0.94 | [0.74-1.19] | 0.61 | 0.14 |
| dog | rs6010620 | S3 | 10703 | 10 | 0.97 | [0.81-1.17] | 0.77 | 1.01 | [0.77-1.29] | 0.93 | 0.11 |
| dog | rs6473227 | I1 | 14660 | 11 | 1.06 | [0.93-1.21] | 0.41 | 1.06 | [0.93-1.21] | 0.41 | 0.95 |
| dog | rs6473227 | I2 | 11780 | 11 | 1.06 | [1.00-1.12] | 4.60E-02 | 1.06 | [1.00-1.12] | 4.60E-02 | 0.77 |
| dog | rs6473227 | I3 | 2880 | 11 | 1.12 | [0.99-1.25] | 6.70E-02 | 1.12 | [0.99-1.25] | 6.70E-02 | 0.96 |
| dog | rs6473227 | S1 | 12875 | 11 | 1.06 | [0.92-1.21] | 0.45 | 1.06 | [0.92-1.21] | 0.45 | 0.94 |
| dog | rs6473227 | S2 | 13050 | 11 | 1.06 | [0.92-1.22] | 0.41 | 1.06 | [0.92-1.22] | 0.41 | 0.85 |
| dog | rs6473227 | S3 | 11293 | 11 | 1.07 | [0.92-1.25] | 0.38 | 1.07 | [0.92-1.25] | 0.38 | 0.78 |
| dog | rs7127307 | I1 | 14659 | 11 | 0.96 | [0.84-1.09] | 0.53 | 0.98 | [0.84-1.13] | 0.83 | 0.27 |
| dog | rs7127307 | I2 | 11780 | 11 | 1.05 | [0.99-1.11] | 7.90E-02 | 1.05 | [0.99-1.12] | 0.13 | 0.37 |
| dog | rs7127307 | I3 | 2879 | 11 | 1.00 | [0.89-1.13] | 0.97 | 1.00 | [0.89-1.13] | 0.97 | 0.49 |
| dog | rs7127307 | S1 | 12874 | 11 | 0.94 | [0.82-1.09] | 0.43 | 0.97 | [0.79-1.16] | 0.75 | 0.13 |
| dog | rs7127307 | S2 | 13049 | 11 | 0.97 | [0.84-1.12] | 0.66 | 1.01 | [0.83-1.18] | 0.91 | 0.15 |
| dog | rs7127307 | S3 | 11292 | 11 | 0.96 | [0.82-1.12] | 0.57 | 0.99 | [0.78-1.22] | 0.97 | 0.08 |
| dog | rs7146581 | I1 | 14652 | 11 | 1.12 | [0.96-1.31] | 0.15 | 1.12 | [0.96-1.31] | 0.15 | 0.89 |
| dog | rs7146581 | I2 | 11773 | 11 | 1.01 | [0.95-1.08] | 0.79 | 1.00 | [0.93-1.09] | 0.92 | 0.23 |
| dog | rs7146581 | I3 | 2879 | 11 | 1.14 | [0.99-1.31] | 6.30E-02 | 1.14 | [0.99-1.31] | 6.30E-02 | 0.98 |
| dog | rs7146581 | S1 | 12869 | 11 | 1.11 | [0.94-1.31] | 0.22 | 1.11 | [0.94-1.31] | 0.22 | 0.83 |
| dog | rs7146581 | S2 | 13042 | 11 | 1.15 | [0.97-1.36] | 0.11 | 1.15 | [0.97-1.36] | 0.11 | 0.85 |
| dog | rs7146581 | S3 | 11287 | 11 | 1.14 | [0.95-1.37] | 0.16 | 1.14 | [0.95-1.37] | 0.16 | 0.81 |
| dog | rs7927894 | I1 | 14031 | 10 | 0.93 | [0.81-1.07] | 0.31 | 0.93 | [0.81-1.07] | 0.31 | 0.88 |
| dog | rs7927894 | I2 | 11324 | 10 | 1.02 | [0.96-1.08] | 0.48 | 1.02 | [0.96-1.08] | 0.52 | 0.42 |
| dog | rs7927894 | I3 | 2707 | 10 | 0.96 | [0.85-1.08] | 0.5 | 0.96 | [0.85-1.08] | 0.5 | 0.98 |
| dog | rs7927894 | S1 | 12285 | 10 | 0.92 | [0.80-1.07] | 0.29 | 0.92 | [0.80-1.07] | 0.29 | 0.72 |
| dog | rs7927894 | S2 | 12421 | 10 | 0.92 | [0.79-1.07] | 0.27 | 0.92 | [0.79-1.07] | 0.27 | 0.79 |
| dog | rs7927894 | S3 | 10703 | 10 | 0.92 | [0.78-1.08] | 0.29 | 0.92 | [0.78-1.08] | 0.29 | 0.58 |
| dog | | M1 | 18045 | 12 | 0.91 | [0.84-0.99] | 0.03 | 0.91 | [0.84-0.99] | 0.03 | 0.68 |
| dog | | M2 | 16226 | 12 | 0.91 | [0.83-0.99] | 3.20E-02 | 0.91 | [0.82-1.00] | 6.60E-02 | 0.34 |
| dog | | M3 | 16174 | 12 | 0.93 | [0.85-1.02] | 0.13 | 0.93 | [0.85-1.02] | 0.13 | 0.86 |
| dog | | M4 | 14398 | 12 | 0.93 | [0.85-1.02] | 0.13 | 0.93 | [0.85-1.02] | 0.13 | 0.51 |
| dura_brfed | flg | I1 | 13468 | 9 | 0.99 | [0.96-1.03] | 0.74 | 0.99 | [0.96-1.03] | 0.74 | 0.56 |
| dura_brfed | flg | S1 | 12828 | 9 | 0.99 | [0.95-1.02] | 0.47 | 0.99 | [0.95-1.02] | 0.47 | 0.86 |
| dura_brfed | flg | S2 | 12763 | 9 | 0.99 | [0.96-1.03] | 0.59 | 0.99 | [0.96-1.03] | 0.59 | 0.51 |
| dura_brfed | flg | S3 | 12147 | 9 | 0.98 | [0.95-1.02] | 0.35 | 0.98 | [0.95-1.02] | 0.35 | 0.85 |
| dura_brfed | rs10214237 | I1 | 15019 | 10 | 1.00 | [0.99-1.02] | 0.57 | 1.01 | [0.99-1.02] | 0.55 | 0.36 |
| dura_brfed | rs10214237 | S1 | 13437 | 10 | 1.00 | [0.98-1.02] | 0.84 | 1.00 | [0.98-1.02] | 0.84 | 0.55 |
| dura_brfed | rs10214237 | S2 | 12260 | 10 | 1.00 | [0.99-1.02] | 0.76 | 1.00 | [0.99-1.02] | 0.7 | 0.34 |
| dura_brfed | rs10214237 | S3 | 10694 | 10 | 1.00 | [0.98-1.02] | 0.97 | 1.00 | [0.98-1.02] | 0.97 | 0.52 |
| dura_brfed | rs1057258 | I1 | 15003 | 10 | 1.01 | [0.99-1.03] | 0.2 | 1.01 | [0.99-1.03] | 0.2 | 0.53 |
| dura_brfed | rs1057258 | S1 | 13438 | 10 | 1.02 | [1.00-1.04] | 0.12 | 1.02 | [1.00-1.04] | 0.12 | 0.43 |
| dura_brfed | rs1057258 | S2 | 12244 | 10 | 1.01 | [0.99-1.03] | 0.16 | 1.01 | [0.99-1.03] | 0.16 | 0.45 |
| dura_brfed | rs1057258 | S3 | 10695 | 10 | 1.02 | [1.00-1.04] | 9.20E-02 | 1.02 | [1.00-1.04] | 0.11 | 0.34 |
| dura_brfed | rs10995251 | I1 | 14471 | 9 | 1.01 | [0.99-1.03] | 0.22 | 1.01 | [0.99-1.03] | 0.24 | 0.30 |
| dura_brfed | rs10995251 | S1 | 12891 | 9 | 1.01 | [0.99-1.02] | 0.5 | 1.01 | [0.99-1.02] | 0.5 | 0.40 |
| dura_brfed | rs10995251 | S2 | 11712 | 9 | 1.01 | [0.99-1.02] | 0.31 | 1.01 | [0.99-1.03] | 0.34 | 0.24 |
| dura_brfed | rs10995251 | S3 | 10148 | 9 | 1.00 | [0.99-1.02] | 0.63 | 1.00 | [0.99-1.02] | 0.64 | 0.27 |
| dura_brfed | rs112111458 | I1 | 14111 | 9 | 1.01 | [0.99-1.03] | 0.44 | 1.00 | [0.98-1.04] | 0.74 | 0.12 |
| dura_brfed | rs112111458 | S1 | 13436 | 9 | 1.01 | [0.99-1.03] | 0.4 | 1.01 | [0.97-1.04] | 0.69 | 0.11 |
| dura_brfed | rs112111458 | S2 | 11352 | 9 | 1.01 | [0.99-1.03] | 0.42 | 1.01 | [0.98-1.03] | 0.64 | 0.24 |
| dura_brfed | rs112111458 | S3 | 10693 | 9 | 1.01 | [0.99-1.03] | 0.33 | 1.01 | [0.98-1.04] | 0.53 | 0.25 |
| dura_brfed | rs11657987 | I1 | 14984 | 10 | 1.00 | [0.99-1.02] | 0.77 | 1.00 | [0.99-1.02] | 0.75 | 0.37 |

| | | | | | | | | | | | |
|---|---|---|---|---|---|---|---|---|---|---|---|
| dura_brfed | rs11657987 | S1 | 13434 | 10 | 1.00 | [0.98-1.01] | 0.58 | 1.00 | [0.98-1.01] | 0.58 | 0.69 |
| dura_brfed | rs11657987 | S2 | 12225 | 10 | 1.00 | [0.98-1.01] | 0.89 | 1.00 | [0.98-1.02] | 0.99 | 0.20 |
| dura_brfed | rs11657987 | S3 | 10691 | 10 | 0.99 | [0.98-1.01] | 0.38 | 0.99 | [0.98-1.01] | 0.38 | 0.43 |
| dura_brfed | rs12153855 | I1 | 13585 | 8 | 0.98 | [0.96-1.01] | 0.21 | 0.99 | [0.96-1.01] | 0.42 | 0.24 |
| dura_brfed | rs12153855 | S1 | 12891 | 8 | 0.98 | [0.95-1.00] | 0.1 | 0.98 | [0.95-1.01] | 0.3 | 0.18 |
| dura_brfed | rs12153855 | S2 | 10826 | 8 | 0.99 | [0.96-1.01] | 0.28 | 0.99 | [0.96-1.01] | 0.43 | 0.32 |
| dura_brfed | rs12153855 | S3 | 10148 | 8 | 0.98 | [0.95-1.01] | 0.14 | 0.98 | [0.95-1.01] | 0.35 | 0.22 |
| dura_brfed | rs12295535 | I1 | 15023 | 10 | 1.00 | [0.95-1.05] | 0.97 | 1.00 | [0.95-1.05] | 0.97 | 0.76 |
| dura_brfed | rs12295535 | S1 | 13438 | 10 | 1.01 | [0.95-1.06] | 0.78 | 1.01 | [0.95-1.06] | 0.78 | 0.56 |
| dura_brfed | rs12295535 | S2 | 12264 | 10 | 0.99 | [0.94-1.05] | 0.84 | 0.99 | [0.94-1.05] | 0.84 | 0.53 |
| dura_brfed | rs12295535 | S3 | 10695 | 10 | 1.01 | [0.95-1.06] | 0.82 | 1.01 | [0.95-1.06] | 0.82 | 0.50 |
| dura_brfed | rs13015714 | I1 | 14474 | 9 | 1.02 | [1.00-1.03] | 4.90E-02 | 1.02 | [1.00-1.04] | 6.80E-02 | 0.32 |
| dura_brfed | rs13015714 | S1 | 12889 | 9 | 1.01 | [0.99-1.03] | 0.17 | 1.02 | [0.99-1.05] | 0.26 | 0.03 |
| dura_brfed | rs13015714 | S2 | 11715 | 9 | 1.02 | [1.00-1.04] | 0.05 | 1.02 | [1.00-1.04] | 7.10E-02 | 0.26 |
| dura_brfed | rs13015714 | S3 | 10146 | 9 | 1.01 | [0.99-1.03] | 0.16 | 1.02 | [0.99-1.05] | 0.23 | 0.02 |
| dura_brfed | rs16948048 | I1 | 14474 | 9 | 1.00 | [0.98-1.01] | 0.86 | 1.00 | [0.98-1.01] | 0.86 | 0.49 |
| dura_brfed | rs16948048 | S1 | 12891 | 9 | 1.00 | [0.98-1.02] | 0.98 | 1.00 | [0.98-1.02] | 0.89 | 0.28 |
| dura_brfed | rs16948048 | S2 | 11715 | 9 | 1.00 | [0.98-1.01] | 0.88 | 1.00 | [0.98-1.01] | 0.86 | 0.39 |
| dura_brfed | rs16948048 | S3 | 10148 | 9 | 1.00 | [0.98-1.02] | 0.93 | 1.00 | [0.98-1.02] | 0.83 | 0.21 |
| dura_brfed | rs17389644 | I1 | 14463 | 9 | 1.01 | [0.99-1.02] | 0.58 | 1.01 | [0.99-1.02] | 0.58 | 0.68 |
| dura_brfed | rs17389644 | S1 | 12891 | 9 | 1.00 | [0.98-1.02] | 0.84 | 1.00 | [0.98-1.02] | 0.84 | 0.56 |
| dura_brfed | rs17389644 | S2 | 11704 | 9 | 1.01 | [0.99-1.02] | 0.59 | 1.01 | [0.99-1.02] | 0.59 | 0.84 |
| dura_brfed | rs17389644 | S3 | 10148 | 9 | 1.00 | [0.98-1.02] | 0.97 | 1.00 | [0.98-1.02] | 0.97 | 0.75 |
| dura_brfed | rs17881320 | I1 | 15004 | 10 | 1.01 | [0.98-1.04] | 0.4 | 1.01 | [0.98-1.04] | 0.46 | 0.31 |
| dura_brfed | rs17881320 | S1 | 13436 | 10 | 1.01 | [0.98-1.04] | 0.71 | 1.01 | [0.98-1.04] | 0.71 | 0.52 |
| dura_brfed | rs17881320 | S2 | 12245 | 10 | 1.02 | [0.99-1.05] | 0.2 | 1.02 | [0.99-1.05] | 0.26 | 0.27 |
| dura_brfed | rs17881320 | S3 | 10693 | 10 | 1.01 | [0.98-1.04] | 0.47 | 1.01 | [0.98-1.04] | 0.47 | 0.52 |
| dura_brfed | rs2041733 | I1 | 14475 | 9 | 0.99 | [0.98-1.01] | 0.38 | 0.99 | [0.98-1.01] | 0.45 | 0.32 |
| dura_brfed | rs2041733 | S1 | 12890 | 9 | 0.99 | [0.97-1.00] | 0.12 | 0.99 | [0.97-1.00] | 0.12 | 0.74 |
| dura_brfed | rs2041733 | S2 | 11716 | 9 | 0.99 | [0.98-1.01] | 0.46 | 0.99 | [0.98-1.01] | 0.52 | 0.35 |
| dura_brfed | rs2041733 | S3 | 10147 | 9 | 0.99 | [0.97-1.00] | 0.18 | 0.99 | [0.97-1.00] | 0.18 | 0.73 |
| dura_brfed | rs2143950 | I1 | 15010 | 10 | 1.01 | [0.99-1.03] | 0.29 | 1.01 | [0.99-1.03] | 0.29 | 0.97 |
| dura_brfed | rs2143950 | S1 | 13429 | 10 | 1.01 | [0.99-1.03] | 0.31 | 1.01 | [0.99-1.03] | 0.31 | 0.92 |
| dura_brfed | rs2143950 | S2 | 12251 | 10 | 1.01 | [0.99-1.03] | 0.39 | 1.01 | [0.99-1.03] | 0.39 | 0.98 |
| dura_brfed | rs2143950 | S3 | 10686 | 10 | 1.01 | [0.99-1.03] | 0.53 | 1.01 | [0.99-1.03] | 0.53 | 0.98 |
| dura_brfed | rs2164983 | I1 | 13585 | 8 | 1.00 | [0.98-1.02] | 0.96 | 1.00 | [0.98-1.02] | 0.96 | 0.93 |
| dura_brfed | rs2164983 | S1 | 12891 | 8 | 1.00 | [0.98-1.02] | 0.92 | 1.00 | [0.98-1.02] | 0.92 | 0.97 |
| dura_brfed | rs2164983 | S2 | 10826 | 8 | 1.00 | [0.98-1.02] | 0.9 | 1.00 | [0.98-1.02] | 0.9 | 0.91 |
| dura_brfed | rs2164983 | S3 | 10148 | 8 | 1.00 | [0.98-1.03] | 0.91 | 1.00 | [0.98-1.03] | 0.91 | 0.99 |
| dura_brfed | rs2227483 | I1 | 15012 | 10 | 0.99 | [0.97-1.00] | 0.13 | 0.99 | [0.97-1.00] | 0.13 | 0.94 |
| dura_brfed | rs2227483 | S1 | 13431 | 10 | 0.99 | [0.98-1.01] | 0.35 | 0.99 | [0.98-1.01] | 0.35 | 0.87 |
| dura_brfed | rs2227483 | S2 | 12253 | 10 | 0.99 | [0.98-1.01] | 0.2 | 0.99 | [0.98-1.01] | 0.2 | 0.92 |
| dura_brfed | rs2227483 | S3 | 10688 | 10 | 0.99 | [0.98-1.01] | 0.47 | 0.99 | [0.98-1.01] | 0.47 | 0.73 |
| dura_brfed | rs2228145 | I1 | 13585 | 8 | 1.00 | [0.98-1.01] | 0.84 | 1.00 | [0.98-1.01] | 0.85 | 0.37 |
| dura_brfed | rs2228145 | S1 | 12891 | 8 | 1.00 | [0.98-1.02] | 0.98 | 1.00 | [0.98-1.02] | 0.98 | 0.44 |
| dura_brfed | rs2228145 | S2 | 10826 | 8 | 1.00 | [0.98-1.01] | 0.62 | 1.00 | [0.98-1.01] | 0.65 | 0.28 |
| dura_brfed | rs2228145 | S3 | 10148 | 8 | 1.00 | [0.98-1.02] | 0.87 | 1.00 | [0.98-1.02] | 0.9 | 0.32 |
| dura_brfed | rs2897442 | I1 | 13343 | 8 | 1.00 | [0.98-1.02] | 0.78 | 1.00 | [0.97-1.03] | 0.95 | 0.16 |
| dura_brfed | rs2897442 | S1 | 11759 | 8 | 1.01 | [0.98-1.03] | 0.64 | 1.00 | [0.97-1.03] | 0.88 | 0.21 |
| dura_brfed | rs2897442 | S2 | 10584 | 8 | 1.00 | [0.98-1.02] | 0.97 | 1.00 | [0.97-1.03] | 0.76 | 0.11 |
| dura_brfed | rs2897442 | S3 | 9016 | 8 | 1.00 | [0.98-1.03] | 0.76 | 1.00 | [0.97-1.03] | 0.96 | 0.18 |
| dura_brfed | rs479844 | I1 | 13344 | 8 | 1.02 | [1.00-1.03] | 8.40E-02 | 1.02 | [1.00-1.03] | 8.40E-02 | 0.97 |
| dura_brfed | rs479844 | S1 | 11759 | 8 | 1.01 | [0.99-1.03] | 0.16 | 1.01 | [0.99-1.03] | 0.16 | 0.98 |
| dura_brfed | rs479844 | S2 | 10585 | 8 | 1.01 | [0.99-1.03] | 0.19 | 1.01 | [0.99-1.03] | 0.19 | 0.99 |
| dura_brfed | rs479844 | S3 | 9016 | 8 | 1.01 | [0.99-1.03] | 0.24 | 1.01 | [0.99-1.03] | 0.24 | 0.99 |
| dura_brfed | rs6010620 | I1 | 14476 | 9 | 1.00 | [0.99-1.02] | 0.74 | 1.00 | [0.99-1.02] | 0.74 | 0.77 |
| dura_brfed | rs6010620 | S1 | 12891 | 9 | 1.00 | [0.98-1.02] | 0.97 | 1.00 | [0.98-1.02] | 0.97 | 0.43 |
| dura_brfed | rs6010620 | S2 | 11717 | 9 | 1.00 | [0.98-1.02] | 0.81 | 1.00 | [0.98-1.02] | 0.81 | 0.96 |
| dura_brfed | rs6010620 | S3 | 10148 | 9 | 1.00 | [0.98-1.02] | 0.98 | 1.00 | [0.98-1.02] | 0.98 | 0.69 |
| dura_brfed | rs6473227 | I1 | 15023 | 10 | 1.00 | [0.99-1.02] | 0.52 | 1.00 | [0.99-1.02] | 0.52 | 0.67 |
| dura_brfed | rs6473227 | S1 | 13438 | 10 | 1.01 | [0.99-1.02] | 0.37 | 1.01 | [0.99-1.03] | 0.44 | 0.23 |
| dura_brfed | rs6473227 | S2 | 12264 | 10 | 1.01 | [0.99-1.02] | 0.45 | 1.01 | [0.99-1.02] | 0.45 | 0.64 |
| dura_brfed | rs6473227 | S3 | 10695 | 10 | 1.01 | [0.99-1.02] | 0.37 | 1.01 | [0.99-1.03] | 0.4 | 0.22 |
| dura_brfed | rs7127307 | I1 | 15022 | 10 | 1.00 | [0.98-1.01] | 0.83 | 1.00 | [0.98-1.02] | 0.98 | 0.17 |
| dura_brfed | rs7127307 | S1 | 13437 | 10 | 1.00 | [0.98-1.01] | 0.59 | 1.00 | [0.98-1.01] | 0.59 | 0.52 |
| dura_brfed | rs7127307 | S2 | 12263 | 10 | 1.00 | [0.98-1.01] | 0.9 | 1.00 | [0.98-1.02] | 0.93 | 0.14 |
| dura_brfed | rs7127307 | S3 | 10694 | 10 | 1.00 | [0.98-1.01] | 0.72 | 1.00 | [0.98-1.01] | 0.73 | 0.40 |
| dura_brfed | rs7146581 | I1 | 15015 | 10 | 0.99 | [0.98-1.01] | 0.45 | 1.00 | [0.97-1.02] | 0.73 | 0.17 |
| dura_brfed | rs7146581 | S1 | 13432 | 10 | 0.99 | [0.97-1.01] | 0.16 | 0.99 | [0.97-1.01] | 0.16 | 0.45 |
| dura_brfed | rs7146581 | S2 | 12256 | 10 | 0.99 | [0.97-1.01] | 0.41 | 1.00 | [0.97-1.01] | 0.65 | 0.24 |
| dura_brfed | rs7146581 | S3 | 10689 | 10 | 0.99 | [0.97-1.01] | 0.19 | 0.99 | [0.97-1.01] | 0.19 | 0.47 |
| dura_brfed | rs7927894 | I1 | 14435 | 9 | 1.00 | [0.99-1.02] | 0.78 | 1.00 | [0.99-1.02] | 0.78 | 0.58 |
| dura_brfed | rs7927894 | S1 | 12891 | 9 | 1.00 | [0.98-1.02] | 0.93 | 1.00 | [0.98-1.02] | 0.99 | 0.34 |
| dura_brfed | rs7927894 | S2 | 11676 | 9 | 1.00 | [0.99-1.02] | 0.63 | 1.00 | [0.99-1.02] | 0.63 | 0.49 |
| dura_brfed | rs7927894 | S3 | 10148 | 9 | 1.00 | [0.99-1.02] | 0.8 | 1.00 | [0.98-1.02] | 0.9 | 0.25 |
| dura_brfed | | M1 | 17101 | 10 | 1.00 | [0.99-1.01] | 0.69 | 1.00 | [0.98-1.02] | 0.81 | 0.00 |
| dura_brfed | | M2 | 15496 | 10 | 1.00 | [0.99-1.01] | 0.66 | 1.00 | [0.98-1.02] | 0.66 | 0.00 |
| dura_brfed | | M3 | 14515 | 9 | 1.00 | [0.99-1.01] | 0.92 | 1.00 | [0.98-1.02] | 0.88 | 0.00 |
| dura_brfed | | M4 | 13825 | 9 | 1.00 | [0.99-1.01] | 0.78 | 1.00 | [0.98-1.02] | 0.99 | 0.00 |
| ever_brfed | flg | I1 | 14091 | 9 | 1.29 | [0.91-1.85] | 0.16 | 1.14 | [0.72-2.06] | 0.58 | 0.27 |
| ever_brfed | flg | I2 | 1733 | 9 | 1.58 | [1.14-2.20] | 6.10E-03 | 1.82 | [1.14-2.53] | 1.20E-02 | 0.20 |
| ever_brfed | flg | I3 | 12358 | 9 | 1.95 | [1.72-2.21] | 7.60E-26 | 1.95 | [1.72-2.21] | 1.50E-25 | 0.43 |
| ever_brfed | flg | S1 | 13434 | 9 | 1.26 | [0.86-1.85] | 0.24 | 1.07 | [0.61-2.20] | 0.82 | 0.23 |
| ever_brfed | flg | S2 | 13312 | 9 | 1.17 | [0.80-1.71] | 0.41 | 1.11 | [0.71-1.83] | 0.65 | 0.34 |
| ever_brfed | flg | S3 | 12690 | 9 | 1.13 | [0.75-1.70] | 0.56 | 1.09 | [0.69-1.78] | 0.7 | 0.39 |
| ever_brfed | rs10214237 | I1 | 14719 | 9 | 1.02 | [0.85-1.22] | 0.85 | 1.02 | [0.85-1.22] | 0.85 | 0.93 |
| ever_brfed | rs10214237 | I2 | 2595 | 9 | 1.11 | [0.94-1.31] | 0.22 | 1.11 | [0.94-1.31] | 0.22 | 0.93 |
| ever_brfed | rs10214237 | I3 | 12124 | 9 | 1.11 | [1.05-1.18] | 8.10E-04 | 1.11 | [1.05-1.18] | 8.10E-04 | 0.82 |
| ever_brfed | rs10214237 | S1 | 14023 | 9 | 1.02 | [0.84-1.24] | 0.84 | 1.02 | [0.84-1.24] | 0.84 | 0.77 |
| ever_brfed | rs10214237 | S2 | 11907 | 9 | 1.01 | [0.83-1.23] | 0.93 | 1.01 | [0.83-1.23] | 0.93 | 0.85 |
| ever_brfed | rs10214237 | S3 | 11233 | 9 | 1.02 | [0.82-1.26] | 0.87 | 1.02 | [0.82-1.26] | 0.87 | 0.66 |

| | | | | | | | | | | | |
|---|---|---|---|---|---|---|---|---|---|---|---|
| ever_brfed | rs1057258 | I1 | 14703 | 9 | 1.16 | [0.93-1.43] | 0.18 | 1.16 | [0.93-1.43] | 0.18 | 0.78 |
| ever_brfed | rs1057258 | I2 | 2588 | 9 | 0.91 | [0.75-1.11] | 0.37 | 0.91 | [0.75-1.11] | 0.37 | 0.66 |
| ever_brfed | rs1057258 | I3 | 12115 | 9 | 1.06 | [0.99-1.14] | 0.11 | 1.06 | [0.99-1.14] | 0.11 | 0.98 |
| ever_brfed | rs1057258 | S1 | 14024 | 9 | 1.12 | [0.89-1.41] | 0.32 | 1.12 | [0.89-1.41] | 0.32 | 0.73 |
| ever_brfed | rs1057258 | S2 | 11891 | 9 | 1.19 | [0.95-1.50] | 0.13 | 1.19 | [0.95-1.50] | 0.13 | 0.75 |
| ever_brfed | rs1057258 | S3 | 11234 | 9 | 1.16 | [0.91-1.48] | 0.23 | 1.16 | [0.91-1.48] | 0.23 | 0.67 |
| ever_brfed | rs10995251 | I1 | 14171 | 8 | 1.07 | [0.90-1.27] | 0.44 | 1.07 | [0.90-1.27] | 0.44 | 0.50 |
| ever_brfed | rs10995251 | I2 | 2511 | 8 | 0.96 | [0.82-1.13] | 0.64 | 0.98 | [0.80-1.18] | 0.86 | 0.22 |
| ever_brfed | rs10995251 | I3 | 11660 | 8 | 1.05 | [0.99-1.11] | 0.1 | 1.06 | [0.96-1.15] | 0.22 | 0.04 |
| ever_brfed | rs10995251 | S1 | 13477 | 8 | 1.07 | [0.89-1.29] | 0.44 | 1.07 | [0.89-1.29] | 0.44 | 0.45 |
| ever_brfed | rs10995251 | S2 | 11359 | 8 | 1.08 | [0.90-1.30] | 0.41 | 1.06 | [0.84-1.35] | 0.63 | 0.25 |
| ever_brfed | rs10995251 | S3 | 10687 | 8 | 1.10 | [0.90-1.34] | 0.36 | 1.09 | [0.86-1.38] | 0.48 | 0.30 |
| ever_brfed | rs112111458 | I1 | 14702 | 9 | 0.97 | [0.77-1.22] | 0.78 | 0.97 | [0.77-1.22] | 0.78 | 0.53 |
| ever_brfed | rs112111458 | I2 | 2590 | 9 | 1.04 | [0.84-1.28] | 0.74 | 1.11 | [0.84-1.36] | 0.46 | 0.21 |
| ever_brfed | rs112111458 | I3 | 12112 | 9 | 1.01 | [0.94-1.10] | 0.74 | 1.02 | [0.93-1.11] | 0.63 | 0.29 |
| ever_brfed | rs112111458 | S1 | 14022 | 9 | 0.98 | [0.77-1.25] | 0.87 | 0.98 | [0.77-1.25] | 0.87 | 0.90 |
| ever_brfed | rs112111458 | S2 | 11890 | 9 | 0.91 | [0.71-1.16] | 0.44 | 0.91 | [0.71-1.16] | 0.44 | 0.52 |
| ever_brfed | rs112111458 | S3 | 11232 | 9 | 0.94 | [0.72-1.21] | 0.62 | 0.94 | [0.72-1.21] | 0.62 | 0.77 |
| ever_brfed | rs11657987 | I1 | 14684 | 9 | 1.01 | [0.86-1.18] | 0.93 | 1.00 | [0.84-1.20] | 0.99 | 0.35 |
| ever_brfed | rs11657987 | I2 | 2586 | 9 | 1.05 | [0.90-1.22] | 0.54 | 1.05 | [0.87-1.27] | 0.6 | 0.20 |
| ever_brfed | rs11657987 | I3 | 12098 | 9 | 1.05 | [0.99-1.11] | 7.80E-02 | 1.04 | [0.96-1.14] | 0.31 | 0.08 |
| ever_brfed | rs11657987 | S1 | 14020 | 9 | 0.97 | [0.82-1.15] | 0.72 | 0.96 | [0.78-1.18] | 0.67 | 0.32 |
| ever_brfed | rs11657987 | S2 | 11872 | 9 | 1.02 | [0.86-1.21] | 0.83 | 1.01 | [0.83-1.25] | 0.89 | 0.30 |
| ever_brfed | rs11657987 | S3 | 11230 | 9 | 0.98 | [0.81-1.18] | 0.8 | 0.97 | [0.77-1.23] | 0.76 | 0.26 |
| ever_brfed | rs12153855 | I1 | 14176 | 8 | 0.96 | [0.73-1.27] | 0.79 | 0.96 | [0.73-1.27] | 0.79 | 0.79 |
| ever_brfed | rs12153855 | I2 | 2512 | 8 | 1.13 | [0.88-1.45] | 0.33 | 1.13 | [0.88-1.45] | 0.33 | 0.99 |
| ever_brfed | rs12153855 | I3 | 11664 | 8 | 1.09 | [0.99-1.20] | 7.60E-02 | 1.09 | [0.99-1.20] | 7.60E-02 | 0.53 |
| ever_brfed | rs12153855 | S1 | 13477 | 8 | 0.95 | [0.70-1.27] | 0.71 | 0.95 | [0.70-1.27] | 0.71 | 0.65 |
| ever_brfed | rs12153855 | S2 | 11364 | 8 | 0.95 | [0.71-1.28] | 0.75 | 0.95 | [0.71-1.28] | 0.75 | 0.91 |
| ever_brfed | rs12153855 | S3 | 10687 | 8 | 0.95 | [0.68-1.31] | 0.74 | 0.95 | [0.68-1.31] | 0.74 | 0.78 |
| ever_brfed | rs12295535 | I1 | 14723 | 9 | 1.09 | [0.66-1.79] | 0.74 | 1.09 | [0.66-1.79] | 0.74 | 0.85 |
| ever_brfed | rs12295535 | I2 | 2594 | 9 | 1.05 | [0.67-1.67] | 0.82 | 1.05 | [0.67-1.67] | 0.82 | 0.71 |
| ever_brfed | rs12295535 | I3 | 12129 | 9 | 1.14 | [0.96-1.35] | 0.13 | 1.14 | [0.96-1.35] | 0.13 | 0.70 |
| ever_brfed | rs12295535 | S1 | 14024 | 9 | 1.07 | [0.63-1.80] | 0.8 | 1.07 | [0.63-1.80] | 0.8 | 0.85 |
| ever_brfed | rs12295535 | S2 | 11911 | 9 | 0.98 | [0.58-1.65] | 0.93 | 0.98 | [0.58-1.65] | 0.93 | 0.72 |
| ever_brfed | rs12295535 | S3 | 11234 | 9 | 0.91 | [0.52-1.59] | 0.75 | 0.91 | [0.52-1.59] | 0.75 | 0.75 |
| ever_brfed | rs13015714 | I1 | 14174 | 8 | 0.93 | [0.77-1.12] | 0.45 | 0.93 | [0.77-1.12] | 0.45 | 0.82 |
| ever_brfed | rs13015714 | I2 | 2512 | 8 | 1.15 | [0.97-1.37] | 0.12 | 1.15 | [0.97-1.37] | 0.12 | 0.88 |
| ever_brfed | rs13015714 | I3 | 11662 | 8 | 1.10 | [1.03-1.18] | 3.90E-03 | 1.10 | [1.02-1.19] | 1.80E-02 | 0.25 |
| ever_brfed | rs13015714 | S1 | 13475 | 8 | 0.86 | [0.70-1.06] | 0.15 | 0.86 | [0.70-1.06] | 0.15 | 0.83 |
| ever_brfed | rs13015714 | S2 | 11362 | 8 | 0.94 | [0.76-1.15] | 0.53 | 0.94 | [0.76-1.15] | 0.53 | 0.80 |
| ever_brfed | rs13015714 | S3 | 10685 | 8 | 0.86 | [0.69-1.07] | 0.19 | 0.86 | [0.69-1.07] | 0.19 | 0.86 |
| ever_brfed | rs16948048 | I1 | 14174 | 8 | 0.98 | [0.83-1.16] | 0.8 | 0.97 | [0.81-1.18] | 0.77 | 0.36 |
| ever_brfed | rs16948048 | I2 | 2511 | 8 | 1.04 | [0.89-1.22] | 0.6 | 1.05 | [0.88-1.25] | 0.58 | 0.31 |
| ever_brfed | rs16948048 | I3 | 11663 | 8 | 1.02 | [0.96-1.08] | 0.5 | 1.02 | [0.96-1.08] | 0.5 | 0.53 |
| ever_brfed | rs16948048 | S1 | 13477 | 8 | 0.96 | [0.80-1.15] | 0.69 | 0.94 | [0.74-1.23] | 0.64 | 0.20 |
| ever_brfed | rs16948048 | S2 | 11362 | 8 | 0.98 | [0.82-1.18] | 0.83 | 0.97 | [0.77-1.23] | 0.78 | 0.23 |
| ever_brfed | rs16948048 | S3 | 10687 | 8 | 0.97 | [0.80-1.18] | 0.79 | 0.96 | [0.73-1.28] | 0.76 | 0.15 |
| ever_brfed | rs17389644 | I1 | 14163 | 8 | 1.04 | [0.85-1.27] | 0.72 | 1.04 | [0.78-1.38] | 0.81 | 0.12 |
| ever_brfed | rs17389644 | I2 | 2511 | 8 | 1.00 | [0.83-1.21] | 0.97 | 0.99 | [0.80-1.24] | 0.95 | 0.32 |
| ever_brfed | rs17389644 | I3 | 11652 | 8 | 1.05 | [0.98-1.12] | 0.17 | 1.01 | [0.90-1.19] | 0.85 | 0.01 |
| ever_brfed | rs17389644 | S1 | 13477 | 8 | 0.96 | [0.78-1.19] | 0.73 | 0.96 | [0.71-1.30] | 0.77 | 0.16 |
| ever_brfed | rs17389644 | S2 | 11351 | 8 | 1.01 | [0.81-1.26] | 0.92 | 1.01 | [0.73-1.41] | 0.94 | 0.07 |
| ever_brfed | rs17389644 | S3 | 10687 | 8 | 0.92 | [0.73-1.16] | 0.48 | 0.92 | [0.65-1.29] | 0.62 | 0.12 |
| ever_brfed | rs17881320 | I1 | 14704 | 9 | 0.85 | [0.63-1.14] | 0.29 | 0.85 | [0.63-1.14] | 0.29 | 0.88 |
| ever_brfed | rs17881320 | I2 | 2587 | 9 | 1.29 | [0.99-1.69] | 0.06 | 1.29 | [0.99-1.69] | 0.06 | 0.96 |
| ever_brfed | rs17881320 | I3 | 12117 | 9 | 1.09 | [0.98-1.20] | 0.11 | 1.09 | [0.95-1.26] | 0.23 | 0.13 |
| ever_brfed | rs17881320 | S1 | 14022 | 9 | 0.82 | [0.59-1.13] | 0.22 | 0.82 | [0.59-1.13] | 0.22 | 0.84 |
| ever_brfed | rs17881320 | S2 | 11892 | 9 | 0.94 | [0.68-1.29] | 0.7 | 0.94 | [0.68-1.29] | 0.7 | 0.96 |
| ever_brfed | rs17881320 | S3 | 11232 | 9 | 0.91 | [0.64-1.29] | 0.58 | 0.91 | [0.64-1.29] | 0.58 | 0.92 |
| ever_brfed | rs2041733 | I1 | 14175 | 8 | 0.86 | [0.73-1.02] | 7.50E-02 | 0.88 | [0.73-1.04] | 0.17 | 0.32 |
| ever_brfed | rs2041733 | I2 | 2512 | 8 | 1.21 | [1.04-1.40] | 1.40E-02 | 1.21 | [1.04-1.40] | 1.40E-02 | 0.58 |
| ever_brfed | rs2041733 | I3 | 11663 | 8 | 1.04 | [0.98-1.10] | 0.22 | 1.04 | [0.98-1.10] | 0.22 | 0.58 |
| ever_brfed | rs2041733 | S1 | 13476 | 8 | 0.81 | [0.68-0.97] | 0.02 | 0.81 | [0.68-0.97] | 0.02 | 0.63 |
| ever_brfed | rs2041733 | S2 | 11363 | 8 | 0.86 | [0.73-1.03] | 0.1 | 0.88 | [0.72-1.05] | 0.2 | 0.32 |
| ever_brfed | rs2041733 | S3 | 10686 | 8 | 0.82 | [0.67-0.99] | 3.50E-02 | 0.82 | [0.67-0.99] | 3.50E-02 | 0.48 |
| ever_brfed | rs2143950 | I1 | 14710 | 9 | 1.06 | [0.86-1.31] | 0.59 | 1.06 | [0.86-1.31] | 0.59 | 0.98 |
| ever_brfed | rs2143950 | I2 | 2591 | 9 | 1.01 | [0.83-1.22] | 0.94 | 1.01 | [0.83-1.22] | 0.94 | 0.96 |
| ever_brfed | rs2143950 | I3 | 12119 | 9 | 1.07 | [1.00-1.15] | 6.30E-02 | 1.07 | [1.00-1.15] | 6.30E-02 | 0.73 |
| ever_brfed | rs2143950 | S1 | 14015 | 9 | 1.02 | [0.81-1.28] | 0.87 | 1.02 | [0.81-1.28] | 0.87 | 0.99 |
| ever_brfed | rs2143950 | S2 | 11898 | 9 | 1.06 | [0.85-1.34] | 0.6 | 1.06 | [0.85-1.34] | 0.6 | 0.97 |
| ever_brfed | rs2143950 | S3 | 11225 | 9 | 1.01 | [0.79-1.29] | 0.94 | 1.01 | [0.79-1.29] | 0.94 | 0.99 |
| ever_brfed | rs2164983 | I1 | 14176 | 8 | 1.05 | [0.84-1.30] | 0.68 | 1.05 | [0.84-1.30] | 0.68 | 0.47 |
| ever_brfed | rs2164983 | I2 | 2512 | 8 | 1.11 | [0.91-1.36] | 0.3 | 1.08 | [0.83-1.46] | 0.56 | 0.19 |
| ever_brfed | rs2164983 | I3 | 11664 | 8 | 1.14 | [1.06-1.24] | 8.00E-04 | 1.14 | [1.06-1.24] | 8.00E-04 | 0.49 |
| ever_brfed | rs2164983 | S1 | 13477 | 8 | 0.98 | [0.77-1.25] | 0.89 | 0.99 | [0.77-1.26] | 0.92 | 0.40 |
| ever_brfed | rs2164983 | S2 | 11364 | 8 | 1.03 | [0.81-1.30] | 0.83 | 1.03 | [0.81-1.30] | 0.83 | 0.43 |
| ever_brfed | rs2164983 | S3 | 10687 | 8 | 0.96 | [0.75-1.24] | 0.77 | 0.97 | [0.72-1.29] | 0.82 | 0.31 |
| ever_brfed | rs2227483 | I1 | 14712 | 9 | 0.94 | [0.80-1.11] | 0.48 | 0.94 | [0.80-1.11] | 0.48 | 0.61 |
| ever_brfed | rs2227483 | I2 | 2593 | 9 | 1.13 | [0.97-1.31] | 0.11 | 1.13 | [0.97-1.32] | 0.13 | 0.41 |
| ever_brfed | rs2227483 | I3 | 12119 | 9 | 1.06 | [1.00-1.12] | 3.40E-02 | 1.06 | [1.00-1.12] | 3.40E-02 | 0.68 |
| ever_brfed | rs2227483 | S1 | 14017 | 9 | 1.02 | [0.85-1.21] | 0.85 | 1.02 | [0.85-1.21] | 0.85 | 0.85 |
| ever_brfed | rs2227483 | S2 | 11900 | 9 | 0.98 | [0.82-1.16] | 0.78 | 0.98 | [0.82-1.16] | 0.78 | 0.44 |
| ever_brfed | rs2227483 | S3 | 11227 | 9 | 1.06 | [0.88-1.28] | 0.51 | 1.06 | [0.88-1.28] | 0.51 | 0.71 |
| ever_brfed | rs2228145 | I1 | 14176 | 8 | 0.96 | [0.81-1.13] | 0.6 | 0.96 | [0.81-1.13] | 0.6 | 0.83 |
| ever_brfed | rs2228145 | I2 | 2512 | 8 | 1.10 | [0.94-1.28] | 0.24 | 1.10 | [0.94-1.28] | 0.24 | 0.77 |
| ever_brfed | rs2228145 | I3 | 11664 | 8 | 1.04 | [0.98-1.10] | 0.18 | 1.04 | [0.98-1.10] | 0.18 | 0.58 |
| ever_brfed | rs2228145 | S1 | 13477 | 8 | 0.99 | [0.83-1.19] | 0.93 | 0.99 | [0.83-1.19] | 0.93 | 0.81 |
| ever_brfed | rs2228145 | S2 | 11364 | 8 | 0.97 | [0.81-1.15] | 0.7 | 0.97 | [0.81-1.15] | 0.7 | 0.75 |
| ever_brfed | rs2228145 | S3 | 10687 | 8 | 1.01 | [0.83-1.22] | 0.94 | 1.01 | [0.83-1.22] | 0.94 | 0.72 |
| ever_brfed | rs2897442 | I1 | 12876 | 7 | 1.05 | [0.87-1.28] | 0.62 | 1.05 | [0.79-1.39] | 0.76 | 0.13 |

| | | | | | | | | | | | |
|---|---|---|---|---|---|---|---|---|---|---|---|
| ever_brfed | rs2897442 | I2 | 2395 | 7 | 1.07 | [0.89-1.28] | 0.47 | 1.07 | [0.82-1.39] | 0.61 | 0.12 |
| ever_brfed | rs2897442 | I3 | 10481 | 7 | 1.10 | [1.03-1.18] | 5.90E-03 | 1.11 | [0.97-1.25] | 0.12 | 0.01 |
| ever_brfed | rs2897442 | S1 | 12178 | 7 | 1.04 | [0.84-1.28] | 0.73 | 1.03 | [0.76-1.41] | 0.86 | 0.15 |
| ever_brfed | rs2897442 | S2 | 10064 | 7 | 1.03 | [0.83-1.27] | 0.8 | 1.02 | [0.75-1.40] | 0.88 | 0.12 |
| ever_brfed | rs2897442 | S3 | 9388 | 7 | 1.02 | [0.81-1.28] | 0.86 | 1.01 | [0.72-1.43] | 0.94 | 0.12 |
| ever_brfed | rs479844 | I1 | 12877 | 7 | 1.11 | [0.94-1.32] | 0.21 | 1.11 | [0.94-1.32] | 0.21 | 0.86 |
| ever_brfed | rs479844 | I2 | 2396 | 7 | 1.04 | [0.89-1.22] | 0.58 | 1.04 | [0.89-1.22] | 0.58 | 0.42 |
| ever_brfed | rs479844 | I3 | 10481 | 7 | 1.14 | [1.07-1.21] | 4.20E-05 | 1.13 | [1.06-1.21] | 9.10E-05 | 0.40 |
| ever_brfed | rs479844 | S1 | 12178 | 7 | 1.10 | [0.91-1.32] | 0.32 | 1.10 | [0.91-1.32] | 0.32 | 0.71 |
| ever_brfed | rs479844 | S2 | 10065 | 7 | 1.10 | [0.92-1.33] | 0.31 | 1.10 | [0.92-1.33] | 0.31 | 0.71 |
| ever_brfed | rs479844 | S3 | 9388 | 7 | 1.10 | [0.90-1.34] | 0.37 | 1.10 | [0.90-1.34] | 0.37 | 0.52 |
| ever_brfed | rs6010620 | I1 | 14176 | 8 | 1.09 | [0.89-1.32] | 0.41 | 1.09 | [0.89-1.32] | 0.41 | 0.58 |
| ever_brfed | rs6010620 | I2 | 2512 | 8 | 1.03 | [0.86-1.23] | 0.76 | 1.03 | [0.86-1.23] | 0.76 | 0.76 |
| ever_brfed | rs6010620 | I3 | 11664 | 8 | 1.12 | [1.05-1.20] | 6.00E-04 | 1.12 | [1.05-1.20] | 6.00E-04 | 0.81 |
| ever_brfed | rs6010620 | S1 | 13477 | 8 | 1.10 | [0.89-1.35] | 0.38 | 1.10 | [0.89-1.35] | 0.38 | 0.52 |
| ever_brfed | rs6010620 | S2 | 11364 | 8 | 1.09 | [0.89-1.35] | 0.39 | 1.09 | [0.89-1.35] | 0.39 | 0.60 |
| ever_brfed | rs6010620 | S3 | 10687 | 8 | 1.11 | [0.89-1.39] | 0.35 | 1.11 | [0.89-1.39] | 0.35 | 0.48 |
| ever_brfed | rs6473227 | I1 | 14723 | 9 | 1.09 | [0.92-1.28] | 0.33 | 1.05 | [0.78-1.47] | 0.74 | 0.01 |
| ever_brfed | rs6473227 | I2 | 2594 | 9 | 1.01 | [0.87-1.18] | 0.86 | 1.01 | [0.80-1.28] | 0.94 | 0.08 |
| ever_brfed | rs6473227 | I3 | 12129 | 9 | 1.09 | [1.03-1.16] | 1.60E-03 | 1.09 | [1.02-1.17] | 1.50E-02 | 0.28 |
| ever_brfed | rs6473227 | S1 | 14024 | 9 | 1.20 | [1.00-1.43] | 4.80E-02 | 1.17 | [0.86-1.63] | 0.31 | 0.03 |
| ever_brfed | rs6473227 | S2 | 11911 | 9 | 1.11 | [0.93-1.33] | 0.23 | 1.14 | [0.88-1.44] | 0.31 | 0.11 |
| ever_brfed | rs6473227 | S3 | 11234 | 9 | 1.23 | [1.02-1.50] | 3.30E-02 | 1.25 | [1.01-1.53] | 4.20E-02 | 0.35 |
| ever_brfed | rs7127307 | I1 | 14722 | 9 | 1.13 | [0.96-1.33] | 0.13 | 1.13 | [0.94-1.37] | 0.19 | 0.33 |
| ever_brfed | rs7127307 | I2 | 2595 | 9 | 0.94 | [0.81-1.09] | 0.41 | 0.94 | [0.81-1.09] | 0.41 | 0.57 |
| ever_brfed | rs7127307 | I3 | 12127 | 9 | 1.06 | [1.00-1.12] | 4.60E-02 | 1.06 | [1.00-1.12] | 4.60E-02 | 0.53 |
| ever_brfed | rs7127307 | S1 | 14023 | 9 | 1.12 | [0.94-1.33] | 0.2 | 1.10 | [0.86-1.44] | 0.45 | 0.15 |
| ever_brfed | rs7127307 | S2 | 11910 | 9 | 1.15 | [0.97-1.38] | 0.11 | 1.15 | [0.97-1.38] | 0.11 | 0.56 |
| ever_brfed | rs7127307 | S3 | 11233 | 9 | 1.14 | [0.95-1.38] | 0.17 | 1.15 | [0.94-1.40] | 0.17 | 0.39 |
| ever_brfed | rs7146581 | I1 | 14715 | 9 | 1.01 | [0.83-1.22] | 0.95 | 0.99 | [0.80-1.25] | 0.95 | 0.33 |
| ever_brfed | rs7146581 | I2 | 2592 | 9 | 1.01 | [0.85-1.21] | 0.89 | 1.02 | [0.83-1.24] | 0.85 | 0.32 |
| ever_brfed | rs7146581 | I3 | 12123 | 9 | 1.01 | [0.95-1.08] | 0.69 | 1.01 | [0.95-1.08] | 0.69 | 0.52 |
| ever_brfed | rs7146581 | S1 | 14018 | 9 | 1.00 | [0.81-1.23] | 0.98 | 0.96 | [0.73-1.31] | 0.75 | 0.20 |
| ever_brfed | rs7146581 | S2 | 11903 | 9 | 0.99 | [0.81-1.22] | 0.94 | 0.99 | [0.80-1.22] | 0.94 | 0.42 |
| ever_brfed | rs7146581 | S3 | 11228 | 9 | 0.98 | [0.79-1.23] | 0.88 | 0.98 | [0.76-1.27] | 0.88 | 0.33 |
| ever_brfed | rs7927894 | I1 | 14135 | 8 | 0.90 | [0.76-1.06] | 0.22 | 0.90 | [0.76-1.06] | 0.22 | 0.97 |
| ever_brfed | rs7927894 | I2 | 2501 | 8 | 1.12 | [0.96-1.31] | 0.14 | 1.12 | [0.96-1.31] | 0.14 | 0.86 |
| ever_brfed | rs7927894 | I3 | 11634 | 8 | 1.00 | [0.95-1.06] | 0.92 | 1.00 | [0.93-1.07] | 0.91 | 0.32 |
| ever_brfed | rs7927894 | S1 | 13477 | 8 | 0.93 | [0.78-1.11] | 0.41 | 0.93 | [0.78-1.11] | 0.41 | 0.97 |
| ever_brfed | rs7927894 | S2 | 11323 | 8 | 0.87 | [0.73-1.04] | 0.14 | 0.87 | [0.73-1.04] | 0.14 | 0.96 |
| ever_brfed | rs7927894 | S3 | 10687 | 8 | 0.90 | [0.74-1.09] | 0.27 | 0.90 | [0.74-1.09] | 0.27 | 0.96 |
| ever_brfed | | M1 | 16894 | 9 | 0.99 | [0.89-1.09] | 0.8 | 0.99 | [0.86-1.14] | 0.9 | 0.13 |
| ever_brfed | | M2 | 16163 | 9 | 0.98 | [0.88-1.10] | 0.74 | 0.98 | [0.83-1.16] | 0.81 | 0.10 |
| ever_brfed | | M3 | 15121 | 9 | 1.00 | [0.90-1.11] | 0.98 | 1.01 | [0.86-1.17] | 0.89 | 0.06 |
| ever_brfed | | M4 | 14425 | 9 | 1.01 | [0.90-1.13] | 0.93 | 1.01 | [0.84-1.21] | 0.88 | 0.04 |
| excl_brfed | flg | I1 | 12097 | 8 | 1.15 | [0.86-1.54] | 0.34 | 1.15 | [0.86-1.54] | 0.34 | 0.82 |
| excl_brfed | flg | I2 | 4176 | 8 | 1.91 | [1.54-2.35] | 2.10E-09 | 1.91 | [1.54-2.35] | 2.10E-09 | 0.51 |
| excl_brfed | flg | I3 | 7921 | 8 | 1.84 | [1.58-2.15] | 5.30E-15 | 1.78 | [1.46-2.25] | 1.10E-08 | 0.28 |
| excl_brfed | flg | S1 | 11449 | 8 | 1.13 | [0.83-1.54] | 0.45 | 1.13 | [0.83-1.54] | 0.45 | 0.78 |
| excl_brfed | flg | S2 | 10728 | 8 | 1.13 | [0.83-1.55] | 0.43 | 1.13 | [0.83-1.55] | 0.43 | 0.81 |
| excl_brfed | flg | S3 | 10115 | 8 | 1.06 | [0.76-1.49] | 0.72 | 1.06 | [0.76-1.49] | 0.72 | 0.79 |
| excl_brfed | rs10214237 | I1 | 11510 | 8 | 1.13 | [0.98-1.32] | 9.20E-02 | 1.13 | [0.90-1.42] | 0.29 | 0.12 |
| excl_brfed | rs10214237 | I2 | 3334 | 8 | 1.03 | [0.92-1.16] | 0.58 | 1.02 | [0.86-1.22] | 0.86 | 0.17 |
| excl_brfed | rs10214237 | I3 | 8176 | 8 | 1.15 | [1.07-1.24] | 2.90E-04 | 1.15 | [1.07-1.24] | 2.90E-04 | 0.46 |
| excl_brfed | rs10214237 | S1 | 10188 | 8 | 1.11 | [0.95-1.31] | 0.18 | 1.11 | [0.95-1.31] | 0.18 | 0.66 |
| excl_brfed | rs10214237 | S2 | 10309 | 8 | 1.11 | [0.95-1.31] | 0.18 | 1.14 | [0.88-1.44] | 0.31 | 0.07 |
| excl_brfed | rs10214237 | S3 | 9009 | 8 | 1.09 | [0.91-1.29] | 0.35 | 1.09 | [0.91-1.29] | 0.35 | 0.70 |
| excl_brfed | rs1057258 | I1 | 11493 | 8 | 1.10 | [0.92-1.31] | 0.3 | 1.10 | [0.92-1.31] | 0.3 | 0.55 |
| excl_brfed | rs1057258 | I2 | 3326 | 8 | 1.00 | [0.88-1.15] | 0.98 | 1.00 | [0.88-1.15] | 0.98 | 0.62 |
| excl_brfed | rs1057258 | I3 | 8167 | 8 | 1.05 | [0.96-1.14] | 0.31 | 1.05 | [0.96-1.14] | 0.31 | 0.76 |
| excl_brfed | rs1057258 | S1 | 10188 | 8 | 1.06 | [0.88-1.28] | 0.56 | 1.06 | [0.88-1.28] | 0.56 | 0.41 |
| excl_brfed | rs1057258 | S2 | 10292 | 8 | 1.09 | [0.90-1.31] | 0.37 | 1.09 | [0.90-1.31] | 0.37 | 0.76 |
| excl_brfed | rs1057258 | S3 | 9009 | 8 | 1.05 | [0.86-1.29] | 0.63 | 1.05 | [0.86-1.29] | 0.63 | 0.61 |
| excl_brfed | rs10995251 | I1 | 11508 | 8 | 1.06 | [0.93-1.22] | 0.4 | 1.06 | [0.93-1.22] | 0.4 | 0.44 |
| excl_brfed | rs10995251 | I2 | 3333 | 8 | 0.97 | [0.87-1.08] | 0.59 | 1.00 | [0.85-1.14] | 0.98 | 0.15 |
| excl_brfed | rs10995251 | I3 | 8175 | 8 | 1.09 | [1.02-1.16] | 1.60E-02 | 1.09 | [1.01-1.18] | 2.90E-02 | 0.29 |
| excl_brfed | rs10995251 | S1 | 10188 | 8 | 1.01 | [0.87-1.17] | 0.89 | 1.01 | [0.87-1.17] | 0.89 | 0.41 |
| excl_brfed | rs10995251 | S2 | 10307 | 8 | 1.05 | [0.90-1.21] | 0.55 | 1.03 | [0.87-1.23] | 0.73 | 0.35 |
| excl_brfed | rs10995251 | S3 | 9009 | 8 | 1.00 | [0.85-1.17] | 0.96 | 0.96 | [0.79-1.21] | 0.71 | 0.26 |
| excl_brfed | rs112111458 | I1 | 10278 | 6 | 1.02 | [0.84-1.24] | 0.83 | 1.02 | [0.80-1.30] | 0.86 | 0.27 |
| excl_brfed | rs112111458 | I2 | 3203 | 6 | 0.96 | [0.83-1.10] | 0.54 | 1.05 | [0.81-1.24] | 0.74 | 0.06 |
| excl_brfed | rs112111458 | I3 | 7075 | 6 | 1.04 | [0.94-1.15] | 0.45 | 1.04 | [0.94-1.15] | 0.45 | 0.43 |
| excl_brfed | rs112111458 | S1 | 9605 | 6 | 1.02 | [0.83-1.25] | 0.88 | 1.03 | [0.82-1.28] | 0.79 | 0.33 |
| excl_brfed | rs112111458 | S2 | 9660 | 6 | 1.02 | [0.83-1.25] | 0.85 | 1.02 | [0.78-1.33] | 0.89 | 0.21 |
| excl_brfed | rs112111458 | S3 | 9009 | 6 | 1.03 | [0.83-1.27] | 0.82 | 1.05 | [0.80-1.34] | 0.72 | 0.26 |
| excl_brfed | rs11657987 | I1 | 11478 | 8 | 0.97 | [0.85-1.11] | 0.67 | 0.97 | [0.85-1.11] | 0.67 | 0.49 |
| excl_brfed | rs11657987 | I2 | 3321 | 8 | 1.05 | [0.95-1.17] | 0.32 | 1.05 | [0.95-1.17] | 0.32 | 0.73 |
| excl_brfed | rs11657987 | I3 | 8157 | 8 | 1.03 | [0.96-1.09] | 0.45 | 1.02 | [0.95-1.10] | 0.61 | 0.35 |
| excl_brfed | rs11657987 | S1 | 10188 | 8 | 0.94 | [0.81-1.08] | 0.36 | 0.94 | [0.79-1.10] | 0.43 | 0.34 |
| excl_brfed | rs11657987 | S2 | 10277 | 8 | 0.94 | [0.82-1.08] | 0.39 | 0.96 | [0.81-1.12] | 0.63 | 0.29 |
| excl_brfed | rs11657987 | S3 | 9009 | 8 | 0.90 | [0.77-1.05] | 0.18 | 0.92 | [0.75-1.11] | 0.41 | 0.21 |
| excl_brfed | rs12153855 | I1 | 10297 | 6 | 1.16 | [0.93-1.44] | 0.2 | 1.16 | [0.93-1.44] | 0.2 | 0.60 |
| excl_brfed | rs12153855 | I2 | 3208 | 6 | 1.03 | [0.87-1.21] | 0.76 | 1.03 | [0.87-1.21] | 0.76 | 0.80 |
| excl_brfed | rs12153855 | I3 | 7089 | 6 | 1.15 | [1.02-1.29] | 1.90E-02 | 1.15 | [1.02-1.29] | 1.90E-02 | 0.62 |
| excl_brfed | rs12153855 | S1 | 9605 | 6 | 1.20 | [0.94-1.52] | 0.14 | 1.17 | [0.90-1.57] | 0.25 | 0.34 |
| excl_brfed | rs12153855 | S2 | 9679 | 6 | 1.11 | [0.88-1.40] | 0.39 | 1.11 | [0.88-1.40] | 0.39 | 0.63 |
| excl_brfed | rs12153855 | S3 | 9009 | 6 | 1.15 | [0.89-1.48] | 0.28 | 1.14 | [0.88-1.50] | 0.32 | 0.38 |
| excl_brfed | rs12295535 | I1 | 10930 | 7 | 1.22 | [0.80-1.86] | 0.35 | 1.22 | [0.80-1.86] | 0.35 | 0.93 |
| excl_brfed | rs12295535 | I2 | 3278 | 7 | 1.02 | [0.75-1.38] | 0.9 | 1.02 | [0.75-1.38] | 0.9 | 0.90 |
| excl_brfed | rs12295535 | I3 | 7652 | 7 | 1.27 | [1.03-1.56] | 2.70E-02 | 1.29 | [0.97-1.68] | 0.08 | 0.17 |
| excl_brfed | rs12295535 | S1 | 9605 | 7 | 1.21 | [0.78-1.89] | 0.4 | 1.21 | [0.78-1.89] | 0.4 | 0.70 |

| | | | | | | | | | | | |
|---|---|---|---|---|---|---|---|---|---|---|---|
| excl_brfed | rs12295535 | S2 | 10312 | 7 | 1.21 | [0.77-1.91] | 0.41 | 1.21 | [0.77-1.91] | 0.41 | 0.84 |
| excl_brfed | rs12295535 | S3 | 9009 | 7 | 1.22 | [0.76-1.97] | 0.42 | 1.22 | [0.76-1.97] | 0.42 | 0.58 |
| excl_brfed | rs13015714 | I1 | 11511 | 8 | 0.92 | [0.78-1.07] | 0.27 | 0.93 | [0.75-1.14] | 0.48 | 0.21 |
| excl_brfed | rs13015714 | I2 | 3334 | 8 | 1.18 | [1.04-1.32] | 8.30E-03 | 1.18 | [0.99-1.40] | 6.30E-02 | 0.21 |
| excl_brfed | rs13015714 | I3 | 8177 | 8 | 1.11 | [1.03-1.20] | 5.20E-03 | 1.11 | [1.03-1.20] | 5.20E-03 | 0.53 |
| excl_brfed | rs13015714 | S1 | 10186 | 8 | 0.89 | [0.75-1.05] | 0.15 | 0.91 | [0.70-1.16] | 0.51 | 0.10 |
| excl_brfed | rs13015714 | S2 | 10310 | 8 | 0.90 | [0.76-1.06] | 0.19 | 0.93 | [0.72-1.16] | 0.6 | 0.09 |
| excl_brfed | rs13015714 | S3 | 9007 | 8 | 0.87 | [0.73-1.04] | 0.12 | 0.93 | [0.67-1.20] | 0.66 | 0.04 |
| excl_brfed | rs16948048 | I1 | 11511 | 8 | 0.96 | [0.84-1.10] | 0.57 | 0.96 | [0.83-1.11] | 0.57 | 0.39 |
| excl_brfed | rs16948048 | I2 | 3333 | 8 | 1.06 | [0.95-1.18] | 0.28 | 1.07 | [0.92-1.23] | 0.41 | 0.20 |
| excl_brfed | rs16948048 | I3 | 8178 | 8 | 1.02 | [0.95-1.09] | 0.6 | 1.02 | [0.95-1.09] | 0.6 | 0.98 |
| excl_brfed | rs16948048 | S1 | 10188 | 8 | 0.96 | [0.83-1.11] | 0.55 | 0.96 | [0.83-1.11] | 0.55 | 0.41 |
| excl_brfed | rs16948048 | S2 | 10310 | 8 | 0.95 | [0.82-1.10] | 0.5 | 0.95 | [0.82-1.10] | 0.5 | 0.43 |
| excl_brfed | rs16948048 | S3 | 9009 | 8 | 0.95 | [0.82-1.11] | 0.54 | 0.95 | [0.82-1.11] | 0.54 | 0.49 |
| excl_brfed | rs17389644 | I1 | 11500 | 8 | 1.05 | [0.90-1.24] | 0.52 | 1.05 | [0.90-1.24] | 0.52 | 0.45 |
| excl_brfed | rs17389644 | I2 | 3332 | 8 | 1.03 | [0.91-1.16] | 0.68 | 0.96 | [0.78-1.26] | 0.7 | 0.09 |
| excl_brfed | rs17389644 | I3 | 8168 | 8 | 1.08 | [1.00-1.17] | 4.80E-02 | 1.08 | [1.00-1.17] | 4.80E-02 | 0.65 |
| excl_brfed | rs17389644 | S1 | 10188 | 8 | 0.99 | [0.83-1.17] | 0.88 | 0.99 | [0.83-1.17] | 0.88 | 0.49 |
| excl_brfed | rs17389644 | S2 | 10299 | 8 | 1.06 | [0.89-1.25] | 0.54 | 1.09 | [0.88-1.31] | 0.43 | 0.27 |
| excl_brfed | rs17389644 | S3 | 9009 | 8 | 0.97 | [0.81-1.17] | 0.75 | 0.98 | [0.79-1.20] | 0.87 | 0.32 |
| excl_brfed | rs17881320 | I1 | 11496 | 8 | 0.91 | [0.71-1.17] | 0.46 | 0.91 | [0.71-1.17] | 0.46 | 0.64 |
| excl_brfed | rs17881320 | I2 | 3326 | 8 | 1.18 | [0.97-1.43] | 9.80E-02 | 1.18 | [0.97-1.43] | 9.80E-02 | 0.89 |
| excl_brfed | rs17881320 | I3 | 8170 | 8 | 1.10 | [0.97-1.24] | 0.14 | 1.10 | [0.92-1.31] | 0.28 | 0.09 |
| excl_brfed | rs17881320 | S1 | 10188 | 8 | 0.85 | [0.65-1.11] | 0.23 | 0.85 | [0.65-1.11] | 0.23 | 0.62 |
| excl_brfed | rs17881320 | S2 | 10295 | 8 | 0.93 | [0.71-1.21] | 0.58 | 0.97 | [0.71-1.26] | 0.83 | 0.34 |
| excl_brfed | rs17881320 | S3 | 9009 | 8 | 0.85 | [0.63-1.13] | 0.27 | 0.89 | [0.61-1.24] | 0.56 | 0.27 |
| excl_brfed | rs2041733 | I1 | 11512 | 8 | 0.97 | [0.85-1.11] | 0.68 | 0.97 | [0.85-1.11] | 0.68 | 0.47 |
| excl_brfed | rs2041733 | I2 | 3334 | 8 | 1.10 | [1.00-1.22] | 5.90E-02 | 1.10 | [1.00-1.22] | 5.90E-02 | 0.69 |
| excl_brfed | rs2041733 | I3 | 8178 | 8 | 1.02 | [0.96-1.09] | 0.49 | 1.02 | [0.95-1.11] | 0.56 | 0.25 |
| excl_brfed | rs2041733 | S1 | 10187 | 8 | 0.92 | [0.80-1.06] | 0.26 | 0.92 | [0.80-1.06] | 0.26 | 0.89 |
| excl_brfed | rs2041733 | S2 | 10311 | 8 | 1.00 | [0.87-1.15] | 0.97 | 1.00 | [0.87-1.15] | 0.97 | 0.46 |
| excl_brfed | rs2041733 | S3 | 9008 | 8 | 0.96 | [0.82-1.11] | 0.55 | 0.96 | [0.82-1.11] | 0.55 | 0.82 |
| excl_brfed | rs2143950 | I1 | 11509 | 8 | 1.12 | [0.95-1.34] | 0.19 | 1.12 | [0.95-1.34] | 0.19 | 0.76 |
| excl_brfed | rs2143950 | I2 | 3333 | 8 | 1.03 | [0.90-1.18] | 0.64 | 1.03 | [0.90-1.18] | 0.64 | 0.67 |
| excl_brfed | rs2143950 | I3 | 8176 | 8 | 1.07 | [0.98-1.16] | 0.14 | 1.06 | [0.95-1.18] | 0.3 | 0.27 |
| excl_brfed | rs2143950 | S1 | 10188 | 8 | 1.12 | [0.93-1.35] | 0.25 | 1.12 | [0.93-1.35] | 0.25 | 0.65 |
| excl_brfed | rs2143950 | S2 | 10308 | 8 | 1.18 | [0.98-1.42] | 8.90E-02 | 1.18 | [0.98-1.42] | 8.90E-02 | 0.69 |
| excl_brfed | rs2143950 | S3 | 9009 | 8 | 1.15 | [0.94-1.41] | 0.16 | 1.15 | [0.94-1.41] | 0.16 | 0.63 |
| excl_brfed | rs2164983 | I1 | 10297 | 6 | 0.97 | [0.81-1.17] | 0.78 | 0.97 | [0.81-1.17] | 0.78 | 0.57 |
| excl_brfed | rs2164983 | I2 | 3208 | 6 | 1.14 | [0.99-1.31] | 6.40E-02 | 1.16 | [0.94-1.41] | 0.18 | 0.16 |
| excl_brfed | rs2164983 | I3 | 7089 | 6 | 1.15 | [1.05-1.27] | 4.40E-03 | 1.15 | [1.05-1.27] | 4.40E-03 | 0.59 |
| excl_brfed | rs2164983 | S1 | 9605 | 6 | 0.93 | [0.76-1.13] | 0.46 | 0.93 | [0.74-1.18] | 0.56 | 0.32 |
| excl_brfed | rs2164983 | S2 | 9679 | 6 | 1.00 | [0.83-1.22] | 0.97 | 1.00 | [0.83-1.22] | 0.97 | 0.49 |
| excl_brfed | rs2164983 | S3 | 9009 | 6 | 0.96 | [0.78-1.18] | 0.71 | 0.95 | [0.73-1.25] | 0.72 | 0.29 |
| excl_brfed | rs2227483 | I1 | 11509 | 8 | 0.97 | [0.85-1.11] | 0.64 | 0.97 | [0.85-1.11] | 0.64 | 0.45 |
| excl_brfed | rs2227483 | I2 | 3334 | 8 | 1.08 | [0.97-1.19] | 0.16 | 1.13 | [0.95-1.28] | 0.17 | 0.09 |
| excl_brfed | rs2227483 | I3 | 8175 | 8 | 1.07 | [1.00-1.14] | 0.05 | 1.06 | [0.99-1.15] | 9.30E-02 | 0.35 |
| excl_brfed | rs2227483 | S1 | 10188 | 8 | 1.05 | [0.91-1.21] | 0.5 | 1.05 | [0.91-1.21] | 0.5 | 0.93 |
| excl_brfed | rs2227483 | S2 | 10308 | 8 | 1.00 | [0.87-1.15] | 0.99 | 0.98 | [0.82-1.19] | 0.8 | 0.28 |
| excl_brfed | rs2227483 | S3 | 9009 | 8 | 1.09 | [0.94-1.27] | 0.26 | 1.09 | [0.94-1.27] | 0.26 | 0.89 |
| excl_brfed | rs2228145 | I1 | 10880 | 7 | 1.06 | [0.93-1.21] | 0.39 | 1.06 | [0.93-1.21] | 0.39 | 0.57 |
| excl_brfed | rs2228145 | I2 | 3264 | 7 | 1.00 | [0.90-1.11] | 0.95 | 1.00 | [0.90-1.11] | 0.95 | 0.55 |
| excl_brfed | rs2228145 | I3 | 7616 | 7 | 1.05 | [0.98-1.13] | 0.14 | 1.05 | [0.96-1.15] | 0.28 | 0.19 |
| excl_brfed | rs2228145 | S1 | 10188 | 7 | 1.06 | [0.92-1.23] | 0.42 | 1.06 | [0.92-1.23] | 0.43 | 0.41 |
| excl_brfed | rs2228145 | S2 | 9679 | 7 | 1.06 | [0.91-1.22] | 0.46 | 1.06 | [0.91-1.22] | 0.46 | 0.47 |
| excl_brfed | rs2228145 | S3 | 9009 | 7 | 1.05 | [0.90-1.23] | 0.52 | 1.04 | [0.87-1.26] | 0.67 | 0.32 |
| excl_brfed | rs2897442 | I1 | 10213 | 7 | 1.07 | [0.92-1.24] | 0.4 | 1.07 | [0.92-1.24] | 0.4 | 0.49 |
| excl_brfed | rs2897442 | I2 | 3208 | 7 | 1.08 | [0.97-1.22] | 0.17 | 1.09 | [0.95-1.25] | 0.23 | 0.32 |
| excl_brfed | rs2897442 | I3 | 7005 | 7 | 1.11 | [1.03-1.21] | 8.10E-03 | 1.13 | [0.98-1.28] | 8.90E-02 | 0.02 |
| excl_brfed | rs2897442 | S1 | 8889 | 7 | 1.09 | [0.93-1.29] | 0.28 | 1.09 | [0.93-1.29] | 0.28 | 0.48 |
| excl_brfed | rs2897442 | S2 | 9012 | 7 | 1.03 | [0.88-1.22] | 0.69 | 1.03 | [0.88-1.22] | 0.69 | 0.51 |
| excl_brfed | rs2897442 | S3 | 7710 | 7 | 1.08 | [0.90-1.28] | 0.4 | 1.08 | [0.90-1.28] | 0.4 | 0.51 |
| excl_brfed | rs479844 | I1 | 10214 | 7 | 1.05 | [0.92-1.21] | 0.45 | 1.05 | [0.92-1.21] | 0.45 | 0.69 |
| excl_brfed | rs479844 | I2 | 3209 | 7 | 1.12 | [1.01-1.24] | 3.50E-02 | 1.12 | [0.99-1.26] | 6.40E-02 | 0.34 |
| excl_brfed | rs479844 | I3 | 7005 | 7 | 1.17 | [1.09-1.25] | 2.50E-05 | 1.17 | [1.09-1.25] | 2.50E-05 | 0.79 |
| excl_brfed | rs479844 | S1 | 8889 | 7 | 1.05 | [0.90-1.21] | 0.54 | 1.05 | [0.90-1.22] | 0.55 | 0.40 |
| excl_brfed | rs479844 | S2 | 9013 | 7 | 1.04 | [0.90-1.21] | 0.57 | 1.04 | [0.90-1.21] | 0.57 | 0.76 |
| excl_brfed | rs479844 | S3 | 7710 | 7 | 1.05 | [0.89-1.23] | 0.56 | 1.05 | [0.89-1.23] | 0.56 | 0.49 |
| excl_brfed | rs6010620 | I1 | 11513 | 8 | 1.05 | [0.89-1.22] | 0.58 | 1.05 | [0.89-1.22] | 0.58 | 0.61 |
| excl_brfed | rs6010620 | I2 | 3334 | 8 | 1.08 | [0.96-1.22] | 0.2 | 1.08 | [0.96-1.22] | 0.2 | 0.58 |
| excl_brfed | rs6010620 | I3 | 8179 | 8 | 1.13 | [1.04-1.22] | 3.40E-03 | 1.13 | [1.04-1.22] | 3.40E-03 | 0.87 |
| excl_brfed | rs6010620 | S1 | 10188 | 8 | 1.03 | [0.87-1.22] | 0.76 | 1.03 | [0.87-1.22] | 0.76 | 0.54 |
| excl_brfed | rs6010620 | S2 | 10312 | 8 | 1.06 | [0.90-1.25] | 0.5 | 1.06 | [0.90-1.25] | 0.5 | 0.44 |
| excl_brfed | rs6010620 | S3 | 9009 | 8 | 1.06 | [0.89-1.27] | 0.51 | 1.03 | [0.83-1.32] | 0.8 | 0.31 |
| excl_brfed | rs6473227 | I1 | 11513 | 8 | 1.06 | [0.93-1.22] | 0.38 | 1.01 | [0.78-1.38] | 0.94 | 0.02 |
| excl_brfed | rs6473227 | I2 | 3334 | 8 | 1.03 | [0.93-1.15] | 0.52 | 1.06 | [0.88-1.25] | 0.57 | 0.06 |
| excl_brfed | rs6473227 | I3 | 8179 | 8 | 1.09 | [1.02-1.17] | 0.01 | 1.08 | [0.99-1.18] | 6.80E-02 | 0.30 |
| excl_brfed | rs6473227 | S1 | 10188 | 8 | 1.11 | [0.96-1.28] | 0.16 | 1.06 | [0.77-1.53] | 0.71 | 0.01 |
| excl_brfed | rs6473227 | S2 | 10312 | 8 | 1.03 | [0.89-1.19] | 0.68 | 0.98 | [0.74-1.37] | 0.92 | 0.01 |
| excl_brfed | rs6473227 | S3 | 9009 | 8 | 1.07 | [0.91-1.24] | 0.43 | 1.03 | [0.72-1.52] | 0.89 | 0.00 |
| excl_brfed | rs7127307 | I1 | 11513 | 8 | 1.03 | [0.90-1.17] | 0.71 | 1.03 | [0.90-1.17] | 0.71 | 0.92 |
| excl_brfed | rs7127307 | I2 | 3334 | 8 | 1.02 | [0.92-1.14] | 0.69 | 1.02 | [0.92-1.14] | 0.69 | 0.80 |
| excl_brfed | rs7127307 | I3 | 8179 | 8 | 1.05 | [0.98-1.12] | 0.19 | 1.05 | [0.98-1.12] | 0.19 | 0.93 |
| excl_brfed | rs7127307 | S1 | 10188 | 8 | 1.00 | [0.87-1.16] | 0.98 | 1.00 | [0.87-1.16] | 0.98 | 0.99 |
| excl_brfed | rs7127307 | S2 | 10312 | 8 | 1.01 | [0.87-1.16] | 0.95 | 1.01 | [0.87-1.16] | 0.95 | 0.75 |
| excl_brfed | rs7127307 | S3 | 9009 | 8 | 0.98 | [0.83-1.14] | 0.76 | 0.98 | [0.83-1.14] | 0.76 | 0.79 |
| excl_brfed | rs7146581 | I1 | 11511 | 8 | 1.05 | [0.90-1.23] | 0.54 | 1.06 | [0.90-1.24] | 0.47 | 0.40 |
| excl_brfed | rs7146581 | I2 | 3333 | 8 | 0.96 | [0.85-1.09] | 0.55 | 0.93 | [0.80-1.13] | 0.35 | 0.27 |
| excl_brfed | rs7146581 | I3 | 8178 | 8 | 1.06 | [0.98-1.15] | 0.14 | 1.06 | [0.98-1.15] | 0.14 | 0.41 |
| excl_brfed | rs7146581 | S1 | 10188 | 8 | 1.01 | [0.85-1.20] | 0.89 | 1.02 | [0.85-1.21] | 0.85 | 0.40 |
| excl_brfed | rs7146581 | S2 | 10310 | 8 | 1.02 | [0.86-1.21] | 0.81 | 1.07 | [0.87-1.25] | 0.53 | 0.29 |

| | | | | | | | | | | | |
|---|---|---|---|---|---|---|---|---|---|---|---|
| excl_brfed | rs7146581 | S3 | 9009 | 8 | 0.99 | [0.83-1.19] | 0.91 | 1.02 | [0.82-1.23] | 0.88 | 0.30 |
| excl_brfed | rs7927894 | I1 | 11472 | 8 | 1.08 | [0.94-1.23] | 0.29 | 1.07 | [0.91-1.26] | 0.42 | 0.34 |
| excl_brfed | rs7927894 | I2 | 3322 | 8 | 0.96 | [0.86-1.07] | 0.44 | 0.96 | [0.86-1.07] | 0.44 | 0.91 |
| excl_brfed | rs7927894 | I3 | 8150 | 8 | 1.05 | [0.98-1.12] | 0.16 | 1.03 | [0.95-1.14] | 0.44 | 0.23 |
| excl_brfed | rs7927894 | S1 | 10188 | 8 | 1.12 | [0.97-1.30] | 0.12 | 1.11 | [0.93-1.35] | 0.25 | 0.29 |
| excl_brfed | rs7927894 | S2 | 10271 | 8 | 1.09 | [0.94-1.26] | 0.26 | 1.07 | [0.89-1.31] | 0.49 | 0.26 |
| excl_brfed | rs7927894 | S3 | 9009 | 8 | 1.12 | [0.96-1.31] | 0.14 | 1.10 | [0.88-1.41] | 0.41 | 0.16 |
| excl_brfed | | M1 | 13789 | 8 | 0.99 | [0.91-1.08] | 0.9 | 1.01 | [0.86-1.17] | 0.87 | 0.02 |
| excl_brfed | | M2 | 13067 | 8 | 0.99 | [0.91-1.09] | 0.88 | 0.99 | [0.85-1.16] | 0.94 | 0.07 |
| excl_brfed | | M3 | 12385 | 8 | 0.98 | [0.89-1.07] | 0.61 | 1.01 | [0.85-1.16] | 0.91 | 0.02 |
| excl_brfed | | M4 | 11698 | 8 | 0.98 | [0.89-1.08] | 0.74 | 1.00 | [0.84-1.16] | 0.97 | 0.05 |
| hdm_1y | flg | I1 | 908 | 2 | 1.05 | [0.90-1.23] | 0.52 | 1.09 | [0.86-1.34] | 0.49 | 0.18 |
| hdm_1y | flg | S1 | 632 | 2 | 1.30 | [0.93-1.82] | 0.13 | 1.30 | [0.93-1.82] | 0.13 | 1.00 |
| hdm_1y | flg | S2 | 908 | 2 | 1.05 | [0.90-1.24] | 0.51 | 1.10 | [0.84-1.37] | 0.49 | 0.15 |
| hdm_1y | flg | S3 | 632 | 2 | 1.30 | [0.94-1.80] | 0.11 | 1.30 | [0.94-1.80] | 0.11 | 1.00 |
| hdm_1y | rs10214237 | I1 | 308 | 1 | 1.01 | [0.96-1.07] | 0.65 | 1.01 | [0.96-1.07] | 0.65 | 1.00 |
| hdm_1y | rs10214237 | S2 | 308 | 1 | 1.01 | [0.96-1.07] | 0.64 | 1.01 | [0.96-1.07] | 0.64 | 1.00 |
| hdm_1y | rs1057258 | I1 | 298 | 1 | 1.03 | [0.97-1.08] | 0.35 | 1.03 | [0.97-1.08] | 0.35 | 1.00 |
| hdm_1y | rs1057258 | S2 | 298 | 1 | 1.03 | [0.98-1.09] | 0.27 | 1.03 | [0.98-1.09] | 0.27 | 1.00 |
| hdm_1y | rs10995251 | I1 | 305 | 1 | 0.97 | [0.93-1.02] | 0.2 | 0.97 | [0.93-1.02] | 0.2 | 1.00 |
| hdm_1y | rs10995251 | S2 | 305 | 1 | 0.97 | [0.93-1.02] | 0.29 | 0.97 | [0.93-1.02] | 0.29 | 1.00 |
| hdm_1y | rs112111458 | I1 | 298 | 1 | 1.05 | [0.96-1.16] | 0.28 | 1.05 | [0.96-1.16] | 0.28 | 1.00 |
| hdm_1y | rs112111458 | S2 | 298 | 1 | 1.05 | [0.96-1.16] | 0.31 | 1.05 | [0.96-1.16] | 0.31 | 1.00 |
| hdm_1y | rs11657987 | I1 | 292 | 1 | 1.01 | [0.95-1.07] | 0.72 | 1.01 | [0.95-1.07] | 0.72 | 1.00 |
| hdm_1y | rs11657987 | S2 | 292 | 1 | 1.01 | [0.95-1.07] | 0.83 | 1.01 | [0.95-1.07] | 0.83 | 1.00 |
| hdm_1y | rs12153855 | I1 | 309 | 1 | 1.06 | [0.96-1.18] | 0.24 | 1.06 | [0.96-1.18] | 0.24 | 1.00 |
| hdm_1y | rs12153855 | S2 | 309 | 1 | 1.07 | [0.95-1.20] | 0.25 | 1.07 | [0.95-1.20] | 0.25 | 1.00 |
| hdm_1y | rs13015714 | I1 | 309 | 1 | 0.94 | [0.88-1.01] | 0.11 | 0.94 | [0.88-1.01] | 0.11 | 1.00 |
| hdm_1y | rs13015714 | S2 | 309 | 1 | 0.94 | [0.88-1.01] | 0.1 | 0.94 | [0.88-1.01] | 0.1 | 1.00 |
| hdm_1y | rs16948048 | I1 | 308 | 1 | 0.97 | [0.92-1.01] | 0.16 | 0.97 | [0.92-1.01] | 0.16 | 1.00 |
| hdm_1y | rs16948048 | S2 | 308 | 1 | 0.97 | [0.92-1.02] | 0.21 | 0.97 | [0.92-1.02] | 0.21 | 1.00 |
| hdm_1y | rs17389644 | I1 | 301 | 1 | 0.99 | [0.90-1.08] | 0.77 | 0.99 | [0.90-1.08] | 0.77 | 1.00 |
| hdm_1y | rs17389644 | S2 | 301 | 1 | 0.98 | [0.90-1.08] | 0.71 | 0.98 | [0.90-1.08] | 0.71 | 1.00 |
| hdm_1y | rs17881320 | I1 | 301 | 1 | 1.02 | [0.96-1.09] | 0.45 | 1.02 | [0.96-1.09] | 0.45 | 1.00 |
| hdm_1y | rs17881320 | S2 | 301 | 1 | 1.01 | [0.95-1.08] | 0.69 | 1.01 | [0.95-1.08] | 0.69 | 1.00 |
| hdm_1y | rs2041733 | I1 | 309 | 1 | 1.02 | [0.99-1.05] | 0.28 | 1.02 | [0.99-1.05] | 0.28 | 1.00 |
| hdm_1y | rs2041733 | S2 | 309 | 1 | 1.02 | [0.99-1.05] | 0.29 | 1.02 | [0.99-1.05] | 0.29 | 1.00 |
| hdm_1y | rs2143950 | I1 | 307 | 1 | 1.08 | [1.01-1.16] | 2.40E-02 | 1.08 | [1.01-1.16] | 2.40E-02 | 1.00 |
| hdm_1y | rs2143950 | S2 | 307 | 1 | 1.09 | [1.01-1.18] | 2.10E-02 | 1.09 | [1.01-1.18] | 2.10E-02 | 1.00 |
| hdm_1y | rs2164983 | I1 | 309 | 1 | 1.00 | [0.95-1.05] | 0.88 | 1.00 | [0.95-1.05] | 0.88 | 1.00 |
| hdm_1y | rs2164983 | S2 | 309 | 1 | 1.00 | [0.95-1.05] | 0.96 | 1.00 | [0.95-1.05] | 0.96 | 1.00 |
| hdm_1y | rs2227483 | I1 | 306 | 1 | 1.02 | [0.96-1.07] | 0.56 | 1.02 | [0.96-1.07] | 0.56 | 1.00 |
| hdm_1y | rs2227483 | S2 | 306 | 1 | 1.02 | [0.96-1.08] | 0.54 | 1.02 | [0.96-1.08] | 0.54 | 1.00 |
| hdm_1y | rs2228145 | I1 | 309 | 1 | 0.97 | [0.93-1.02] | 0.22 | 0.97 | [0.93-1.02] | 0.22 | 1.00 |
| hdm_1y | rs2228145 | S2 | 309 | 1 | 0.98 | [0.93-1.02] | 0.33 | 0.98 | [0.93-1.02] | 0.33 | 1.00 |
| hdm_1y | rs2897442 | I1 | 308 | 1 | 1.01 | [0.98-1.05] | 0.4 | 1.01 | [0.98-1.05] | 0.4 | 1.00 |
| hdm_1y | rs2897442 | S2 | 308 | 1 | 1.02 | [0.98-1.05] | 0.37 | 1.02 | [0.98-1.05] | 0.37 | 1.00 |
| hdm_1y | rs479844 | I1 | 309 | 1 | 0.99 | [0.95-1.03] | 0.59 | 0.99 | [0.95-1.03] | 0.59 | 1.00 |
| hdm_1y | rs479844 | S2 | 309 | 1 | 0.98 | [0.93-1.02] | 0.29 | 0.98 | [0.93-1.02] | 0.29 | 1.00 |
| hdm_1y | rs6010620 | I1 | 309 | 1 | 0.94 | [0.88-1.01] | 8.50E-02 | 0.94 | [0.88-1.01] | 8.50E-02 | 1.00 |
| hdm_1y | rs6010620 | S2 | 309 | 1 | 0.95 | [0.88-1.01] | 0.12 | 0.95 | [0.88-1.01] | 0.12 | 1.00 |
| hdm_1y | rs6473227 | I1 | 309 | 1 | 1.00 | [0.97-1.03] | 0.92 | 1.00 | [0.97-1.03] | 0.92 | 1.00 |
| hdm_1y | rs6473227 | S2 | 309 | 1 | 1.01 | [0.97-1.04] | 0.76 | 1.01 | [0.97-1.04] | 0.76 | 1.00 |
| hdm_1y | rs7127307 | I1 | 309 | 1 | 0.96 | [0.92-1.01] | 0.1 | 0.96 | [0.92-1.01] | 0.1 | 1.00 |
| hdm_1y | rs7127307 | S2 | 309 | 1 | 0.96 | [0.92-1.01] | 9.50E-02 | 0.96 | [0.92-1.01] | 9.50E-02 | 1.00 |
| hdm_1y | rs7146581 | I1 | 309 | 1 | 0.97 | [0.93-1.01] | 0.16 | 0.97 | [0.93-1.01] | 0.16 | 1.00 |
| hdm_1y | rs7146581 | S2 | 309 | 1 | 0.97 | [0.93-1.01] | 0.17 | 0.97 | [0.93-1.01] | 0.17 | 1.00 |
| hdm_1y | rs7927894 | I1 | 291 | 1 | 1.01 | [0.96-1.06] | 0.74 | 1.01 | [0.96-1.06] | 0.74 | 1.00 |
| hdm_1y | rs7927894 | S2 | 291 | 1 | 1.00 | [0.95-1.06] | 0.94 | 1.00 | [0.95-1.06] | 0.94 | 1.00 |
| hdm_1y | | M1 | 1084 | 2 | 1.00 | [0.98-1.02] | 0.77 | 1.00 | [0.98-1.02] | 0.77 | 0.34 |
| hdm_1y | | M2 | 775 | 2 | 1.01 | [0.99-1.04] | 0.33 | 1.01 | [0.99-1.04] | 0.33 | 1.00 |
| hdm_1y | | M3 | 1084 | 2 | 1.01 | [0.99-1.03] | 0.5 | 1.01 | [0.99-1.03] | 0.5 | 0.41 |
| hdm_1y | | M4 | 775 | 2 | 1.02 | [0.99-1.05] | 0.26 | 1.02 | [0.99-1.05] | 0.26 | 1.00 |
| hdm_birth | flg | I1 | 2377 | 5 | 1.00 | [0.99-1.02] | 0.67 | 1.00 | [0.99-1.02] | 0.67 | 0.73 |
| hdm_birth | flg | S1 | 1852 | 5 | 1.02 | [0.96-1.08] | 0.5 | 1.02 | [0.96-1.08] | 0.5 | 0.67 |
| hdm_birth | flg | S2 | 2377 | 5 | 1.00 | [0.99-1.02] | 0.76 | 1.00 | [0.99-1.02] | 0.76 | 0.81 |
| hdm_birth | flg | S3 | 1852 | 5 | 1.02 | [0.96-1.08] | 0.6 | 1.02 | [0.96-1.08] | 0.6 | 0.76 |
| hdm_birth | rs10214237 | I1 | 1834 | 4 | 1.00 | [0.99-1.01] | 0.65 | 0.99 | [0.98-1.01] | 0.45 | 0.18 |
| hdm_birth | rs10214237 | S1 | 1245 | 4 | 0.99 | [0.97-1.00] | 0.14 | 0.99 | [0.96-1.01] | 0.26 | 0.27 |
| hdm_birth | rs10214237 | S2 | 1834 | 4 | 1.00 | [0.99-1.01] | 0.71 | 0.99 | [0.98-1.01] | 0.44 | 0.16 |
| hdm_birth | rs10214237 | S3 | 1245 | 4 | 0.99 | [0.97-1.00] | 0.12 | 0.98 | [0.95-1.02] | 0.27 | 0.22 |
| hdm_birth | rs1057258 | I1 | 1818 | 4 | 1.00 | [0.99-1.01] | 0.59 | 1.00 | [0.99-1.01] | 0.59 | 0.56 |
| hdm_birth | rs1057258 | S1 | 1245 | 4 | 0.98 | [0.96-1.01] | 0.21 | 0.98 | [0.96-1.01] | 0.21 | 0.60 |
| hdm_birth | rs1057258 | S2 | 1818 | 4 | 1.00 | [0.99-1.01] | 0.65 | 1.00 | [0.99-1.01] | 0.65 | 0.46 |
| hdm_birth | rs1057258 | S3 | 1245 | 4 | 0.99 | [0.96-1.01] | 0.28 | 0.99 | [0.96-1.01] | 0.28 | 0.53 |
| hdm_birth | rs10995251 | I1 | 1832 | 4 | 1.00 | [0.99-1.00] | 0.22 | 0.99 | [0.98-1.01] | 0.31 | 0.23 |
| hdm_birth | rs10995251 | S1 | 1245 | 4 | 0.99 | [0.98-1.00] | 0.21 | 0.99 | [0.97-1.02] | 0.41 | 0.14 |
| hdm_birth | rs10995251 | S2 | 1832 | 4 | 1.00 | [0.99-1.00] | 0.28 | 1.00 | [0.99-1.01] | 0.34 | 0.29 |
| hdm_birth | rs10995251 | S3 | 1245 | 4 | 0.99 | [0.98-1.01] | 0.26 | 0.99 | [0.97-1.01] | 0.43 | 0.20 |
| hdm_birth | rs112111458 | I1 | 1820 | 4 | 1.00 | [0.99-1.01] | 0.88 | 0.99 | [0.98-1.02] | 0.53 | 0.14 |
| hdm_birth | rs112111458 | S1 | 1245 | 4 | 0.99 | [0.97-1.00] | 0.14 | 0.98 | [0.96-1.01] | 0.22 | 0.33 |
| hdm_birth | rs112111458 | S2 | 1820 | 4 | 1.00 | [1.00-1.01] | 0.58 | 1.00 | [0.98-1.02] | 0.68 | 0.18 |
| hdm_birth | rs112111458 | S3 | 1245 | 4 | 0.99 | [0.97-1.01] | 0.19 | 0.99 | [0.96-1.01] | 0.26 | 0.34 |
| hdm_birth | rs11657987 | I1 | 1805 | 4 | 1.00 | [0.99-1.01] | 0.44 | 1.00 | [0.99-1.01] | 0.44 | 0.72 |
| hdm_birth | rs11657987 | S1 | 1245 | 4 | 1.01 | [0.99-1.03] | 0.51 | 1.01 | [0.99-1.03] | 0.51 | 0.90 |
| hdm_birth | rs11657987 | S2 | 1805 | 4 | 1.00 | [0.99-1.00] | 0.31 | 1.00 | [0.99-1.00] | 0.31 | 0.71 |
| hdm_birth | rs11657987 | S3 | 1245 | 4 | 1.01 | [0.98-1.03] | 0.61 | 1.01 | [0.98-1.03] | 0.61 | 0.83 |
| hdm_birth | rs12153855 | I1 | 1837 | 4 | 1.00 | [0.99-1.02] | 0.47 | 1.00 | [0.99-1.02] | 0.47 | 0.75 |
| hdm_birth | rs12153855 | S1 | 1245 | 4 | 0.98 | [0.95-1.02] | 0.36 | 0.98 | [0.95-1.02] | 0.36 | 0.87 |
| hdm_birth | rs12153855 | S2 | 1837 | 4 | 1.00 | [0.99-1.02] | 0.45 | 1.00 | [0.99-1.02] | 0.45 | 0.79 |
| hdm_birth | rs12153855 | S3 | 1245 | 4 | 0.99 | [0.95-1.02] | 0.53 | 0.99 | [0.95-1.02] | 0.53 | 0.96 |

| | | | | | | | | | | | |
|---|---|---|---|---|---|---|---|---|---|---|---|
| hdm_birth | rs12295535 | I1 | 1245 | 3 | 1.06 | [0.92-1.22] | 0.44 | 1.06 | [0.92-1.22] | 0.44 | 0.79 |
| hdm_birth | rs12295535 | S1 | 1245 | 3 | 1.06 | [0.91-1.24] | 0.44 | 1.06 | [0.91-1.24] | 0.44 | 0.82 |
| hdm_birth | rs12295535 | S2 | 1245 | 3 | 1.11 | [0.93-1.32] | 0.24 | 1.11 | [0.93-1.32] | 0.24 | 0.54 |
| hdm_birth | rs12295535 | S3 | 1245 | 3 | 1.10 | [0.92-1.31] | 0.29 | 1.10 | [0.92-1.31] | 0.29 | 0.73 |
| hdm_birth | rs13015714 | I1 | 1837 | 4 | 1.00 | [0.99-1.01] | 0.45 | 1.00 | [0.99-1.01] | 0.45 | 0.52 |
| hdm_birth | rs13015714 | S1 | 1245 | 4 | 1.01 | [0.99-1.02] | 0.51 | 1.01 | [0.99-1.02] | 0.51 | 0.63 |
| hdm_birth | rs13015714 | S2 | 1837 | 4 | 1.00 | [0.99-1.00] | 0.32 | 1.00 | [0.99-1.00] | 0.32 | 0.58 |
| hdm_birth | rs13015714 | S3 | 1245 | 4 | 1.00 | [0.99-1.02] | 0.74 | 1.00 | [0.99-1.02] | 0.74 | 0.63 |
| hdm_birth | rs16948048 | I1 | 1835 | 4 | 1.00 | [1.00-1.01] | 0.29 | 1.00 | [1.00-1.01] | 0.29 | 0.54 |
| hdm_birth | rs16948048 | S1 | 1245 | 4 | 1.00 | [0.98-1.02] | 0.96 | 1.00 | [0.98-1.02] | 0.9 | 0.34 |
| hdm_birth | rs16948048 | S2 | 1835 | 4 | 1.00 | [1.00-1.01] | 0.29 | 1.00 | [1.00-1.01] | 0.29 | 0.45 |
| hdm_birth | rs16948048 | S3 | 1245 | 4 | 1.00 | [0.98-1.02] | 0.83 | 1.00 | [0.98-1.02] | 0.83 | 0.37 |
| hdm_birth | rs17389644 | I1 | 1825 | 4 | 1.00 | [0.99-1.01] | 0.63 | 1.00 | [0.99-1.02] | 0.8 | 0.33 |
| hdm_birth | rs17389644 | S1 | 1245 | 4 | 1.00 | [0.97-1.03] | 0.92 | 1.00 | [0.95-1.05] | 0.96 | 0.17 |
| hdm_birth | rs17389644 | S2 | 1825 | 4 | 1.00 | [0.99-1.01] | 0.66 | 1.00 | [0.99-1.02] | 0.82 | 0.31 |
| hdm_birth | rs17389644 | S3 | 1245 | 4 | 1.00 | [0.97-1.03] | 0.95 | 1.00 | [0.95-1.05] | 0.96 | 0.18 |
| hdm_birth | rs17881320 | I1 | 1820 | 4 | 1.00 | [0.99-1.02] | 0.49 | 1.00 | [0.99-1.02] | 0.49 | 0.89 |
| hdm_birth | rs17881320 | S1 | 1245 | 4 | 1.02 | [0.98-1.06] | 0.41 | 1.02 | [0.98-1.06] | 0.41 | 0.92 |
| hdm_birth | rs17881320 | S2 | 1820 | 4 | 1.00 | [0.99-1.02] | 0.62 | 1.00 | [0.99-1.02] | 0.62 | 0.89 |
| hdm_birth | rs17881320 | S3 | 1245 | 4 | 1.02 | [0.98-1.07] | 0.38 | 1.02 | [0.98-1.07] | 0.38 | 0.87 |
| hdm_birth | rs2041733 | I1 | 1837 | 4 | 1.00 | [1.00-1.01] | 0.27 | 1.00 | [0.99-1.01] | 0.49 | 0.35 |
| hdm_birth | rs2041733 | S1 | 1245 | 4 | 0.99 | [0.98-1.01] | 0.36 | 0.99 | [0.98-1.01] | 0.36 | 0.93 |
| hdm_birth | rs2041733 | S2 | 1837 | 4 | 1.01 | [1.00-1.01] | 0.2 | 1.00 | [0.99-1.02] | 0.8 | 0.25 |
| hdm_birth | rs2041733 | S3 | 1245 | 4 | 0.99 | [0.97-1.01] | 0.28 | 0.99 | [0.97-1.01] | 0.28 | 0.87 |
| hdm_birth | rs2143950 | I1 | 1833 | 4 | 1.00 | [0.99-1.01] | 0.65 | 1.02 | [0.99-1.03] | 0.27 | 0.13 |
| hdm_birth | rs2143950 | S1 | 1245 | 4 | 1.03 | [1.00-1.07] | 0.05 | 1.03 | [1.00-1.07] | 7.40E-02 | 0.30 |
| hdm_birth | rs2143950 | S2 | 1833 | 4 | 1.00 | [0.99-1.01] | 0.43 | 1.02 | [0.99-1.03] | 0.23 | 0.15 |
| hdm_birth | rs2143950 | S3 | 1245 | 4 | 1.04 | [1.00-1.07] | 0.04 | 1.04 | [1.00-1.07] | 4.20E-02 | 0.36 |
| hdm_birth | rs2164983 | I1 | 1837 | 4 | 1.00 | [0.99-1.01] | 0.79 | 1.00 | [0.99-1.01] | 0.79 | 0.85 |
| hdm_birth | rs2164983 | S1 | 1245 | 4 | 1.00 | [0.98-1.02] | 0.99 | 1.00 | [0.98-1.02] | 0.99 | 0.87 |
| hdm_birth | rs2164983 | S2 | 1837 | 4 | 1.00 | [0.99-1.01] | 0.96 | 1.00 | [0.99-1.01] | 0.96 | 0.88 |
| hdm_birth | rs2164983 | S3 | 1245 | 4 | 1.00 | [0.98-1.02] | 0.94 | 1.00 | [0.98-1.02] | 0.94 | 0.83 |
| hdm_birth | rs2227483 | I1 | 1833 | 4 | 0.99 | [0.99-1.00] | 0.11 | 0.99 | [0.99-1.00] | 0.11 | 0.59 |
| hdm_birth | rs2227483 | S1 | 1245 | 4 | 0.99 | [0.98-1.01] | 0.24 | 0.99 | [0.98-1.01] | 0.24 | 0.46 |
| hdm_birth | rs2227483 | S2 | 1833 | 4 | 0.99 | [0.99-1.00] | 0.16 | 0.99 | [0.99-1.00] | 0.16 | 0.64 |
| hdm_birth | rs2227483 | S3 | 1245 | 4 | 0.99 | [0.98-1.01] | 0.25 | 0.99 | [0.98-1.01] | 0.25 | 0.48 |
| hdm_birth | rs2228145 | I1 | 1837 | 4 | 1.00 | [0.99-1.01] | 0.88 | 1.01 | [0.99-1.02] | 0.39 | 0.22 |
| hdm_birth | rs2228145 | S1 | 1245 | 4 | 1.02 | [1.00-1.04] | 5.10E-02 | 1.02 | [1.00-1.04] | 5.10E-02 | 0.93 |
| hdm_birth | rs2228145 | S2 | 1837 | 4 | 1.00 | [0.99-1.01] | 0.93 | 1.00 | [0.99-1.01] | 0.5 | 0.28 |
| hdm_birth | rs2228145 | S3 | 1245 | 4 | 1.02 | [1.00-1.04] | 7.50E-02 | 1.02 | [1.00-1.04] | 7.50E-02 | 0.89 |
| hdm_birth | rs2897442 | I1 | 1836 | 4 | 1.00 | [1.00-1.01] | 0.24 | 1.00 | [1.00-1.01] | 0.24 | 0.68 |
| hdm_birth | rs2897442 | S1 | 1245 | 4 | 1.00 | [0.99-1.02] | 0.74 | 1.00 | [0.99-1.02] | 0.74 | 0.53 |
| hdm_birth | rs2897442 | S2 | 1836 | 4 | 1.01 | [1.00-1.01] | 0.16 | 1.01 | [1.00-1.01] | 0.16 | 0.50 |
| hdm_birth | rs2897442 | S3 | 1245 | 4 | 1.00 | [0.99-1.02] | 0.77 | 1.00 | [0.99-1.02] | 0.77 | 0.38 |
| hdm_birth | rs479844 | I1 | 1837 | 4 | 1.00 | [0.99-1.00] | 0.65 | 1.00 | [0.99-1.00] | 0.65 | 0.89 |
| hdm_birth | rs479844 | S1 | 1245 | 4 | 1.00 | [0.98-1.02] | 0.89 | 1.00 | [0.98-1.02] | 0.89 | 0.82 |
| hdm_birth | rs479844 | S2 | 1837 | 4 | 1.00 | [0.99-1.00] | 0.63 | 1.00 | [0.99-1.00] | 0.63 | 0.80 |
| hdm_birth | rs479844 | S3 | 1245 | 4 | 1.00 | [0.98-1.02] | 0.94 | 1.00 | [0.98-1.02] | 0.94 | 0.72 |
| hdm_birth | rs6010620 | I1 | 1837 | 4 | 0.99 | [0.98-1.00] | 0.24 | 0.99 | [0.98-1.00] | 0.24 | 1.00 |
| hdm_birth | rs6010620 | S1 | 1245 | 4 | 0.99 | [0.97-1.02] | 0.7 | 0.99 | [0.97-1.02] | 0.7 | 0.95 |
| hdm_birth | rs6010620 | S2 | 1837 | 4 | 0.99 | [0.98-1.00] | 0.11 | 0.99 | [0.98-1.00] | 0.11 | 0.99 |
| hdm_birth | rs6010620 | S3 | 1245 | 4 | 0.99 | [0.97-1.02] | 0.57 | 0.99 | [0.97-1.02] | 0.57 | 0.94 |
| hdm_birth | rs6473227 | I1 | 1837 | 4 | 1.01 | [1.00-1.02] | 0.15 | 1.01 | [1.00-1.02] | 0.15 | 0.92 |
| hdm_birth | rs6473227 | S1 | 1245 | 4 | 1.01 | [0.99-1.03] | 0.27 | 1.01 | [0.99-1.03] | 0.27 | 0.81 |
| hdm_birth | rs6473227 | S2 | 1837 | 4 | 1.01 | [1.00-1.02] | 0.21 | 1.01 | [1.00-1.02] | 0.21 | 0.90 |
| hdm_birth | rs6473227 | S3 | 1245 | 4 | 1.01 | [0.99-1.03] | 0.37 | 1.01 | [0.99-1.03] | 0.37 | 0.71 |
| hdm_birth | rs7127307 | I1 | 1837 | 4 | 1.00 | [0.99-1.01] | 0.54 | 1.00 | [0.99-1.01] | 0.54 | 0.51 |
| hdm_birth | rs7127307 | S1 | 1245 | 4 | 1.00 | [0.98-1.03] | 0.72 | 1.00 | [0.98-1.03] | 0.72 | 0.37 |
| hdm_birth | rs7127307 | S2 | 1837 | 4 | 1.00 | [0.99-1.01] | 0.58 | 1.00 | [0.99-1.01] | 0.58 | 0.57 |
| hdm_birth | rs7127307 | S3 | 1245 | 4 | 1.00 | [0.98-1.03] | 0.79 | 1.00 | [0.98-1.03] | 0.79 | 0.40 |
| hdm_birth | rs7146581 | I1 | 1835 | 4 | 1.00 | [0.99-1.01] | 0.83 | 1.00 | [0.99-1.01] | 0.83 | 0.70 |
| hdm_birth | rs7146581 | S1 | 1245 | 4 | 1.00 | [0.99-1.02] | 0.66 | 1.00 | [0.99-1.02] | 0.66 | 0.72 |
| hdm_birth | rs7146581 | S2 | 1835 | 4 | 1.00 | [0.99-1.01] | 0.76 | 1.00 | [0.99-1.01] | 0.76 | 0.83 |
| hdm_birth | rs7146581 | S3 | 1245 | 4 | 1.00 | [0.99-1.02] | 0.82 | 1.00 | [0.99-1.02] | 0.82 | 0.74 |
| hdm_birth | rs7927894 | I1 | 1799 | 4 | 1.00 | [0.99-1.01] | 0.92 | 0.99 | [0.96-1.03] | 0.48 | 0.03 |
| hdm_birth | rs7927894 | S1 | 1245 | 4 | 0.98 | [0.96-1.00] | 1.50E-02 | 0.98 | [0.96-1.00] | 1.50E-02 | 0.99 |
| hdm_birth | rs7927894 | S2 | 1799 | 4 | 1.00 | [0.99-1.01] | 0.9 | 0.99 | [0.97-1.03] | 0.53 | 0.04 |
| hdm_birth | rs7927894 | S3 | 1245 | 4 | 0.98 | [0.96-1.00] | 2.20E-02 | 0.98 | [0.96-1.00] | 2.20E-02 | 0.96 |
| hdm_birth | | M1 | 2877 | 5 | 1.00 | [0.99-1.00] | 0.31 | 1.00 | [0.99-1.00] | 0.41 | 0.24 |
| hdm_birth | | M2 | 2285 | 5 | 1.00 | [0.99-1.00] | 0.26 | 0.99 | [0.98-1.01] | 0.46 | 0.16 |
| hdm_birth | | M3 | 2877 | 5 | 1.00 | [0.99-1.00] | 0.32 | 1.00 | [0.99-1.00] | 0.4 | 0.25 |
| hdm_birth | | M4 | 2285 | 5 | 1.00 | [0.99-1.00] | 0.26 | 1.00 | [0.98-1.01] | 0.45 | 0.17 |
| no2 | flg | I1 | 4727 | 5 | 1.03 | [0.99-1.07] | 0.16 | 1.03 | [0.97-1.09] | 0.34 | 0.16 |
| no2 | flg | S1 | 4275 | 5 | 1.02 | [0.98-1.06] | 0.38 | 1.01 | [0.95-1.08] | 0.7 | 0.14 |
| no2 | flg | S2 | 4727 | 5 | 1.03 | [0.99-1.08] | 0.12 | 1.03 | [0.97-1.10] | 0.31 | 0.13 |
| no2 | flg | S3 | 4275 | 5 | 1.02 | [0.98-1.06] | 0.35 | 1.01 | [0.95-1.09] | 0.72 | 0.10 |
| no2 | rs10214237 | I1 | 5330 | 5 | 1.00 | [0.99-1.01] | 0.87 | 1.00 | [0.98-1.01] | 0.59 | 0.26 |
| no2 | rs10214237 | S1 | 4813 | 5 | 1.00 | [0.99-1.01] | 0.99 | 1.00 | [0.99-1.01] | 0.92 | 0.36 |
| no2 | rs10214237 | S2 | 5330 | 5 | 1.00 | [0.99-1.01] | 0.84 | 0.99 | [0.98-1.02] | 0.49 | 0.18 |
| no2 | rs10214237 | S3 | 4813 | 5 | 1.00 | [0.99-1.01] | 0.96 | 1.00 | [0.99-1.01] | 0.79 | 0.31 |
| no2 | rs1057258 | I1 | 5315 | 5 | 0.99 | [0.98-1.01] | 0.31 | 0.99 | [0.98-1.01] | 0.31 | 0.48 |
| no2 | rs1057258 | S1 | 4813 | 5 | 0.99 | [0.98-1.01] | 0.29 | 0.99 | [0.98-1.01] | 0.29 | 0.89 |
| no2 | rs1057258 | S2 | 5315 | 5 | 0.99 | [0.98-1.01] | 0.26 | 0.99 | [0.98-1.01] | 0.26 | 0.56 |
| no2 | rs1057258 | S3 | 4813 | 5 | 0.99 | [0.98-1.01] | 0.25 | 0.99 | [0.98-1.01] | 0.25 | 0.89 |
| no2 | rs10995251 | I1 | 5328 | 5 | 1.01 | [1.00-1.02] | 0.13 | 1.01 | [1.00-1.02] | 0.13 | 0.95 |
| no2 | rs10995251 | S1 | 4813 | 5 | 1.01 | [1.00-1.02] | 0.18 | 1.01 | [1.00-1.02] | 0.18 | 0.91 |
| no2 | rs10995251 | S2 | 5328 | 5 | 1.01 | [1.00-1.02] | 0.22 | 1.01 | [1.00-1.02] | 0.22 | 0.98 |
| no2 | rs10995251 | S3 | 4813 | 5 | 1.01 | [0.99-1.02] | 0.29 | 1.01 | [0.99-1.02] | 0.29 | 0.96 |
| no2 | rs112111458 | I1 | 5316 | 5 | 0.99 | [0.98-1.01] | 0.46 | 0.99 | [0.98-1.01] | 0.46 | 0.99 |
| no2 | rs112111458 | S1 | 4813 | 5 | 1.00 | [0.98-1.01] | 0.55 | 1.00 | [0.98-1.01] | 0.55 | 0.99 |
| no2 | rs112111458 | S2 | 5316 | 5 | 0.99 | [0.98-1.01] | 0.32 | 0.99 | [0.98-1.01] | 0.32 | 0.99 |

| | | | | | | | | | | | |
|---|---|---|---|---|---|---|---|---|---|---|---|
| no2 | rs112111458 | S3 | 4813 | 5 | 0.99 | [0.98-1.01] | 0.38 | 0.99 | [0.98-1.01] | 0.38 | 0.98 |
| no2 | rs11657987 | I1 | 5304 | 5 | 1.01 | [1.00-1.02] | 0.11 | 1.01 | [1.00-1.02] | 0.11 | 0.39 |
| no2 | rs11657987 | S1 | 4813 | 5 | 1.01 | [1.00-1.02] | 9.30E-02 | 1.01 | [1.00-1.02] | 0.11 | 0.32 |
| no2 | rs11657987 | S2 | 5304 | 5 | 1.01 | [1.00-1.02] | 0.12 | 1.01 | [1.00-1.02] | 0.12 | 0.45 |
| no2 | rs11657987 | S3 | 4813 | 5 | 1.01 | [1.00-1.02] | 0.1 | 1.01 | [1.00-1.02] | 0.1 | 0.39 |
| no2 | rs12153855 | I1 | 5332 | 5 | 1.00 | [0.98-1.01] | 0.62 | 1.00 | [0.97-1.03] | 0.75 | 0.06 |
| no2 | rs12153855 | S1 | 4813 | 5 | 1.00 | [0.98-1.01] | 0.65 | 0.99 | [0.97-1.02] | 0.6 | 0.18 |
| no2 | rs12153855 | S2 | 5332 | 5 | 0.99 | [0.98-1.01] | 0.54 | 0.99 | [0.96-1.03] | 0.71 | 0.04 |
| no2 | rs12153855 | S3 | 4813 | 5 | 1.00 | [0.98-1.01] | 0.57 | 0.99 | [0.97-1.02] | 0.51 | 0.19 |
| no2 | rs12295535 | I1 | 5332 | 5 | 1.01 | [0.98-1.05] | 0.48 | 1.01 | [0.98-1.05] | 0.48 | 0.70 |
| no2 | rs12295535 | S1 | 4813 | 5 | 1.02 | [0.98-1.06] | 0.4 | 1.02 | [0.98-1.06] | 0.4 | 0.56 |
| no2 | rs12295535 | S2 | 5332 | 5 | 1.02 | [0.98-1.06] | 0.45 | 1.02 | [0.98-1.06] | 0.45 | 0.63 |
| no2 | rs12295535 | S3 | 4813 | 5 | 1.02 | [0.98-1.06] | 0.36 | 1.02 | [0.98-1.06] | 0.36 | 0.49 |
| no2 | rs13015714 | I1 | 5332 | 5 | 1.01 | [0.99-1.02] | 0.37 | 1.01 | [0.99-1.02] | 0.37 | 0.67 |
| no2 | rs13015714 | S1 | 4813 | 5 | 1.01 | [0.99-1.02] | 0.41 | 1.01 | [0.99-1.02] | 0.41 | 0.46 |
| no2 | rs13015714 | S2 | 5332 | 5 | 1.01 | [0.99-1.02] | 0.36 | 1.01 | [0.99-1.02] | 0.36 | 0.66 |
| no2 | rs13015714 | S3 | 4813 | 5 | 1.01 | [0.99-1.02] | 0.4 | 1.01 | [0.99-1.02] | 0.4 | 0.46 |
| no2 | rs16948048 | I1 | 5330 | 5 | 1.00 | [0.99-1.01] | 0.63 | 1.00 | [0.99-1.01] | 0.63 | 0.80 |
| no2 | rs16948048 | S1 | 4813 | 5 | 1.00 | [0.99-1.01] | 0.64 | 1.00 | [0.99-1.01] | 0.64 | 0.67 |
| no2 | rs16948048 | S2 | 5330 | 5 | 1.00 | [0.99-1.01] | 0.65 | 1.00 | [0.99-1.01] | 0.65 | 0.68 |
| no2 | rs16948048 | S3 | 4813 | 5 | 1.00 | [0.99-1.01] | 0.67 | 1.00 | [0.99-1.01] | 0.67 | 0.52 |
| no2 | rs17389644 | I1 | 5324 | 5 | 1.00 | [0.99-1.01] | 0.71 | 1.00 | [0.99-1.01] | 0.71 | 0.66 |
| no2 | rs17389644 | S1 | 4813 | 5 | 1.00 | [0.98-1.01] | 0.65 | 1.00 | [0.98-1.01] | 0.65 | 0.59 |
| no2 | rs17389644 | S2 | 5324 | 5 | 1.00 | [0.99-1.01] | 0.81 | 1.00 | [0.99-1.01] | 0.81 | 0.57 |
| no2 | rs17389644 | S3 | 4813 | 5 | 1.00 | [0.98-1.01] | 0.71 | 1.00 | [0.98-1.01] | 0.71 | 0.55 |
| no2 | rs17881320 | I1 | 5317 | 5 | 1.00 | [0.98-1.02] | 0.9 | 1.00 | [0.97-1.03] | 0.77 | 0.11 |
| no2 | rs17881320 | S1 | 4813 | 5 | 1.00 | [0.98-1.02] | 0.96 | 1.01 | [0.97-1.04] | 0.64 | 0.06 |
| no2 | rs17881320 | S2 | 5317 | 5 | 1.00 | [0.98-1.02] | 0.9 | 1.00 | [0.97-1.03] | 0.79 | 0.13 |
| no2 | rs17881320 | S3 | 4813 | 5 | 1.00 | [0.98-1.02] | 0.98 | 1.01 | [0.97-1.04] | 0.67 | 0.05 |
| no2 | rs2041733 | I1 | 5332 | 5 | 1.01 | [1.00-1.02] | 0.19 | 1.00 | [0.98-1.03] | 0.88 | 0.03 |
| no2 | rs2041733 | S1 | 4813 | 5 | 1.01 | [1.00-1.02] | 0.17 | 1.00 | [0.98-1.03] | 0.89 | 0.02 |
| no2 | rs2041733 | S2 | 5332 | 5 | 1.01 | [1.00-1.02] | 0.23 | 1.00 | [0.98-1.03] | 0.93 | 0.04 |
| no2 | rs2041733 | S3 | 4813 | 5 | 1.01 | [1.00-1.02] | 0.18 | 1.00 | [0.98-1.03] | 0.9 | 0.02 |
| no2 | rs2143950 | I1 | 5328 | 5 | 0.99 | [0.98-1.00] | 0.12 | 0.99 | [0.98-1.00] | 0.12 | 0.50 |
| no2 | rs2143950 | S1 | 4813 | 5 | 0.99 | [0.97-1.00] | 8.80E-02 | 0.99 | [0.97-1.00] | 8.80E-02 | 0.49 |
| no2 | rs2143950 | S2 | 5328 | 5 | 0.99 | [0.97-1.00] | 0.11 | 0.99 | [0.97-1.00] | 0.11 | 0.47 |
| no2 | rs2143950 | S3 | 4813 | 5 | 0.99 | [0.97-1.00] | 7.70E-02 | 0.99 | [0.97-1.00] | 7.70E-02 | 0.52 |
| no2 | rs2164983 | I1 | 5332 | 5 | 1.00 | [0.99-1.01] | 0.81 | 1.00 | [0.99-1.01] | 0.81 | 0.96 |
| no2 | rs2164983 | S1 | 4813 | 5 | 1.00 | [0.98-1.01] | 0.77 | 1.00 | [0.98-1.01] | 0.77 | 0.89 |
| no2 | rs2164983 | S2 | 5332 | 5 | 1.00 | [0.98-1.01] | 0.8 | 1.00 | [0.98-1.01] | 0.8 | 0.98 |
| no2 | rs2164983 | S3 | 4813 | 5 | 1.00 | [0.98-1.01] | 0.79 | 1.00 | [0.98-1.01] | 0.79 | 0.93 |
| no2 | rs2227483 | I1 | 5329 | 5 | 1.00 | [0.99-1.01] | 0.73 | 1.00 | [0.99-1.01] | 0.73 | 0.46 |
| no2 | rs2227483 | S1 | 4813 | 5 | 1.00 | [0.99-1.01] | 0.77 | 1.00 | [0.99-1.01] | 0.77 | 0.45 |
| no2 | rs2227483 | S2 | 5329 | 5 | 1.00 | [0.99-1.01] | 0.73 | 1.00 | [0.99-1.01] | 0.73 | 0.41 |
| no2 | rs2227483 | S3 | 4813 | 5 | 1.00 | [0.99-1.01] | 0.82 | 1.00 | [0.99-1.01] | 0.82 | 0.46 |
| no2 | rs2228145 | I1 | 5332 | 5 | 1.01 | [1.00-1.02] | 0.19 | 1.01 | [0.99-1.02] | 0.45 | 0.10 |
| no2 | rs2228145 | S1 | 4813 | 5 | 1.01 | [1.00-1.02] | 0.18 | 1.01 | [0.99-1.03] | 0.44 | 0.06 |
| no2 | rs2228145 | S2 | 5332 | 5 | 1.01 | [1.00-1.02] | 0.16 | 1.01 | [0.99-1.03] | 0.4 | 0.08 |
| no2 | rs2228145 | S3 | 4813 | 5 | 1.01 | [1.00-1.02] | 0.15 | 1.01 | [0.99-1.03] | 0.41 | 0.06 |
| no2 | rs2897442 | I1 | 5332 | 5 | 1.00 | [0.99-1.01] | 0.86 | 1.00 | [0.99-1.01] | 0.86 | 0.80 |
| no2 | rs2897442 | S1 | 4813 | 5 | 1.00 | [0.99-1.02] | 0.58 | 1.00 | [0.99-1.02] | 0.58 | 0.81 |
| no2 | rs2897442 | S2 | 5332 | 5 | 1.00 | [0.99-1.01] | 0.83 | 1.00 | [0.99-1.01] | 0.83 | 0.68 |
| no2 | rs2897442 | S3 | 4813 | 5 | 1.00 | [0.99-1.02] | 0.62 | 1.00 | [0.99-1.02] | 0.62 | 0.80 |
| no2 | rs479844 | I1 | 5332 | 5 | 1.00 | [0.99-1.01] | 0.61 | 1.00 | [0.99-1.01] | 0.61 | 0.47 |
| no2 | rs479844 | S1 | 4813 | 5 | 1.00 | [0.99-1.01] | 0.53 | 1.00 | [0.99-1.01] | 0.53 | 0.45 |
| no2 | rs479844 | S2 | 5332 | 5 | 1.00 | [0.99-1.01] | 0.7 | 1.00 | [0.99-1.01] | 0.78 | 0.38 |
| no2 | rs479844 | S3 | 4813 | 5 | 1.00 | [0.99-1.01] | 0.6 | 1.00 | [0.99-1.01] | 0.6 | 0.50 |
| no2 | rs6010620 | I1 | 5332 | 5 | 0.99 | [0.98-1.00] | 0.22 | 0.99 | [0.98-1.01] | 0.41 | 0.25 |
| no2 | rs6010620 | S1 | 4813 | 5 | 0.99 | [0.98-1.00] | 0.14 | 0.99 | [0.97-1.01] | 0.36 | 0.13 |
| no2 | rs6010620 | S2 | 5332 | 5 | 0.99 | [0.98-1.00] | 0.16 | 0.99 | [0.98-1.01] | 0.38 | 0.21 |
| no2 | rs6010620 | S3 | 4813 | 5 | 0.99 | [0.98-1.00] | 0.11 | 0.99 | [0.97-1.01] | 0.3 | 0.15 |
| no2 | rs6473227 | I1 | 5332 | 5 | 1.00 | [0.99-1.01] | 0.83 | 1.00 | [0.99-1.01] | 0.93 | 0.34 |
| no2 | rs6473227 | S1 | 4813 | 5 | 1.00 | [0.99-1.01] | 0.73 | 1.00 | [0.99-1.02] | 0.96 | 0.19 |
| no2 | rs6473227 | S2 | 5332 | 5 | 1.00 | [0.99-1.01] | 0.94 | 1.00 | [0.99-1.01] | 0.95 | 0.32 |
| no2 | rs6473227 | S3 | 4813 | 5 | 1.00 | [0.99-1.01] | 0.82 | 1.00 | [0.99-1.02] | 0.99 | 0.21 |
| no2 | rs7127307 | I1 | 5332 | 5 | 1.00 | [0.99-1.01] | 0.86 | 1.00 | [0.99-1.01] | 0.86 | 0.43 |
| no2 | rs7127307 | S1 | 4813 | 5 | 1.00 | [0.99-1.01] | 0.86 | 1.00 | [0.99-1.01] | 0.86 | 0.50 |
| no2 | rs7127307 | S2 | 5332 | 5 | 1.00 | [0.99-1.01] | 0.98 | 1.00 | [0.99-1.01] | 0.94 | 0.33 |
| no2 | rs7127307 | S3 | 4813 | 5 | 1.00 | [0.99-1.01] | 0.97 | 1.00 | [0.99-1.01] | 0.97 | 0.48 |
| no2 | rs7146581 | I1 | 5331 | 5 | 1.00 | [0.98-1.01] | 0.48 | 0.99 | [0.97-1.02] | 0.48 | 0.02 |
| no2 | rs7146581 | S1 | 4813 | 5 | 1.00 | [0.98-1.01] | 0.59 | 0.99 | [0.97-1.02] | 0.67 | 0.03 |
| no2 | rs7146581 | S2 | 5331 | 5 | 1.00 | [0.98-1.01] | 0.5 | 0.99 | [0.96-1.02] | 0.53 | 0.01 |
| no2 | rs7146581 | S3 | 4813 | 5 | 1.00 | [0.98-1.01] | 0.65 | 1.00 | [0.97-1.02] | 0.74 | 0.02 |
| no2 | rs7927894 | I1 | 5300 | 5 | 1.00 | [0.99-1.01] | 0.38 | 1.00 | [0.99-1.01] | 0.38 | 0.50 |
| no2 | rs7927894 | S1 | 4813 | 5 | 1.00 | [0.99-1.01] | 0.38 | 1.00 | [0.99-1.01] | 0.39 | 0.39 |
| no2 | rs7927894 | S2 | 5300 | 5 | 0.99 | [0.98-1.01] | 0.32 | 0.99 | [0.98-1.01] | 0.32 | 0.50 |
| no2 | rs7927894 | S3 | 4813 | 5 | 0.99 | [0.98-1.01] | 0.34 | 0.99 | [0.98-1.01] | 0.35 | 0.38 |
| no2 | | M1 | 5753 | 5 | 1.01 | [1.00-1.02] | 3.50E-02 | 1.01 | [0.99-1.02] | 0.2 | 0.03 |
| no2 | | M2 | 5234 | 5 | 1.01 | [1.00-1.02] | 2.30E-02 | 1.01 | [0.99-1.03] | 0.19 | 0.01 |
| no2 | | M3 | 5753 | 5 | 1.01 | [1.00-1.02] | 0.04 | 1.01 | [0.99-1.02] | 0.21 | 0.03 |
| no2 | | M4 | 5234 | 5 | 1.01 | [1.00-1.02] | 2.60E-02 | 1.01 | [0.99-1.03] | 0.2 | 0.01 |
| pm10 | flg | I1 | 3237 | 4 | 1.02 | [0.91-1.13] | 0.77 | 0.98 | [0.80-1.24] | 0.83 | 0.19 |
| pm10 | flg | S1 | 2785 | 4 | 1.01 | [0.90-1.13] | 0.88 | 0.94 | [0.74-1.29] | 0.63 | 0.09 |
| pm10 | flg | S2 | 3237 | 4 | 1.03 | [0.92-1.14] | 0.64 | 0.99 | [0.81-1.26] | 0.93 | 0.18 |
| pm10 | flg | S3 | 2785 | 4 | 1.02 | [0.91-1.14] | 0.75 | 0.95 | [0.73-1.33] | 0.73 | 0.07 |
| pm10 | rs10214237 | I1 | 3502 | 4 | 0.98 | [0.94-1.02] | 0.38 | 0.98 | [0.93-1.03] | 0.47 | 0.28 |
| pm10 | rs10214237 | S1 | 2985 | 4 | 0.99 | [0.95-1.03] | 0.54 | 0.99 | [0.95-1.03] | 0.54 | 0.56 |
| pm10 | rs10214237 | S2 | 3502 | 4 | 0.98 | [0.94-1.02] | 0.35 | 0.98 | [0.93-1.03] | 0.45 | 0.26 |
| pm10 | rs10214237 | S3 | 2985 | 4 | 0.99 | [0.94-1.03] | 0.5 | 0.99 | [0.94-1.03] | 0.5 | 0.42 |
| pm10 | rs1057258 | I1 | 3487 | 4 | 0.98 | [0.94-1.03] | 0.46 | 0.98 | [0.94-1.03] | 0.46 | 0.63 |
| pm10 | rs1057258 | S1 | 2985 | 4 | 0.98 | [0.93-1.02] | 0.3 | 0.98 | [0.93-1.02] | 0.3 | 0.42 |

| | | | | | | | | | | | |
|---|---|---|---|---|---|---|---|---|---|---|---|
| pm10 | rs1057258 | S2 | 3487 | 4 | 0.98 | [0.93-1.03] | 0.41 | 0.98 | [0.93-1.03] | 0.41 | 0.65 |
| pm10 | rs1057258 | S3 | 2985 | 4 | 0.97 | [0.93-1.02] | 0.25 | 0.97 | [0.93-1.02] | 0.25 | 0.41 |
| pm10 | rs10995251 | I1 | 3500 | 4 | 1.01 | [0.97-1.04] | 0.78 | 1.01 | [0.97-1.04] | 0.78 | 0.98 |
| pm10 | rs10995251 | S1 | 2985 | 4 | 1.01 | [0.97-1.04] | 0.75 | 1.01 | [0.97-1.04] | 0.75 | 0.92 |
| pm10 | rs10995251 | S2 | 3500 | 4 | 1.00 | [0.97-1.04] | 0.86 | 1.00 | [0.97-1.04] | 0.86 | 0.98 |
| pm10 | rs10995251 | S3 | 2985 | 4 | 1.00 | [0.97-1.04] | 0.84 | 1.00 | [0.97-1.04] | 0.84 | 0.94 |
| pm10 | rs112111458 | I1 | 3488 | 4 | 1.00 | [0.95-1.05] | 0.95 | 1.01 | [0.94-1.07] | 0.79 | 0.25 |
| pm10 | rs112111458 | S1 | 2985 | 4 | 1.00 | [0.95-1.04] | 0.85 | 1.01 | [0.93-1.07] | 0.88 | 0.19 |
| pm10 | rs112111458 | S2 | 3488 | 4 | 0.99 | [0.95-1.04] | 0.82 | 1.01 | [0.93-1.07] | 0.83 | 0.21 |
| pm10 | rs112111458 | S3 | 2985 | 4 | 0.99 | [0.94-1.04] | 0.76 | 1.01 | [0.93-1.08] | 0.88 | 0.15 |
| pm10 | rs11657987 | I1 | 3476 | 4 | 1.00 | [0.96-1.04] | 0.93 | 1.00 | [0.96-1.04] | 0.93 | 0.51 |
| pm10 | rs11657987 | S1 | 2985 | 4 | 1.00 | [0.97-1.04] | 0.91 | 1.00 | [0.97-1.04] | 0.91 | 0.64 |
| pm10 | rs11657987 | S2 | 3476 | 4 | 1.00 | [0.96-1.04] | 0.94 | 1.00 | [0.96-1.04] | 0.94 | 0.50 |
| pm10 | rs11657987 | S3 | 2985 | 4 | 1.00 | [0.97-1.04] | 0.93 | 1.00 | [0.97-1.04] | 0.93 | 0.65 |
| pm10 | rs12153855 | I1 | 3504 | 4 | 1.01 | [0.95-1.07] | 0.77 | 1.04 | [0.93-1.13] | 0.53 | 0.06 |
| pm10 | rs12153855 | S1 | 2985 | 4 | 1.01 | [0.95-1.07] | 0.83 | 1.01 | [0.95-1.07] | 0.83 | 0.38 |
| pm10 | rs12153855 | S2 | 3504 | 4 | 1.01 | [0.95-1.07] | 0.76 | 1.04 | [0.92-1.13] | 0.53 | 0.05 |
| pm10 | rs12153855 | S3 | 2985 | 4 | 1.01 | [0.95-1.07] | 0.83 | 1.01 | [0.95-1.07] | 0.83 | 0.46 |
| pm10 | rs12295535 | I1 | 3504 | 4 | 1.12 | [0.97-1.29] | 0.11 | 1.11 | [0.93-1.35] | 0.25 | 0.32 |
| pm10 | rs12295535 | S1 | 2985 | 4 | 1.12 | [0.97-1.29] | 0.13 | 1.10 | [0.87-1.41] | 0.41 | 0.21 |
| pm10 | rs12295535 | S2 | 3504 | 4 | 1.13 | [0.98-1.30] | 8.70E-02 | 1.13 | [0.96-1.33] | 0.14 | 0.36 |
| pm10 | rs12295535 | S3 | 2985 | 4 | 1.13 | [0.98-1.31] | 9.30E-02 | 1.13 | [0.98-1.31] | 9.30E-02 | 0.38 |
| pm10 | rs13015714 | I1 | 3504 | 4 | 0.99 | [0.95-1.03] | 0.62 | 0.99 | [0.95-1.03] | 0.62 | 0.51 |
| pm10 | rs13015714 | S1 | 2985 | 4 | 0.99 | [0.95-1.03] | 0.58 | 0.99 | [0.95-1.03] | 0.58 | 0.38 |
| pm10 | rs13015714 | S2 | 3504 | 4 | 0.99 | [0.95-1.03] | 0.58 | 0.99 | [0.95-1.03] | 0.58 | 0.58 |
| pm10 | rs13015714 | S3 | 2985 | 4 | 0.99 | [0.95-1.03] | 0.54 | 0.99 | [0.95-1.03] | 0.54 | 0.41 |
| pm10 | rs16948048 | I1 | 3502 | 4 | 1.01 | [0.97-1.05] | 0.72 | 1.01 | [0.97-1.05] | 0.72 | 0.46 |
| pm10 | rs16948048 | S1 | 2985 | 4 | 1.01 | [0.97-1.04] | 0.8 | 1.00 | [0.96-1.05] | 0.92 | 0.29 |
| pm10 | rs16948048 | S2 | 3502 | 4 | 1.01 | [0.97-1.05] | 0.58 | 1.01 | [0.97-1.05] | 0.58 | 0.42 |
| pm10 | rs16948048 | S3 | 2985 | 4 | 1.01 | [0.97-1.05] | 0.61 | 1.01 | [0.96-1.06] | 0.82 | 0.23 |
| pm10 | rs17389644 | I1 | 3496 | 4 | 1.01 | [0.97-1.05] | 0.69 | 1.01 | [0.97-1.05] | 0.69 | 0.52 |
| pm10 | rs17389644 | S1 | 2985 | 4 | 1.01 | [0.97-1.06] | 0.52 | 1.01 | [0.97-1.06] | 0.52 | 0.67 |
| pm10 | rs17389644 | S2 | 3496 | 4 | 1.01 | [0.97-1.06] | 0.63 | 1.01 | [0.97-1.06] | 0.63 | 0.50 |
| pm10 | rs17389644 | S3 | 2985 | 4 | 1.01 | [0.97-1.06] | 0.55 | 1.01 | [0.97-1.06] | 0.55 | 0.58 |
| pm10 | rs17881320 | I1 | 3489 | 4 | 1.00 | [0.92-1.07] | 0.91 | 0.99 | [0.88-1.12] | 0.8 | 0.18 |
| pm10 | rs17881320 | S1 | 2985 | 4 | 1.01 | [0.93-1.09] | 0.86 | 1.01 | [0.90-1.13] | 0.93 | 0.19 |
| pm10 | rs17881320 | S2 | 3489 | 4 | 0.99 | [0.92-1.07] | 0.87 | 0.99 | [0.89-1.10] | 0.82 | 0.25 |
| pm10 | rs17881320 | S3 | 2985 | 4 | 1.00 | [0.93-1.08] | 0.94 | 1.00 | [0.90-1.11] | 0.99 | 0.23 |
| pm10 | rs2041733 | I1 | 3504 | 4 | 0.99 | [0.95-1.03] | 0.56 | 1.04 | [0.94-1.09] | 0.48 | 0.00 |
| pm10 | rs2041733 | S1 | 2985 | 4 | 0.99 | [0.95-1.02] | 0.46 | 1.01 | [0.92-1.08] | 0.86 | 0.01 |
| pm10 | rs2041733 | S2 | 3504 | 4 | 0.99 | [0.95-1.02] | 0.54 | 1.04 | [0.94-1.10] | 0.46 | 0.00 |
| pm10 | rs2041733 | S3 | 2985 | 4 | 0.99 | [0.95-1.02] | 0.44 | 1.01 | [0.92-1.08] | 0.85 | 0.01 |
| pm10 | rs2143950 | I1 | 3500 | 4 | 1.00 | [0.95-1.06] | 0.9 | 0.98 | [0.87-1.13] | 0.75 | 0.02 |
| pm10 | rs2143950 | S1 | 2985 | 4 | 1.00 | [0.95-1.05] | 0.94 | 0.96 | [0.84-1.14] | 0.5 | 0.01 |
| pm10 | rs2143950 | S2 | 3500 | 4 | 0.99 | [0.94-1.05] | 0.85 | 0.97 | [0.86-1.13] | 0.66 | 0.01 |
| pm10 | rs2143950 | S3 | 2985 | 4 | 0.99 | [0.94-1.04] | 0.7 | 0.95 | [0.83-1.13] | 0.43 | 0.01 |
| pm10 | rs2164983 | I1 | 3504 | 4 | 1.01 | [0.96-1.07] | 0.62 | 1.01 | [0.95-1.08] | 0.75 | 0.33 |
| pm10 | rs2164983 | S1 | 2985 | 4 | 1.01 | [0.96-1.07] | 0.61 | 1.01 | [0.93-1.09] | 0.83 | 0.19 |
| pm10 | rs2164983 | S2 | 3504 | 4 | 1.02 | [0.96-1.07] | 0.54 | 1.01 | [0.96-1.08] | 0.63 | 0.35 |
| pm10 | rs2164983 | S3 | 2985 | 4 | 1.02 | [0.96-1.07] | 0.56 | 1.01 | [0.94-1.10] | 0.77 | 0.20 |
| pm10 | rs2227483 | I1 | 3501 | 4 | 0.99 | [0.96-1.03] | 0.6 | 0.99 | [0.96-1.03] | 0.6 | 0.89 |
| pm10 | rs2227483 | S1 | 2985 | 4 | 0.99 | [0.96-1.03] | 0.67 | 0.99 | [0.96-1.03] | 0.67 | 0.78 |
| pm10 | rs2227483 | S2 | 3501 | 4 | 0.99 | [0.96-1.03] | 0.6 | 0.99 | [0.96-1.03] | 0.6 | 0.92 |
| pm10 | rs2227483 | S3 | 2985 | 4 | 0.99 | [0.96-1.03] | 0.68 | 0.99 | [0.96-1.03] | 0.68 | 0.81 |
| pm10 | rs2228145 | I1 | 3504 | 4 | 1.00 | [0.97-1.04] | 0.85 | 1.00 | [0.97-1.04] | 0.84 | 0.38 |
| pm10 | rs2228145 | S1 | 2985 | 4 | 1.01 | [0.97-1.04] | 0.77 | 1.01 | [0.96-1.05] | 0.73 | 0.24 |
| pm10 | rs2228145 | S2 | 3504 | 4 | 1.01 | [0.97-1.05] | 0.64 | 1.01 | [0.97-1.05] | 0.64 | 0.45 |
| pm10 | rs2228145 | S3 | 2985 | 4 | 1.01 | [0.97-1.05] | 0.66 | 1.01 | [0.97-1.06] | 0.65 | 0.26 |
| pm10 | rs2897442 | I1 | 3504 | 4 | 1.00 | [0.96-1.04] | 0.97 | 1.00 | [0.96-1.04] | 0.97 | 0.79 |
| pm10 | rs2897442 | S1 | 2985 | 4 | 1.00 | [0.96-1.04] | 0.96 | 1.00 | [0.96-1.04] | 0.96 | 0.83 |
| pm10 | rs2897442 | S2 | 3504 | 4 | 0.99 | [0.95-1.04] | 0.77 | 0.99 | [0.95-1.04] | 0.77 | 0.88 |
| pm10 | rs2897442 | S3 | 2985 | 4 | 1.00 | [0.96-1.04] | 0.88 | 1.00 | [0.96-1.04] | 0.88 | 0.89 |
| pm10 | rs479844 | I1 | 3504 | 4 | 0.97 | [0.93-1.00] | 8.30E-02 | 0.95 | [0.89-1.03] | 0.17 | 0.10 |
| pm10 | rs479844 | S1 | 2985 | 4 | 0.97 | [0.93-1.01] | 9.70E-02 | 0.95 | [0.88-1.05] | 0.2 | 0.03 |
| pm10 | rs479844 | S2 | 3504 | 4 | 0.97 | [0.93-1.00] | 7.40E-02 | 0.95 | [0.89-1.04] | 0.16 | 0.08 |
| pm10 | rs479844 | S3 | 2985 | 4 | 0.97 | [0.93-1.00] | 8.30E-02 | 0.94 | [0.87-1.05] | 0.19 | 0.02 |
| pm10 | rs6010620 | I1 | 3504 | 4 | 0.98 | [0.94-1.02] | 0.32 | 0.98 | [0.94-1.02] | 0.32 | 0.75 |
| pm10 | rs6010620 | S1 | 2985 | 4 | 0.97 | [0.93-1.02] | 0.24 | 0.97 | [0.93-1.02] | 0.24 | 0.98 |
| pm10 | rs6010620 | S2 | 3504 | 4 | 0.98 | [0.93-1.02] | 0.3 | 0.98 | [0.93-1.02] | 0.3 | 0.70 |
| pm10 | rs6010620 | S3 | 2985 | 4 | 0.97 | [0.93-1.02] | 0.22 | 0.97 | [0.93-1.02] | 0.22 | 0.97 |
| pm10 | rs6473227 | I1 | 3504 | 4 | 1.01 | [0.97-1.05] | 0.65 | 1.02 | [0.96-1.08] | 0.54 | 0.08 |
| pm10 | rs6473227 | S1 | 2985 | 4 | 1.00 | [0.97-1.04] | 0.96 | 1.00 | [0.97-1.04] | 0.96 | 0.72 |
| pm10 | rs6473227 | S2 | 3504 | 4 | 1.01 | [0.97-1.05] | 0.63 | 1.02 | [0.96-1.07] | 0.59 | 0.17 |
| pm10 | rs6473227 | S3 | 2985 | 4 | 1.00 | [0.97-1.04] | 0.84 | 1.00 | [0.97-1.04] | 0.84 | 0.75 |
| pm10 | rs7127307 | I1 | 3504 | 4 | 1.01 | [0.97-1.05] | 0.74 | 1.01 | [0.97-1.05] | 0.74 | 0.69 |
| pm10 | rs7127307 | S1 | 2985 | 4 | 1.01 | [0.97-1.05] | 0.6 | 1.01 | [0.97-1.05] | 0.6 | 0.46 |
| pm10 | rs7127307 | S2 | 3504 | 4 | 1.00 | [0.97-1.04] | 0.83 | 1.00 | [0.97-1.04] | 0.83 | 0.61 |
| pm10 | rs7127307 | S3 | 2985 | 4 | 1.01 | [0.97-1.05] | 0.65 | 1.01 | [0.97-1.05] | 0.65 | 0.41 |
| pm10 | rs7146581 | I1 | 3503 | 4 | 1.02 | [0.98-1.07] | 0.28 | 1.04 | [0.96-1.11] | 0.34 | 0.09 |
| pm10 | rs7146581 | S1 | 2985 | 4 | 1.03 | [0.98-1.07] | 0.24 | 1.04 | [0.96-1.12] | 0.32 | 0.05 |
| pm10 | rs7146581 | S2 | 3503 | 4 | 1.02 | [0.98-1.07] | 0.27 | 1.04 | [0.96-1.11] | 0.32 | 0.07 |
| pm10 | rs7146581 | S3 | 2985 | 4 | 1.03 | [0.98-1.07] | 0.25 | 1.05 | [0.95-1.13] | 0.33 | 0.03 |
| pm10 | rs7927894 | I1 | 3472 | 4 | 0.99 | [0.95-1.02] | 0.46 | 0.99 | [0.95-1.02] | 0.46 | 0.49 |
| pm10 | rs7927894 | S1 | 2985 | 4 | 0.99 | [0.95-1.03] | 0.59 | 0.99 | [0.95-1.03] | 0.59 | 0.91 |
| pm10 | rs7927894 | S2 | 3472 | 4 | 0.99 | [0.95-1.02] | 0.49 | 0.99 | [0.95-1.02] | 0.49 | 0.42 |
| pm10 | rs7927894 | S3 | 2985 | 4 | 0.99 | [0.95-1.03] | 0.66 | 0.99 | [0.95-1.03] | 0.66 | 0.94 |
| pm10 | | M1 | 3884 | 4 | 1.01 | [0.99-1.03] | 0.45 | 0.99 | [0.95-1.06] | 0.81 | 0.03 |
| pm10 | | M2 | 3365 | 4 | 1.01 | [0.99-1.04] | 0.39 | 1.00 | [0.95-1.06] | 0.9 | 0.02 |
| pm10 | | M3 | 3884 | 4 | 1.01 | [0.99-1.03] | 0.4 | 1.00 | [0.95-1.06] | 0.88 | 0.04 |
| pm10 | | M4 | 3365 | 4 | 1.01 | [0.99-1.04] | 0.35 | 1.00 | [0.95-1.06] | 0.94 | 0.02 |
| siblings | flg | I1 | 16745 | 10 | 0.98 | [0.77-1.24] | 0.86 | 1.01 | [0.70-1.42] | 0.96 | 0.08 |

| | | | | | | | | | | | |
|---|---|---|---|---|---|---|---|---|---|---|---|
| siblings | flg | I2 | 7375 | 10 | 1.93 | [1.62-2.30] | 1.80E-13 | 1.91 | [1.53-2.41] | 8.80E-09 | 0.26 |
| siblings | flg | I3 | 9370 | 10 | 1.87 | [1.59-2.19] | 1.70E-14 | 1.87 | [1.57-2.23] | 5.80E-12 | 0.37 |
| siblings | flg | S1 | 12529 | 10 | 0.93 | [0.72-1.19] | 0.55 | 0.93 | [0.72-1.19] | 0.55 | 0.47 |
| siblings | flg | S2 | 15415 | 10 | 0.95 | [0.73-1.24] | 0.72 | 0.98 | [0.64-1.47] | 0.93 | 0.05 |
| siblings | flg | S3 | 11223 | 10 | 0.90 | [0.68-1.18] | 0.44 | 0.89 | [0.65-1.22] | 0.45 | 0.36 |
| siblings | rs10214237 | I1 | 19155 | 11 | 0.84 | [0.75-0.94] | 3.00E-03 | 0.84 | [0.75-0.94] | 3.00E-03 | 0.99 |
| siblings | rs10214237 | I2 | 9142 | 11 | 1.20 | [1.11-1.30] | 8.40E-06 | 1.20 | [1.11-1.30] | 8.40E-06 | 0.99 |
| siblings | rs10214237 | I3 | 10013 | 11 | 1.02 | [0.94-1.10] | 0.63 | 1.02 | [0.94-1.10] | 0.63 | 0.86 |
| siblings | rs10214237 | S1 | 13402 | 11 | 0.84 | [0.74-0.95] | 6.20E-03 | 0.84 | [0.74-0.95] | 6.20E-03 | 0.93 |
| siblings | rs10214237 | S2 | 15682 | 11 | 0.84 | [0.74-0.95] | 6.20E-03 | 0.84 | [0.74-0.95] | 6.20E-03 | 0.96 |
| siblings | rs10214237 | S3 | 9945 | 11 | 0.83 | [0.73-0.96] | 8.80E-03 | 0.83 | [0.73-0.96] | 8.80E-03 | 0.74 |
| siblings | rs1057258 | I1 | 18974 | 11 | 0.97 | [0.84-1.11] | 0.64 | 0.97 | [0.81-1.15] | 0.7 | 0.24 |
| siblings | rs1057258 | I2 | 9102 | 11 | 1.07 | [0.97-1.18] | 0.15 | 1.07 | [0.96-1.20] | 0.21 | 0.25 |
| siblings | rs1057258 | I3 | 9872 | 11 | 1.03 | [0.94-1.13] | 0.52 | 1.03 | [0.94-1.13] | 0.52 | 0.85 |
| siblings | rs1057258 | S1 | 13403 | 11 | 1.00 | [0.86-1.16] | 0.97 | 1.00 | [0.86-1.16] | 0.97 | 0.44 |
| siblings | rs1057258 | S2 | 15501 | 11 | 0.93 | [0.80-1.08] | 0.33 | 0.92 | [0.74-1.14] | 0.42 | 0.12 |
| siblings | rs1057258 | S3 | 9946 | 11 | 0.96 | [0.82-1.13] | 0.66 | 0.98 | [0.79-1.20] | 0.86 | 0.15 |
| siblings | rs10995251 | I1 | 18608 | 10 | 0.89 | [0.80-1.00] | 4.20E-02 | 0.89 | [0.80-1.00] | 4.20E-02 | 0.44 |
| siblings | rs10995251 | I2 | 8908 | 10 | 1.07 | [1.00-1.15] | 6.70E-02 | 1.07 | [1.00-1.15] | 6.70E-02 | 0.58 |
| siblings | rs10995251 | I3 | 9700 | 10 | 0.97 | [0.90-1.04] | 0.4 | 0.97 | [0.89-1.06] | 0.51 | 0.27 |
| siblings | rs10995251 | S1 | 12847 | 10 | 0.87 | [0.77-0.98] | 2.20E-02 | 0.87 | [0.76-1.00] | 3.80E-02 | 0.31 |
| siblings | rs10995251 | S2 | 15135 | 10 | 0.89 | [0.79-1.00] | 5.10E-02 | 0.89 | [0.79-1.00] | 5.10E-02 | 0.52 |
| siblings | rs10995251 | S3 | 9390 | 10 | 0.86 | [0.76-0.98] | 2.20E-02 | 0.86 | [0.76-0.98] | 2.20E-02 | 0.41 |
| siblings | rs112111458 | I1 | 16911 | 9 | 1.07 | [0.92-1.24] | 0.39 | 1.07 | [0.92-1.24] | 0.39 | 0.96 |
| siblings | rs112111458 | I2 | 7460 | 9 | 0.98 | [0.88-1.09] | 0.73 | 0.98 | [0.88-1.10] | 0.78 | 0.40 |
| siblings | rs112111458 | I3 | 9451 | 9 | 1.05 | [0.94-1.16] | 0.39 | 1.07 | [0.95-1.18] | 0.28 | 0.29 |
| siblings | rs112111458 | S1 | 12817 | 9 | 1.05 | [0.90-1.24] | 0.52 | 1.05 | [0.90-1.24] | 0.52 | 0.98 |
| siblings | rs112111458 | S2 | 14022 | 9 | 1.04 | [0.89-1.22] | 0.59 | 1.04 | [0.89-1.22] | 0.59 | 0.90 |
| siblings | rs112111458 | S3 | 9944 | 9 | 1.03 | [0.87-1.21] | 0.76 | 1.03 | [0.87-1.21] | 0.76 | 0.93 |
| siblings | rs11657987 | I1 | 19002 | 11 | 1.00 | [0.90-1.11] | 0.99 | 1.00 | [0.90-1.12] | 0.96 | 0.41 |
| siblings | rs11657987 | I2 | 9099 | 11 | 1.06 | [0.99-1.14] | 8.80E-02 | 1.06 | [0.97-1.16] | 0.21 | 0.21 |
| siblings | rs11657987 | I3 | 9903 | 11 | 1.07 | [1.00-1.15] | 4.50E-02 | 1.07 | [1.00-1.15] | 4.50E-02 | 0.44 |
| siblings | rs11657987 | S1 | 13399 | 11 | 0.97 | [0.87-1.09] | 0.6 | 0.97 | [0.86-1.11] | 0.7 | 0.31 |
| siblings | rs11657987 | S2 | 15529 | 11 | 0.98 | [0.88-1.10] | 0.79 | 0.98 | [0.88-1.10] | 0.79 | 0.61 |
| siblings | rs11657987 | S3 | 9942 | 11 | 0.95 | [0.85-1.08] | 0.45 | 0.95 | [0.85-1.08] | 0.45 | 0.54 |
| siblings | rs12153855 | I1 | 16533 | 8 | 1.05 | [0.87-1.26] | 0.64 | 1.05 | [0.86-1.28] | 0.66 | 0.36 |
| siblings | rs12153855 | I2 | 7277 | 8 | 1.09 | [0.96-1.24] | 0.19 | 1.09 | [0.96-1.24] | 0.19 | 0.76 |
| siblings | rs12153855 | I3 | 9256 | 8 | 1.16 | [1.02-1.32] | 2.10E-02 | 1.16 | [1.02-1.32] | 2.10E-02 | 0.50 |
| siblings | rs12153855 | S1 | 12263 | 8 | 1.01 | [0.83-1.23] | 0.92 | 1.01 | [0.83-1.23] | 0.92 | 0.48 |
| siblings | rs12153855 | S2 | 13644 | 8 | 1.03 | [0.85-1.25] | 0.77 | 1.03 | [0.81-1.30] | 0.83 | 0.27 |
| siblings | rs12153855 | S3 | 9390 | 8 | 0.98 | [0.79-1.21] | 0.84 | 0.98 | [0.79-1.21] | 0.84 | 0.48 |
| siblings | rs12295535 | I1 | 19186 | 11 | 1.04 | [0.76-1.43] | 0.79 | 1.04 | [0.76-1.43] | 0.79 | 0.97 |
| siblings | rs12295535 | I2 | 9153 | 11 | 1.16 | [0.94-1.43] | 0.18 | 1.16 | [0.94-1.43] | 0.18 | 0.56 |
| siblings | rs12295535 | I3 | 10033 | 11 | 1.17 | [0.94-1.44] | 0.16 | 1.17 | [0.94-1.44] | 0.16 | 0.94 |
| siblings | rs12295535 | S1 | 13403 | 11 | 1.15 | [0.81-1.62] | 0.43 | 1.15 | [0.81-1.62] | 0.43 | 0.98 |
| siblings | rs12295535 | S2 | 15713 | 11 | 1.14 | [0.81-1.62] | 0.45 | 1.14 | [0.81-1.62] | 0.45 | 0.97 |
| siblings | rs12295535 | S3 | 9946 | 11 | 1.32 | [0.91-1.94] | 0.15 | 1.32 | [0.91-1.94] | 0.15 | 0.94 |
| siblings | rs13015714 | I1 | 18630 | 10 | 0.98 | [0.87-1.11] | 0.76 | 0.98 | [0.87-1.11] | 0.76 | 0.50 |
| siblings | rs13015714 | I2 | 8916 | 10 | 1.11 | [1.02-1.21] | 1.10E-02 | 1.11 | [1.02-1.21] | 1.10E-02 | 0.88 |
| siblings | rs13015714 | I3 | 9714 | 10 | 1.06 | [0.97-1.15] | 0.19 | 1.05 | [0.96-1.16] | 0.27 | 0.37 |
| siblings | rs13015714 | S1 | 12847 | 10 | 0.99 | [0.86-1.13] | 0.84 | 0.98 | [0.84-1.16] | 0.85 | 0.27 |
| siblings | rs13015714 | S2 | 15157 | 10 | 0.97 | [0.85-1.11] | 0.69 | 0.97 | [0.84-1.13] | 0.73 | 0.31 |
| siblings | rs13015714 | S3 | 9390 | 10 | 0.97 | [0.84-1.13] | 0.72 | 0.98 | [0.79-1.21] | 0.83 | 0.10 |
| siblings | rs16948048 | I1 | 18598 | 10 | 0.99 | [0.88-1.10] | 0.79 | 0.99 | [0.88-1.10] | 0.79 | 0.68 |
| siblings | rs16948048 | I2 | 8910 | 10 | 1.03 | [0.96-1.11] | 0.37 | 1.03 | [0.96-1.11] | 0.37 | 0.58 |
| siblings | rs16948048 | I3 | 9688 | 10 | 1.01 | [0.93-1.08] | 0.86 | 1.01 | [0.93-1.08] | 0.86 | 0.66 |
| siblings | rs16948048 | S1 | 12847 | 10 | 1.00 | [0.89-1.12] | 0.98 | 1.00 | [0.89-1.12] | 0.98 | 0.65 |
| siblings | rs16948048 | S2 | 15125 | 10 | 0.98 | [0.87-1.11] | 0.79 | 0.98 | [0.87-1.11] | 0.79 | 0.70 |
| siblings | rs16948048 | S3 | 9390 | 10 | 1.01 | [0.89-1.14] | 0.93 | 1.01 | [0.89-1.14] | 0.93 | 0.74 |
| siblings | rs17389644 | I1 | 18529 | 10 | 0.98 | [0.86-1.11] | 0.77 | 1.00 | [0.85-1.15] | 0.95 | 0.21 |
| siblings | rs17389644 | I2 | 8890 | 10 | 1.06 | [0.97-1.15] | 0.17 | 1.04 | [0.93-1.18] | 0.49 | 0.22 |
| siblings | rs17389644 | I3 | 9639 | 10 | 1.08 | [0.99-1.17] | 0.1 | 1.06 | [0.92-1.23] | 0.43 | 0.06 |
| siblings | rs17389644 | S1 | 12847 | 10 | 1.01 | [0.88-1.15] | 0.94 | 1.04 | [0.86-1.21] | 0.69 | 0.16 |
| siblings | rs17389644 | S2 | 15056 | 10 | 0.94 | [0.82-1.08] | 0.38 | 0.94 | [0.82-1.08] | 0.38 | 0.74 |
| siblings | rs17389644 | S3 | 9390 | 10 | 0.97 | [0.84-1.13] | 0.72 | 0.97 | [0.84-1.13] | 0.72 | 0.93 |
| siblings | rs17881320 | I1 | 19073 | 11 | 0.98 | [0.81-1.19] | 0.85 | 0.98 | [0.81-1.19] | 0.85 | 0.67 |
| siblings | rs17881320 | I2 | 9127 | 11 | 1.09 | [0.96-1.24] | 0.17 | 1.09 | [0.96-1.24] | 0.17 | 0.49 |
| siblings | rs17881320 | I3 | 9946 | 11 | 1.07 | [0.94-1.22] | 0.28 | 1.08 | [0.94-1.24] | 0.28 | 0.33 |
| siblings | rs17881320 | S1 | 13401 | 11 | 0.95 | [0.77-1.17] | 0.63 | 0.95 | [0.77-1.17] | 0.63 | 0.67 |
| siblings | rs17881320 | S2 | 15600 | 11 | 1.03 | [0.83-1.27] | 0.81 | 1.03 | [0.83-1.27] | 0.81 | 0.66 |
| siblings | rs17881320 | S3 | 9944 | 11 | 1.01 | [0.81-1.27] | 0.92 | 1.01 | [0.81-1.27] | 0.92 | 0.55 |
| siblings | rs2041733 | I1 | 18630 | 10 | 1.06 | [0.95-1.18] | 0.28 | 1.06 | [0.95-1.18] | 0.28 | 0.81 |
| siblings | rs2041733 | I2 | 8916 | 10 | 1.03 | [0.96-1.11] | 0.37 | 1.03 | [0.96-1.11] | 0.37 | 0.98 |
| siblings | rs2041733 | I3 | 9714 | 10 | 1.10 | [1.03-1.19] | 7.90E-03 | 1.11 | [0.99-1.23] | 6.80E-02 | 0.06 |
| siblings | rs2041733 | S1 | 12847 | 10 | 1.04 | [0.93-1.17] | 0.45 | 1.04 | [0.93-1.17] | 0.45 | 0.84 |
| siblings | rs2041733 | S2 | 15157 | 10 | 1.05 | [0.94-1.18] | 0.38 | 1.05 | [0.94-1.18] | 0.38 | 0.75 |
| siblings | rs2041733 | S3 | 9390 | 10 | 1.03 | [0.91-1.16] | 0.64 | 1.03 | [0.91-1.16] | 0.64 | 0.71 |
| siblings | rs2143950 | I1 | 19129 | 11 | 1.09 | [0.95-1.25] | 0.24 | 1.13 | [0.95-1.30] | 0.17 | 0.19 |
| siblings | rs2143950 | I2 | 9141 | 11 | 1.04 | [0.95-1.14] | 0.37 | 1.01 | [0.89-1.18] | 0.9 | 0.13 |
| siblings | rs2143950 | I3 | 9988 | 11 | 1.13 | [1.03-1.25] | 8.80E-03 | 1.13 | [1.03-1.25] | 8.80E-03 | 0.83 |
| siblings | rs2143950 | S1 | 13394 | 11 | 1.10 | [0.95-1.27] | 0.21 | 1.19 | [0.94-1.39] | 0.15 | 0.06 |
| siblings | rs2143950 | S2 | 15656 | 11 | 1.06 | [0.92-1.23] | 0.41 | 1.09 | [0.91-1.28] | 0.36 | 0.21 |
| siblings | rs2143950 | S3 | 9937 | 11 | 1.07 | [0.91-1.25] | 0.41 | 1.13 | [0.87-1.38] | 0.35 | 0.05 |
| siblings | rs2164983 | I1 | 16531 | 8 | 0.99 | [0.85-1.15] | 0.88 | 0.99 | [0.85-1.15] | 0.88 | 0.99 |
| siblings | rs2164983 | I2 | 7277 | 8 | 1.12 | [1.01-1.25] | 3.40E-02 | 1.12 | [1.01-1.25] | 3.40E-02 | 0.44 |
| siblings | rs2164983 | I3 | 9254 | 8 | 1.12 | [1.01-1.24] | 3.30E-02 | 1.12 | [1.01-1.24] | 3.30E-02 | 0.66 |
| siblings | rs2164983 | S1 | 12263 | 8 | 1.01 | [0.86-1.18] | 0.92 | 1.01 | [0.86-1.18] | 0.92 | 0.98 |
| siblings | rs2164983 | S2 | 13642 | 8 | 0.97 | [0.83-1.13] | 0.7 | 0.97 | [0.83-1.13] | 0.7 | 0.95 |
| siblings | rs2164983 | S3 | 9390 | 8 | 0.99 | [0.83-1.17] | 0.89 | 0.99 | [0.83-1.17] | 0.89 | 0.88 |
| siblings | rs2227483 | I1 | 19125 | 11 | 0.96 | [0.87-1.07] | 0.47 | 0.96 | [0.86-1.08] | 0.53 | 0.37 |
| siblings | rs2227483 | I2 | 9135 | 11 | 1.08 | [1.00-1.16] | 3.60E-02 | 1.08 | [1.00-1.16] | 3.60E-02 | 0.84 |

| | | | | | | | | | | | |
|---|---|---|---|---|---|---|---|---|---|---|---|
| siblings | rs2227483 | I3 | 9990 | 11 | 1.03 | [0.96-1.11] | 0.42 | 1.02 | [0.91-1.15] | 0.68 | 0.03 |
| siblings | rs2227483 | S1 | 13397 | 11 | 0.94 | [0.84-1.05] | 0.25 | 0.94 | [0.83-1.06] | 0.28 | 0.38 |
| siblings | rs2227483 | S2 | 15652 | 11 | 0.93 | [0.83-1.04] | 0.22 | 0.95 | [0.82-1.08] | 0.45 | 0.17 |
| siblings | rs2227483 | S3 | 9940 | 11 | 0.90 | [0.80-1.02] | 8.60E-02 | 0.90 | [0.76-1.06] | 0.2 | 0.16 |
| siblings | rs2228145 | I1 | 17111 | 9 | 1.03 | [0.93-1.15] | 0.56 | 1.03 | [0.93-1.15] | 0.56 | 0.70 |
| siblings | rs2228145 | I2 | 7508 | 9 | 1.00 | [0.93-1.09] | 0.91 | 1.00 | [0.93-1.09] | 0.91 | 0.73 |
| siblings | rs2228145 | I3 | 9603 | 9 | 1.04 | [0.97-1.12] | 0.26 | 1.04 | [0.97-1.12] | 0.26 | 0.72 |
| siblings | rs2228145 | S1 | 12847 | 9 | 1.03 | [0.92-1.16] | 0.56 | 1.03 | [0.92-1.16] | 0.56 | 0.43 |
| siblings | rs2228145 | S2 | 13638 | 9 | 0.99 | [0.88-1.11] | 0.82 | 0.99 | [0.88-1.11] | 0.82 | 0.85 |
| siblings | rs2228145 | S3 | 9390 | 9 | 0.98 | [0.87-1.11] | 0.76 | 0.98 | [0.87-1.11] | 0.76 | 0.58 |
| siblings | rs2897442 | I1 | 18623 | 10 | 0.98 | [0.87-1.11] | 0.78 | 0.98 | [0.85-1.13] | 0.76 | 0.29 |
| siblings | rs2897442 | I2 | 8914 | 10 | 1.12 | [1.03-1.21] | 8.50E-03 | 1.10 | [0.96-1.28] | 0.17 | 0.02 |
| siblings | rs2897442 | I3 | 9709 | 10 | 1.07 | [0.99-1.16] | 9.40E-02 | 1.07 | [0.99-1.16] | 9.40E-02 | 0.56 |
| siblings | rs2897442 | S1 | 12847 | 10 | 0.97 | [0.85-1.10] | 0.59 | 0.97 | [0.85-1.10] | 0.59 | 0.68 |
| siblings | rs2897442 | S2 | 15150 | 10 | 0.97 | [0.86-1.11] | 0.69 | 0.96 | [0.82-1.14] | 0.61 | 0.25 |
| siblings | rs2897442 | S3 | 9390 | 10 | 0.96 | [0.83-1.10] | 0.54 | 0.96 | [0.83-1.10] | 0.54 | 0.52 |
| siblings | rs479844 | I1 | 18630 | 10 | 0.98 | [0.88-1.09] | 0.71 | 1.00 | [0.87-1.12] | 0.99 | 0.24 |
| siblings | rs479844 | I2 | 8916 | 10 | 1.15 | [1.07-1.23] | 2.30E-04 | 1.15 | [1.02-1.29] | 1.90E-02 | 0.02 |
| siblings | rs479844 | I3 | 9714 | 10 | 1.15 | [1.07-1.24] | 2.00E-04 | 1.15 | [1.07-1.24] | 2.00E-04 | 0.45 |
| siblings | rs479844 | S1 | 12847 | 10 | 1.02 | [0.91-1.14] | 0.77 | 1.02 | [0.91-1.14] | 0.77 | 0.51 |
| siblings | rs479844 | S2 | 15157 | 10 | 0.96 | [0.86-1.08] | 0.5 | 0.97 | [0.85-1.10] | 0.66 | 0.28 |
| siblings | rs479844 | S3 | 9390 | 10 | 1.00 | [0.88-1.13] | 0.95 | 1.00 | [0.88-1.13] | 0.95 | 0.52 |
| siblings | rs6010620 | I1 | 18630 | 10 | 1.00 | [0.88-1.13] | 1 | 1.07 | [0.89-1.20] | 0.47 | 0.09 |
| siblings | rs6010620 | I2 | 8916 | 10 | 1.10 | [1.01-1.20] | 2.10E-02 | 1.10 | [1.01-1.20] | 2.10E-02 | 0.54 |
| siblings | rs6010620 | I3 | 9714 | 10 | 1.09 | [1.00-1.19] | 0.05 | 1.10 | [1.00-1.20] | 5.50E-02 | 0.32 |
| siblings | rs6010620 | S1 | 12847 | 10 | 0.96 | [0.84-1.10] | 0.52 | 1.01 | [0.83-1.16] | 0.92 | 0.13 |
| siblings | rs6010620 | S2 | 15157 | 10 | 0.99 | [0.87-1.13] | 0.89 | 1.05 | [0.87-1.20] | 0.61 | 0.11 |
| siblings | rs6010620 | S3 | 9390 | 10 | 0.94 | [0.81-1.09] | 0.4 | 0.97 | [0.80-1.14] | 0.8 | 0.18 |
| siblings | rs6473227 | I1 | 19173 | 11 | 0.92 | [0.83-1.02] | 0.12 | 0.92 | [0.83-1.03] | 0.15 | 0.42 |
| siblings | rs6473227 | I2 | 9151 | 11 | 1.09 | [1.02-1.17] | 1.40E-02 | 1.08 | [0.99-1.19] | 7.30E-02 | 0.27 |
| siblings | rs6473227 | I3 | 10022 | 11 | 1.03 | [0.96-1.11] | 0.37 | 1.03 | [0.96-1.11] | 0.37 | 0.78 |
| siblings | rs6473227 | S1 | 13403 | 11 | 0.90 | [0.81-1.01] | 8.20E-02 | 0.90 | [0.81-1.01] | 8.20E-02 | 0.72 |
| siblings | rs6473227 | S2 | 15700 | 11 | 0.91 | [0.81-1.02] | 0.11 | 0.94 | [0.81-1.06] | 0.44 | 0.14 |
| siblings | rs6473227 | S3 | 9946 | 11 | 0.89 | [0.79-1.01] | 6.60E-02 | 0.91 | [0.79-1.03] | 0.21 | 0.26 |
| siblings | rs7127307 | I1 | 19185 | 11 | 1.02 | [0.91-1.13] | 0.76 | 1.05 | [0.93-1.15] | 0.44 | 0.28 |
| siblings | rs7127307 | I2 | 9153 | 11 | 1.05 | [0.97-1.12] | 0.22 | 1.05 | [0.97-1.12] | 0.22 | 0.64 |
| siblings | rs7127307 | I3 | 10032 | 11 | 1.09 | [1.01-1.17] | 2.50E-02 | 1.09 | [1.01-1.17] | 2.50E-02 | 0.50 |
| siblings | rs7127307 | S1 | 13402 | 11 | 1.01 | [0.90-1.13] | 0.87 | 1.07 | [0.92-1.18] | 0.37 | 0.16 |
| siblings | rs7127307 | S2 | 15712 | 11 | 1.02 | [0.91-1.14] | 0.74 | 1.04 | [0.91-1.16] | 0.55 | 0.30 |
| siblings | rs7127307 | S3 | 9945 | 11 | 1.02 | [0.90-1.15] | 0.81 | 1.07 | [0.90-1.21] | 0.46 | 0.15 |
| siblings | rs7146581 | I1 | 19176 | 11 | 1.25 | [1.10-1.42] | 4.20E-04 | 1.24 | [1.08-1.44] | 3.10E-03 | 0.32 |
| siblings | rs7146581 | I2 | 9149 | 11 | 0.95 | [0.88-1.04] | 0.28 | 0.96 | [0.86-1.07] | 0.54 | 0.13 |
| siblings | rs7146581 | I3 | 10027 | 11 | 1.15 | [1.06-1.26] | 1.00E-03 | 1.14 | [1.03-1.28] | 1.10E-02 | 0.26 |
| siblings | rs7146581 | S1 | 13399 | 11 | 1.23 | [1.08-1.41] | 2.20E-03 | 1.20 | [1.01-1.46] | 3.40E-02 | 0.23 |
| siblings | rs7146581 | S2 | 15703 | 11 | 1.27 | [1.11-1.45] | 4.80E-04 | 1.24 | [1.04-1.52] | 1.90E-02 | 0.15 |
| siblings | rs7146581 | S3 | 9942 | 11 | 1.25 | [1.08-1.45] | 2.30E-03 | 1.20 | [0.98-1.54] | 0.08 | 0.12 |
| siblings | rs7927894 | I1 | 18263 | 10 | 1.13 | [1.01-1.26] | 3.10E-02 | 1.13 | [1.01-1.26] | 3.10E-02 | 0.53 |
| siblings | rs7927894 | I2 | 8829 | 10 | 0.96 | [0.90-1.04] | 0.33 | 0.95 | [0.86-1.07] | 0.36 | 0.09 |
| siblings | rs7927894 | I3 | 9434 | 10 | 1.09 | [1.02-1.18] | 1.60E-02 | 1.09 | [1.02-1.18] | 1.60E-02 | 0.94 |
| siblings | rs7927894 | S1 | 12847 | 10 | 1.12 | [0.99-1.25] | 6.20E-02 | 1.12 | [0.99-1.27] | 8.30E-02 | 0.35 |
| siblings | rs7927894 | S2 | 14790 | 10 | 1.11 | [0.99-1.24] | 8.40E-02 | 1.11 | [0.99-1.24] | 8.40E-02 | 0.84 |
| siblings | rs7927894 | S3 | 9390 | 10 | 1.10 | [0.97-1.25] | 0.14 | 1.10 | [0.97-1.25] | 0.14 | 0.72 |
| siblings | | M1 | 21045 | 11 | 1.00 | [0.93-1.07] | 0.99 | 1.00 | [0.93-1.08] | 0.96 | 0.38 |
| siblings | | M2 | 15241 | 11 | 0.99 | [0.92-1.06] | 0.75 | 0.99 | [0.92-1.06] | 0.75 | 0.53 |
| siblings | | M3 | 17383 | 11 | 0.98 | [0.91-1.05] | 0.51 | 0.97 | [0.89-1.06] | 0.52 | 0.31 |
| siblings | | M4 | 12821 | 10 | 0.98 | [0.91-1.06] | 0.58 | 0.98 | [0.90-1.06] | 0.58 | 0.36 |
| smoke_child | flg | I1 | 15618 | 12 | 1.33 | [1.05-1.68] | 1.80E-02 | 1.37 | [1.05-1.74] | 2.20E-02 | 0.36 |
| smoke_child | flg | I2 | 9567 | 12 | 1.73 | [1.50-1.99] | 2.00E-14 | 1.68 | [1.37-2.13] | 1.10E-06 | 0.07 |
| smoke_child | flg | I3 | 6051 | 12 | 2.15 | [1.80-2.56] | 1.10E-17 | 2.15 | [1.80-2.56] | 1.10E-17 | 0.63 |
| smoke_child | flg | S1 | 14259 | 12 | 1.31 | [1.02-1.69] | 3.40E-02 | 1.34 | [1.00-1.76] | 4.60E-02 | 0.35 |
| smoke_child | flg | S2 | 13896 | 12 | 1.39 | [1.08-1.81] | 1.20E-02 | 1.39 | [1.08-1.81] | 1.20E-02 | 0.53 |
| smoke_child | flg | S3 | 12562 | 12 | 1.39 | [1.05-1.83] | 2.10E-02 | 1.39 | [1.05-1.83] | 2.10E-02 | 0.47 |
| smoke_child | rs10214237 | I1 | 16519 | 12 | 1.00 | [0.89-1.13] | 0.99 | 1.00 | [0.89-1.13] | 0.99 | 0.62 |
| smoke_child | rs10214237 | I2 | 10252 | 12 | 1.11 | [1.04-1.19] | 2.80E-03 | 1.11 | [1.04-1.19] | 2.80E-03 | 0.70 |
| smoke_child | rs10214237 | I3 | 6267 | 12 | 1.09 | [1.00-1.20] | 6.20E-02 | 1.09 | [1.00-1.20] | 6.20E-02 | 0.69 |
| smoke_child | rs10214237 | S1 | 14942 | 12 | 1.00 | [0.88-1.14] | 0.99 | 1.00 | [0.88-1.14] | 0.99 | 0.46 |
| smoke_child | rs10214237 | S2 | 12681 | 12 | 1.01 | [0.89-1.16] | 0.85 | 1.01 | [0.89-1.16] | 0.85 | 0.72 |
| smoke_child | rs10214237 | S3 | 11123 | 12 | 1.00 | [0.87-1.16] | 0.96 | 1.00 | [0.87-1.16] | 0.96 | 0.67 |
| smoke_child | rs1057258 | I1 | 16502 | 12 | 0.99 | [0.86-1.15] | 0.92 | 0.99 | [0.82-1.20] | 0.96 | 0.16 |
| smoke_child | rs1057258 | I2 | 10244 | 12 | 1.05 | [0.97-1.14] | 0.25 | 1.05 | [0.97-1.14] | 0.25 | 0.45 |
| smoke_child | rs1057258 | I3 | 6258 | 12 | 1.02 | [0.91-1.14] | 0.71 | 1.02 | [0.91-1.14] | 0.71 | 0.62 |
| smoke_child | rs1057258 | S1 | 14943 | 12 | 0.98 | [0.84-1.14] | 0.8 | 0.98 | [0.80-1.21] | 0.87 | 0.16 |
| smoke_child | rs1057258 | S2 | 12664 | 12 | 0.96 | [0.82-1.13] | 0.65 | 0.95 | [0.79-1.16] | 0.6 | 0.29 |
| smoke_child | rs1057258 | S3 | 11124 | 12 | 0.95 | [0.80-1.12] | 0.53 | 0.94 | [0.79-1.13] | 0.51 | 0.40 |
| smoke_child | rs10995251 | I1 | 15928 | 11 | 0.99 | [0.88-1.11] | 0.88 | 0.99 | [0.88-1.11] | 0.88 | 0.44 |
| smoke_child | rs10995251 | I2 | 9868 | 11 | 1.03 | [0.96-1.10] | 0.38 | 1.05 | [0.96-1.12] | 0.28 | 0.16 |
| smoke_child | rs10995251 | I3 | 6060 | 11 | 1.03 | [0.94-1.12] | 0.53 | 1.04 | [0.91-1.18] | 0.6 | 0.04 |
| smoke_child | rs10995251 | S1 | 14353 | 11 | 1.01 | [0.89-1.14] | 0.86 | 1.00 | [0.85-1.19] | 0.97 | 0.19 |
| smoke_child | rs10995251 | S2 | 12090 | 11 | 0.98 | [0.86-1.11] | 0.7 | 0.97 | [0.85-1.12] | 0.69 | 0.36 |
| smoke_child | rs10995251 | S3 | 10534 | 11 | 0.99 | [0.87-1.13] | 0.89 | 0.98 | [0.82-1.19] | 0.86 | 0.14 |
| smoke_child | rs112111458 | I1 | 14664 | 9 | 1.04 | [0.88-1.22] | 0.65 | 1.04 | [0.88-1.22] | 0.65 | 0.95 |
| smoke_child | rs112111458 | I2 | 9251 | 9 | 0.98 | [0.90-1.08] | 0.74 | 1.01 | [0.90-1.11] | 0.91 | 0.18 |
| smoke_child | rs112111458 | I3 | 5413 | 9 | 1.05 | [0.92-1.19] | 0.45 | 1.05 | [0.92-1.19] | 0.45 | 0.75 |
| smoke_child | rs112111458 | S1 | 13997 | 9 | 1.07 | [0.90-1.26] | 0.47 | 1.07 | [0.90-1.26] | 0.47 | 0.98 |
| smoke_child | rs112111458 | S2 | 11770 | 9 | 1.04 | [0.88-1.23] | 0.65 | 1.04 | [0.88-1.23] | 0.65 | 0.90 |
| smoke_child | rs112111458 | S3 | 11122 | 9 | 1.06 | [0.89-1.27] | 0.51 | 1.06 | [0.89-1.27] | 0.51 | 0.93 |
| smoke_child | rs11657987 | I1 | 16487 | 12 | 1.06 | [0.95-1.18] | 0.32 | 1.06 | [0.95-1.18] | 0.32 | 0.59 |
| smoke_child | rs11657987 | I2 | 10237 | 12 | 1.03 | [0.97-1.10] | 0.32 | 1.03 | [0.95-1.12] | 0.49 | 0.11 |
| smoke_child | rs11657987 | I3 | 6250 | 12 | 1.09 | [1.00-1.18] | 4.80E-02 | 1.09 | [1.00-1.18] | 4.80E-02 | 0.59 |
| smoke_child | rs11657987 | S1 | 14939 | 12 | 1.09 | [0.97-1.22] | 0.15 | 1.09 | [0.97-1.22] | 0.15 | 0.54 |
| smoke_child | rs11657987 | S2 | 12649 | 12 | 1.05 | [0.93-1.18] | 0.43 | 1.05 | [0.93-1.18] | 0.43 | 0.48 |

| | | | | | | | | | | | |
|---|---|---|---|---|---|---|---|---|---|---|---|
| smoke_child | rs11657987 | S3 | 11120 | 12 | 1.08 | [0.95-1.22] | 0.24 | 1.08 | [0.94-1.25] | 0.29 | 0.31 |
| smoke_child | rs12153855 | I1 | 14096 | 8 | 0.97 | [0.80-1.18] | 0.76 | 0.97 | [0.80-1.18] | 0.76 | 0.90 |
| smoke_child | rs12153855 | I2 | 8879 | 8 | 1.11 | [0.99-1.24] | 7.20E-02 | 1.11 | [0.99-1.24] | 7.20E-02 | 0.84 |
| smoke_child | rs12153855 | I3 | 5217 | 8 | 1.09 | [0.93-1.26] | 0.29 | 1.09 | [0.93-1.26] | 0.29 | 0.91 |
| smoke_child | rs12153855 | S1 | 13409 | 8 | 0.95 | [0.77-1.17] | 0.62 | 0.95 | [0.77-1.17] | 0.62 | 0.83 |
| smoke_child | rs12153855 | S2 | 11202 | 8 | 0.95 | [0.78-1.17] | 0.64 | 0.95 | [0.78-1.17] | 0.64 | 0.77 |
| smoke_child | rs12153855 | S3 | 10534 | 8 | 0.94 | [0.75-1.16] | 0.55 | 0.94 | [0.75-1.16] | 0.55 | 0.64 |
| smoke_child | rs12295535 | I1 | 16523 | 12 | 1.02 | [0.73-1.42] | 0.91 | 0.89 | [0.56-1.62] | 0.61 | 0.16 |
| smoke_child | rs12295535 | I2 | 10254 | 12 | 1.15 | [0.96-1.39] | 0.13 | 1.20 | [0.95-1.46] | 0.13 | 0.19 |
| smoke_child | rs12295535 | I3 | 6269 | 12 | 1.17 | [0.90-1.51] | 0.24 | 1.12 | [0.84-1.57] | 0.44 | 0.34 |
| smoke_child | rs12295535 | S1 | 14943 | 12 | 0.98 | [0.67-1.42] | 0.92 | 0.77 | [0.41-1.86] | 0.43 | 0.03 |
| smoke_child | rs12295535 | S2 | 12685 | 12 | 1.10 | [0.76-1.60] | 0.6 | 1.10 | [0.76-1.60] | 0.6 | 0.58 |
| smoke_child | rs12295535 | S3 | 11124 | 12 | 1.11 | [0.73-1.67] | 0.62 | 1.04 | [0.63-1.83] | 0.87 | 0.26 |
| smoke_child | rs13015714 | I1 | 15931 | 11 | 0.94 | [0.82-1.07] | 0.33 | 0.94 | [0.82-1.07] | 0.33 | 0.57 |
| smoke_child | rs13015714 | I2 | 9870 | 11 | 1.14 | [1.06-1.22] | 6.20E-04 | 1.14 | [1.06-1.22] | 6.20E-04 | 0.53 |
| smoke_child | rs13015714 | I3 | 6061 | 11 | 1.08 | [0.98-1.19] | 0.12 | 1.08 | [0.98-1.19] | 0.12 | 0.67 |
| smoke_child | rs13015714 | S1 | 14351 | 11 | 0.96 | [0.84-1.10] | 0.57 | 0.93 | [0.78-1.15] | 0.44 | 0.19 |
| smoke_child | rs13015714 | S2 | 12093 | 11 | 0.95 | [0.83-1.09] | 0.48 | 0.95 | [0.83-1.09] | 0.48 | 0.53 |
| smoke_child | rs13015714 | S3 | 10532 | 11 | 0.97 | [0.84-1.13] | 0.73 | 0.93 | [0.75-1.20] | 0.48 | 0.14 |
| smoke_child | rs16948048 | I1 | 15932 | 11 | 1.02 | [0.91-1.14] | 0.8 | 1.01 | [0.90-1.14] | 0.8 | 0.43 |
| smoke_child | rs16948048 | I2 | 9871 | 11 | 1.03 | [0.96-1.10] | 0.42 | 1.03 | [0.95-1.11] | 0.46 | 0.27 |
| smoke_child | rs16948048 | I3 | 6061 | 11 | 1.04 | [0.95-1.13] | 0.37 | 1.04 | [0.95-1.13] | 0.37 | 0.82 |
| smoke_child | rs16948048 | S1 | 14353 | 11 | 0.99 | [0.88-1.12] | 0.93 | 0.99 | [0.88-1.12] | 0.93 | 0.70 |
| smoke_child | rs16948048 | S2 | 12094 | 11 | 0.99 | [0.88-1.12] | 0.9 | 0.99 | [0.88-1.12] | 0.9 | 0.60 |
| smoke_child | rs16948048 | S3 | 10534 | 11 | 0.96 | [0.84-1.10] | 0.57 | 0.96 | [0.84-1.10] | 0.57 | 0.85 |
| smoke_child | rs17389644 | I1 | 15922 | 11 | 0.91 | [0.80-1.04] | 0.18 | 0.91 | [0.80-1.04] | 0.18 | 0.54 |
| smoke_child | rs17389644 | I2 | 9865 | 11 | 1.09 | [1.01-1.17] | 3.30E-02 | 1.07 | [0.96-1.21] | 0.2 | 0.10 |
| smoke_child | rs17389644 | I3 | 6057 | 11 | 1.01 | [0.91-1.11] | 0.91 | 1.01 | [0.91-1.11] | 0.91 | 0.53 |
| smoke_child | rs17389644 | S1 | 14353 | 11 | 0.94 | [0.82-1.09] | 0.43 | 0.93 | [0.80-1.10] | 0.39 | 0.36 |
| smoke_child | rs17389644 | S2 | 12084 | 11 | 0.91 | [0.79-1.05] | 0.2 | 0.91 | [0.79-1.05] | 0.2 | 0.66 |
| smoke_child | rs17389644 | S3 | 10534 | 11 | 0.95 | [0.81-1.11] | 0.5 | 0.95 | [0.81-1.11] | 0.5 | 0.50 |
| smoke_child | rs17881320 | I1 | 16505 | 12 | 0.89 | [0.73-1.09] | 0.25 | 0.89 | [0.73-1.09] | 0.25 | 0.83 |
| smoke_child | rs17881320 | I2 | 10243 | 12 | 1.14 | [1.02-1.28] | 2.30E-02 | 1.14 | [0.98-1.33] | 8.40E-02 | 0.13 |
| smoke_child | rs17881320 | I3 | 6262 | 12 | 1.02 | [0.87-1.18] | 0.83 | 1.02 | [0.87-1.18] | 0.83 | 0.95 |
| smoke_child | rs17881320 | S1 | 14941 | 12 | 0.92 | [0.74-1.14] | 0.44 | 0.92 | [0.74-1.14] | 0.44 | 0.72 |
| smoke_child | rs17881320 | S2 | 12667 | 12 | 0.84 | [0.67-1.05] | 0.13 | 0.84 | [0.67-1.05] | 0.13 | 0.57 |
| smoke_child | rs17881320 | S3 | 11122 | 12 | 0.88 | [0.69-1.11] | 0.28 | 0.88 | [0.69-1.11] | 0.28 | 0.48 |
| smoke_child | rs2041733 | I1 | 15932 | 11 | 1.03 | [0.92-1.15] | 0.66 | 1.03 | [0.92-1.15] | 0.66 | 0.70 |
| smoke_child | rs2041733 | I2 | 9871 | 11 | 1.03 | [0.97-1.10] | 0.3 | 1.03 | [0.97-1.10] | 0.3 | 0.91 |
| smoke_child | rs2041733 | I3 | 6061 | 11 | 1.04 | [0.96-1.13] | 0.35 | 1.04 | [0.96-1.13] | 0.35 | 0.54 |
| smoke_child | rs2041733 | S1 | 14352 | 11 | 1.01 | [0.90-1.14] | 0.82 | 1.01 | [0.90-1.14] | 0.82 | 0.68 |
| smoke_child | rs2041733 | S2 | 12094 | 11 | 1.03 | [0.92-1.17] | 0.59 | 1.03 | [0.92-1.17] | 0.59 | 0.75 |
| smoke_child | rs2041733 | S3 | 10533 | 11 | 1.02 | [0.89-1.16] | 0.81 | 1.02 | [0.89-1.16] | 0.81 | 0.63 |
| smoke_child | rs2143950 | I1 | 16510 | 12 | 1.00 | [0.87-1.16] | 0.97 | 1.00 | [0.87-1.16] | 0.97 | 0.91 |
| smoke_child | rs2143950 | I2 | 10242 | 12 | 1.05 | [0.97-1.14] | 0.26 | 1.05 | [0.97-1.14] | 0.26 | 0.90 |
| smoke_child | rs2143950 | I3 | 6268 | 12 | 1.04 | [0.93-1.16] | 0.5 | 1.04 | [0.93-1.16] | 0.5 | 0.63 |
| smoke_child | rs2143950 | S1 | 14934 | 12 | 0.99 | [0.85-1.15] | 0.85 | 0.99 | [0.85-1.15] | 0.85 | 0.84 |
| smoke_child | rs2143950 | S2 | 12672 | 12 | 1.03 | [0.88-1.20] | 0.74 | 1.03 | [0.88-1.20] | 0.74 | 0.74 |
| smoke_child | rs2143950 | S3 | 11115 | 12 | 1.02 | [0.86-1.20] | 0.82 | 1.02 | [0.86-1.20] | 0.82 | 0.59 |
| smoke_child | rs2164983 | I1 | 14096 | 8 | 0.97 | [0.83-1.13] | 0.68 | 0.97 | [0.83-1.13] | 0.68 | 0.97 |
| smoke_child | rs2164983 | I2 | 8879 | 8 | 1.14 | [1.04-1.25] | 4.50E-03 | 1.14 | [1.04-1.25] | 4.50E-03 | 0.81 |
| smoke_child | rs2164983 | I3 | 5217 | 8 | 1.13 | [1.00-1.28] | 5.80E-02 | 1.13 | [1.00-1.28] | 5.80E-02 | 0.51 |
| smoke_child | rs2164983 | S1 | 13409 | 8 | 0.98 | [0.83-1.17] | 0.85 | 0.98 | [0.83-1.17] | 0.85 | 0.95 |
| smoke_child | rs2164983 | S2 | 11202 | 8 | 0.96 | [0.81-1.14] | 0.64 | 0.96 | [0.81-1.14] | 0.64 | 0.95 |
| smoke_child | rs2164983 | S3 | 10534 | 8 | 0.98 | [0.82-1.17] | 0.8 | 0.98 | [0.82-1.17] | 0.8 | 0.92 |
| smoke_child | rs2227483 | I1 | 16512 | 12 | 1.00 | [0.90-1.12] | 0.99 | 1.00 | [0.90-1.12] | 0.99 | 0.49 |
| smoke_child | rs2227483 | I2 | 10248 | 12 | 1.06 | [0.99-1.13] | 8.30E-02 | 1.06 | [0.99-1.13] | 8.30E-02 | 0.90 |
| smoke_child | rs2227483 | I3 | 6264 | 12 | 1.05 | [0.96-1.14] | 0.29 | 1.03 | [0.91-1.19] | 0.65 | 0.03 |
| smoke_child | rs2227483 | S1 | 14936 | 12 | 0.95 | [0.85-1.07] | 0.43 | 0.95 | [0.85-1.07] | 0.43 | 0.49 |
| smoke_child | rs2227483 | S2 | 12674 | 12 | 1.03 | [0.91-1.15] | 0.67 | 1.03 | [0.91-1.15] | 0.67 | 0.81 |
| smoke_child | rs2227483 | S3 | 11117 | 12 | 0.98 | [0.87-1.12] | 0.79 | 0.98 | [0.87-1.12] | 0.79 | 0.68 |
| smoke_child | rs2228145 | I1 | 15040 | 10 | 0.96 | [0.86-1.08] | 0.54 | 0.96 | [0.86-1.08] | 0.54 | 0.65 |
| smoke_child | rs2228145 | I2 | 9137 | 10 | 1.05 | [0.99-1.13] | 0.12 | 1.05 | [0.99-1.13] | 0.12 | 0.55 |
| smoke_child | rs2228145 | I3 | 5903 | 10 | 1.01 | [0.93-1.10] | 0.81 | 1.01 | [0.93-1.10] | 0.81 | 0.52 |
| smoke_child | rs2228145 | S1 | 14353 | 10 | 0.96 | [0.85-1.08] | 0.51 | 0.96 | [0.85-1.08] | 0.51 | 0.63 |
| smoke_child | rs2228145 | S2 | 11202 | 10 | 0.99 | [0.88-1.12] | 0.89 | 0.99 | [0.88-1.12] | 0.89 | 0.78 |
| smoke_child | rs2228145 | S3 | 10534 | 10 | 0.98 | [0.86-1.12] | 0.8 | 0.98 | [0.86-1.12] | 0.8 | 0.60 |
| smoke_child | rs2897442 | I1 | 14631 | 10 | 0.96 | [0.85-1.10] | 0.59 | 0.96 | [0.85-1.10] | 0.59 | 0.44 |
| smoke_child | rs2897442 | I2 | 9136 | 10 | 1.10 | [1.02-1.19] | 0.01 | 1.09 | [0.99-1.22] | 9.30E-02 | 0.15 |
| smoke_child | rs2897442 | I3 | 5495 | 10 | 1.09 | [0.98-1.20] | 0.1 | 1.10 | [0.96-1.24] | 0.18 | 0.16 |
| smoke_child | rs2897442 | S1 | 13052 | 10 | 0.93 | [0.81-1.07] | 0.3 | 0.93 | [0.80-1.08] | 0.38 | 0.37 |
| smoke_child | rs2897442 | S2 | 10793 | 10 | 1.01 | [0.87-1.16] | 0.92 | 1.01 | [0.87-1.17] | 0.86 | 0.38 |
| smoke_child | rs2897442 | S3 | 9233 | 10 | 0.97 | [0.83-1.14] | 0.72 | 0.99 | [0.81-1.18] | 0.92 | 0.23 |
| smoke_child | rs479844 | I1 | 14632 | 10 | 0.97 | [0.86-1.09] | 0.57 | 0.96 | [0.82-1.14] | 0.66 | 0.13 |
| smoke_child | rs479844 | I2 | 9136 | 10 | 1.15 | [1.07-1.23] | 4.50E-05 | 1.15 | [1.06-1.24] | 4.20E-04 | 0.30 |
| smoke_child | rs479844 | I3 | 5496 | 10 | 1.12 | [1.03-1.23] | 1.20E-02 | 1.12 | [0.99-1.27] | 6.10E-02 | 0.14 |
| smoke_child | rs479844 | S1 | 13052 | 10 | 0.94 | [0.83-1.07] | 0.36 | 0.93 | [0.78-1.12] | 0.39 | 0.19 |
| smoke_child | rs479844 | S2 | 10794 | 10 | 1.01 | [0.89-1.15] | 0.83 | 1.01 | [0.89-1.15] | 0.83 | 0.52 |
| smoke_child | rs479844 | S3 | 9233 | 10 | 0.99 | [0.86-1.14] | 0.89 | 0.99 | [0.86-1.14] | 0.89 | 0.76 |
| smoke_child | rs6010620 | I1 | 15933 | 11 | 0.90 | [0.79-1.02] | 0.1 | 0.90 | [0.79-1.02] | 0.1 | 0.78 |
| smoke_child | rs6010620 | I2 | 9871 | 11 | 1.16 | [1.08-1.25] | 9.90E-05 | 1.16 | [1.08-1.25] | 9.90E-05 | 1.00 |
| smoke_child | rs6010620 | I3 | 6062 | 11 | 1.04 | [0.94-1.15] | 0.43 | 1.04 | [0.94-1.15] | 0.43 | 0.69 |
| smoke_child | rs6010620 | S1 | 14353 | 11 | 0.91 | [0.79-1.04] | 0.17 | 0.91 | [0.79-1.04] | 0.17 | 0.61 |
| smoke_child | rs6010620 | S2 | 12095 | 11 | 0.87 | [0.76-1.01] | 6.30E-02 | 0.87 | [0.76-1.01] | 6.30E-02 | 0.70 |
| smoke_child | rs6010620 | S3 | 10534 | 11 | 0.88 | [0.75-1.02] | 0.09 | 0.88 | [0.75-1.02] | 0.09 | 0.42 |
| smoke_child | rs6473227 | I1 | 16523 | 12 | 1.03 | [0.92-1.15] | 0.59 | 1.03 | [0.92-1.15] | 0.59 | 0.61 |
| smoke_child | rs6473227 | I2 | 10254 | 12 | 1.04 | [0.98-1.11] | 0.21 | 1.04 | [0.98-1.11] | 0.21 | 0.84 |
| smoke_child | rs6473227 | I3 | 6269 | 12 | 1.09 | [1.00-1.18] | 4.60E-02 | 1.09 | [1.00-1.18] | 4.60E-02 | 0.63 |
| smoke_child | rs6473227 | S1 | 14943 | 12 | 0.99 | [0.88-1.11] | 0.87 | 0.99 | [0.88-1.11] | 0.87 | 0.89 |
| smoke_child | rs6473227 | S2 | 12685 | 12 | 1.01 | [0.90-1.14] | 0.81 | 1.01 | [0.90-1.14] | 0.81 | 0.50 |
| smoke_child | rs6473227 | S3 | 11124 | 12 | 0.97 | [0.85-1.10] | 0.63 | 0.97 | [0.85-1.10] | 0.63 | 0.90 |

| | | | | | | | | | | | |
|---|---|---|---|---|---|---|---|---|---|---|---|
| smoke_child | rs7127307 | I1 | 16522 | 12 | 0.99 | [0.89-1.11] | 0.88 | 0.99 | [0.89-1.11] | 0.88 | 0.93 |
| smoke_child | rs7127307 | I2 | 10253 | 12 | 1.04 | [0.98-1.11] | 0.21 | 1.04 | [0.98-1.11] | 0.21 | 1.00 |
| smoke_child | rs7127307 | I3 | 6269 | 12 | 1.06 | [0.98-1.16] | 0.15 | 1.06 | [0.98-1.16] | 0.15 | 0.45 |
| smoke_child | rs7127307 | S1 | 14942 | 12 | 1.01 | [0.90-1.14] | 0.85 | 1.01 | [0.90-1.14] | 0.85 | 0.95 |
| smoke_child | rs7127307 | S2 | 12684 | 12 | 0.98 | [0.87-1.11] | 0.75 | 0.98 | [0.87-1.11] | 0.75 | 0.78 |
| smoke_child | rs7127307 | S3 | 11123 | 12 | 1.01 | [0.88-1.15] | 0.94 | 1.01 | [0.88-1.15] | 0.94 | 0.82 |
| smoke_child | rs7146581 | I1 | 16515 | 12 | 0.97 | [0.85-1.11] | 0.67 | 0.97 | [0.85-1.11] | 0.67 | 0.74 |
| smoke_child | rs7146581 | I2 | 10249 | 12 | 1.04 | [0.97-1.12] | 0.3 | 1.04 | [0.97-1.12] | 0.3 | 0.53 |
| smoke_child | rs7146581 | I3 | 6266 | 12 | 1.01 | [0.92-1.11] | 0.85 | 1.01 | [0.92-1.11] | 0.85 | 0.89 |
| smoke_child | rs7146581 | S1 | 14937 | 12 | 0.97 | [0.84-1.11] | 0.65 | 0.97 | [0.84-1.11] | 0.65 | 0.61 |
| smoke_child | rs7146581 | S2 | 12677 | 12 | 1.00 | [0.87-1.14] | 0.95 | 1.00 | [0.87-1.14] | 0.95 | 0.76 |
| smoke_child | rs7146581 | S3 | 11118 | 12 | 0.98 | [0.85-1.14] | 0.81 | 0.98 | [0.85-1.14] | 0.81 | 0.63 |
| smoke_child | rs7927894 | I1 | 15892 | 11 | 0.99 | [0.89-1.11] | 0.9 | 0.99 | [0.88-1.12] | 0.86 | 0.39 |
| smoke_child | rs7927894 | I2 | 9846 | 11 | 1.02 | [0.96-1.09] | 0.5 | 1.02 | [0.95-1.10] | 0.53 | 0.36 |
| smoke_child | rs7927894 | I3 | 6046 | 11 | 1.01 | [0.93-1.10] | 0.82 | 1.00 | [0.90-1.12] | 0.96 | 0.24 |
| smoke_child | rs7927894 | S1 | 14353 | 11 | 0.99 | [0.88-1.12] | 0.92 | 0.99 | [0.88-1.12] | 0.92 | 0.67 |
| smoke_child | rs7927894 | S2 | 12054 | 11 | 0.99 | [0.88-1.12] | 0.92 | 0.99 | [0.87-1.14] | 0.93 | 0.35 |
| smoke_child | rs7927894 | S3 | 10534 | 11 | 0.99 | [0.87-1.13] | 0.88 | 0.99 | [0.87-1.13] | 0.88 | 0.79 |
| smoke_child | | M1 | 18722 | 12 | 0.93 | [0.87-1.00] | 3.80E-02 | 0.93 | [0.86-1.00] | 4.90E-02 | 0.39 |
| smoke_child | | M2 | 17120 | 12 | 0.92 | [0.85-0.99] | 2.70E-02 | 0.92 | [0.85-0.99] | 2.70E-02 | 0.87 |
| smoke_child | | M3 | 15850 | 12 | 0.93 | [0.86-1.00] | 4.20E-02 | 0.92 | [0.84-1.01] | 8.20E-02 | 0.21 |
| smoke_child | | M4 | 14273 | 12 | 0.92 | [0.85-0.99] | 2.90E-02 | 0.92 | [0.85-0.99] | 2.90E-02 | 0.72 |
| smoke_utero | flg | I1 | 15669 | 12 | 1.24 | [0.95-1.62] | 0.11 | 1.24 | [0.95-1.62] | 0.11 | 0.88 |
| smoke_utero | flg | I2 | 11691 | 12 | 1.85 | [1.63-2.09] | 8.80E-22 | 1.80 | [1.47-2.26] | 1.30E-08 | 0.03 |
| smoke_utero | flg | I3 | 3978 | 12 | 2.12 | [1.70-2.65] | 3.70E-11 | 2.12 | [1.70-2.65] | 3.70E-11 | 0.96 |
| smoke_utero | flg | S1 | 13769 | 12 | 1.28 | [0.96-1.71] | 9.80E-02 | 1.28 | [0.96-1.71] | 9.80E-02 | 0.74 |
| smoke_utero | flg | S2 | 13884 | 12 | 1.29 | [0.96-1.73] | 9.60E-02 | 1.29 | [0.96-1.73] | 9.60E-02 | 0.81 |
| smoke_utero | flg | S3 | 12011 | 12 | 1.31 | [0.94-1.82] | 0.11 | 1.31 | [0.94-1.82] | 0.11 | 0.62 |
| smoke_utero | rs10214237 | I1 | 16650 | 12 | 1.12 | [0.98-1.28] | 9.70E-02 | 1.12 | [0.98-1.28] | 9.70E-02 | 0.82 |
| smoke_utero | rs10214237 | I2 | 12472 | 12 | 1.08 | [1.01-1.15] | 2.20E-02 | 1.08 | [1.01-1.15] | 2.20E-02 | 0.98 |
| smoke_utero | rs10214237 | I3 | 4178 | 12 | 1.19 | [1.07-1.34] | 1.80E-03 | 1.19 | [1.07-1.34] | 1.80E-03 | 0.66 |
| smoke_utero | rs10214237 | S1 | 14429 | 12 | 1.11 | [0.96-1.29] | 0.17 | 1.11 | [0.96-1.29] | 0.17 | 0.78 |
| smoke_utero | rs10214237 | S2 | 12784 | 12 | 1.12 | [0.97-1.30] | 0.13 | 1.12 | [0.97-1.30] | 0.13 | 0.74 |
| smoke_utero | rs10214237 | S3 | 10581 | 12 | 1.11 | [0.94-1.30] | 0.23 | 1.11 | [0.94-1.30] | 0.23 | 0.75 |
| smoke_utero | rs1057258 | I1 | 16633 | 12 | 1.10 | [0.94-1.29] | 0.22 | 1.10 | [0.94-1.29] | 0.22 | 0.91 |
| smoke_utero | rs1057258 | I2 | 12460 | 12 | 1.03 | [0.95-1.11] | 0.46 | 1.03 | [0.95-1.11] | 0.46 | 0.85 |
| smoke_utero | rs1057258 | I3 | 4173 | 12 | 1.11 | [0.98-1.27] | 0.11 | 1.11 | [0.98-1.27] | 0.11 | 0.91 |
| smoke_utero | rs1057258 | S1 | 14430 | 12 | 1.13 | [0.95-1.34] | 0.15 | 1.13 | [0.95-1.34] | 0.15 | 0.67 |
| smoke_utero | rs1057258 | S2 | 12767 | 12 | 1.09 | [0.92-1.30] | 0.3 | 1.09 | [0.92-1.30] | 0.3 | 0.73 |
| smoke_utero | rs1057258 | S3 | 10582 | 12 | 1.12 | [0.93-1.35] | 0.23 | 1.12 | [0.93-1.36] | 0.24 | 0.40 |
| smoke_utero | rs10995251 | I1 | 16059 | 11 | 1.02 | [0.90-1.16] | 0.75 | 1.02 | [0.90-1.16] | 0.75 | 0.59 |
| smoke_utero | rs10995251 | I2 | 11982 | 11 | 1.00 | [0.94-1.07] | 0.9 | 1.00 | [0.94-1.07] | 0.9 | 0.45 |
| smoke_utero | rs10995251 | I3 | 4077 | 11 | 1.06 | [0.95-1.18] | 0.28 | 1.06 | [0.94-1.19] | 0.38 | 0.32 |
| smoke_utero | rs10995251 | S1 | 13840 | 11 | 1.04 | [0.90-1.19] | 0.61 | 1.04 | [0.90-1.19] | 0.61 | 0.70 |
| smoke_utero | rs10995251 | S2 | 12193 | 11 | 1.02 | [0.89-1.18] | 0.76 | 1.02 | [0.89-1.18] | 0.76 | 0.49 |
| smoke_utero | rs10995251 | S3 | 9992 | 11 | 1.03 | [0.88-1.20] | 0.74 | 1.03 | [0.88-1.20] | 0.74 | 0.60 |
| smoke_utero | rs112111458 | I1 | 14154 | 9 | 1.14 | [0.95-1.36] | 0.16 | 1.14 | [0.95-1.36] | 0.16 | 0.45 |
| smoke_utero | rs112111458 | I2 | 10597 | 9 | 0.96 | [0.88-1.05] | 0.4 | 0.99 | [0.87-1.10] | 0.93 | 0.10 |
| smoke_utero | rs112111458 | I3 | 3557 | 9 | 1.11 | [0.96-1.29] | 0.16 | 1.11 | [0.96-1.29] | 0.16 | 0.65 |
| smoke_utero | rs112111458 | S1 | 13463 | 9 | 1.13 | [0.94-1.37] | 0.19 | 1.13 | [0.94-1.37] | 0.19 | 0.50 |
| smoke_utero | rs112111458 | S2 | 11253 | 9 | 1.17 | [0.97-1.42] | 9.20E-02 | 1.17 | [0.97-1.42] | 9.30E-02 | 0.42 |
| smoke_utero | rs112111458 | S3 | 10580 | 9 | 1.16 | [0.95-1.41] | 0.15 | 1.16 | [0.95-1.41] | 0.15 | 0.40 |
| smoke_utero | rs11657987 | I1 | 16616 | 12 | 0.95 | [0.84-1.07] | 0.41 | 0.95 | [0.84-1.07] | 0.41 | 0.61 |
| smoke_utero | rs11657987 | I2 | 12448 | 12 | 1.06 | [1.00-1.13] | 3.60E-02 | 1.06 | [0.97-1.15] | 0.19 | 0.09 |
| smoke_utero | rs11657987 | I3 | 4168 | 12 | 1.00 | [0.91-1.11] | 0.98 | 1.00 | [0.91-1.11] | 0.98 | 0.46 |
| smoke_utero | rs11657987 | S1 | 14426 | 12 | 0.94 | [0.82-1.07] | 0.34 | 0.94 | [0.82-1.07] | 0.34 | 0.62 |
| smoke_utero | rs11657987 | S2 | 12750 | 12 | 0.96 | [0.84-1.09] | 0.52 | 0.96 | [0.84-1.09] | 0.52 | 0.89 |
| smoke_utero | rs11657987 | S3 | 10578 | 12 | 0.94 | [0.81-1.08] | 0.36 | 0.94 | [0.81-1.08] | 0.36 | 0.68 |
| smoke_utero | rs12153855 | I1 | 13586 | 8 | 0.94 | [0.75-1.17] | 0.57 | 0.94 | [0.75-1.17] | 0.57 | 0.97 |
| smoke_utero | rs12153855 | I2 | 10121 | 8 | 1.14 | [1.02-1.27] | 1.90E-02 | 1.14 | [1.02-1.27] | 1.90E-02 | 0.70 |
| smoke_utero | rs12153855 | I3 | 3465 | 8 | 1.05 | [0.88-1.26] | 0.56 | 1.05 | [0.88-1.26] | 0.56 | 0.99 |
| smoke_utero | rs12153855 | S1 | 12875 | 8 | 0.93 | [0.74-1.18] | 0.55 | 0.93 | [0.74-1.18] | 0.55 | 0.89 |
| smoke_utero | rs12153855 | S2 | 10685 | 8 | 0.91 | [0.73-1.15] | 0.45 | 0.91 | [0.73-1.15] | 0.45 | 0.95 |
| smoke_utero | rs12153855 | S3 | 9992 | 8 | 0.91 | [0.72-1.17] | 0.48 | 0.91 | [0.72-1.17] | 0.48 | 0.81 |
| smoke_utero | rs12295535 | I1 | 16654 | 12 | 1.01 | [0.70-1.47] | 0.95 | 1.01 | [0.70-1.47] | 0.95 | 0.96 |
| smoke_utero | rs12295535 | I2 | 12475 | 12 | 1.14 | [0.96-1.35] | 0.13 | 1.14 | [0.96-1.35] | 0.13 | 0.69 |
| smoke_utero | rs12295535 | I3 | 4179 | 12 | 1.15 | [0.84-1.56] | 0.39 | 1.15 | [0.84-1.56] | 0.39 | 0.95 |
| smoke_utero | rs12295535 | S1 | 14430 | 12 | 0.94 | [0.62-1.44] | 0.79 | 0.94 | [0.62-1.44] | 0.79 | 0.88 |
| smoke_utero | rs12295535 | S2 | 12788 | 12 | 1.09 | [0.72-1.64] | 0.68 | 1.09 | [0.72-1.64] | 0.68 | 1.00 |
| smoke_utero | rs12295535 | S3 | 10582 | 12 | 1.09 | [0.68-1.73] | 0.73 | 1.09 | [0.68-1.73] | 0.73 | 0.96 |
| smoke_utero | rs13015714 | I1 | 16062 | 11 | 0.92 | [0.79-1.06] | 0.23 | 0.92 | [0.78-1.08] | 0.3 | 0.30 |
| smoke_utero | rs13015714 | I2 | 11983 | 11 | 1.13 | [1.06-1.21] | 4.70E-04 | 1.13 | [1.03-1.25] | 1.10E-02 | 0.10 |
| smoke_utero | rs13015714 | I3 | 4079 | 11 | 1.08 | [0.96-1.22] | 0.21 | 1.08 | [0.96-1.22] | 0.21 | 0.71 |
| smoke_utero | rs13015714 | S1 | 13838 | 11 | 0.94 | [0.80-1.10] | 0.43 | 0.94 | [0.78-1.13] | 0.49 | 0.29 |
| smoke_utero | rs13015714 | S2 | 12196 | 11 | 0.93 | [0.80-1.09] | 0.37 | 0.93 | [0.76-1.13] | 0.46 | 0.18 |
| smoke_utero | rs13015714 | S3 | 9990 | 11 | 0.96 | [0.81-1.14] | 0.61 | 0.95 | [0.75-1.21] | 0.67 | 0.13 |
| smoke_utero | rs16948048 | I1 | 16062 | 11 | 1.15 | [1.02-1.31] | 2.80E-02 | 1.15 | [1.02-1.31] | 2.80E-02 | 0.53 |
| smoke_utero | rs16948048 | I2 | 11984 | 11 | 0.99 | [0.93-1.05] | 0.7 | 0.99 | [0.93-1.06] | 0.82 | 0.33 |
| smoke_utero | rs16948048 | I3 | 4078 | 11 | 1.13 | [1.02-1.25] | 2.50E-02 | 1.13 | [1.02-1.25] | 2.50E-02 | 0.74 |
| smoke_utero | rs16948048 | S1 | 13840 | 11 | 1.15 | [1.00-1.32] | 4.50E-02 | 1.15 | [1.00-1.33] | 5.30E-02 | 0.41 |
| smoke_utero | rs16948048 | S2 | 12196 | 11 | 1.17 | [1.02-1.34] | 2.70E-02 | 1.17 | [1.02-1.34] | 2.70E-02 | 0.57 |
| smoke_utero | rs16948048 | S3 | 9992 | 11 | 1.16 | [1.00-1.35] | 5.30E-02 | 1.16 | [0.98-1.38] | 8.20E-02 | 0.31 |
| smoke_utero | rs17389644 | I1 | 16051 | 11 | 0.96 | [0.83-1.11] | 0.59 | 0.96 | [0.83-1.11] | 0.59 | 0.72 |
| smoke_utero | rs17389644 | I2 | 11976 | 11 | 1.07 | [0.99-1.14] | 7.00E-02 | 1.05 | [0.97-1.16] | 0.25 | 0.29 |
| smoke_utero | rs17389644 | I3 | 4075 | 11 | 0.99 | [0.88-1.12] | 0.89 | 0.99 | [0.88-1.12] | 0.89 | 0.50 |
| smoke_utero | rs17389644 | S1 | 13840 | 11 | 0.96 | [0.82-1.14] | 0.67 | 0.96 | [0.82-1.14] | 0.67 | 0.73 |
| smoke_utero | rs17389644 | S2 | 12185 | 11 | 0.92 | [0.78-1.09] | 0.33 | 0.92 | [0.78-1.09] | 0.33 | 0.55 |
| smoke_utero | rs17389644 | S3 | 9992 | 11 | 0.94 | [0.79-1.13] | 0.51 | 0.94 | [0.79-1.13] | 0.51 | 0.61 |
| smoke_utero | rs17881320 | I1 | 16634 | 12 | 1.03 | [0.82-1.29] | 0.79 | 1.05 | [0.82-1.31] | 0.71 | 0.38 |
| smoke_utero | rs17881320 | I2 | 12462 | 12 | 1.07 | [0.97-1.19] | 0.19 | 1.05 | [0.90-1.26] | 0.52 | 0.05 |
| smoke_utero | rs17881320 | I3 | 4172 | 12 | 1.11 | [0.92-1.34] | 0.28 | 1.11 | [0.92-1.34] | 0.28 | 0.74 |

| | | | | | | | | | | | |
|---|---|---|---|---|---|---|---|---|---|---|---|
| smoke_utero | rs17881320 | S1 | 14428 | 12 | 1.03 | [0.80-1.32] | 0.84 | 1.09 | [0.79-1.43] | 0.6 | 0.18 |
| smoke_utero | rs17881320 | S2 | 12768 | 12 | 0.91 | [0.71-1.18] | 0.49 | 0.94 | [0.69-1.24] | 0.69 | 0.21 |
| smoke_utero | rs17881320 | S3 | 10580 | 12 | 0.90 | [0.67-1.19] | 0.46 | 0.94 | [0.65-1.30] | 0.73 | 0.16 |
| smoke_utero | rs2041733 | I1 | 16063 | 11 | 1.10 | [0.98-1.25] | 0.11 | 1.10 | [0.98-1.25] | 0.11 | 0.54 |
| smoke_utero | rs2041733 | I2 | 11985 | 11 | 1.01 | [0.95-1.07] | 0.77 | 1.01 | [0.95-1.07] | 0.77 | 0.54 |
| smoke_utero | rs2041733 | I3 | 4078 | 11 | 1.11 | [1.01-1.23] | 3.70E-02 | 1.11 | [1.01-1.23] | 3.70E-02 | 0.71 |
| smoke_utero | rs2041733 | S1 | 13839 | 11 | 1.14 | [1.00-1.31] | 5.30E-02 | 1.14 | [1.00-1.31] | 5.30E-02 | 0.58 |
| smoke_utero | rs2041733 | S2 | 12197 | 11 | 1.10 | [0.97-1.26] | 0.15 | 1.10 | [0.97-1.26] | 0.15 | 0.42 |
| smoke_utero | rs2041733 | S3 | 9991 | 11 | 1.14 | [0.98-1.32] | 7.90E-02 | 1.14 | [0.98-1.33] | 8.20E-02 | 0.40 |
| smoke_utero | rs2143950 | I1 | 16641 | 12 | 0.98 | [0.84-1.15] | 0.81 | 0.99 | [0.81-1.20] | 0.91 | 0.18 |
| smoke_utero | rs2143950 | I2 | 12463 | 12 | 1.08 | [1.00-1.16] | 4.80E-02 | 1.08 | [1.00-1.17] | 6.30E-02 | 0.42 |
| smoke_utero | rs2143950 | I3 | 4178 | 12 | 1.02 | [0.89-1.16] | 0.8 | 1.02 | [0.87-1.19] | 0.81 | 0.22 |
| smoke_utero | rs2143950 | S1 | 14421 | 12 | 0.92 | [0.77-1.10] | 0.36 | 0.92 | [0.77-1.10] | 0.36 | 0.63 |
| smoke_utero | rs2143950 | S2 | 12775 | 12 | 0.98 | [0.83-1.16] | 0.83 | 0.98 | [0.79-1.21] | 0.81 | 0.21 |
| smoke_utero | rs2143950 | S3 | 10573 | 12 | 0.93 | [0.77-1.12] | 0.45 | 0.93 | [0.77-1.12] | 0.45 | 0.62 |
| smoke_utero | rs2164983 | I1 | 13586 | 8 | 0.90 | [0.75-1.07] | 0.23 | 0.90 | [0.75-1.07] | 0.23 | 0.60 |
| smoke_utero | rs2164983 | I2 | 10121 | 8 | 1.17 | [1.07-1.27] | 4.90E-04 | 1.17 | [1.07-1.27] | 4.90E-04 | 0.55 |
| smoke_utero | rs2164983 | I3 | 3465 | 8 | 1.06 | [0.91-1.22] | 0.46 | 1.06 | [0.90-1.24] | 0.47 | 0.35 |
| smoke_utero | rs2164983 | S1 | 12875 | 8 | 0.91 | [0.75-1.10] | 0.31 | 0.92 | [0.74-1.12] | 0.42 | 0.36 |
| smoke_utero | rs2164983 | S2 | 10685 | 8 | 0.88 | [0.73-1.06] | 0.18 | 0.88 | [0.73-1.06] | 0.18 | 0.48 |
| smoke_utero | rs2164983 | S3 | 9992 | 8 | 0.89 | [0.73-1.09] | 0.26 | 0.91 | [0.71-1.14] | 0.46 | 0.24 |
| smoke_utero | rs2227483 | I1 | 16643 | 12 | 0.94 | [0.84-1.06] | 0.34 | 0.94 | [0.84-1.06] | 0.34 | 0.73 |
| smoke_utero | rs2227483 | I2 | 12467 | 12 | 1.06 | [1.00-1.13] | 4.20E-02 | 1.06 | [1.00-1.13] | 4.70E-02 | 0.43 |
| smoke_utero | rs2227483 | I3 | 4176 | 12 | 1.03 | [0.93-1.14] | 0.58 | 1.03 | [0.92-1.15] | 0.63 | 0.36 |
| smoke_utero | rs2227483 | S1 | 14423 | 12 | 0.91 | [0.80-1.04] | 0.17 | 0.91 | [0.80-1.04] | 0.17 | 0.53 |
| smoke_utero | rs2227483 | S2 | 12777 | 12 | 0.92 | [0.81-1.05] | 0.23 | 0.92 | [0.81-1.05] | 0.23 | 0.54 |
| smoke_utero | rs2227483 | S3 | 10575 | 12 | 0.89 | [0.77-1.03] | 0.11 | 0.90 | [0.76-1.05] | 0.18 | 0.30 |
| smoke_utero | rs2228145 | I1 | 14551 | 10 | 0.96 | [0.84-1.09] | 0.5 | 0.96 | [0.84-1.09] | 0.5 | 0.76 |
| smoke_utero | rs2228145 | I2 | 10906 | 10 | 1.05 | [0.99-1.12] | 0.11 | 1.05 | [0.99-1.12] | 0.11 | 0.55 |
| smoke_utero | rs2228145 | I3 | 3645 | 10 | 1.05 | [0.94-1.16] | 0.4 | 1.05 | [0.94-1.16] | 0.4 | 0.69 |
| smoke_utero | rs2228145 | S1 | 13840 | 10 | 0.95 | [0.83-1.09] | 0.49 | 0.95 | [0.83-1.09] | 0.49 | 0.65 |
| smoke_utero | rs2228145 | S2 | 10685 | 10 | 0.94 | [0.81-1.08] | 0.37 | 0.94 | [0.81-1.08] | 0.37 | 0.76 |
| smoke_utero | rs2228145 | S3 | 9992 | 10 | 0.93 | [0.80-1.08] | 0.37 | 0.93 | [0.80-1.08] | 0.37 | 0.56 |
| smoke_utero | rs2897442 | I1 | 14701 | 10 | 0.98 | [0.85-1.14] | 0.83 | 0.94 | [0.77-1.20] | 0.58 | 0.14 |
| smoke_utero | rs2897442 | I2 | 11374 | 10 | 1.12 | [1.05-1.20] | 1.10E-03 | 1.12 | [1.03-1.22] | 8.50E-03 | 0.23 |
| smoke_utero | rs2897442 | I3 | 3327 | 10 | 1.11 | [0.98-1.26] | 0.11 | 1.07 | [0.89-1.33] | 0.45 | 0.08 |
| smoke_utero | rs2897442 | S1 | 12478 | 10 | 0.96 | [0.81-1.13] | 0.62 | 0.90 | [0.70-1.23] | 0.39 | 0.09 |
| smoke_utero | rs2897442 | S2 | 10835 | 10 | 1.00 | [0.84-1.17] | 0.96 | 0.97 | [0.77-1.24] | 0.77 | 0.14 |
| smoke_utero | rs2897442 | S3 | 8630 | 10 | 0.98 | [0.81-1.18] | 0.82 | 0.92 | [0.68-1.32] | 0.6 | 0.06 |
| smoke_utero | rs479844 | I1 | 14702 | 10 | 1.05 | [0.92-1.20] | 0.47 | 1.05 | [0.92-1.20] | 0.47 | 0.77 |
| smoke_utero | rs479844 | I2 | 11375 | 10 | 1.14 | [1.07-1.21] | 2.10E-05 | 1.14 | [1.07-1.21] | 2.40E-05 | 0.43 |
| smoke_utero | rs479844 | I3 | 3327 | 10 | 1.17 | [1.05-1.31] | 6.30E-03 | 1.17 | [1.05-1.31] | 6.30E-03 | 0.63 |
| smoke_utero | rs479844 | S1 | 12478 | 10 | 1.12 | [0.97-1.30] | 0.13 | 1.12 | [0.97-1.30] | 0.13 | 0.66 |
| smoke_utero | rs479844 | S2 | 10836 | 10 | 1.03 | [0.89-1.19] | 0.69 | 1.03 | [0.89-1.19] | 0.69 | 0.80 |
| smoke_utero | rs479844 | S3 | 8630 | 10 | 1.10 | [0.93-1.29] | 0.28 | 1.10 | [0.93-1.29] | 0.28 | 0.74 |
| smoke_utero | rs6010620 | I1 | 16064 | 11 | 0.99 | [0.85-1.14] | 0.84 | 0.99 | [0.85-1.14] | 0.84 | 0.89 |
| smoke_utero | rs6010620 | I2 | 11985 | 11 | 1.10 | [1.02-1.18] | 8.50E-03 | 1.10 | [1.02-1.18] | 8.50E-03 | 0.95 |
| smoke_utero | rs6010620 | I3 | 4079 | 11 | 1.07 | [0.95-1.21] | 0.25 | 1.07 | [0.95-1.21] | 0.25 | 0.96 |
| smoke_utero | rs6010620 | S1 | 13840 | 11 | 1.02 | [0.87-1.19] | 0.81 | 1.02 | [0.87-1.19] | 0.81 | 0.81 |
| smoke_utero | rs6010620 | S2 | 12198 | 11 | 1.00 | [0.85-1.16] | 0.97 | 1.00 | [0.85-1.16] | 0.97 | 0.88 |
| smoke_utero | rs6010620 | S3 | 9992 | 11 | 1.05 | [0.88-1.24] | 0.6 | 1.05 | [0.88-1.24] | 0.6 | 0.83 |
| smoke_utero | rs6473227 | I1 | 16654 | 12 | 0.97 | [0.86-1.10] | 0.68 | 0.97 | [0.84-1.12] | 0.65 | 0.29 |
| smoke_utero | rs6473227 | I2 | 12475 | 12 | 1.08 | [1.01-1.14] | 1.50E-02 | 1.08 | [1.01-1.14] | 1.50E-02 | 0.95 |
| smoke_utero | rs6473227 | I3 | 4179 | 12 | 1.05 | [0.95-1.16] | 0.35 | 1.03 | [0.90-1.20] | 0.69 | 0.17 |
| smoke_utero | rs6473227 | S1 | 14430 | 12 | 0.92 | [0.81-1.05] | 0.23 | 0.92 | [0.81-1.05] | 0.23 | 0.59 |
| smoke_utero | rs6473227 | S2 | 12788 | 12 | 0.98 | [0.86-1.12] | 0.79 | 0.97 | [0.83-1.16] | 0.75 | 0.20 |
| smoke_utero | rs6473227 | S3 | 10582 | 12 | 0.93 | [0.80-1.07] | 0.3 | 0.93 | [0.80-1.07] | 0.3 | 0.52 |
| smoke_utero | rs7127307 | I1 | 16653 | 12 | 1.01 | [0.89-1.14] | 0.93 | 1.01 | [0.89-1.14] | 0.93 | 0.72 |
| smoke_utero | rs7127307 | I2 | 12474 | 12 | 1.05 | [0.99-1.11] | 0.12 | 1.05 | [0.99-1.11] | 0.12 | 0.86 |
| smoke_utero | rs7127307 | I3 | 4179 | 12 | 1.06 | [0.96-1.17] | 0.27 | 1.06 | [0.96-1.17] | 0.27 | 0.67 |
| smoke_utero | rs7127307 | S1 | 14429 | 12 | 1.03 | [0.90-1.18] | 0.67 | 1.03 | [0.90-1.18] | 0.67 | 0.80 |
| smoke_utero | rs7127307 | S2 | 12787 | 12 | 1.02 | [0.89-1.16] | 0.82 | 1.02 | [0.89-1.16] | 0.82 | 0.64 |
| smoke_utero | rs7127307 | S3 | 10581 | 12 | 1.04 | [0.90-1.21] | 0.58 | 1.04 | [0.90-1.21] | 0.58 | 0.63 |
| smoke_utero | rs7146581 | I1 | 16646 | 12 | 0.94 | [0.82-1.08] | 0.4 | 0.95 | [0.81-1.11] | 0.57 | 0.27 |
| smoke_utero | rs7146581 | I2 | 12469 | 12 | 1.05 | [0.97-1.12] | 0.21 | 1.04 | [0.97-1.12] | 0.23 | 0.43 |
| smoke_utero | rs7146581 | I3 | 4177 | 12 | 0.99 | [0.88-1.11] | 0.85 | 0.99 | [0.88-1.12] | 0.89 | 0.38 |
| smoke_utero | rs7146581 | S1 | 14424 | 12 | 0.91 | [0.78-1.07] | 0.25 | 0.91 | [0.74-1.13] | 0.42 | 0.11 |
| smoke_utero | rs7146581 | S2 | 12780 | 12 | 1.01 | [0.86-1.17] | 0.95 | 1.01 | [0.86-1.17] | 0.95 | 0.62 |
| smoke_utero | rs7146581 | S3 | 10576 | 12 | 0.98 | [0.83-1.16] | 0.85 | 0.98 | [0.83-1.16] | 0.85 | 0.53 |
| smoke_utero | rs7927894 | I1 | 16024 | 11 | 1.02 | [0.90-1.16] | 0.75 | 1.02 | [0.90-1.16] | 0.75 | 0.48 |
| smoke_utero | rs7927894 | I2 | 11956 | 11 | 1.01 | [0.95-1.07] | 0.81 | 1.01 | [0.93-1.09] | 0.85 | 0.18 |
| smoke_utero | rs7927894 | I3 | 4068 | 11 | 1.02 | [0.92-1.13] | 0.77 | 1.02 | [0.92-1.13] | 0.77 | 0.50 |
| smoke_utero | rs7927894 | S1 | 13840 | 11 | 1.06 | [0.93-1.22] | 0.39 | 1.06 | [0.92-1.23] | 0.45 | 0.39 |
| smoke_utero | rs7927894 | S2 | 12158 | 11 | 0.99 | [0.87-1.14] | 0.91 | 0.99 | [0.87-1.14] | 0.91 | 0.45 |
| smoke_utero | rs7927894 | S3 | 9992 | 11 | 1.03 | [0.89-1.20] | 0.68 | 1.03 | [0.88-1.20] | 0.7 | 0.40 |
| smoke_utero | | M1 | 18696 | 12 | 0.94 | [0.87-1.02] | 0.15 | 0.94 | [0.87-1.02] | 0.15 | 0.64 |
| smoke_utero | | M2 | 16445 | 12 | 0.96 | [0.88-1.05] | 0.35 | 0.96 | [0.88-1.05] | 0.35 | 0.85 |
| smoke_utero | | M3 | 15706 | 12 | 0.94 | [0.87-1.02] | 0.14 | 0.94 | [0.87-1.02] | 0.14 | 0.44 |
| smoke_utero | | M4 | 13482 | 12 | 0.95 | [0.87-1.04] | 0.27 | 0.95 | [0.87-1.04] | 0.27 | 0.74 |
| wash_2y | flg | I1 | 6962 | 3 | 0.71 | [0.51-0.99] | 4.50E-02 | 0.65 | [0.38-1.22] | 0.12 | 0.31 |
| wash_2y | flg | I2 | 3072 | 3 | 2.52 | [2.00-3.17] | 5.10E-15 | 2.52 | [2.00-3.17] | 5.10E-15 | 0.40 |
| wash_2y | flg | I3 | 3890 | 3 | 1.75 | [1.40-2.19] | 6.70E-07 | 1.75 | [1.40-2.19] | 6.70E-07 | 0.58 |
| wash_2y | flg | S1 | 6879 | 3 | 0.70 | [0.50-0.98] | 3.50E-02 | 0.64 | [0.37-1.20] | 0.1 | 0.32 |
| wash_2y | flg | S2 | 6301 | 3 | 0.67 | [0.47-0.95] | 2.50E-02 | 0.67 | [0.47-0.95] | 2.50E-02 | 0.37 |
| wash_2y | flg | S3 | 6243 | 3 | 0.64 | [0.45-0.92] | 1.70E-02 | 0.63 | [0.42-0.96] | 2.60E-02 | 0.36 |
| wash_2y | rs10214237 | I1 | 5593 | 2 | 1.00 | [0.83-1.19] | 0.97 | 1.00 | [0.83-1.19] | 0.97 | 0.75 |
| wash_2y | rs10214237 | I2 | 2311 | 2 | 1.10 | [0.96-1.26] | 0.17 | 1.08 | [0.89-1.34] | 0.46 | 0.22 |
| wash_2y | rs10214237 | I3 | 3282 | 2 | 1.10 | [0.98-1.23] | 0.11 | 1.10 | [0.98-1.23] | 0.11 | 0.36 |
| wash_2y | rs10214237 | S1 | 5532 | 2 | 0.99 | [0.83-1.19] | 0.93 | 0.99 | [0.83-1.19] | 0.93 | 0.69 |
| wash_2y | rs10214237 | S2 | 5062 | 2 | 0.98 | [0.81-1.19] | 0.83 | 0.98 | [0.81-1.19] | 0.83 | 0.65 |
| wash_2y | rs10214237 | S3 | 5019 | 2 | 0.97 | [0.80-1.18] | 0.74 | 0.97 | [0.80-1.18] | 0.74 | 0.59 |

| | | | | | | | | | | | |
|---|---|---|---|---|---|---|---|---|---|---|---|
| wash_2y | rs1057258 | I1 | 5593 | 2 | 1.05 | [0.85-1.30] | 0.65 | 1.05 | [0.85-1.30] | 0.65 | 0.57 |
| wash_2y | rs1057258 | I2 | 2311 | 2 | 0.99 | [0.84-1.16] | 0.88 | 0.99 | [0.84-1.16] | 0.88 | 0.71 |
| wash_2y | rs1057258 | I3 | 3282 | 2 | 1.04 | [0.90-1.19] | 0.61 | 1.04 | [0.90-1.19] | 0.61 | 0.65 |
| wash_2y | rs1057258 | S1 | 5532 | 2 | 1.04 | [0.84-1.29] | 0.7 | 1.04 | [0.84-1.29] | 0.7 | 0.59 |
| wash_2y | rs1057258 | S2 | 5062 | 2 | 1.06 | [0.84-1.33] | 0.62 | 1.06 | [0.84-1.33] | 0.62 | 0.56 |
| wash_2y | rs1057258 | S3 | 5019 | 2 | 1.06 | [0.84-1.33] | 0.62 | 1.06 | [0.84-1.33] | 0.62 | 0.55 |
| wash_2y | rs10995251 | I1 | 5593 | 2 | 1.09 | [0.92-1.28] | 0.32 | 1.09 | [0.92-1.28] | 0.32 | 0.97 |
| wash_2y | rs10995251 | I2 | 2311 | 2 | 0.96 | [0.84-1.09] | 0.48 | 0.96 | [0.84-1.09] | 0.48 | 0.81 |
| wash_2y | rs10995251 | I3 | 3282 | 2 | 1.04 | [0.94-1.16] | 0.47 | 1.04 | [0.94-1.16] | 0.47 | 0.80 |
| wash_2y | rs10995251 | S1 | 5532 | 2 | 1.08 | [0.92-1.28] | 0.35 | 1.08 | [0.92-1.28] | 0.35 | 0.93 |
| wash_2y | rs10995251 | S2 | 5062 | 2 | 1.07 | [0.90-1.28] | 0.45 | 1.07 | [0.90-1.28] | 0.45 | 0.82 |
| wash_2y | rs10995251 | S3 | 5019 | 2 | 1.07 | [0.89-1.28] | 0.47 | 1.07 | [0.89-1.28] | 0.47 | 0.85 |
| wash_2y | rs112111458 | I1 | 5593 | 2 | 0.84 | [0.67-1.05] | 0.13 | 0.84 | [0.67-1.05] | 0.13 | 0.99 |
| wash_2y | rs112111458 | I2 | 2311 | 2 | 1.11 | [0.93-1.33] | 0.25 | 1.13 | [0.90-1.39] | 0.28 | 0.25 |
| wash_2y | rs112111458 | I3 | 3282 | 2 | 0.92 | [0.80-1.07] | 0.27 | 0.96 | [0.76-1.17] | 0.71 | 0.18 |
| wash_2y | rs112111458 | S1 | 5532 | 2 | 0.82 | [0.65-1.04] | 0.1 | 0.82 | [0.65-1.04] | 0.1 | 0.89 |
| wash_2y | rs112111458 | S2 | 5062 | 2 | 0.85 | [0.67-1.08] | 0.19 | 0.85 | [0.67-1.08] | 0.19 | 1.00 |
| wash_2y | rs112111458 | S3 | 5019 | 2 | 0.83 | [0.65-1.06] | 0.14 | 0.83 | [0.65-1.06] | 0.14 | 0.91 |
| wash_2y | rs11657987 | I1 | 5593 | 2 | 1.09 | [0.93-1.28] | 0.3 | 1.09 | [0.93-1.28] | 0.3 | 0.95 |
| wash_2y | rs11657987 | I2 | 2311 | 2 | 1.04 | [0.92-1.18] | 0.52 | 1.04 | [0.92-1.18] | 0.52 | 0.78 |
| wash_2y | rs11657987 | I3 | 3282 | 2 | 1.13 | [1.02-1.25] | 1.80E-02 | 1.13 | [1.02-1.25] | 1.80E-02 | 0.69 |
| wash_2y | rs11657987 | S1 | 5532 | 2 | 1.09 | [0.92-1.28] | 0.32 | 1.09 | [0.92-1.28] | 0.32 | 0.95 |
| wash_2y | rs11657987 | S2 | 5062 | 2 | 1.08 | [0.91-1.28] | 0.4 | 1.08 | [0.91-1.28] | 0.4 | 0.91 |
| wash_2y | rs11657987 | S3 | 5019 | 2 | 1.08 | [0.91-1.28] | 0.41 | 1.08 | [0.91-1.28] | 0.41 | 0.92 |
| wash_2y | rs12153855 | I1 | 5593 | 2 | 0.87 | [0.66-1.14] | 0.31 | 0.74 | [0.37-1.74] | 0.39 | 0.04 |
| wash_2y | rs12153855 | I2 | 2311 | 2 | 1.26 | [1.02-1.55] | 3.40E-02 | 1.35 | [0.92-1.85] | 0.13 | 0.13 |
| wash_2y | rs12153855 | I3 | 3282 | 2 | 1.09 | [0.92-1.30] | 0.32 | 1.04 | [0.78-1.46] | 0.77 | 0.15 |
| wash_2y | rs12153855 | S1 | 5532 | 2 | 0.89 | [0.68-1.18] | 0.43 | 0.76 | [0.37-1.82] | 0.44 | 0.04 |
| wash_2y | rs12153855 | S2 | 5062 | 2 | 0.86 | [0.65-1.15] | 0.31 | 0.74 | [0.36-1.77] | 0.41 | 0.04 |
| wash_2y | rs12153855 | S3 | 5019 | 2 | 0.89 | [0.66-1.19] | 0.43 | 0.76 | [0.36-1.85] | 0.46 | 0.03 |
| wash_2y | rs12295535 | I1 | 5593 | 2 | 1.14 | [0.70-1.86] | 0.6 | 1.14 | [0.70-1.86] | 0.6 | 0.80 |
| wash_2y | rs12295535 | I2 | 2311 | 2 | 1.08 | [0.72-1.61] | 0.72 | 1.08 | [0.72-1.61] | 0.72 | 0.92 |
| wash_2y | rs12295535 | I3 | 3282 | 2 | 1.23 | [0.93-1.63] | 0.15 | 1.23 | [0.93-1.63] | 0.15 | 0.62 |
| wash_2y | rs12295535 | S1 | 5532 | 2 | 1.17 | [0.71-1.94] | 0.53 | 1.17 | [0.71-1.94] | 0.53 | 0.73 |
| wash_2y | rs12295535 | S2 | 5062 | 2 | 1.23 | [0.72-2.12] | 0.44 | 1.23 | [0.72-2.12] | 0.44 | 0.73 |
| wash_2y | rs12295535 | S3 | 5019 | 2 | 1.17 | [0.68-2.02] | 0.57 | 1.17 | [0.68-2.02] | 0.57 | 0.74 |
| wash_2y | rs13015714 | I1 | 5593 | 2 | 0.98 | [0.81-1.18] | 0.84 | 0.98 | [0.81-1.18] | 0.84 | 0.46 |
| wash_2y | rs13015714 | I2 | 2311 | 2 | 1.11 | [0.96-1.28] | 0.17 | 1.11 | [0.96-1.28] | 0.17 | 0.67 |
| wash_2y | rs13015714 | I3 | 3282 | 2 | 1.08 | [0.96-1.22] | 0.19 | 1.08 | [0.96-1.22] | 0.19 | 0.47 |
| wash_2y | rs13015714 | S1 | 5532 | 2 | 0.98 | [0.81-1.19] | 0.86 | 0.98 | [0.81-1.19] | 0.86 | 0.55 |
| wash_2y | rs13015714 | S2 | 5062 | 2 | 0.97 | [0.79-1.18] | 0.75 | 0.97 | [0.79-1.18] | 0.75 | 0.58 |
| wash_2y | rs13015714 | S3 | 5019 | 2 | 0.96 | [0.78-1.17] | 0.67 | 0.96 | [0.78-1.17] | 0.67 | 0.64 |
| wash_2y | rs16948048 | I1 | 5593 | 2 | 1.14 | [0.96-1.34] | 0.13 | 1.14 | [0.96-1.34] | 0.13 | 0.37 |
| wash_2y | rs16948048 | I2 | 2311 | 2 | 0.92 | [0.81-1.05] | 0.2 | 0.92 | [0.81-1.05] | 0.2 | 0.52 |
| wash_2y | rs16948048 | I3 | 3282 | 2 | 1.05 | [0.94-1.16] | 0.4 | 1.05 | [0.94-1.16] | 0.4 | 0.51 |
| wash_2y | rs16948048 | S1 | 5532 | 2 | 1.13 | [0.96-1.34] | 0.14 | 1.13 | [0.96-1.34] | 0.14 | 0.42 |
| wash_2y | rs16948048 | S2 | 5062 | 2 | 1.10 | [0.92-1.31] | 0.31 | 1.10 | [0.92-1.31] | 0.31 | 0.55 |
| wash_2y | rs16948048 | S3 | 5019 | 2 | 1.09 | [0.92-1.31] | 0.32 | 1.09 | [0.92-1.31] | 0.32 | 0.56 |
| wash_2y | rs17389644 | I1 | 5593 | 2 | 0.99 | [0.82-1.20] | 0.91 | 0.99 | [0.82-1.20] | 0.91 | 0.67 |
| wash_2y | rs17389644 | I2 | 2311 | 2 | 1.13 | [0.97-1.31] | 0.11 | 1.04 | [0.74-1.59] | 0.82 | 0.07 |
| wash_2y | rs17389644 | I3 | 3282 | 2 | 1.12 | [0.99-1.27] | 6.40E-02 | 1.08 | [0.87-1.40] | 0.47 | 0.17 |
| wash_2y | rs17389644 | S1 | 5532 | 2 | 0.98 | [0.81-1.19] | 0.86 | 0.98 | [0.81-1.19] | 0.86 | 0.58 |
| wash_2y | rs17389644 | S2 | 5062 | 2 | 1.02 | [0.83-1.25] | 0.85 | 1.02 | [0.83-1.25] | 0.85 | 0.78 |
| wash_2y | rs17389644 | S3 | 5019 | 2 | 1.01 | [0.82-1.24] | 0.92 | 1.01 | [0.82-1.24] | 0.92 | 0.66 |
| wash_2y | rs17881320 | I1 | 5593 | 2 | 0.89 | [0.67-1.20] | 0.46 | 0.89 | [0.67-1.20] | 0.46 | 0.75 |
| wash_2y | rs17881320 | I2 | 2311 | 2 | 1.13 | [0.90-1.42] | 0.29 | 1.03 | [0.65-1.78] | 0.9 | 0.14 |
| wash_2y | rs17881320 | I3 | 3282 | 2 | 1.04 | [0.86-1.25] | 0.69 | 1.01 | [0.77-1.36] | 0.96 | 0.26 |
| wash_2y | rs17881320 | S1 | 5532 | 2 | 0.91 | [0.68-1.23] | 0.54 | 0.91 | [0.68-1.23] | 0.54 | 0.69 |
| wash_2y | rs17881320 | S2 | 5062 | 2 | 0.90 | [0.65-1.24] | 0.52 | 0.90 | [0.65-1.24] | 0.52 | 0.69 |
| wash_2y | rs17881320 | S3 | 5019 | 2 | 0.91 | [0.66-1.25] | 0.55 | 0.91 | [0.66-1.25] | 0.55 | 0.66 |
| wash_2y | rs2041733 | I1 | 5593 | 2 | 1.08 | [0.92-1.27] | 0.37 | 1.08 | [0.92-1.27] | 0.37 | 0.40 |
| wash_2y | rs2041733 | I2 | 2311 | 2 | 0.98 | [0.87-1.12] | 0.81 | 0.98 | [0.85-1.13] | 0.77 | 0.29 |
| wash_2y | rs2041733 | I3 | 3282 | 2 | 1.06 | [0.96-1.18] | 0.26 | 1.06 | [0.96-1.18] | 0.26 | 0.90 |
| wash_2y | rs2041733 | S1 | 5532 | 2 | 1.07 | [0.91-1.27] | 0.39 | 1.07 | [0.91-1.27] | 0.39 | 0.36 |
| wash_2y | rs2041733 | S2 | 5062 | 2 | 1.11 | [0.93-1.32] | 0.24 | 1.11 | [0.93-1.32] | 0.24 | 0.49 |
| wash_2y | rs2041733 | S3 | 5019 | 2 | 1.11 | [0.93-1.32] | 0.24 | 1.11 | [0.93-1.32] | 0.24 | 0.45 |
| wash_2y | rs2143950 | I1 | 5593 | 2 | 0.83 | [0.68-1.03] | 8.60E-02 | 0.92 | [0.59-1.29] | 0.69 | 0.10 |
| wash_2y | rs2143950 | I2 | 2311 | 2 | 1.20 | [1.02-1.40] | 2.30E-02 | 1.08 | [0.70-1.84] | 0.74 | 0.04 |
| wash_2y | rs2143950 | I3 | 3282 | 2 | 1.00 | [0.88-1.15] | 0.97 | 1.00 | [0.88-1.15] | 0.97 | 0.91 |
| wash_2y | rs2143950 | S1 | 5532 | 2 | 0.83 | [0.68-1.02] | 8.20E-02 | 0.91 | [0.59-1.28] | 0.68 | 0.10 |
| wash_2y | rs2143950 | S2 | 5062 | 2 | 0.84 | [0.67-1.05] | 0.12 | 0.91 | [0.59-1.30] | 0.68 | 0.11 |
| wash_2y | rs2143950 | S3 | 5019 | 2 | 0.84 | [0.67-1.05] | 0.12 | 0.91 | [0.59-1.30] | 0.67 | 0.10 |
| wash_2y | rs2164983 | I1 | 5593 | 2 | 0.94 | [0.76-1.17] | 0.59 | 0.94 | [0.76-1.17] | 0.59 | 0.64 |
| wash_2y | rs2164983 | I2 | 2311 | 2 | 1.20 | [1.02-1.41] | 2.80E-02 | 1.20 | [1.02-1.41] | 2.80E-02 | 0.42 |
| wash_2y | rs2164983 | I3 | 3282 | 2 | 1.14 | [0.99-1.31] | 6.20E-02 | 1.07 | [0.81-1.51] | 0.64 | 0.11 |
| wash_2y | rs2164983 | S1 | 5532 | 2 | 0.94 | [0.76-1.17] | 0.6 | 0.94 | [0.76-1.17] | 0.6 | 0.71 |
| wash_2y | rs2164983 | S2 | 5062 | 2 | 0.93 | [0.74-1.16] | 0.52 | 0.93 | [0.74-1.16] | 0.52 | 0.67 |
| wash_2y | rs2164983 | S3 | 5019 | 2 | 0.93 | [0.74-1.17] | 0.55 | 0.93 | [0.74-1.17] | 0.55 | 0.72 |
| wash_2y | rs2227483 | I1 | 5593 | 2 | 0.95 | [0.81-1.12] | 0.54 | 0.92 | [0.73-1.20] | 0.51 | 0.22 |
| wash_2y | rs2227483 | I2 | 2311 | 2 | 1.12 | [0.99-1.27] | 8.40E-02 | 1.12 | [0.98-1.28] | 9.60E-02 | 0.30 |
| wash_2y | rs2227483 | I3 | 3282 | 2 | 1.06 | [0.96-1.18] | 0.26 | 1.06 | [0.96-1.18] | 0.26 | 0.50 |
| wash_2y | rs2227483 | S1 | 5532 | 2 | 0.97 | [0.82-1.14] | 0.68 | 0.92 | [0.70-1.28] | 0.57 | 0.16 |
| wash_2y | rs2227483 | S2 | 5062 | 2 | 0.94 | [0.79-1.12] | 0.5 | 0.92 | [0.72-1.21] | 0.49 | 0.21 |
| wash_2y | rs2227483 | S3 | 5019 | 2 | 0.96 | [0.81-1.15] | 0.67 | 0.92 | [0.68-1.30] | 0.58 | 0.15 |
| wash_2y | rs2228145 | I1 | 5593 | 2 | 1.06 | [0.90-1.25] | 0.46 | 1.06 | [0.90-1.25] | 0.46 | 0.73 |
| wash_2y | rs2228145 | I2 | 2311 | 2 | 0.97 | [0.85-1.10] | 0.61 | 0.97 | [0.85-1.10] | 0.61 | 0.78 |
| wash_2y | rs2228145 | I3 | 3282 | 2 | 1.03 | [0.93-1.14] | 0.59 | 1.03 | [0.93-1.14] | 0.59 | 0.86 |
| wash_2y | rs2228145 | S1 | 5532 | 2 | 1.06 | [0.90-1.25] | 0.47 | 1.06 | [0.90-1.25] | 0.47 | 0.80 |
| wash_2y | rs2228145 | S2 | 5062 | 2 | 1.04 | [0.87-1.23] | 0.68 | 1.04 | [0.87-1.23] | 0.68 | 0.62 |
| wash_2y | rs2228145 | S3 | 5019 | 2 | 1.03 | [0.87-1.23] | 0.72 | 1.03 | [0.87-1.23] | 0.72 | 0.67 |
| wash_2y | rs2897442 | I1 | 5593 | 2 | 1.00 | [0.84-1.21] | 0.97 | 1.00 | [0.84-1.21] | 0.97 | 0.34 |

| | | | | | | | | | | | |
|---|---|---|---|---|---|---|---|---|---|---|---|
| wash_2y | rs2897442 | I2 | 2311 | 2 | 1.10 | [0.95-1.26] | 0.19 | 1.10 | [0.95-1.26] | 0.19 | 0.89 |
| wash_2y | rs2897442 | I3 | 3282 | 2 | 1.10 | [0.98-1.24] | 0.11 | 1.03 | [0.77-1.46] | 0.86 | 0.08 |
| wash_2y | rs2897442 | S1 | 5532 | 2 | 1.03 | [0.86-1.24] | 0.72 | 1.03 | [0.86-1.24] | 0.72 | 0.35 |
| wash_2y | rs2897442 | S2 | 5062 | 2 | 1.02 | [0.84-1.24] | 0.86 | 1.02 | [0.84-1.24] | 0.87 | 0.32 |
| wash_2y | rs2897442 | S3 | 5019 | 2 | 1.04 | [0.85-1.26] | 0.71 | 1.04 | [0.85-1.27] | 0.71 | 0.32 |
| wash_2y | rs479844 | I1 | 5593 | 2 | 0.97 | [0.83-1.15] | 0.75 | 0.97 | [0.83-1.15] | 0.75 | 0.85 |
| wash_2y | rs479844 | I2 | 2311 | 2 | 1.15 | [1.01-1.31] | 2.90E-02 | 1.15 | [1.01-1.31] | 2.90E-02 | 0.33 |
| wash_2y | rs479844 | I3 | 3282 | 2 | 1.13 | [1.01-1.25] | 2.70E-02 | 1.13 | [1.01-1.25] | 2.70E-02 | 0.38 |
| wash_2y | rs479844 | S1 | 5532 | 2 | 0.97 | [0.82-1.15] | 0.72 | 0.97 | [0.82-1.15] | 0.72 | 0.86 |
| wash_2y | rs479844 | S2 | 5062 | 2 | 0.96 | [0.81-1.15] | 0.69 | 0.96 | [0.81-1.15] | 0.69 | 0.82 |
| wash_2y | rs479844 | S3 | 5019 | 2 | 0.97 | [0.81-1.15] | 0.7 | 0.97 | [0.81-1.15] | 0.7 | 0.83 |
| wash_2y | rs6010620 | I1 | 5593 | 2 | 0.99 | [0.82-1.20] | 0.92 | 0.99 | [0.82-1.20] | 0.92 | 0.87 |
| wash_2y | rs6010620 | I2 | 2311 | 2 | 1.14 | [0.98-1.32] | 8.90E-02 | 1.14 | [0.98-1.32] | 8.90E-02 | 0.72 |
| wash_2y | rs6010620 | I3 | 3282 | 2 | 1.13 | [1.00-1.27] | 5.70E-02 | 1.13 | [1.00-1.27] | 5.70E-02 | 0.85 |
| wash_2y | rs6010620 | S1 | 5532 | 2 | 0.97 | [0.80-1.18] | 0.76 | 0.97 | [0.80-1.18] | 0.76 | 0.77 |
| wash_2y | rs6010620 | S2 | 5062 | 2 | 0.98 | [0.80-1.20] | 0.85 | 0.98 | [0.80-1.20] | 0.85 | 0.88 |
| wash_2y | rs6010620 | S3 | 5019 | 2 | 0.97 | [0.79-1.19] | 0.74 | 0.97 | [0.79-1.19] | 0.74 | 0.79 |
| wash_2y | rs6473227 | I1 | 5593 | 2 | 1.01 | [0.86-1.19] | 0.88 | 1.01 | [0.86-1.19] | 0.88 | 0.40 |
| wash_2y | rs6473227 | I2 | 2311 | 2 | 1.10 | [0.97-1.24] | 0.16 | 1.10 | [0.97-1.24] | 0.16 | 0.55 |
| wash_2y | rs6473227 | I3 | 3282 | 2 | 1.11 | [1.00-1.23] | 5.30E-02 | 1.11 | [1.00-1.23] | 5.30E-02 | 0.55 |
| wash_2y | rs6473227 | S1 | 5532 | 2 | 1.01 | [0.86-1.20] | 0.86 | 1.02 | [0.86-1.20] | 0.85 | 0.31 |
| wash_2y | rs6473227 | S2 | 5062 | 2 | 1.02 | [0.86-1.21] | 0.82 | 1.02 | [0.86-1.21] | 0.82 | 0.36 |
| wash_2y | rs6473227 | S3 | 5019 | 2 | 1.02 | [0.86-1.21] | 0.83 | 1.03 | [0.84-1.24] | 0.78 | 0.28 |
| wash_2y | rs7127307 | I1 | 5593 | 2 | 0.88 | [0.75-1.04] | 0.13 | 0.88 | [0.73-1.06] | 0.15 | 0.29 |
| wash_2y | rs7127307 | I2 | 2311 | 2 | 1.15 | [1.02-1.31] | 2.80E-02 | 1.18 | [0.98-1.39] | 8.60E-02 | 0.21 |
| wash_2y | rs7127307 | I3 | 3282 | 2 | 1.02 | [0.92-1.13] | 0.76 | 1.02 | [0.92-1.13] | 0.76 | 0.92 |
| wash_2y | rs7127307 | S1 | 5532 | 2 | 0.88 | [0.75-1.04] | 0.13 | 0.87 | [0.70-1.09] | 0.18 | 0.26 |
| wash_2y | rs7127307 | S2 | 5062 | 2 | 0.88 | [0.74-1.05] | 0.15 | 0.88 | [0.73-1.06] | 0.16 | 0.30 |
| wash_2y | rs7127307 | S3 | 5019 | 2 | 0.88 | [0.74-1.05] | 0.14 | 0.87 | [0.70-1.09] | 0.18 | 0.26 |
| wash_2y | rs7146581 | I1 | 5593 | 2 | 0.91 | [0.75-1.10] | 0.33 | 0.91 | [0.75-1.10] | 0.33 | 0.75 |
| wash_2y | rs7146581 | I2 | 2311 | 2 | 1.12 | [0.96-1.30] | 0.14 | 1.11 | [0.94-1.32] | 0.22 | 0.29 |
| wash_2y | rs7146581 | I3 | 3282 | 2 | 1.02 | [0.90-1.15] | 0.79 | 1.02 | [0.90-1.15] | 0.79 | 0.45 |
| wash_2y | rs7146581 | S1 | 5532 | 2 | 0.91 | [0.75-1.11] | 0.35 | 0.91 | [0.75-1.11] | 0.35 | 0.61 |
| wash_2y | rs7146581 | S2 | 5062 | 2 | 0.90 | [0.73-1.10] | 0.3 | 0.90 | [0.73-1.10] | 0.3 | 0.69 |
| wash_2y | rs7146581 | S3 | 5019 | 2 | 0.91 | [0.74-1.12] | 0.37 | 0.91 | [0.74-1.12] | 0.37 | 0.59 |
| wash_2y | rs7927894 | I1 | 5593 | 2 | 1.04 | [0.89-1.23] | 0.61 | 1.04 | [0.89-1.23] | 0.61 | 0.63 |
| wash_2y | rs7927894 | I2 | 2311 | 2 | 1.05 | [0.93-1.19] | 0.41 | 1.05 | [0.93-1.19] | 0.41 | 0.77 |
| wash_2y | rs7927894 | I3 | 3282 | 2 | 1.10 | [0.99-1.22] | 7.70E-02 | 1.10 | [0.99-1.22] | 7.70E-02 | 0.74 |
| wash_2y | rs7927894 | S1 | 5532 | 2 | 1.04 | [0.89-1.23] | 0.61 | 1.04 | [0.89-1.23] | 0.61 | 0.62 |
| wash_2y | rs7927894 | S2 | 5062 | 2 | 1.02 | [0.86-1.21] | 0.84 | 1.02 | [0.86-1.21] | 0.84 | 0.70 |
| wash_2y | rs7927894 | S3 | 5019 | 2 | 1.02 | [0.86-1.21] | 0.81 | 1.02 | [0.86-1.21] | 0.81 | 0.68 |
| wash_2y | | M1 | 7747 | 3 | 0.98 | [0.89-1.08] | 0.7 | 0.86 | [0.64-1.31] | 0.3 | 0.01 |
| wash_2y | | M2 | 7663 | 3 | 0.98 | [0.88-1.08] | 0.64 | 0.86 | [0.64-1.30] | 0.29 | 0.01 |
| wash_2y | | M3 | 7062 | 3 | 0.99 | [0.89-1.10] | 0.82 | 0.88 | [0.67-1.30] | 0.35 | 0.02 |
| wash_2y | | M4 | 7003 | 3 | 0.99 | [0.89-1.10] | 0.8 | 0.88 | [0.66-1.30] | 0.35 | 0.02 |
| wash_6m | flg | I1 | 7135 | 3 | 0.75 | [0.55-1.03] | 7.20E-02 | 0.75 | [0.55-1.03] | 7.20E-02 | 0.99 |
| wash_6m | flg | I2 | 3432 | 3 | 2.30 | [1.85-2.85] | 6.10E-14 | 2.30 | [1.85-2.85] | 6.10E-14 | 0.75 |
| wash_6m | flg | I3 | 3703 | 3 | 1.70 | [1.36-2.13] | 3.30E-06 | 1.70 | [1.36-2.13] | 3.30E-06 | 0.72 |
| wash_6m | flg | S1 | 7049 | 3 | 0.72 | [0.52-1.00] | 4.90E-02 | 0.72 | [0.52-1.00] | 4.90E-02 | 0.98 |
| wash_6m | flg | S2 | 6414 | 3 | 0.68 | [0.48-0.95] | 2.50E-02 | 0.68 | [0.48-0.95] | 2.50E-02 | 0.95 |
| wash_6m | flg | S3 | 6352 | 3 | 0.64 | [0.45-0.92] | 1.50E-02 | 0.64 | [0.45-0.92] | 1.50E-02 | 1.00 |
| wash_6m | rs10214237 | I1 | 5718 | 2 | 0.94 | [0.79-1.13] | 0.51 | 0.94 | [0.79-1.13] | 0.51 | 0.49 |
| wash_6m | rs10214237 | I2 | 2517 | 2 | 1.17 | [1.02-1.34] | 2.30E-02 | 1.17 | [1.02-1.34] | 2.30E-02 | 0.86 |
| wash_6m | rs10214237 | I3 | 3201 | 2 | 1.07 | [0.96-1.20] | 0.23 | 1.04 | [0.85-1.31] | 0.7 | 0.12 |
| wash_6m | rs10214237 | S1 | 5653 | 2 | 0.93 | [0.78-1.12] | 0.47 | 0.93 | [0.78-1.12] | 0.47 | 0.52 |
| wash_6m | rs10214237 | S2 | 5127 | 2 | 0.95 | [0.78-1.16] | 0.61 | 0.95 | [0.78-1.16] | 0.61 | 0.46 |
| wash_6m | rs10214237 | S3 | 5078 | 2 | 0.94 | [0.77-1.15] | 0.57 | 0.94 | [0.77-1.15] | 0.57 | 0.47 |
| wash_6m | rs1057258 | I1 | 5718 | 2 | 1.09 | [0.88-1.35] | 0.44 | 0.93 | [0.52-1.94] | 0.8 | 0.06 |
| wash_6m | rs1057258 | I2 | 2517 | 2 | 1.00 | [0.86-1.17] | 0.98 | 1.08 | [0.77-1.41] | 0.66 | 0.17 |
| wash_6m | rs1057258 | I3 | 3201 | 2 | 1.08 | [0.94-1.24] | 0.28 | 1.05 | [0.86-1.33] | 0.62 | 0.18 |
| wash_6m | rs1057258 | S1 | 5653 | 2 | 1.08 | [0.87-1.34] | 0.48 | 0.90 | [0.47-2.05] | 0.74 | 0.05 |
| wash_6m | rs1057258 | S2 | 5127 | 2 | 1.03 | [0.82-1.29] | 0.82 | 0.91 | [0.54-1.73] | 0.72 | 0.10 |
| wash_6m | rs1057258 | S3 | 5078 | 2 | 1.02 | [0.81-1.29] | 0.85 | 0.88 | [0.49-1.83] | 0.67 | 0.07 |
| wash_6m | rs10995251 | I1 | 5718 | 2 | 1.05 | [0.89-1.24] | 0.55 | 1.05 | [0.89-1.24] | 0.55 | 0.48 |
| wash_6m | rs10995251 | I2 | 2517 | 2 | 0.98 | [0.86-1.11] | 0.74 | 0.98 | [0.86-1.11] | 0.74 | 0.45 |
| wash_6m | rs10995251 | I3 | 3201 | 2 | 1.03 | [0.93-1.15] | 0.55 | 1.03 | [0.93-1.15] | 0.55 | 0.86 |
| wash_6m | rs10995251 | S1 | 5653 | 2 | 1.05 | [0.88-1.24] | 0.6 | 1.05 | [0.88-1.24] | 0.6 | 0.41 |
| wash_6m | rs10995251 | S2 | 5127 | 2 | 1.00 | [0.83-1.19] | 0.96 | 1.00 | [0.83-1.19] | 0.96 | 0.66 |
| wash_6m | rs10995251 | S3 | 5078 | 2 | 0.99 | [0.83-1.19] | 0.94 | 0.99 | [0.83-1.19] | 0.94 | 0.58 |
| wash_6m | rs112111458 | I1 | 5718 | 2 | 1.08 | [0.86-1.36] | 0.49 | 1.12 | [0.82-1.48] | 0.48 | 0.26 |
| wash_6m | rs112111458 | I2 | 2517 | 2 | 0.93 | [0.78-1.10] | 0.39 | 0.93 | [0.78-1.10] | 0.39 | 0.87 |
| wash_6m | rs112111458 | I3 | 3201 | 2 | 1.05 | [0.90-1.21] | 0.55 | 1.11 | [0.80-1.44] | 0.53 | 0.05 |
| wash_6m | rs112111458 | S1 | 5653 | 2 | 1.06 | [0.84-1.34] | 0.6 | 1.10 | [0.80-1.46] | 0.56 | 0.26 |
| wash_6m | rs112111458 | S2 | 5127 | 2 | 1.05 | [0.82-1.34] | 0.71 | 1.10 | [0.76-1.51] | 0.62 | 0.22 |
| wash_6m | rs112111458 | S3 | 5078 | 2 | 1.03 | [0.80-1.32] | 0.82 | 1.08 | [0.75-1.49] | 0.68 | 0.22 |
| wash_6m | rs11657987 | I1 | 5718 | 2 | 1.11 | [0.94-1.30] | 0.21 | 1.11 | [0.94-1.30] | 0.21 | 0.82 |
| wash_6m | rs11657987 | I2 | 2517 | 2 | 1.05 | [0.93-1.18] | 0.44 | 1.05 | [0.93-1.18] | 0.44 | 0.87 |
| wash_6m | rs11657987 | I3 | 3201 | 2 | 1.16 | [1.05-1.29] | 4.50E-03 | 1.16 | [1.05-1.29] | 4.50E-03 | 0.90 |
| wash_6m | rs11657987 | S1 | 5653 | 2 | 1.12 | [0.95-1.31] | 0.18 | 1.12 | [0.95-1.31] | 0.18 | 0.83 |
| wash_6m | rs11657987 | S2 | 5127 | 2 | 1.08 | [0.91-1.28] | 0.4 | 1.08 | [0.91-1.28] | 0.4 | 0.71 |
| wash_6m | rs11657987 | S3 | 5078 | 2 | 1.08 | [0.91-1.29] | 0.37 | 1.08 | [0.91-1.29] | 0.37 | 0.71 |
| wash_6m | rs12153855 | I1 | 5718 | 2 | 1.09 | [0.83-1.43] | 0.55 | 1.07 | [0.78-1.49] | 0.67 | 0.29 |
| wash_6m | rs12153855 | I2 | 2517 | 2 | 1.12 | [0.91-1.38] | 0.27 | 1.12 | [0.91-1.38] | 0.27 | 0.48 |
| wash_6m | rs12153855 | I3 | 3201 | 2 | 1.21 | [1.02-1.44] | 3.20E-02 | 1.21 | [1.02-1.44] | 3.20E-02 | 0.42 |
| wash_6m | rs12153855 | S1 | 5653 | 2 | 1.11 | [0.84-1.46] | 0.48 | 1.08 | [0.78-1.54] | 0.64 | 0.28 |
| wash_6m | rs12153855 | S2 | 5127 | 2 | 1.08 | [0.80-1.44] | 0.62 | 1.05 | [0.73-1.54] | 0.79 | 0.27 |
| wash_6m | rs12153855 | S3 | 5078 | 2 | 1.09 | [0.81-1.47] | 0.55 | 1.07 | [0.74-1.57] | 0.72 | 0.27 |
| wash_6m | rs12295535 | I1 | 5718 | 2 | 0.92 | [0.57-1.49] | 0.74 | 0.92 | [0.57-1.49] | 0.74 | 0.79 |
| wash_6m | rs12295535 | I2 | 2517 | 2 | 1.25 | [0.86-1.83] | 0.24 | 1.25 | [0.86-1.83] | 0.24 | 0.65 |
| wash_6m | rs12295535 | I3 | 3201 | 2 | 1.14 | [0.85-1.52] | 0.39 | 1.14 | [0.85-1.52] | 0.39 | 0.78 |
| wash_6m | rs12295535 | S1 | 5653 | 2 | 0.94 | [0.57-1.55] | 0.81 | 0.94 | [0.57-1.55] | 0.81 | 0.56 |

| | | | | | | | | | | | |
|---|---|---|---|---|---|---|---|---|---|---|---|
| wash_6m | rs12295535 | S2 | 5127 | 2 | 1.03 | [0.61-1.73] | 0.91 | 1.03 | [0.61-1.73] | 0.91 | 0.61 |
| wash_6m | rs12295535 | S3 | 5078 | 2 | 1.00 | [0.59-1.72] | 0.99 | 1.00 | [0.59-1.72] | 0.99 | 0.63 |
| wash_6m | rs13015714 | I1 | 5718 | 2 | 0.87 | [0.72-1.05] | 0.15 | 0.86 | [0.68-1.09] | 0.19 | 0.29 |
| wash_6m | rs13015714 | I2 | 2517 | 2 | 1.20 | [1.04-1.38] | 1.10E-02 | 1.20 | [1.04-1.38] | 1.10E-02 | 0.44 |
| wash_6m | rs13015714 | I3 | 3201 | 2 | 1.04 | [0.92-1.18] | 0.5 | 1.04 | [0.92-1.18] | 0.5 | 0.57 |
| wash_6m | rs13015714 | S1 | 5653 | 2 | 0.85 | [0.71-1.04] | 0.11 | 0.85 | [0.71-1.04] | 0.11 | 0.33 |
| wash_6m | rs13015714 | S2 | 5127 | 2 | 0.80 | [0.65-0.98] | 3.10E-02 | 0.80 | [0.65-0.98] | 3.10E-02 | 0.53 |
| wash_6m | rs13015714 | S3 | 5078 | 2 | 0.77 | [0.63-0.95] | 1.50E-02 | 0.77 | [0.63-0.95] | 1.50E-02 | 0.56 |
| wash_6m | rs16948048 | I1 | 5718 | 2 | 1.01 | [0.86-1.19] | 0.89 | 1.01 | [0.86-1.19] | 0.89 | 0.73 |
| wash_6m | rs16948048 | I2 | 2517 | 2 | 1.01 | [0.89-1.14] | 0.9 | 1.01 | [0.89-1.14] | 0.9 | 0.68 |
| wash_6m | rs16948048 | I3 | 3201 | 2 | 1.01 | [0.91-1.13] | 0.8 | 1.01 | [0.91-1.13] | 0.8 | 0.93 |
| wash_6m | rs16948048 | S1 | 5653 | 2 | 1.01 | [0.86-1.20] | 0.87 | 1.01 | [0.86-1.20] | 0.87 | 0.77 |
| wash_6m | rs16948048 | S2 | 5127 | 2 | 1.01 | [0.84-1.20] | 0.95 | 1.01 | [0.84-1.20] | 0.95 | 0.67 |
| wash_6m | rs16948048 | S3 | 5078 | 2 | 1.01 | [0.84-1.20] | 0.93 | 1.01 | [0.84-1.20] | 0.93 | 0.71 |
| wash_6m | rs17389644 | I1 | 5718 | 2 | 1.01 | [0.83-1.23] | 0.9 | 1.01 | [0.83-1.23] | 0.9 | 0.45 |
| wash_6m | rs17389644 | I2 | 2517 | 2 | 1.14 | [0.99-1.32] | 7.00E-02 | 1.14 | [0.99-1.32] | 7.00E-02 | 0.80 |
| wash_6m | rs17389644 | I3 | 3201 | 2 | 1.10 | [0.98-1.25] | 0.12 | 1.03 | [0.74-1.54] | 0.88 | 0.02 |
| wash_6m | rs17389644 | S1 | 5653 | 2 | 0.99 | [0.81-1.20] | 0.92 | 0.99 | [0.81-1.20] | 0.92 | 0.53 |
| wash_6m | rs17389644 | S2 | 5127 | 2 | 1.06 | [0.86-1.31] | 0.56 | 1.06 | [0.86-1.31] | 0.56 | 0.33 |
| wash_6m | rs17389644 | S3 | 5078 | 2 | 1.03 | [0.84-1.27] | 0.79 | 1.03 | [0.84-1.27] | 0.79 | 0.42 |
| wash_6m | rs17881320 | I1 | 5718 | 2 | 0.90 | [0.67-1.20] | 0.46 | 0.90 | [0.67-1.20] | 0.46 | 0.69 |
| wash_6m | rs17881320 | I2 | 2517 | 2 | 1.13 | [0.92-1.41] | 0.25 | 1.03 | [0.65-1.80] | 0.91 | 0.17 |
| wash_6m | rs17881320 | I3 | 3201 | 2 | 1.00 | [0.83-1.21] | 0.99 | 0.98 | [0.76-1.29] | 0.86 | 0.26 |
| wash_6m | rs17881320 | S1 | 5653 | 2 | 0.89 | [0.67-1.20] | 0.45 | 0.89 | [0.67-1.20] | 0.45 | 0.71 |
| wash_6m | rs17881320 | S2 | 5127 | 2 | 0.98 | [0.72-1.35] | 0.91 | 0.98 | [0.72-1.35] | 0.91 | 0.73 |
| wash_6m | rs17881320 | S3 | 5078 | 2 | 0.96 | [0.70-1.33] | 0.82 | 0.96 | [0.70-1.33] | 0.82 | 0.79 |
| wash_6m | rs2041733 | I1 | 5718 | 2 | 0.87 | [0.74-1.03] | 0.1 | 0.87 | [0.74-1.03] | 0.1 | 0.71 |
| wash_6m | rs2041733 | I2 | 2517 | 2 | 1.10 | [0.98-1.24] | 0.1 | 1.10 | [0.98-1.24] | 0.1 | 0.76 |
| wash_6m | rs2041733 | I3 | 3201 | 2 | 0.96 | [0.87-1.07] | 0.45 | 0.96 | [0.87-1.07] | 0.45 | 0.97 |
| wash_6m | rs2041733 | S1 | 5653 | 2 | 0.87 | [0.74-1.02] | 9.10E-02 | 0.87 | [0.74-1.02] | 9.10E-02 | 0.72 |
| wash_6m | rs2041733 | S2 | 5127 | 2 | 0.88 | [0.74-1.05] | 0.16 | 0.88 | [0.74-1.05] | 0.16 | 0.76 |
| wash_6m | rs2041733 | S3 | 5078 | 2 | 0.88 | [0.74-1.05] | 0.15 | 0.88 | [0.74-1.05] | 0.15 | 0.78 |
| wash_6m | rs2143950 | I1 | 5718 | 2 | 1.00 | [0.81-1.23] | 0.99 | 1.00 | [0.81-1.23] | 0.99 | 0.93 |
| wash_6m | rs2143950 | I2 | 2517 | 2 | 1.12 | [0.96-1.30] | 0.14 | 1.12 | [0.96-1.30] | 0.14 | 0.71 |
| wash_6m | rs2143950 | I3 | 3201 | 2 | 1.08 | [0.94-1.23] | 0.28 | 1.06 | [0.88-1.29] | 0.56 | 0.21 |
| wash_6m | rs2143950 | S1 | 5653 | 2 | 0.99 | [0.80-1.22] | 0.9 | 0.99 | [0.80-1.22] | 0.9 | 0.87 |
| wash_6m | rs2143950 | S2 | 5127 | 2 | 1.01 | [0.81-1.27] | 0.91 | 1.01 | [0.81-1.27] | 0.91 | 0.85 |
| wash_6m | rs2143950 | S3 | 5078 | 2 | 1.00 | [0.79-1.25] | 0.98 | 1.00 | [0.79-1.25] | 0.98 | 0.82 |
| wash_6m | rs2164983 | I1 | 5718 | 2 | 0.99 | [0.80-1.23] | 0.93 | 0.99 | [0.80-1.23] | 0.93 | 0.81 |
| wash_6m | rs2164983 | I2 | 2517 | 2 | 1.21 | [1.03-1.42] | 1.70E-02 | 1.21 | [1.02-1.44] | 3.10E-02 | 0.31 |
| wash_6m | rs2164983 | I3 | 3201 | 2 | 1.15 | [1.00-1.32] | 5.10E-02 | 1.11 | [0.88-1.45] | 0.38 | 0.14 |
| wash_6m | rs2164983 | S1 | 5653 | 2 | 0.99 | [0.80-1.23] | 0.93 | 0.99 | [0.80-1.23] | 0.93 | 0.81 |
| wash_6m | rs2164983 | S2 | 5127 | 2 | 0.98 | [0.78-1.23] | 0.83 | 0.98 | [0.78-1.23] | 0.83 | 0.76 |
| wash_6m | rs2164983 | S3 | 5078 | 2 | 0.98 | [0.77-1.23] | 0.83 | 0.98 | [0.77-1.23] | 0.83 | 0.79 |
| wash_6m | rs2227483 | I1 | 5718 | 2 | 0.95 | [0.81-1.12] | 0.55 | 0.95 | [0.81-1.12] | 0.55 | 0.68 |
| wash_6m | rs2227483 | I2 | 2517 | 2 | 1.11 | [0.99-1.26] | 8.30E-02 | 1.11 | [0.99-1.26] | 8.30E-02 | 0.62 |
| wash_6m | rs2227483 | I3 | 3201 | 2 | 1.06 | [0.96-1.18] | 0.24 | 1.06 | [0.96-1.18] | 0.24 | 0.99 |
| wash_6m | rs2227483 | S1 | 5653 | 2 | 0.96 | [0.82-1.14] | 0.67 | 0.96 | [0.82-1.14] | 0.67 | 0.59 |
| wash_6m | rs2227483 | S2 | 5127 | 2 | 0.94 | [0.79-1.12] | 0.47 | 0.94 | [0.79-1.12] | 0.47 | 0.73 |
| wash_6m | rs2227483 | S3 | 5078 | 2 | 0.96 | [0.80-1.14] | 0.61 | 0.96 | [0.80-1.14] | 0.61 | 0.63 |
| wash_6m | rs2228145 | I1 | 5718 | 2 | 1.11 | [0.94-1.31] | 0.21 | 1.11 | [0.94-1.31] | 0.21 | 0.84 |
| wash_6m | rs2228145 | I2 | 2517 | 2 | 0.95 | [0.85-1.08] | 0.45 | 0.95 | [0.85-1.08] | 0.45 | 0.94 |
| wash_6m | rs2228145 | I3 | 3201 | 2 | 1.05 | [0.95-1.17] | 0.33 | 1.05 | [0.95-1.17] | 0.33 | 0.61 |
| wash_6m | rs2228145 | S1 | 5653 | 2 | 1.11 | [0.94-1.31] | 0.21 | 1.11 | [0.94-1.31] | 0.21 | 0.78 |
| wash_6m | rs2228145 | S2 | 5127 | 2 | 1.11 | [0.93-1.32] | 0.25 | 1.11 | [0.93-1.32] | 0.25 | 0.85 |
| wash_6m | rs2228145 | S3 | 5078 | 2 | 1.11 | [0.93-1.32] | 0.26 | 1.11 | [0.93-1.32] | 0.26 | 0.79 |
| wash_6m | rs2897442 | I1 | 5718 | 2 | 1.03 | [0.86-1.24] | 0.74 | 0.79 | [0.36-2.26] | 0.55 | 0.01 |
| wash_6m | rs2897442 | I2 | 2517 | 2 | 1.10 | [0.96-1.26] | 0.19 | 1.23 | [0.82-1.63] | 0.31 | 0.11 |
| wash_6m | rs2897442 | I3 | 3201 | 2 | 1.11 | [0.99-1.25] | 7.90E-02 | 1.02 | [0.72-1.58] | 0.92 | 0.01 |
| wash_6m | rs2897442 | S1 | 5653 | 2 | 1.04 | [0.86-1.25] | 0.7 | 0.78 | [0.35-2.33] | 0.56 | 0.01 |
| wash_6m | rs2897442 | S2 | 5127 | 2 | 1.07 | [0.88-1.30] | 0.51 | 0.80 | [0.34-2.51] | 0.61 | 0.00 |
| wash_6m | rs2897442 | S3 | 5078 | 2 | 1.08 | [0.88-1.31] | 0.47 | 0.81 | [0.34-2.55] | 0.62 | 0.00 |
| wash_6m | rs479844 | I1 | 5718 | 2 | 0.98 | [0.83-1.16] | 0.82 | 1.05 | [0.76-1.35] | 0.78 | 0.17 |
| wash_6m | rs479844 | I2 | 2517 | 2 | 1.16 | [1.03-1.31] | 1.80E-02 | 1.04 | [0.72-1.68] | 0.83 | 0.08 |
| wash_6m | rs479844 | I3 | 3201 | 2 | 1.12 | [1.01-1.24] | 3.80E-02 | 1.12 | [1.01-1.24] | 3.80E-02 | 0.79 |
| wash_6m | rs479844 | S1 | 5653 | 2 | 0.98 | [0.83-1.16] | 0.81 | 1.05 | [0.75-1.38] | 0.76 | 0.15 |
| wash_6m | rs479844 | S2 | 5127 | 2 | 0.99 | [0.83-1.18] | 0.92 | 1.04 | [0.77-1.34] | 0.79 | 0.19 |
| wash_6m | rs479844 | S3 | 5078 | 2 | 1.00 | [0.84-1.20] | 1 | 1.05 | [0.77-1.36] | 0.74 | 0.19 |
| wash_6m | rs6010620 | I1 | 5718 | 2 | 1.05 | [0.87-1.27] | 0.62 | 1.05 | [0.87-1.27] | 0.62 | 0.33 |
| wash_6m | rs6010620 | I2 | 2517 | 2 | 1.09 | [0.94-1.25] | 0.25 | 1.09 | [0.94-1.25] | 0.25 | 0.60 |
| wash_6m | rs6010620 | I3 | 3201 | 2 | 1.14 | [1.01-1.29] | 3.40E-02 | 1.14 | [1.01-1.29] | 3.40E-02 | 0.44 |
| wash_6m | rs6010620 | S1 | 5653 | 2 | 1.05 | [0.86-1.27] | 0.64 | 1.05 | [0.85-1.29] | 0.64 | 0.31 |
| wash_6m | rs6010620 | S2 | 5127 | 2 | 1.06 | [0.87-1.31] | 0.55 | 1.06 | [0.87-1.31] | 0.55 | 0.37 |
| wash_6m | rs6010620 | S3 | 5078 | 2 | 1.07 | [0.87-1.32] | 0.5 | 1.07 | [0.87-1.32] | 0.5 | 0.36 |
| wash_6m | rs6473227 | I1 | 5718 | 2 | 0.92 | [0.78-1.09] | 0.32 | 1.08 | [0.64-1.55] | 0.78 | 0.04 |
| wash_6m | rs6473227 | I2 | 2517 | 2 | 1.16 | [1.03-1.31] | 1.50E-02 | 1.06 | [0.75-1.64] | 0.76 | 0.10 |
| wash_6m | rs6473227 | I3 | 3201 | 2 | 1.07 | [0.97-1.19] | 0.18 | 1.09 | [0.95-1.23] | 0.24 | 0.23 |
| wash_6m | rs6473227 | S1 | 5653 | 2 | 0.92 | [0.78-1.08] | 0.3 | 1.07 | [0.64-1.52] | 0.8 | 0.04 |
| wash_6m | rs6473227 | S2 | 5127 | 2 | 0.94 | [0.79-1.12] | 0.5 | 1.10 | [0.65-1.59] | 0.73 | 0.04 |
| wash_6m | rs6473227 | S3 | 5078 | 2 | 0.94 | [0.79-1.12] | 0.49 | 1.09 | [0.65-1.57] | 0.75 | 0.05 |
| wash_6m | rs7127307 | I1 | 5718 | 2 | 0.99 | [0.84-1.17] | 0.92 | 0.99 | [0.84-1.17] | 0.92 | 0.42 |
| wash_6m | rs7127307 | I2 | 2517 | 2 | 1.09 | [0.97-1.23] | 0.14 | 1.09 | [0.97-1.23] | 0.14 | 0.68 |
| wash_6m | rs7127307 | I3 | 3201 | 2 | 1.09 | [0.98-1.21] | 9.60E-02 | 1.09 | [0.98-1.21] | 9.60E-02 | 0.44 |
| wash_6m | rs7127307 | S1 | 5653 | 2 | 0.99 | [0.84-1.17] | 0.88 | 0.99 | [0.84-1.17] | 0.88 | 0.38 |
| wash_6m | rs7127307 | S2 | 5127 | 2 | 0.99 | [0.83-1.18] | 0.92 | 0.99 | [0.83-1.18] | 0.92 | 0.39 |
| wash_6m | rs7127307 | S3 | 5078 | 2 | 1.00 | [0.83-1.19] | 0.98 | 1.00 | [0.83-1.19] | 0.98 | 0.39 |
| wash_6m | rs7146581 | I1 | 5718 | 2 | 0.94 | [0.77-1.14] | 0.53 | 0.90 | [0.66-1.28] | 0.5 | 0.22 |
| wash_6m | rs7146581 | I2 | 2517 | 2 | 1.11 | [0.96-1.28] | 0.15 | 1.11 | [0.96-1.28] | 0.15 | 0.61 |
| wash_6m | rs7146581 | I3 | 3201 | 2 | 1.02 | [0.90-1.16] | 0.73 | 0.99 | [0.79-1.27] | 0.91 | 0.12 |
| wash_6m | rs7146581 | S1 | 5653 | 2 | 0.93 | [0.76-1.13] | 0.45 | 0.91 | [0.71-1.19] | 0.46 | 0.28 |
| wash_6m | rs7146581 | S2 | 5127 | 2 | 0.88 | [0.72-1.09] | 0.25 | 0.88 | [0.72-1.09] | 0.25 | 0.36 |

| | | | | | | | | | | | |
|---|---|---|---|---|---|---|---|---|---|---|---|
| wash_6m | rs7146581 | S3 | 5078 | 2 | 0.88 | [0.72-1.09] | 0.25 | 0.88 | [0.72-1.09] | 0.25 | 0.36 |
| wash_6m | rs7927894 | I1 | 5718 | 2 | 1.02 | [0.87-1.20] | 0.78 | 1.02 | [0.87-1.20] | 0.78 | 0.57 |
| wash_6m | rs7927894 | I2 | 2517 | 2 | 1.06 | [0.94-1.20] | 0.33 | 1.06 | [0.94-1.20] | 0.33 | 0.67 |
| wash_6m | rs7927894 | I3 | 3201 | 2 | 1.08 | [0.98-1.20] | 0.13 | 1.08 | [0.98-1.20] | 0.13 | 0.81 |
| wash_6m | rs7927894 | S1 | 5653 | 2 | 1.03 | [0.88-1.22] | 0.69 | 1.03 | [0.88-1.22] | 0.69 | 0.65 |
| wash_6m | rs7927894 | S2 | 5127 | 2 | 1.00 | [0.84-1.19] | 0.97 | 1.00 | [0.84-1.19] | 0.97 | 0.52 |
| wash_6m | rs7927894 | S3 | 5078 | 2 | 1.00 | [0.84-1.20] | 0.97 | 1.00 | [0.84-1.20] | 0.97 | 0.56 |
| wash_6m | | M1 | 8040 | 3 | 1.05 | [0.95-1.15] | 0.34 | 1.02 | [0.88-1.21] | 0.8 | 0.24 |
| wash_6m | | M2 | 7953 | 3 | 1.05 | [0.95-1.16] | 0.33 | 1.03 | [0.90-1.20] | 0.68 | 0.28 |
| wash_6m | | M3 | 7289 | 3 | 1.03 | [0.93-1.14] | 0.61 | 1.01 | [0.87-1.18] | 0.94 | 0.26 |
| wash_6m | | M4 | 7226 | 3 | 1.03 | [0.93-1.15] | 0.53 | 1.01 | [0.88-1.19] | 0.85 | 0.27 |

**Additional file 3: Estimation of statistical power.** Posthoc power calculations performed to facilitate interpretation of negative findings.

| Exposure | SNV or gene | OR $_{GxE}$ (discovery) | N (replication) | Power (replication)* | N (for 0.80 power)* | Interaction effect with 80% power (replication)* |
|---|---|---|---|---|---|---|
| Siblings | rs7146581 | 1.25 | 7,529 | 0.37 | 16,276 | 1.39 |
| Siblings | rs10214237 | 0.84 | 5,049 | 0.13 | 23,624 | 1.48 |
| Dog | rs10214237 | 0.83 | 47,185 | 0.88 | 39,416 | 1.17 |
| Cat | rs16948048 | 0.86 | 49,212 | 0.88 | 41,548 | 1.14 |
| Environmental tobacco smoke 0-2y | *FLG* | 1.33 | 147,880 | 0.99 | 23,272 | 1.12 |
| Environmental tobacco smoke in utero | rs16948048 | 1.15 | 8,078 | 0.11 | 40,952 | 1.36 |
| Siblings | rs7927894 | 1.13 | 7,529 | 0.1 | 42,716 | 1.33 |
| Cat | rs2227483 | 1.13 | 49,212 | 0.73 | 55,456 | 1.14 |
| Cat | rs2228145 | 1.14 | 49,212 | 0.72 | 56,428 | 1.15 |
| Siblings | rs10995251 | 0.89 | 7,529 | 0.08 | 46,248 | 1.34 |
| Cat | rs2897442 | 0.87 | 49,212 | 0.76 | 53,228 | 1.14 |
| Antibiotics in utero | rs10995251 | 1.19 | 2,666 | 0.04 | 30,656 | 1.78 |
| Washing practices 0-2y | *FLG* | 0.71 | 1,061 | 0.03 | 17,192 | 3.77 |
| Duration of breastfeeding † | rs13015714 | 1.02 | 4,252 | 0.04 | 45,040 | 1.08 |
| Breastfeeding ever | rs13015714 | 0.93 | 199,300 | 0.56 | 301,108 | 1.10 |
| **Previously reported interactions** [1] | | | | | | |
| Cat | *FLG* | 11.11 [2] | 48,246 | 0.99 | 516 | 1.26 |
| Siblings | *FLG* | 1.94 [3] | 6,185 | 0.99 | 4,152 | 1.72 |
| Duration of breastfeeding † | *FLG* | 0.99 | 3,056 | 0.006 | 385,008 | 1.08 |
| Breastfeeding ever | *FLG* | 1.29 | 195,879 | 0.99 | 54,184 | 1.15 |

* Replication alpha = 0.004

†power calculation based on GxE estimate per 3.5 SD

[1] Blakeway, H, Van-de-Velde, V, Allen, V. B et al. What is the evidence for interactions between filaggrin null mutations and environmental exposures in the aetiology of atopic dermatitis? A systematic review. *British J Dermatol*, 183(3), 443–451.

[2] Bisgaard H, Simpson A, Palmer CN et al. Gene-environment interaction in the onset of eczema in infancy: filaggrin loss-of-function mutations enhanced by neonatal cat exposure. *PLOS Med* 2008; 5:e131.

[3] Cramer C, Link E, Horster M et al. Elder siblings enhance the effect of filaggrin mutations on childhood eczema: results from the 2 birth cohort studies LISAplus and GINIplus. *J Allergy Clin Immunol*; 125:1254–60.e5. Estimate includes children who attended daycare.

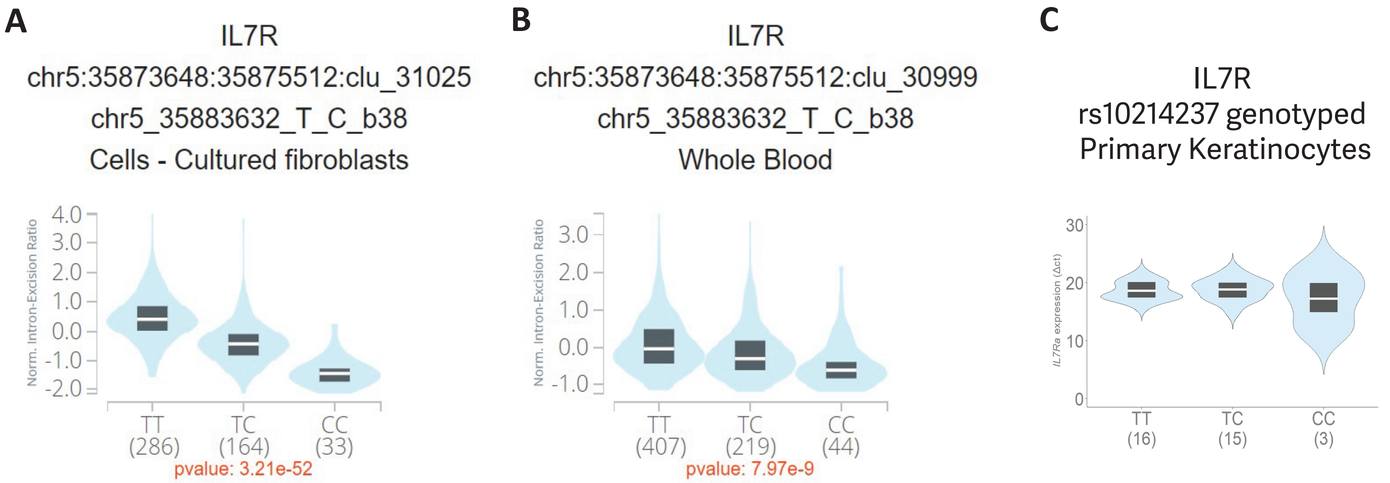
A
IL7R
chr5:35873648:35875512:clu_31025
chr5_35883632_T_C_b38
Cells - Cultured fibroblasts
Norm. Intron-Excision Ratio
4.0
3.0
2.0
1.0
0.0
-1.0
-2.0
TT
(286)
TC
(164)
CC
(33)
pvalue: 3.21e-52
B
IL7R
chr5:35873648:35875512:clu_30999
chr5_35883632_T_C_b38
Whole Blood
Norm. Intron-Excision Ratio
3.0
2.0
1.0
0.0
-1.0
TT
(407)
TC
(219)
CC
(44)
pvalue: 7.97e-9
C
IL7R
rs10214237 genotyped
Primary Keratinocytes
IL7Ra expression (Δct)
30
20
10
0
TT
(16)
TC
(15)
CC
(3)

| Cytokines in ELISA panel (n=28) |
|---|
| Amphiregulin |
| CCL1/I-309 |
| CCL11/Eotaxin |
| CCL17/TARC |
| CCL2/JE/MCP-1 |
| CCL22/MDC |
| CCL27/CTACK |
| CCL28 |
| CCL3/MIP-1 alpha |
| CCL5/RANTES |
| Chemerin |
| CRP |
| CXCL10/IP-10/CRG-2 |
| CXCL11/I-TAC |
| CXCL16 |
| CXCL4/PF4 |
| CXCL9/MIG |
| DPPIV/CD26 |
| Endoglin/CD105 |
| Fas Ligand / TNFSF6 |
| FLT3 (Duo) |
| GM-CSF |
| ICAM-1/ CD54 |
| IFN-gamma |
| IGF-I |
| IGFBP-3 |
| IGFBP-5 |
| IL-1 alpha / IL-1F1 |
| IL-1 beta IL-1F2 |
| IL-10 |
| IL-11 |
| IL-12 p70 |
| IL-17 / IL-17A |
| IL-2 |
| IL-23 (duoset) |
| IL-3 |
| IL-6 |
| Lipocalin-2/NGAL |
| MMP-3 |
| MMP-7 |
| MMP-9 |
| Osteopontin |
| PlGF-2 |
| Trance / Rank L (Duo) |
| Thrombospondin-2 |
| TIMP-1 |
| TNF-alpha |
| TREM-1 |
| VEGF |
| CD80 / B7-1 |
| granzyme B (duoset) |
| CXCL8/IL8 |
| CCL19/MIP-3 beta |
| CCL4/MIP-1 beta |
| CXCL1/GRO alpha |
| IL-12/IL-23 p40 Monomer |
| IL-15 |
| IL-18/IL-1F4 |
| IL-22 |
| IL-31 |
| Lymphotoxin-alpha/TNF-beta |
| IL-33 |
| TSLP |
| IL-38 |

| Cytokine expression detected (n=28) |
|---|
| Amphiregulin |
| CCL1/I-309 |
| CCL2/JE/MCP-1 |
| CCL27/CTACK |
| CCL5/RANTES |
| CXCL10/IP-10/CRG-2 |
| CXCL11/I-TAC |
| CXCL16 |
| CXCL4/PF4 |
| GM-CSF |
| ICAM-1/ CD54 |
| IGFBP-3 |
| IL-1 alpha / IL-1F1 |
| IL-1 beta IL-1F2 |
| IL-6 |
| Lipocalin-2/NGAL |
| MMP-3 |
| MMP-9 |
| PlGF-2 |
| Thrombospondin-2 |
| TIMP-1 |
| TNF-alpha |
| TREM-1 |
| VEGF |
| CXCL8/IL8 |
| CCL4/MIP-1 beta |
| CXCL1/GRO alpha |
| IL-18/IL-1F4 |

| Significant increase in expression (n=9) | p-value |
|---|---|
| TREM-1 | 0.00015 |
| TNF-alpha | 0.003763 |
| CXCL1/GRO alpha | 0.006245 |
| IL-6 | 0.016428 |
| MMP-9 | 0.033438 |
| CXCL10/IP-10/CRG-2 | 0.037395 |
| CCL27/CTACK | 0.040939 |
| CXCL8/IL8 | 0.044355 |
| CXCL11/I-TAC | 0.045492 |

| No change in expression (n=19) | p-value |
|---|---|
| Amphiregulin | >0.05 |
| CCL1/I-309 | >0.05 |
| CCL2/JE/MCP-1 | >0.05 |
| CCL5/RANTES | >0.05 |
| CXCL16 | >0.05 |
| CXCL4/PF4 | >0.05 |
| GM-CSF | >0.05 |
| ICAM-1/ CD54 | >0.05 |
| IGFBP-3 | >0.05 |
| IL-1 alpha / IL-1F1 | >0.05 |
| IL-1 beta IL-1F2 | >0.05 |
| Lipocalin-2/NGAL | >0.05 |
| MMP-3 | >0.05 |
| PlGF-2 | >0.05 |
| Thrombospondin-2 | >0.05 |
| TIMP-1 | >0.05 |
| VEGF | >0.05 |
| CCL4/MIP-1 beta | >0.05 |
| IL-18/IL-1F4 | >0.05 |

**Additional file 5. Results of cytokine, chemokine and receptor expression on human primary keratinocytes following dog allergen exposure.**

**(A)** Proteins not detected; differential expression analysed using paired t-test.

Protein detection by ELISA

***Method***

Primary human keratinocytes were grown to confluency and treated with 500ng/ml dog epithelium (Lofarma, Milan, Italy) for 16 hours. The carrier solution for dog epithelium was used as a negative control (Lofarma, Milan, Italy). Two duplicate wells were treated for each condition, and media then pooled per condition for use in a 64 cytokine DuoSet ELISA assay (cytokines selected based on relevance to atopic or allergic inflammation) (Biotechne, Minneapolis, USA).

The ELISA array spotting was carried out with the Quanterix 2470 Arrayer platform (Quanterix, Billerica, USA) using 185µM pins. Each capture antibody was spotted as four replicate spots on each subarray onto single pad ONCYTE® SuperNOVA nitrocellulose slides (Grace Bio-Labs, Bend, USA). Following array printing, slides were blocked and then incubated with sample overnight at 4°C. After washing and blocking steps each slide was incubated with the appropriate biotin-labelled detection antibody. A final incubation with fluorescently labelled streptavidin followed by slide scanning using an InnoScan710IR scanner (Innopsys, Chicago, USA) generated array images. Images were analysed and signals quantified using Mapix software (Innopsys). The feature (spot) diameter of the grid was set to 270 µm. The average signal intensity was determined for each individual feature and the median background from the adjacent area subtracted from each feature signal leading to a net signal per feature.

**(B)** STRING network analysis of upregulated genes.

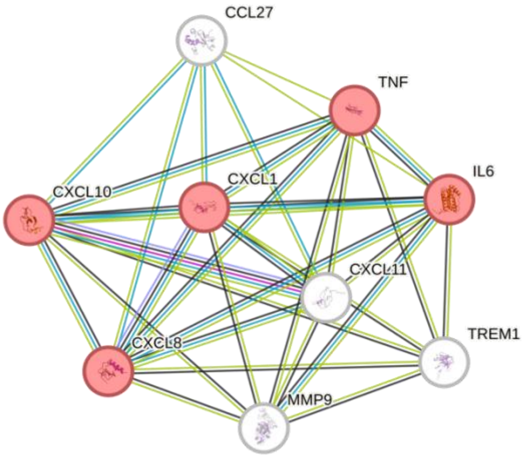

STRING v12.0 network analysis of upregulated genes forms a network with significant enrichment for Reactome Pathway HSA-6783783 IL-10 signalling (5/45 proteins, FDR 2.44e-08).

**(C)** Cytokine expression from primary human keratinocytes.

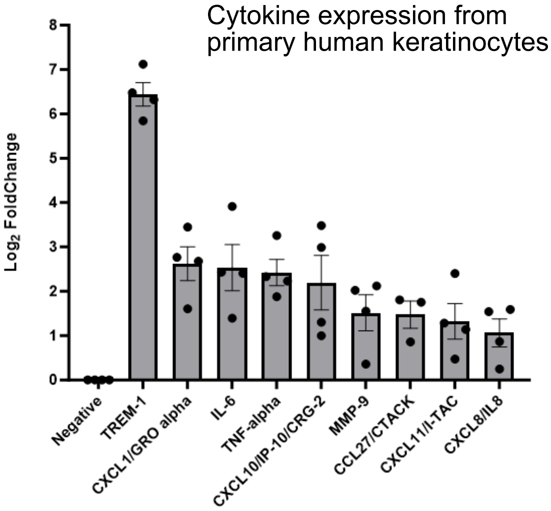

Graph represents the mean log2 fold change in cytokine expression +/- SEM from 3-4 individual human primary keratinocyte donors.

**(D)** STRING network analysis of genes showing no significant change in expression.

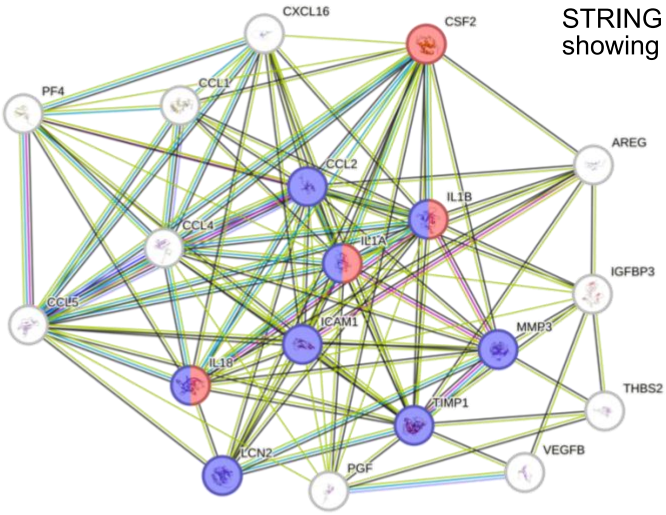

STRING v12.0 network analysis of genes showing no significant change in expression forms a network enriched for Reactome Pathway HSA-6785807 IL-4 and IL-13 signalling (8/107 proteins, FDR 7.93e-11) and local network cluster (STRING) CL:15940 JAK-STAT signalling pathway and IL-1 family (4/138 proteins, FDR 0.0026).

**Additional file 6:**

**Cohort descriptions for 'Gene-environment interaction analysis in atopic eczema: evidence from large population datasets and modelling *in vitro*'**

**Standl *et al.* 2024.**

***ALSPAC***

**Recruitment**

Pregnant women resident in Avon, UK with expected dates of delivery between 1st April 1991 and 31st December 1992 were invited to take part in the study. 20,248 pregnancies have been identified as being eligible and the initial number of pregnancies enrolled was 14,541. Of the initial pregnancies, there was a total of 14,676 foetuses, resulting in 14,062 live births and 13,988 children who were alive at 1 year of age. When the oldest children were approximately 7 years of age, an attempt was made to bolster the initial sample with eligible cases who had failed to join the study originally. As a result, when considering variables collected from the age of seven onwards (and potentially abstracted from obstetric notes) there are data available for more than the 14,541 pregnancies mentioned above: The number of new pregnancies not in the initial sample (known as Phase I enrolment) that are currently represented in the released data and reflecting enrolment status at the age of 24 is 906, resulting in an additional 913 children being enrolled (456, 262 and 195 recruited during Phases II, III and IV respectively). The phases of enrolment are described in more detail in the cohort profile paper and its update (see footnote 5 below). The total sample size for analyses using any data collected after the age of seven is therefore 15,447 pregnancies, resulting in 15,658 foetuses. Of these 14,901 children were alive at 1 year of age. Further details of enrolment have been described previously[1,2]. "Please note that the study website contains details of all the data that is available through a fully searchable data dictionary and variable search tool" and reference the following webpage: http://www.bristol.ac.uk/alspac/researchers/our-data/. Ethical approval for the study was obtained from the ALSPAC Ethics and Law Committee and the Local Research Ethics Committees. Consent for biological samples has been collected in accordance with the Human Tissue Act (2004). Informed consent for the use of data collected via questionnaires and clinics was obtained from participants following the recommendations of the ALSPAC Ethics and Law Committee at the time.

**Definition of eczema cases and controls**

The children have been followed up with regular questionnaires and clinic visits. Data collected from the questionnaires was used to classify children as AD cases or controls. When the children were approximately 81, 91, 103 months, 10, 13, 14 years, parents were asked the following questions [possible answers]:

1. Has your child in the past 12 months had eczema? [Yes and saw a Dr; Yes, but did not see a Dr; No]
2. Has a doctor ever actually said that your child has eczema? [yes; no] (10 & 14 years only)

We defined cases as the children whose parents answered “Yes and saw a Dr” to Q1 or “yes” to Q2. We defined controls as the children who were not a case and whose parents answered “No” to Q2 at 14 years.

**Definition of environmental exposures**

<u>Antibiotics (in utero / up to 6 months / up to 12 months)</u>

At 32 weeks gestation, pregnant mothers were asked if they had taken medication for infection in the last 3 months, where parents answered either “yes” or “no”. When children were approximately 6 months parents were asked if their child had taken antibiotics in the past 6 months. Again at 15 months, parents were asked of the frequency of antibiotic use, where they indicated that the child had never taken antibiotics, or had been administered for 1 episode, or more than 1 episode.

<u>Cesarean section</u>

When children were approximately 8 weeks old mothers were asked if they had delivered via a caesarean section, to which they answered “yes” or “no”.

Cat ownership

At 8 weeks gestation pregnant mothers were asked of the number of cats in their household.

Dog ownership

At 8 weeks gestation pregnant mothers were asked of the number of dogs in their household.

Breastfeeding (duration / ever any / ever exclusive)

When children were approximately 4 weeks old parents were asked if their child was exclusively breastfed during the 2nd, 3rd and 4th week of life, to which they indicated “bottle and breast”, “bottle only”, “breast only” or other.

When children were approximately 4 weeks parents were asked if their child had ever been breastfed to which they answered “yes” or “no”. Again at around 6 months, parents were asked if their child had ever been breastfed where they answered “never”, yes and still breastfeeding, or yes and had stopped breastfeeding.

When children were roughly 6 months parents were asked of the duration of breastfeeding where they indicated “never”, “less than 1 month”, “1 to 3 months”, “3 to 6 months” or “6 or more months”.

Presence of older siblings

When children were approximately 6 months old, parents were asked of the number of other children in the household.

Environmental tobacco smoke exposure (in utero / up to 2 years)

At 18 weeks gestation pregnant mothers were asked if they had smoked tobacco during the first 3 months of pregnancy and in the last 2 weeks. When children were around 8 weeks old mothers were asked if they had smoked cigarettes during the last 2 months of pregnancy and if they had smoked since giving birth.

Washing practices (up to 6 months / up to 2 years)

When children were approximately 6months and 24 months, parents were asked of the frequency that the child bathed or showered.

**Definition of parental education (as a measure of socioeconomic status)**

Parental education was defined as the highest qualification of the mother at 32 weeks pregnant. Levels included “CSE”, “Vocational qualification”, “O-levels”, “A-levels”, “Degree”.

**Definition of family history of atopic disease**

Mothers were asked if they had ever had eczema where they responded “no” or “yes” to having had eczema currently or in the past.

**Genotyping**

ALSPAC children were genotyped using the Illumina HumanHap550 quad chip genotyping platforms by 23andme subcontracting the Wellcome Trust Sanger Institute, Cambridge, UK and the Laboratory Corporation of America, Burlington, NC, US. The resulting raw genome-wide data were subjected to standard quality control methods. Individuals were excluded on the basis of gender mismatches; minimal or excessive heterozygosity; disproportionate levels of individual missingness (>3%) and insufficient sample replication (IBD < 0.8). Population stratification was assessed by multidimensional scaling analysis and compared with Hapmap II (release 22) European descent (CEU), Han Chinese, Japanese and Yoruba reference populations; all individuals with non-European ancestry were removed. SNPs with a minor allele frequency of < 1%, a call rate of < 95% or evidence for violations of Hardy-Weinberg equilibrium (P < 5E-7) were removed. Cryptic relatedness was measured as proportion of identity by descent (IBD > 0.1). Related subjects that passed all other quality control thresholds were retained during subsequent phasing and imputation. 9,115 subjects and 500,527 SNPs passed these quality control filters.

ALSPAC mothers were genotyped using the Illumina human660W-quad array at Centre National de Génotypage (CNG) and genotypes were called with Illumina GenomeStudio. PLINK (v1.07) was used to carry out quality control measures on an initial set of 10,015 subjects and 557,124 directly genotyped SNPs. SNPs were removed if they displayed more than 5% missingness or a Hardy-Weinberg equilibrium P value of less than 1.0E-06. Additionally, SNPs with a minor allele frequency of less than 1% were removed. Samples were excluded if they displayed more than 5% missingness, had indeterminate X chromosome heterozygosity or extreme autosomal heterozygosity. Samples showing evidence of population stratification were identified by multidimensional scaling of genome-wide identity by state pairwise distances using the four HapMap populations as a reference, and then excluded. Cryptic relatedness was assessed using a IBD estimate of more than 0.125 which is expected to correspond to roughly 12.5% alleles shared IBD or a relatedness at the first cousin level. Related subjects that passed all other quality control thresholds were retained during subsequent phasing and imputation. 9,048 subjects and 526,688 SNPs passed these quality control filters.

We combined 477,482 SNP genotypes in common between the sample of mothers and sample of children. We removed SNPs with genotype missingness above 1% due to poor quality (11,396 SNPs removed) and removed a further 321 subjects due to potential ID mismatches. This resulted in a dataset of 17,842 subjects containing 6,305 duos and 465,740 SNPs (112 were removed during liftover and 234 were out of HWE after combination). We estimated haplotypes using ShapeIT (v2.r644) which utilises relatedness during phasing. The phased haplotypes were then imputed to the Haplotype Reference Consortium (HRCr1.1, 2016) panel of approximately 31,000 phased whole genomes. The HRC panel was phased using ShapeIt v2, and the imputation was performed using the Michigan imputation server.

This gave 8,237 eligible children and 8,196 eligible mothers with available genotype data after exclusion of related subjects using cryptic relatedness measures described previously.

**Acknowledgments and Funding**

We are extremely grateful to all the families who took part in this study, the midwives for their help in recruiting them, and the whole ALSPAC team, which includes interviewers, computer and laboratory technicians, clerical workers, research scientists, volunteers, managers, receptionists and nurses.

The UK Medical Research Council and Wellcome (Grant ref: 217065/Z/19/Z) and the University of Bristol provide core support for ALSPAC. This publication is the work of the authors and LP will serve as guarantors for the contents of this paper.

Genome-wide genotyping data was generated by Sample Logistics and Genotyping Facilities at Wellcome Sanger Institute and LabCorp (Laboratory Corporation of America) using support from 23andMe. A comprehensive list of grants funding is available on the ALSPAC website (http://www.bristol.ac.uk/alspac/external/documents/grant-acknowledgements.pdf).

### *COPSAC2000 & COPSAC2010*

#### Recruitment

The COPSAC2000 birth cohort study is a prospective clinical study of a birth cohort of 411 infants born to mothers with a history of asthma. The newborns were enrolled at the age of 1 month, the recruitment of which was previously described in detail (1-3). The study was approved by the Ethics Committee for Copenhagen (KF 01-289/96) and The Danish Data Protection Agency (2008-41-1754) and informed consent was obtained from both parents.

COPSAC2010 is a mother child cohort comprising 700 children born to unselected mothers from Denmark as described previously in detail (4). The study is conducted in accordance with the Declaration of Helsinki and was approved by the Danish Ethics Committee (H-B-2008-093) and the Danish Data Protection Agency (2008-41-2599).

#### Definition of eczema cases and controls

The families used doctors employed at the clinical research unit, and not the family practitioner, for diagnosis and treatment of AD and other skin-related symptoms. Skin lesions were described at both scheduled visits at 6-monthly intervals and acute visits with skin symptoms according to pre-defined morphology and localization; AD was defined based on the Hanifin-Rajka criteria as previously detailed (5).

Children without any eczema diagnosis were used as controls. Cases from the COPSAC2000 dataset is based on an eczema diagnosis during the first 17 years of life. Cases from the COPSAC2010 dataset is based on an eczema diagnosis during the first 7 years of life.

#### Definition of environmental exposures

<u>Antibiotics (in utero / up to 6 months / up to 12 months)</u>

Use of antibiotics in utero was determined if the mother had used any antibiotics during one of the three trimesters. The 6- and 12-months variables were made similar using medical prescription history for the child. Antibiotic was classified using the following atc codes: J01+P01AB01 and J01+G04A C01.

<u>Cesarean section</u>

Mode of delivery is based on the mother's information and validated using medical records. 1 indicate that a cesarean section occurred, regardless of it being planned or acute. Natural births are 0.

<u>Cat ownership</u>

Using data on how many days a child was exposed to a cat at home during the first year of life. Exposure is being in the same room if indoor, touching if outdoor. Children with recordings of a cat in their home during the first year of life was set to 1, else 0.

<u>Dog ownership</u>

Similar to the cat ownership variable.

<u>Breastfeeding (duration / ever any / ever exclusive)</u>

Based on information from the mother. The duration of breastfeeding was calculated as the difference between the birthday and the reported end date for breastfeeding. The ever-breastfeeding variable is 1 if the difference between the birthday and the breastfeeding end date is more than 7 days. If the children were breastfed more than 7 days, then the exclusive breastfed variable is 1 otherwise it is 0.

Presence of older siblings

Sibling information is based on whether any older children is in the household. The presence of older children isn't' necessarily blood relatives of the children.

Environmental tobacco smoke exposure (in utero / up to 2 years)

Passive smoking for the child in the first 2 years is based on the parent's own answer to "Days of smoking" and "child exposed to passive smoking". For each child we have the number of days with exposure to passive smoking, and the binary variable is 1 if the child had any passive smoke exposure, and 0 otherwise. The in-utero exposure is based on whether the mother was actively smoking during pregnancy. If any active smoking was recorded, the variable was set to 1, otherwise 0.

**Definition of parental education (as a measure of socioeconomic status)**

The educational definition is based on the mother's longest completed education at time of 6-year visit. If the mothers had completed university, then these are coded as 1, otherwise 0.

**Genotyping**

COPSAC cohorts were genotyped on the Illumina HumanOmniExpressExome 8 v1-2 BeadChip (951,117 SNPs) at the AROS Applied Biotechnology AS center, Aarhus, Denmark. SNP genotype calling for COPSAC cohorts was performed using the GenCall [51] followed by zCall [52]. SNPs with a) minor allele frequency (MAF) > 0.01, b) cluster separation score ≥ 0.3, c) individual call rate > 99%, d) SNP call rate > 99% and e) Hardy Weinberg Equilibrium (HWE) p-values > 1e-6 were included. Individuals with a sex mismatch along with genetic duplicates or related individuals were excluded using identity by descent (IBD) analyses. In addition, heterozygosity outliers (>0.37 and <0.27) were removed from the analysis. Individuals not clustering with the European ancestry (Utah residents with ancestry from Northern or Western Europe (CEU)) were excluded using the multidimensional scaling analyses seeded with individuals from the International HapMap 3 project.

**Acknowledgments and Funding**

We express our deepest gratitude to the children and families of the COPSAC2000 and COPSAC2010 cohorts' study for all their support and commitment. We acknowledge and appreciate the unique efforts of the COPSAC research team**.**

All funding received by COPSAC is listed on www.copsac.com. The Lundbeck Foundation (Grant no R16-A1694); The Ministry of Health (Grant no 903516); Danish Council for Strategic Research (Grant no 0603-00280B) and The Capital Region Research Foundation have provided core support to the COPSAC research center.

### *Danish National Birth Cohort (DNBC)*

**Recruitment**

Cited from previous paper:[1]

The Danish National Birth Cohort (DNBC) is a population-based cohort of more than 100,000 pregnancies, recruited in the years 1996-2002[2] Extensive phenotype information was collected by computer-assisted telephone interviews twice during pregnancy as well as 6 and 18 months after delivery. Additional questionnaire-based follow-up surveys are conducted at regular intervals. The DNBC mothers provided written informed consent on behalf of themselves and their children. The study protocol was approved by the Danish Scientific Ethical Committee and the Danish Data Protection Agency.

**Definition of eczema cases and controls**

Cited from previous paper:[1]

Cases with early onset eczema were identified from the 18 months telephone interview data using an algorithm specifically developed for this purpose.[3] In addition, children with a positive response to both of the following two questions from the 7 year survey were included in the case group: 1) "Has a doctor ever said that your child had eczema, also known as allergic rash?" and 2) "Has your child ever had an itchy *rash which was coming and going for at least 6 months?*". Finally, children with ICD10 diagnosis code L20 in the Danish Hospital Discharge Register were also included in the case group. Controls were required not to have any eczema or eczema symptoms recorded in interview, questionnaire, or register data.

**Definition of environmental exposures**

<u>Cesarean section</u>

Information on mode of delivery (specifically cesarean and vaginal delivery) was obtained by person-linkage of the DNBC with the Danish Medical Birth Register. Since 1973, the Medical Birth Register has contained nationwide computerized information about birth characteristics, including mode of delivery, which are required to be recorded shortly after birth.[4]

<u>Cat and/or dog ownership</u>

Children with cat and/or dog ownership since birth was identified from the 18 months telephone interview data, as children whose mothers chose "cat(s)" and/or "dog(s) as affirmative answer to the question (D033) "What animals or pets have you had since his/her birth" excluding "Animals not in contact with the child" or "Animals kept outside".

<u>Breastfeeding (duration / ever any / ever exclusive)</u>

Information on breastfeeding was based on responses to the following questions from the 6 months telephone interview data, available for >70% of cases and controls: (C001) "Do you breast feed your boy/girl now?", (C004) "How old was your child when you stopped breast feeding her/him every day?" (C003) "For how long have you been breast feeding him/her without giving him/her anything else except for water and vitamins?"

<u>Presence of older siblings</u>

Information on presence of older siblings at birth was obtained from the Danish family relations database, which is based on individual kinship information in the Danish Civil Registration system, and permits the identification of relatives for persons with family in Denmark, e.g. siblings to all persons born since 1950.[5]

Environmental tobacco smoke exposure (in utero/up to 2 years)

Exposure up to 18 months (~2 years) was based on positive responses to the following questions from the 6 and/or 18 months telephone interview data, respectively: (C062) "Smoking in the home while the child is present?" and/or (D031) "After your child has turned 6 months old, has there been anyone smoking daily in your home (for periods of at least one week) while the child was present?".

In utero exposure was based on positive responses to the following questions in the telephone interview data from twice in pregnancy and at 18 months of age: ~ Week 12, (A127) "Did you smoke during pregnancy – please also think back to the very beginning of the pregnancy? ", (A128) "Do you smoke now?"; ~week 30, (B074) "Have you smoked since last interview?", (B075) "Do you smoke now?"; child aged 18 months, (P094) "Did you smoke during the last part of pregnancy or after the birth?" (yes, last part).

**Genotyping and imputation**

Cited from previous paper:[1]

GWAS data were generated for 3,840 individuals from the DNBC (mothers and their children) in a study of prematurity and its complications within the Gene Environment Association Studies (GENEVA) consortium. Genotyping was performed using the Illumina Human660W-Quad BeadChip. Prior to imputation, we required participants to have a genotype call rate >97%, and we excluded SNPs based on a missing rate >2%, deviation from Hardy-Weinberg equilibrium in controls ($P<10^{-3}$), minor allele frequency <0.5%. We also converted the genotype data from NCBI build 36 to NCBI build 37 and aligned all genotypes to the forward strand. Finally, we excluded SNPs that did not match known variant positions in the 1000 Genomes project reference data. The remaining 529128 SNPs were used for imputation.

We used a two-step procedure to impute unobserved genotypes using phased haplotypes from the integrated Phase I release of the 1000 Genomes Project[6] (v3.20101123, ALL populations, no monomorphic/singletons). In a first prephasing step, we used SHAPEIT[7] to estimate the haplotypes for our study samples. In a second step, we imputed the missing alleles for additional SNPs directly onto these phased haplotypes using IMPUTE2[8]. Eczema information and genome-wide genotype and imputed data were available for 1,631 children.

Four filaggrin mutations (R501X, 2282del4, R2447X and S3247X) were genotyped by LGC Genomics using KASP™ genotyping technology, as described for ALSPAC.

Statistical Analysis

Cited from previous paper:[1]

Genome-wide association analysis was carried out using SNPTEST[9] for the 224 cases and 1407 controls with genetic and phenotypic data. Summary statistics were available for 30,071,690 variants that were successfully analyzed.

**Acknowledgments and Funding**

Cited from previous paper:[1]

We are very grateful to the women and children taking part in the DNBC. The DNBC was established with the support of a major grant from the Danish National Research Foundation. Additional support for the DNBC has been obtained from the Danish Pharmacists' Fund, the Egmont Foundation, the March of Dimes Birth Defects Foundation, the Augustinus Foundation and the Health Fund of the Danish Health Insurance Societies. The generation of GWAS genotype data for the DNBC samples was carried out within the GENEVA consortium, with funding provided through the NIH Genes, Environment and Health Initiative (GEI) (U01HG004423, U01HG004438, U01HG004446). FLG genotyping was funded by an MRC centenary award (awarded to L Paternoster).

### *Generation R*

**Recruitment**

The Generation R Study is a population-based prospective cohort study of pregnant women and their children from fetal life onwards in Rotterdam, The Netherlands. All children were born between April 2002 and January 2006, and currently followed until young adulthood. Of all eligible children in the study area, 61% were participating in the study at birth.

**Definition of eczema cases and controls**

Postnatal data about eczema was collected via questionnaires at the ages of 1 to 5 years, and at the age of 9 years. Cases of eczema were defined by answering 'Yes, my child has eczema and has seen a doctor' to the questions 'Has your child in the past 6 months or 12 months had eczema?' at age 6 months, or age 1, 2, 3, 4 and 9 years, respectively. We defined controls as those children of whom parents answered that their child 'Yes, had eczema, but did not see a doctor/No, did not have eczema' at the same ages as cases of eczema.

**Definition of environmental exposures**

<u>Antibiotics (during pregnancy / up to 6 months / up to 12 months)</u>

Via questionnaires.

Have you used antibiotic during pregnancy? (y/n)

Has your child ever used antibiotic/penicillin at the age of age 1 year? (y/n)

Has your child ever used antibiotic/penicillin at the age of age 6 months? ( y/n)

<u>Cesarean section</u>

Information on mode of  delivery was obtained from midwife and hospital records at birth. Mode of delivery was categorized as (1) vaginal delivery or (2) caesarean section.

<u>Cat ownership</u>

Via a questionnaire at enrolment.

Do you have a cat at home? (y/n)

<u>Dog ownership</u>

Via questionnaires at enrolment.

Prenatal. Do you have a dog at home? (y/n)

<u>Breastfeeding (duration / ever any / ever exclusive)</u>

Detailed information on breastfeeding initiation and continuation was obtained from parental postal questionnaires at the ages of 2, 6 and 12 months after birth. Mothers were asked whether they ever breastfed their child (no; yes) and at what age (in months) they stopped breastfeeding.

<u>Presence of older siblings</u>

Via a questionnaires at enrolment.

How many living children have you given birth to? (none or 1 or more).

<u>Environmental tobacco smoke exposure (during pregnancy / up to 2 years)</u>

Via multiple questionnaires during pregnancy and at the age of 2 years.

Do you smoke (never, until pregnancy was known/continuously during pregnancy)? (y/n

Do people smoke occasionally in your house? (y/n)

Air pollution ($NO_2$, $PM_{10}$)

Individual exposures to $PM_{10}$ and $NO_2$ between 0 and 6 months after birth were assessed at the home address, using a combination of continuous monitoring data and dispersion modeling techniques, taking into account both the spatial and temporal variation in air pollution (van den Hooven, de Kluizenaar, Pierik, Hofman, van Ratingen, ZandveldAir pollution, blood pressure, and the risk of hypertensive complications during pregnancy: the generation R study).

**Definition of parental education (as a measure of socioeconomic status)**

Maternal education was asked by a questionnaire at enrollment (primary or secondary school versus higher than secondary school).

**Definition of family history of atopic disease**

Parental history of eczema, allergy or asthma (no; yes) was available from a parental questionnaire obtained at enrolment.

**Genotyping and Imputation**

The most prevalent FLG mutations in Caucasians (2282del4, R2447X, R501X, and S3247X) were genotyped by modified Taqman allelic discrimination assays, using previously described primers (Irvine, McLean and Leung Filaggrin mutations associated with skin and allergic diseases; Kezic, O'Regan, Yau, Sandilands, Chen, Campbell Levels of filaggrin degradation products are influenced by both filaggrin genotype and atopic dermatitis severity). Children without any mutant alleles were classified as wild type. Information on single nucleotide polymorphisms (SNPs) was available from the GWA screen performed on DNA isolated from cord blood leukocytes and genotyped in the Illumina HumanHap 610 microarray or, in a small minority of children with missing cord blood samples, at age 6 years using the Illumina 670K microarray. Stringent quality control of the genotype data was performed (Medina-Gomez, Felix, Estrada, Peters, Herrera, Kruithof Challenges in conducting genome-wide association studies in highly admixed multi-ethnic populations: the Generation R Study). Imputation to the HRC1.1. panel was performed in the MICHIGAN server using Phasing:Eagle v2.3/ imputation minimac 3.

**Analysis**

Preparation of input files and association analysis were performed following the distributed analysis plan. The analysis was run using rvtest adjusting for sex and 20 genomic principal components (as calculated by multi-dimentional scaling in PLINK as previously described (Medina-Gomez C, Felix JF, Estrada K, Peters MJ, Herrera L, Kruithof CJ, et al. Challenges in conducting genome-wide association studies in highly admixed multi-ethnic populations: the Generation R Study. Eur J Epidemiol. 2015;30(4):317-30.)).

**Acknowledgments and Funding**

The Generation R Study is conducted by the Erasmus Medical Center in close collaboration with the School of Law and Faculty of Social Sciences of the Erasmus University Rotterdam, the Municipal Health Service Rotterdam area, Rotterdam, the Rotterdam Homecare Foundation, Rotterdam, and the Stichting Trombosedienst & Artsenlaboratorium Rijnmond (STAR-MDC), Rotterdam. We gratefully acknowledge the contribution of children and parents, general practitioners, hospitals, midwives, and pharmacies in Rotterdam. The Generation R Study is made possible by financial support from the Erasmus Medical Center, Rotterdam, the Erasmus University Rotterdam and the Netherlands Organization for Health Research and Development. The study received funding from the European Union's Horizon 2020 research and innovation programme (LIFECYCLE, grant agreement No 733206, 2016; EUCAN-Connect grant agreement No 824989; ATHLETE, grant agreement No 874583; ENDOMIX, grant agreement No 101136566). Prof. Tamar Nijsten and Liesbeth Duijts received funding from Nestlé Skin Health - Galderma R&D (grant agreement no 35195; 2016).

### *GINIplus / LISA*

#### Recruitment

The influence of Life-style factors on the development of the Immune System and Allergies in East and West Germany (LISA) Study is a population-based birth cohort study. A total of 3094 healthy, full-term neonates were recruited between 1997 and 1999 in Munich, Leipzig, Wesel and Bad Honnef. The participants were not pre-selected based on family history of allergic diseases.

A total of 5991 mothers and their newborns were recruited into the German Infant study on the influence of Nutrition Intervention PLUS environmental and genetic influences on allergy development (GINIplus) between September 1995 and June 1998 in Munich and Wesel. Infants with at least one allergic parent and/or sibling were allocated to the interventional study arm investigating the effect of different hydrolysed formulas for allergy prevention in the first year of life. All children without a family history of allergic diseases and children whose parents did not give consent for the intervention were allocated to the non-interventional arm. Detailed descriptions of the LISA and GINIplus studies have been published elsewhere [DOI: 10.5414/ALX01455E and DOI: 10.1038/s41467-023-41180-2]. DNA was collected at the age 6 and 10 years. For both studies, approval by the local Ethics Committees and written consent from participant's families were obtained.

The data from the Munich study center was included in the discovery step and the data from the Wesel study center was part of the replication.

#### Definition of eczema cases and controls

Information on ever having physician-diagnosed eczema was collected using self-administered questionnaires completed by the parents. The questionnaires were completed at 6, 12, 18 and 24 months and 4, 5, 6, 10 and 15 years of age in the LISA study and 1, 2, 3, 4, 6 and 10 and 15 years in the GINIplus study asking for each year of age since the previous follow-up. Cases were defined as subjects who reported having a diagnosis at any time point, and controls were defined as those reporting no diagnosis at every time point.

#### Definition of environmental exposures

Antibiotics in utero

Information on maternal use of antibiotics during pregnancy was collected at birth in the LISA study.

Cesarean section

Information whether the child was delivered by cesarean section was collected at birth.

Cat ownership

In the GINIplus cohort, cat ownership was defined as having a cat in the household between birth and one year of age. In LISA, cat ownership was defined as having a cat in the household at birth.

Dog ownership

In the GINIplus cohort, dog ownership was defined as having a dog in the household between birth and one year of age. In LISA, dog ownership was defined as having a dog in the household at birth.

Breastfeeding (duration / ever any / ever exclusive)

Information on exclusive breastfeeding, exclusive bottle feeding or mixed feeding was available for each of the first six months of life. Breastfeeding duration was defined as the number of months receiving any breastmilk. Ever any breastfeeding was defined as receiving any breastmilk for at least one month of life and ever exclusive breastfeeding was defined as being exclusively breastfed for at least one month.

Presence of older siblings

Presence of older siblings was defined as having any biological siblings at birth.

Environmental tobacco smoke exposure (in utero / up to 2 years)

Environmental tobacco smoke exposure in utero was defined as active cigarette smoking by the mother in any pregnancy trimester (only LISA).

Information on any smoking in the household was used to define environmental tobacco smoke exposure up to 2 years. In LISA, this information was collected every 6 months from birth to 2 years and any smoking was defined as yes. In GINIplus, this information was collected for the first 4 months of life and from ages 13-24 months, with the time period from 5-12 months only covered for the intervention arm.

Air pollution (NO2, PM10)

The annual average concentrations of the air pollutants NO2 and PM10 at participants' residential addresses were derived from land-use regression models assigned within the European Study of Cohorts for Air Pollution Effects (ESCAPE).

House dust mite: Der p1 concentration (at birth / 1 year)

House dust mite concentrations were measured in dust collected shortly after birth in the LISA cohort.

**Definition of parental education (as a measure of socioeconomic status)**

Parental education level was defined as the highest degree of education (defined as the number of school years) received by either the mother or the father.

**Genotyping**

*Munich study center*

1511 children from Munich from both studies were included (835 (55%) children from the GINIplus study and 676 (45%) children from the LISA study). 1423 individuals (835 from the GINIplus study and 588 from the LISA study) were analyzed using the Affymetrix Human SNP Array 5.0 and 88 individuals from the LISA study were analyzed using Affymetrix Human SNP Array 6.0. Genotypes were called using BRLMM-P algorithm (5.0), respectively BIRDSEED V2 algorithm (6.0). In each of the two data sets, criteria for exclusion of individuals were: a call rate below 95%, a heterozygosity outside mean +/- 4sd, a failure of the sex check or a failure of the similarity quality control using MDS analysis based on IBS. Criteria for exclusion of variants were: a call rate below 95%, a MAF < 0.01 and a HWE p-value < 0.00001. The filtered data sets were prephased using SHAPEIT V2 and imputation was done using IMPUTE2.3 considering the haplotypes from the 1000 Genomes Project Phase I v3 as a reference (March 2012 release, updated version from 26 Aug 2012, all ancestries, limited to variants with more than one minor allele copy).

Genotyping was performed for the two most common variants of the filaggrin gene (*FLG*), R501X and 2282del4. Participants with at least one FLG mutation were classified as having the FLG loss-of-function mutation. Participants with missing data for a particular variant were excluded if the remaining variant was wild-type.

*GINIplus/LISA North*

Genome-wide genotyping provided by the German Research Center for Environmental Health at the Helmholtz Zentrum München in April 2019 was performed in 883 blood samples (collected in follow-up at age 10 years) using the Infinium Global Screening Array GSA v2 MD (GRCh37/hg19) with GenomeStudio Version 2.0.

Pre-imputation Quality Control in R 3.6.1 included (Reed et al. 2015 DOI: 10.1002/sim.6605):

1) SNP level 1: Exclusion of variats on chromosome 0, insert/deletion variants, MAF (<0.01), and call rates (<0.95)

2) Sample level: Removal of duplicated individuals, individuals with sex-mismatch, call rates (<0.95), heterozygosity (iinbreeding coefficient 0.1), highly related individuals (identity-by-descent analysis with ld.tresh=0.2 and kin.tresh=0.1), individuals belonging to different ancestry group (Tukey's rule based on the 1-10 eigenvectors from Principal Component Analysis), and Hardy-Weinberg ($p<10^{-6}$)

3) SNP level 2: Hardy-Weinberg ($p<10^{-6}$)

4) Strand designation/ strand flips correction

To find haplotype segments that are shared by study individuals and the HRC r1.1 2016 (GRCh37/hg19), we did a genotype imputation with minimac4 using the Michigan Imputation Server (settings: European population, no rsq filter, eagle v2.4 phasing in output, and no AES 256 encryption).

**Acknowledgments and Funding**

M.S. has received funding from the European Research Council (ERC) under the European Union's Horizon 2020 research and innovation programme (Grant Agreement No. 949906).

### *INMA*

#### Recruitment

Population-based birth cohorts were established from 1997 to 2008 as part of the INMA – INfancia y Medio Ambiente (Environment and Childhood) Project in several regions of Spain following a common protocol. INMA aims to study the associations between pre- and postnatal environmental exposures and growth, health and development from early life until adolescence and has been described previously in detail (Guxens, M. *et al.* Cohort Profile: the INMA--INfancia y Medio Ambiente--(Environment and Childhood) Project. *Int J Epidemiol* **41**, 930-40 (2012). Pregnant women were enrolled during the 1st trimester of pregnancy at public primary health care centers or public hospitals. Detailed measurements were performed using ultrasound and physical examinations and biological samples were collected. Informed consent was obtained from all participants and the study was approved by the Hospital Ethics Committees in each participating region.

#### Definition of eczema cases and controls

Children from the subcohorts of INMA Gipuzkoa, Menorca, Sabadell and Valencia were included in the present study. Atopic eczema cases were those children that had answered "yes" to having had eczema at least in one of the three follow-up visits (14 months, 48 months and 7 years). Control children were those that had never had eczema. Questions were slightly different among subcohorts (**Table**).

**Table**. Eczema information in each subcohort and follow-up visit.

| Follow-up visit | Questionnaire | Menorca | Valencia | Sabadell | Gipuzkoa |
|---|---|---|---|---|---|
| 14 months | Has your child had atopic eczema? (0-14 months) | | X | | |
| | Has your child ever been diagnosed by doctor with having atopic eczema? | X | | X | X |
| 48 months | In the last 12 months, has your child had ever suffered atopic eczema? (36-48 months) | | | X | |
| | Doctor diagnosis of eczema | X | | | |
| | Has your child had suffered atopic eczema? | | X | | X |
| 7 years | Doctor diagnosis of eczema | X | X | X | X |

#### Definition of environmental exposures

<u>Antibiotics (in utero / up to 6 months / up to 12 months)</u>

The use of antibiotics was obtained using interviewer-administered questionnaires with the mothers during pregnancy at 12 and 32 weeks (Valencia, Sabadell and Gipuzkoa), and when the children were 6 mo (only Sabadell) and 1 yo (Menorca, Sabadell and Gipuzkoa). It was defined as yes or no in utero, at 6 mo and at 12mo

<u>Cesarean section</u>

Information on delivery by Cesarean sections was obtained from clinical records at birth for all cohorts and defined as y/n.

Cat ownership:

Cat ownership was obtained from interviewer-administered questionnaires with the mothers during pregnancy (Menorca) and at 32 weeks (Valencia, Sabadell and Gipuzkoa), and was defined as y/n owning a pet in utero.

Dog ownership

Cat ownership was obtained from interviewer-administered questionnaires with the mothers during pregnancy (Menorca) and at 32 weeks (Valencia, Sabadell and Gipuzkoa), and was defined as y/n owning a pet in utero.

Breastfeeding (duration / ever any / ever exclusive)

Detailed information about the child's diet, including breastfeeding, was obtained using interviewer-administered questionnaires with the mothers when children were 6 mo old (only Sabadell sub-cohort) and 14 mo (all sub-cohorts).

For our analysis breastfeeding was defined in three different way: (i) ever breastfeeding y/n, (ii) predominant breastfeeding y/n and (iii) duration of any breasfeeding in weeks.

NB: Predominant breastfeeding Accept water, juice and infusion; exclussive breastfeeding was missing for VALENCIA as water introduction was not available

Presence of older siblings

The presence of older siblings was obtained using interviewer-administered questionnaires with the mothers during pregnancy and was defined as yes or no.

Environmental tobacco smoke exposure (in utero / up to 2 years)

Information on environmental tobacco smoke exposure in the household was obtained using interviewer-administered questionnaires with the mothers during pregnancy (Menorca) and at 32 weeks (Valencia, Sabadell, and Gipuzkoa), and when the children were 1yo (Valencia, Sabadell, Gipuzkoa and Menorca) and was defined as y/n at any moment during the first year of life.

Washing practices (up to 6 months / up to 2 years)

Detailed information about the child's showering/bathing frequency was obtained using interviewer-administered questionnaires with the mothers when the children were 14 mo old for all sub-cohorts. Current (as a proxy for up to 2 years) and first months of life (as a proxy for up to 6 months) washing practices were asked separately, and was defined as y/n comparing daily or more frequent versus less than showering daily.

Air pollution (NO2, PM10)

Air pollution concentrations at each participant's home address at birth were estimated for the whole pregnancy period by land-use regression models following the ESCAPE standardized procedure). Briefly, air pollution monitoring campaigns were performed in the study areas between October 2008 and January 2011. In all areas, three week measurements of NO2 were performed within 1 year. In Sabadell, simultaneous measurements of PM10 were performed. Land-use regression models were developed for each pollutant metric using all measurement sites to estimate annual average air pollution concentration at each participants' home address at birth. We used a back-extrapolation procedure to estimate pregnancy-average concentrations from annual average concentrations using routine data from background sites of air quality monitoring networks in the study areas. Air pollutants are described in ug/m3 and the analyses were performed for an increase of 10 ug/m3.

House dust mite: Der p1 concentration (at birth / 1 year)

Dust samples were collected from children's mattresses during the first three months of life. House dust mite allergen (Der p1) concentration was estimated in a single laboratory by enzyme-linked immunoabsorbent assay (ELISA) following an standard protocol. Der p1 concentration is described in µg/g.

**Definition of parental education (as a measure of socioeconomic status)**

Highest level of education attained by the child's mother and father, that is, primary or less than primary (reference category), secondary or higher education.

**Definition of family history of atopic disease**

Information on family history of atopic disease was obtained using interviewer-administered questionnaires with the mothers during pregnancy in all cohorts, including information on parental asthma, atopic dermatitis, and atopic rhinitis. It was defined as y/n.

**Genotyping**

DNA was obtained from cord blood, whole blood collected at 4y or saliva using the Chemagen protocol at the Spanish National Genotyping Centre (CEGEN), Spain. Children of European ancestry whose parents reported to be born in Spain or in European countries and that were not lost during the follow-up were selected for genotyping. Genome-wide genotyping was performed using the HumanOmni1-Quad Beadchip (Illumina) at CEGEN (Menorca, Sabadell, Valencia subcohorts) and Infinium Global Screening Array (GSA) Beadchip (Illumina) at the Human Genotyping Facility (HuGeF), Erasmus MC, The Netherlands (Gipuzkoa subcohort). Genotype calling was done using the GeneTrain2.0 algorithm based on HapMap clusters implemented in the GenomeStudio software. Quality control was done using PLINK and following standard criteria. First of all, SNPs were flipped to the human genome + strand. We applied the following initial quality control thresholds: sample call rate >98% and/or Log R Ratio (LRR) standard deviation (SD) <0.3. Then, we checked sex, relatedness, heterozygosity and population stratification. Genetic variants were filtered for SNP call rate >95%, MAF >1% and Hardy-Weinberg equilibrium P-value >1.10E-06. Imputation of genetic variants was done using IMPUTE V2 and the cosmopolitan 1000G panel (release March 2012) (Menorca, Sabadell, Valencia) and the Michigan imputation server using the Haplotype Reference Consortium HRC v1.1 reference panel (Gipuzkoa).

Candidate single nucleotide polymorphisms (SNPs) were retrieved from genome-wide genotyping data and their genotypic frequencies were checked between genotyping platforms (Omni1 vs. GSA). No substantial differences were observed and data from the different subcohrots was pooled and models were adjusted by a variable including the cohort (Sabadell+Valencia+Menorca vs. Gipuzkoa).

Additionally, in Menorca, Sabadell and Valencia subcohorts, four mutations (R501X, 2282del4, R2447X and S3247X) in the *Filaggrin* gene (*FLG*) were genotyped by LGC Genomics using KASP™ genotyping technology.

**Acknowledgments and Funding**

The authors would particularly like to thank all the participants for their generous collaboration. A full roster of the INMA Project Investigators can be found at http://www.proyectoinma.org/presentacion-inma/listado-investigadores/en_listado-investigadores.html.

This study was funded by grants from Instituto de Salud Carlos III (CB06/02/0041, G03/176, FIS PI041436, PI081151, PI041705, PI061756, PI091958, and PS09/00432, FIS-FEDER 03/1615, 04/1509, 04/1112, 04/1931 , 05/1079, 05/1052, 06/0867, 06/1213, 07/0314, 09/02647, 09/00090, 11/01007, 11/02591, 11/02038, 13/1944, 13/2032, 13/02187 and CP11/0178), Spanish Ministry of Science and Innovation (SAF2008-00357), CIBERESP, European Commission (HEALTH.2010.2.4.5-1, FP7-ENV-2011 cod 282957), Fundació La Marató de TV3, Generalitat de Catalunya-CIRIT 1999SGR 00241, Conselleria de Sanitat Generalitat Valenciana, Department of Health of the Basque Government (2005111093, 2009111069, 2013111089 and 2015111065), the Provincial Government of Gipuzkoa (DFG06/002, DFG08/001 and DFG15/221) and annual agreements with the municipalities of the study area (Zumarraga, Urretxu , Legazpi, Azkoitia y Azpeitia y Beasain).

Part of the DNA extractions and genotyping was performed at the Spanish National Genotyping Centre (CEGEN-Barcelona). FLG genotyping in Menorca, Valencia and Sabadell was funded by an MRC centenary award (awarded to L Paternoster).

### *Isle of Wight Birth Cohort - IOW*

**Recruitment**

The IOW birth cohort study, a population-based sample of 1456 infants, initially enrolled study participants (new borns) between January 1989 and February 1990. Study participants have been prospectively assessed at ages 1, 2, 4, 10 and 18 years, as detailed elsewhere1,2.

**Definition of eczema cases and controls**

In all assessments of the Isle of Wight birth cohort, eczema was defined as chronic or chronically relapsing itchy dermatitis lasting more than 6 weeks with characteristic morphology and distribution, according to the criteria of Hanifin and Rajka.

**Definition of environmental exposures**

<u>Cesarean section</u>

Child's birth was recorded as vaginal or C-section

<u>Cat ownership</u>

Cat ownership was report at birth by parents

<u>Dog ownership</u>

Dog ownership was report at birth by parents

<u>Breastfeeding (duration / ever any / ever exclusive)</u>

Duration of any breastfeeding was reported by the mother in months

Ever breastfeeding was reported by the mother

<u>Presence of older siblings</u>

Presence of older siblings was reported by parents at birth

<u>Environmental tobacco smoke exposure (in utero / up to 2 years)</u>

Exposure to ETS in utero was reported by the mother at birth

Exposure to household ETS during the first 2 years of life was reported by parents

**Definition of parental education (as a measure of socioeconomic status)**

Mother have had formal education for at least 16 years

**Definition of family history of atopic disease**

Maternal or/and paternal history of asthma, eczema, or rhinitis

**Genotyping**

*FLG**

rs10214237*

rs1057258

rs112111458

rs11657987

rs12295535

rs17881320

rs2143950

rs2227483*

rs6473227

rs7127307

rs7146581*

*Were included for in the replication analysis

**Acknowledgments and Funding**

We thank the participants and their families who have helped us with this project over the last two decades. We would like to acknowledge the help of all the staff at The David Hide Asthma and Allergy Research Centre in undertaking the 18-year and previous assessments of the 1989 Isle of Wight birth cohort. The 18-year assessment and other analyses were funded by grants from the National Institute of Health, USA (R01 HL082925 and R01AI091905).

### *MAAS*

**Recruitment**

The Manchester Asthma and Allergy Study is an unselected (i.e. population-based), birth cohort study (Custovic et al., 2002, Lowe et al., 2005, Murray et al., 2002, Nicolaou et al., 2006, Nicolaou et al., 2008). The setting is the maternity catchment area of Wythenshawe and Stepping Hill Hospitals, comprising of 50 square miles of South Manchester and Cheshire, UK, a stable mixed urban-rural population. Study was approved by the Local Research Ethics Committee. Informed consent was obtained from all parents.

*Screening & Recruitment*

All pregnant women were screened for eligibility at antenatal visits (8th-10th week of pregnancy). The study was explained to the parents, and informed consent for initial questionnaires and skin prick testing was obtained. Both parents completed a questionnaire about their and their partner's history of asthma and allergic diseases and smoking habits. If the pregnant woman's partner was not present at the antenatal clinic visit, an invitation was sent for him to attend an open-access evening clinic for skin prick testing and questionnaire. Once both parents had completed questionnaires and skin prick testing, a full explanation of the proposed future follow-up for the child was given. Of the 1499 couples who met the inclusion criteria (<10 weeks of pregnancy, maternal age >18 years, questionnaire and skin test data available for both parents), 288 declined to take part in the study. A total of 1185 participants had at least some evaluable data.

*Follow-up*

The children have been followed prospectively, and attended review clinics at ages 1, 3, 5, 8 and 11 years.

**Definition of eczema cases and controls**

Cases were defined as a doctor diagnosis of atopic dermatitis on the day that the child came to the follow-up clinic, at any timepoint (1y, 3y, 5y, 8y)

Controls were a Parental report of "no" at all timepoints (1y, 3y, 5y, 8y) to the question "Has your child ever suffered from atopic dermatitis", 'No'

**Definition of environmental exposures**

<u>Antibiotics (up to 6 months / up to 12 months)</u>

<u>Cesarean section</u>

<u>Cat ownership</u>

<u>Dog ownership</u>

<u>Breastfeeding (duration / ever any / ever exclusive)</u>

<u>Presence of older siblings</u>

<u>Environmental tobacco smoke exposure (in utero / up to 2 years)</u>

<u>Air pollution (NO2, PM10)</u>

<u>House dust mite: Der p1 concentration (at birth / 1 year)</u>

**Definition of parental education (as a measure of socioeconomic status)**

Parental income at the child's recruitment.

**Definition of family history of atopic disease**

Mother and/or Father has atopy at birth (skin prick test positive)

**Genotyping**

DNA samples were genotyping on an Illumina 610 quad chip. The Illumina genotypes were called using the Illumina GenCall application following the manufacturer's instructions. Quality control criteria for samples included: 97% call rate, exclusion of samples with an outlier autosomal heterozygosity (scree-plot visualization) gender validation and sequenome genotype concordance. Quality control criteria for SNPs included a 95% call rate, HWE > $5.9x10^{-7}$, minor allele frequency > 0.005. Genotypes were prephased with SHAPEIT[20] shapeit.v2.837 and imputed with IMPUTE2[7] version 2.3.2 using the March2012 release of the 1 1000 Genomes Phase 3 reference genotypes.

FLG genotyping was carried out as previously described in Bisgaard et al.[57] and Sandilands et al.[23]. Briefly genotyping The R501X[57], R2447X and S3247X[23] mutations was performed using a TAQMAN-based allelic discrimination assay (Applied Biosystems). The 2282del4 Mutation was genotyped on an Applied Biosystems 3100 or 3730 DNA sequencer[57].

**Acknowledgments and Funding**

We would like to thank the children and their parents for their continued support and enthusiasm. We greatly appreciate the commitment they have given to the project. We would also like to acknowledge the hard work and dedication of the study team (post-doctoral scientists, research fellows, nurses, physiologists, technicians and clerical staff). This report includes independent research supported by National Institute for Health Research Respiratory Clinical Research Facility at Manchester University NHS Foundation Trust (Wythenshawe). The views expressed in this publication are those of the author(s) and not necessarily those of the NHS, the National Institute for Health Research or the Department of Health. MAAS was supported by the Asthma UK Grants No 301 (1995-1998), No 362 (1998-2001), No 01/012 (2001-2004), No 04/014 (2004-2007), the BMA James Trust and Medical Research Council, UK (G0601361) and The Moulton Charitable Foundation (2004-current); the Medical Research Council (MRC) Grants G0601361, MR/K002449/1 and MR/L012693/1, and Angela Simpson is supported by the NIHR Manchester Biomedical Research Centre. The authors would like to acknowledge the North West Lung Centre Charity for supporting this project.

### *Multicentre Allergy Study (MAS)*

#### Recruitment

The Multicentre Allergy Study (MAS) is a German birth cohort which has been described in detail previously.[1,2] Briefly, in 1990, the MAS cohort recruited 1,314 newborns, who were regularly followed-up. Data were collected from examinations and questionnaires at birth, at 1, 3, 6, 12, and 18 months, and yearly from age 2 to age 13. The institutional review boards of the participating centers approved the study, and written informed consent was obtained.

#### Definition of eczema cases and controls

Eczema was defined by the presence of either i) a reported physician's diagnosis, ii) parental report of eczema symptoms, or iii) visible eczema at the time of follow-up.[3]

#### Definition of environmental exposures

Environmental exposures were defined based on the answers to the following questions (from questionnaires at the indicated time points).

<u>Antibiotics (in utero / up to 6 months / up to 12 months)</u>

Have you used antibiotics since the last questionnaire? (in utero not available / at 1, 3, or 6 months / at 1, 3, 6 or 12 months)

<u>Cesarean section</u>

(at birth)

<u>Cat ownership</u>

Do you keep cats? (at 3 months)

<u>Dog ownership</u>

Do you keep dogs? (at 3 months)

<u>Breastfeeding (duration / ever any / ever exclusive)</u>

Duration of breastfeeding (at 12 months) / Are you breastfeeding? (at 1, 3, 6 or 12 months) / Have you introduced food other than breast milk? (at 1 month)

<u>Presence of older siblings</u>

Number of siblings (at birth)

<u>Environmental tobacco smoke exposure (in utero / up to 2 years)</u>

Have you (mother) been smoking during pregnancy? (at 1 month) / Is someone smoking in your household? (at 1 month or at 18 months)

<u>Washing practices (up to 6 months / up to 2 years)</u>

How often do you bathe your child; >=7 times/week or <7 times/week? (at 3 months / at 9 months)

<u>Air pollution (NO2, PM10)</u>

Where do you live; urban environment or village? (at 3 months)

<u>House dust mite: Der p1 concentration (at birth / 1 year)</u>

Collected at 9 months / 18 months (Dpt in µg/g)

#### Definition of parental education (as a measure of socioeconomic status)

Highest education of the father or mother (at birth)

**Definition of family history of atopic disease**

Asthma, eczema or allergic rhinitis of the father or mother (at birth)

**Genotyping**

DNA samples of 871 children were available for genotyping. Genomic DNA was prepared from whole blood using standard methods. In all individuals the FLG mutations were genotyped by using TaqMan allelic discrimination (p.R501X, p.R2447X and p.S3247X)[4] or fluorescence-based semiautomated allele-sizing technology (c.2282del4).[5]

**Acknowledgments and Funding**

We wish to thank the initiators Ulrich Wahn, Renate L. Bergmann, and Karl E. Bergmann. Furthermore we thank Carl-Peter Bauer, Johannes Forster, Volker Wahn, Antje Schuster, Fred Zepp, Ute Hoffmann, Christoph Grueber, Renate Krueger, Bodo Niggemann, Petra Wagner, Monika Goetze, Andreas Reich, and Gabriele Schulz. We also thank all MAS children and their parents, all collaborators and staff not listed here. The MAS study up to adolescence was funded by several grants from the German Federal Ministry of Education and Research (07015633, 07 ALE 27, 01EE9405/5, and 01EE9406).

#### *NTR twins*

**Recruitment**

The Netherlands Twin Register (NTR) collects data on development and growth in twins who are registered at birth by their parents. In the NTR, data on eczema and the environmental exposures were obtained from surveys mailed to parents when twins were aged 1, 2, 3, and 5 years. Shortly after registration of their newborn twins, mothers receive the first survey in which they are asked to report on mode of delivery, the use of medication during pregnancy and on smoking behavior of both parents during pregnancy. The survey when the twins are 2 years of age includes questions on whether the children have ever used medication, which kind of medication they have used, the duration of breastfeeding, and the current smoking status of both parents. Data on parental education attainment are obtained at age 3 of the twins. At age 5, the parents are asked to report (yes/no) whether a physician ever diagnosed eczema in their children. From the same survey, information on current parental smoking and the number of older sibs is obtained.

**Definition of eczema cases and controls**

-Eczema cases are defined based on a yes reply to the question whether a physician ever diagnosed eczema (dichotomous question at age 5 years of the child).

-Controls are defined as a no reply to the question.

**Definition of environmental exposures**

Antibiotics (in utero / up to 6 months / up to 12 months): Antibiotic use in utero was based on the question of prescribed medication or nutritional supplements during the pregnancy in the first survey of mothers.

Antibiotics up to 2 years was based on the question: Did your child ever use medication?

All medications with a ‘J01’ Anatomical Therapeutic Chemical (ATC) code were assigned as antibiotics use.

Cesarean section: Information on mode of delivery was collected from mother after registration of new born twins. Cesarean section included planned and urgent cesarean sections. All other modes of delivery were assigned as controls

Breastfeeding (duration / ever any / ever exclusive). At age 2 years of twins mothers reported the duration of breastfeeding for each child using the following categories: ‘no’, ‘less than two weeks’, ‘2-6 weeks’, ‘6 weeks – 3 months’, ‘3 – 6 months’ and ‘more than 6 months’.

Presence of older siblings: At age 5, the parents report the following information on the sibs of twin pairs: birth date, sex, relationship, and whether the sib lives together with the twin pairs. The number of older sibs is obtained by counting the older sibs that live together with the twins regardless their biological relationship.

Environmental tobacco smoke exposure (in utero / up to 2 years): Information on smoking exposure in utero was collected from mother after registration of new born twins and on current smoking behavior in the surveys sent at age 2 and 5 of the twins. In these surveys the following categories were used: ‘no’, ‘yes, pipes/cigars’, ‘yes, less than 10 cigarettes’ and ‘yes, more than 10 cigarettes’.

**Definition of parental education (as a measure of socioeconomic status**) Parental education was assessed in he surveys at ages 3, 7, and 10 on a 13-point scale ranging from primary to post-doctoral education. The most recent education measure was used and collapsed into 4 categories: ‘primary school’, ‘lower vocational school and lower secondary school’, ‘intermediate vocational school and intermediate or higher secondary school’, and ‘higher vocational school and university”.

**Genotyping**

Genotyping was done on multiple platforms, with a number of overlapping participants. The following platforms were chronologically used: Affymetrix-Perlegen, Illumina 660 , Illumina Omni Express 1 M and Affymetrix 6.0. Genotype calls were made with the platform specific software (Birdseed, APT-Genotyper, Beadstudio) following manufacturers' protocols. For the Affymetrix-Perlegen and Illumina 660 platforms, the SNPs were lifted over to build 37 (HG19) of the Human reference genome. Per platform, a sample was removed if the call rate for this person was <90%, the Plink 1.07 value F was <-0.075 or >0.075, the gender of the person did not match the DNA of the person, the IBD status did not match the expected familial relations, or the sample had more than mean+5sd Mendelian errors. For the Affymetrix 6.0 platform all samples with a CQC value < 0.40 were removed. Afterwards, in case a subject, was genotyped on multiple platforms, only the platform with the highest number of SNPs was selected when concordance between platforms was over 97%. Allele - and strand alignment of SNPs was done against the Dutch GONL reference panel for each platform [1]. SNPs were removed in each platform when MAF < 0.005, HWE< $10^{-12}$ and the call rate of the SNP was < 95%[2]. Then SNPs were selected if the allele frequency of the SNP deviated <0.10 as compared to the GONL data. Subsequently, the individual platform data were merged into a single dataset. In this single dataset, the sample IBD, on a common backbone of ~70K SNPs, was re-compared with their expected familial relations and samples were removed if they did not match. The single merged dataset was imputed with mach-admix, using GONL as a reference panel, for only the SNPs that survived QC and were present on at least one platform, forcing missing genotype imputation for all SNPs. Best guess genotypes were generated from these data and from these cross-platform imputed SNPs, the following SNPs were selected: SNPs with a R2>0.90, with HWE p > 0.00001, with a Mendelian error rate < 2% and if the association of one platform=case vs. the other platforms=controls p-value > 0.00001 (of course applied for each platform). This left 1.2M SNPs. These SNPs were then re-aligned against the HRC 1.1 reference panel and then imputed on the Michigan imputation server [3]. From the resulting VCF files, best guess genotypes were calculated and the SNPs required for these analyses were selected.

**References**

1: PMC3895638

2: PMC4236550

3: PMC5157836

**Acknowledgments and Funding**

We would like to thank all twin families for taking part in the study. DIB acknowledges the Royal Netherlands Academy of Science Professor Award (PAH/6635).

We thank the Amsterdam Public Health (APH) institute for awards "Elucidating factors influencing the genetic background estimation in family members" and “Personalized Medicine in Pediatric Asthma”; the Netherlands Organization for Scientific Research (NWO) and The Netherlands Organisation for Health Research and Development (ZonMW) : NWO-Groot 480-15-001/674: Netherlands Twin Registry Repository; the Biomolecular Resources Research Infrastructure (BBMRI–NL, 184.021.007; 184.033.111); the European Community's Seventh Framework Program Grant 602768: ACTION). Genotyping was funded by the National Institutes of Health (NIH, , Grand Opportunity grant 1RC2 MH089995) and the Avera Institute for Human Genetics, Sioux Falls, South Dakota (USA).

#### *PIAMA (Prevention and Incidence of Asthma and Mite Allergy)*

**Recruitment**

The PIAMA-study is a large ongoing population-based birth cohort study with prenatal inclusion and follow-up until the current age of 23/24 years. The study was designed to investigate the influence of lifestyle and environment on the development of asthma, allergy and lung function. It therefore contains a detailed characterization of lifestyle and environmental exposures in all stages of childhood. The study started with 3,963 newborns, who were born in 1996/97 after prenatal recruitment through prenatal clinics in the northern, middle and southwestern part of the Netherlands.

After inclusion, allergic and non-allergic mothers were identified through a validated screening questionnaire. Pregnant women identified as allergic were primarily allocated to an intervention study with a random subset allocated to a "natural history study". The intervention involved the use of mite-impermeable mattress and pillow covers on the children's and parents' beds. Non-allergic pregnant women were invited to participate in a "natural history" study arm.

- More information on the design and methods: **Brunekreef et al. 2002**
- More information on the intervention study: **Gehring et al. 2012**

**Definition of eczema cases and controls**

Questionnaire information on eczema was obtained at ages 3m,1y,2y,3y,4y,5y,6y,

7y,8y. Cases were defined as a positive response to one or more of these three questions:

1. Has your child ever had atopic dermatitis? 2. Did a doctor ever diagnose atopic dermatitis in your child? 3. Did your child have atopic dermatitis during the past 12 months?

Controls were defined a negative response to these questions at ages 2 – 8 years.

**Definition of environmental exposures**

Cesarean section

Born by Cesarean section (self-reported)

Cat ownership

Cat present in the house during pregnancy (self-reported)

Dog ownership

Dog present in the house during pregnancy (self-reported)

Breastfeeding (duration / ever any / ever exclusive)

Ever breastfeeding and duration of breastfeeding is self-reported. Duration of breastfeeding in months is calculated as ‘duration of breastfeeding in weeks/4.35.

Presence of older siblings

Self-reported

Environmental tobacco smoke exposure (in utero / up to 2 years)

ETS in utero is defined as having a smoking mother during pregnancy (irrespective of the trimester)(self-reported)

ETS up to 2 years is defined as smoking indoors by the father or the mother (self-reported at 6 months, 1 year, or 2 year)

Air pollution (NO2, PM10)

PM10 and NO2 exposure at the birth address. Estimated using Land Use Regression models as developed in the ESCAPE project (**Beelen et al. 2013**; **Eeftens et al. 2012a**; **Gehring et al., 2013**).

House dust mite: Der p1 concentration (at birth / 1 year)

House dust mite Der p1 is measured before birth, at 3 months and at 1 year in the mattress of the child.

**Definition of parental education (as a measure of socioeconomic status)**

Highest education of father or mother (coded as low/medium vs high)

**Definition of family history of atopic disease**

Family history of asthma (mother and father), hayfever (mother and father), and allergy (siblings)

**Genotyping**

DNA was collected from 2,162 children. Genome-wide genotyping was performed in three phases. The first phase was performed within the framework of the GABRIEL Consortium using an Illumina Human 610K quad array37. Genotypes were available from 172 children with asthma and from 187 controls after quality control. A second group of 268 children who were more extensively examined during follow up was genotyped with an Illumina HumanOmniExpress array. A final group of 1377 children was genotyped with the Illumina Human Omni Express Exome Array. SNPs were harmonized by base pair position annotated to genome build 37, name and annotation of strand for each platform. Discordant or duplicate SNPs or SNPs that showed large differences in allele frequencies (> 15 %) were removed. After quality control, a total of 1968 individuals remained and imputation was performed per platform using IMPUTE 2,0 against the reference data set of the CEU panel of the 1000 Genomes project (version March 2012). SNPs of high quality (info-score IMPUTE ≥ 0.7) were merged into one dataset using GTOOL and used for further analysis.

**Acknowledgments and Funding**

The PIAMA study is supported by The Netherlands Organization for Health Research and Development; The Netherlands Organization for Scientific Research; The Lung Foundation of the Netherlands (grant number AF 45.1.14.001 supported the nasal methylation assays); The Netherlands Ministry of Spatial Planning, Housing, and the Environment; and The Netherlands Ministry of Health, Welfare, and Sport.

We thank all the children and their parents for their cooperation. We also thank all the field workers and laboratory personnel involved for their efforts, and Marjan Tewis and Marjan Kerkhof for data management.

### *The Raine Study*

**Recruitment**

The Raine Study is a prospective pregnancy cohort where 2900 mothers were recruited between 1989-1992 (https://rainestudy.org.au/). Recruitment took place at Western Australia's major perinatal centre, King Edward Memorial Hospital, and nearby private practices. Women who had sufficient English language skills, an expectation to deliver at King Edward Memorial Hospital, and an intention to reside in Western Australia to allow for future follow-up of their child were eligible for the study.[1]

**Definition of eczema cases and controls**

A study individual is defined as a case if he/she was diagnosed with eczema by a doctor at any follow-up by 16 years of age (conducted at 5, 8, 10, 13/14, 16/17).

A study individual is defined as a control if he/she was never ever diagnosed with eczema at any follow-up by 16 years of age.

**Definition of environmental exposures**

<u>Antibiotics (in utero / up to 6 months / up to 12 months)</u>

Self-reported usage of antibiotics from the Raine Study antenatal data at 18- and 34-weeks gestation, and the Raine Study Gen2 1- and 2-year follow-ups.

<u>Cesarean section</u>

Retrieved from medical records (Mode of delivery)

<u>Cat ownership</u>

Self-reported number of cats owned

<u>Dog ownership</u>

Self-reported number of dogs owned

<u>Breastfeeding (duration / ever any / ever exclusive)</u>

Duration - Self-reported time at which Raine Study Gen1 mothers stopped breastfeeding Raine Study Gen2 index participants. This question was included in questionnaires at the Raine Study Gen2 1-, 2- and 3-year follow-ups.

<u>Environmental tobacco smoke exposure (in utero / up to 2 years)</u>

Self-reported smoking from the Raine Study antenatal data at 18- and 34-weeks gestation, and the Raine Study Gen2 1- and 2-year follow-ups.

**Definition of parental education (as a measure of socioeconomic status)**

Completed year 12 (yes/no) - equivalent to A levels

**Definition of family history of atopic disease**

Self-reported

**Genotyping**

Illumina 660W

Illumina Omni

**Acknowledgments and Funding**

The Raine Study acknowledges the Raine Study participants and their families for their ongoing participation in the study and the Raine Study team for study co-ordination and data collection. We also thank the National Health and Medical Research Council (NHMRC) and the Raine Medical Research Foundation for their long-term contribution to funding the study over the last 30 years. The core management of the Raine Study is funded by The University of Western Australia (UWA), Curtin University, The Kids Research Institute Australia, Women and Infants Research Foundation, Edith Cowan University, Murdoch University, and The University of Notre Dame Australia. This study was supported by the National Health and Medical Research Council of Australia [grant numbers 572613, 403981, 003209, 211912, and 353514] and the Canadian Institutes of Health Research [grant number MOP-82893]. The authors gratefully acknowledge the assistance of the Western Australian DNA Bank (National Health and Medical Research Council of Australia National Enabling Facility). All analytic work was supported by resources provided by the Pawsey Supercomputing Centre with funding from the Australian Government and the Government of Western Australia.

#### *TwinsUK*

**Recruitment**

The TwinsUK adult twin registry includes about 14,000 subjects, predominately middle-aged females, unselected for any specific disease and with similar disease and lifestyle characteristic to the general population [PMID:11780939]. St. Thomas' Hospital Research Ethics Committee approved the study, and all twins provided informed written consent.

**Definition of eczema cases and controls**

Self-reported eczema data were collated from multiple questionnaires completed by TwinsUK participants. All questions asked: *"Have you ever had eczema?"* or "*Has a doctor ever told you that you have/had any of the following conditions? (Eczema)"*. Participants were considered as affected by eczema if they replied yes to any questionnaire, unaffected if they only replied no. Participants who did not provide any answer were discarded. Only participants of European ancestry were considered in this analysis.

**Definition of environmental exposures**

<u>Cesarean section</u>

Self-reported method of delivery (naturally/Cesarean section) was collated from multiple questionnaires. All questions asked: *"How were you delivered?"*. Participants who provided discordant answers were discarded.

<u>Presence of older siblings</u>

Presence of older siblings was derived from self-reported family information ("*Year of birth of all siblings"*).

**Definition of family history of atopic disease**

Self-reported family history of eczema was collected via questionnaire. Participants were asked, via multiple questions, whether any parent, sibling, or child has or had suffered from eczema.

**Genotyping**

Microarray genotyping was conducted using a combination of Illumina arrays (HumanHap300, HumanHap610, 1M-Duo and 1.2M-Duo 1 M) and imputation was performed using the IMPUTE2 software using haplotype information from the 1000 Genomes Project (Phase 1, integrated variant set across 1092 individuals, v2, March 2012), as previously described [PMID:21572415].

**Acknowledgments and Funding**

TwinsUK is funded by the Wellcome Trust, Medical Research Council, European Union, the National Institute for Health Research (NIHR)-funded BioResource, Clinical Research Facility and Biomedical Research Centre based at Guy's and St Thomas' NHS Foundation Trust in partnership with King's College London.

#### *BABY Skin birth cohort*

**Recruitment**

300 term children were recruited from the maternity ward at Copenhagen University Hospital – Rigshospitalet, Denmark, during the first 3 days of life.

**Definition of eczema cases and controls**

If a child had any signs of eczema during the first 2 years of life, they attended a clinical visit where atopic dermatitis was diagnosed by a physician.

**Definition of environmental exposures**

Cat ownership: Defined as answering "yes, a cat" to the question: "Have you been living with a pet while you were pregnant?". [Parental questionnaire when child was 2 months of age].

Breastfeeding (duration / ever any / ever exclusive): Ever breastfeeding defined as answering "breastfeeding" to the question: "Primary method of feeding?" or an affirmative response to the question: "If not breastfeeding now, have you been breastfeeding when your child was younger?". Duration of breastfeeding was defined as number of months the child was breastfed. [Parental telephone interview when child was 18 and 24 months of age].

Presence of older siblings: Defined as answering: "1 child/2 children/3 children/more than 3 children" to the question: "How many biological children do you already have?" [Parental questionnaire when child was 2 months of age].

Environmental tobacco smoke exposure (in utero / up to 2 years): Environmental tobacco exposure up to 2 years defined as an affirmative reply to the question: "Do you or other person smoke inside your home?". [Parental telephone interview when child was 18 and 24 months of age].

Washing practices (up to 6 months / up to 2 years): Bathing every day defined as answering "every day" to the question: "How many times a week do you bathe you child?". [Interview at clinical visits when child was 2, 6, and 12 months of age].

**Definition of family history of atopic disease**

Family history of atopic disease was defined as self-reported physician-diagnosed atopic dermatitis, asthma, hay fever and/or self-reported allergy to selected allergens (birch, grass, mugwort, horse, dog, cat, house dust mites or molds).

**Genotyping**

DNA was collected via a buccal swab (Isohelix, Herrietsham, UK) and analyzed for the three common filaggrin gene mutations ((R501X, 2282del4 and R2447X) using TaqMan genotyping assay.

**Acknowledgments and Funding**

The BABY Skin birth cohort received financial support from The Leo Foundation, The Lundbeck Foundation, The Novo Nordisk Foundation, Aage Bangs fond, 'Savværksejer Jeppe Juhl og hustru Ovita Juhls Mindelegat' and Herlev and Gentofte Hospital Research Foundation. The study was conducted independent of these, and they had no influence on study design or data analysis.

#### *BAMSE*

**Recruitment**

BAMSE (Swedish abbreviation for Children, Allergy, Milieu, Stockholm, Epidemiology) is a Swedish longitudinal birth cohort study. A total of 4,089 newborn infants were recruited in the BAMSE study between 1994 and 1996 in the north an central area of Stockholm(1). Questionnaire data on baseline study characteristics were obtained when the children were about 2 months. At approximately one, two, four, eight, twelve and sixteen years old, parents completed questionnaires on their children's symptoms related to asthma and other allergic diseases.

**Definition of eczema cases and controls**

The children have been followed with regular questionnaires and clinic visits. For the current study, data collected from the questionnaires was used to classify children as eczema cases or controls. Eczema case status at 8 years old was defined based on an affirmative response at any of the following questions in questionnaires up to 8 years: "Has a doctor ever diagnosed your child as having eczema?" [yes/no], "Has a doctor diagnosed your child as having eczema after the age of one year?" [yes/no], "Has a doctor diagnosed your child as having eczema after the age of two years?" [yes/no] or "Has your child been diagnosed with eczema by a doctor since age 4?" [yes/no]. Controls were defined as children who were not cases and whose answer was negative for the questions above.

**Definition of environmental exposures**

<u>Cat ownership</u>

Presence of cat at home at the time of questionnaire 0.

<u>Breastfeeding (duration / ever any / ever exclusive)</u>

Total breastfeeding duration in number of months.

<u>Presence of older siblings</u>

Presence of elder sibling(s) at the time of questionnaire 0 was defined as a dichotomous variable [yes=1; no=0].

<u>Environmental tobacco smoke exposure (in utero / up to 2 years)</u>

Binary environmental tobacco exposure was defined based on an affirmative or negative response to any of the following questions, independent of how many cigarettes for the following questions: "Did the father smoke when the mother was pregnant with the "BAMSE child"?" [1 = No; 2 = Yes, rarely; 3 = Yes, daily but not in the home; 4 = Yes, daily in the home] or "Did the mother smoke while pregnant with the "BAMSE child"?· [1 = No; 2 = Yes, rarely; 3 = Yes, daily but not in the home; 4 = Yes, daily in the home].

**Definition of parental education (as a measure of socioeconomic status)**

Highest education level of the household at the time of questionnaire 0, based on maternal and paternal education level, in two categories: a) Up to high school (gymnasium) or b) University (högskola/universitet).

**Definition of family history of atopic disease**

Mother and/or father with doctor's diagnosis of asthma and asthma medication and/or doctor's diagnosis of hay fever in combination with furred pets- and/or pollen allergy at the time of questionnaire 0.

**Genotyping**

DNA was extracted from peripheral blood and genotyping was described elsewhere(2). Recently, a total of 2,378 16-year-old children were additionally genotyped with the Ilumina Infinium Global Screening Array-24 BeadChip following the same sample collection procedures, protocols and questionnaires. Quality control (QC) was performed following the Ricopili pipeline(3). Data was imputed using the Haplotype Reference Consortium(4) 1.1 reference panel. Variants with imputation quality (Rsq) >= 0.3 were retained. After QC, the imputed or genotyped variants of interest were extracted for analysis.

**Acknowledgments and Funding**

This study was supported by grants from the Swedish Research Council, the Swedish Research Council for Health, Working Life and Welfare, Formas, the Swedish Heart-Lung Foundation, the European Research Council (TRIBAL, grant agreement 757919), and Region Stockholm (ALF project, and for cohort and database maintenance).  We thank the children and parents participating in the BAMSE cohort and all staff involved in the study through the years.

***BASELINE***

Cork BASELINE Birth Cohort Study

**Recruitment**

Infants recruited to the Cork Babies After Scope: Evaluating the Longitudinal Impact Using Neurological and Nutritional Endpoints (BASELINE) Birth Cohort study. The purpose of the BASELINE study was to examine the effects of environmental factors during pregnancy and infancy on childhood health and development. It was developed as the pediatric follow on from the Cork Centre for the Screening for Pregnancy Endpoints (SCOPE) study, a multicenter international study evaluating diseases of pregnancy in primigravidous women. The Cork BASELINE Birth Cohort study recruited from August 2008 through to October 2011. A total of 1537 infants were recruited antenatally (stream1). These women were subject to the inclusion criteria of the SCOPE study, namely, first-time, low-risk mothers with singleton pregnancies delivered at or near term. A second recruitment stream began in July 2010 that recruited mothers and babies on the postnatal ward (stream2). A total of 600 infants were recruited postnatally. These mothers were enrolled independently of the SCOPE study.

Cohort profile paper - https://academic.oup.com/ije/article/44/3/764/629555

**Definition of eczema cases and controls**

Questions below asked at 6, 12, 24 months and 5 years appointments, UK eczema diagnostic criteria (Carried out by a trained research assistant or nurse (UK diagnostic criteria))

- Has your child had an itchy skin condition in the last 6 months?
- If yes, is there a history of flexural involvement?
- Does your child have a history of generally dry skin?
- Is there a history of atopic disease in a first degree relative?
- On assessment, is there visible flexural dermatitis as per photographic protocol?
- Does the child meet the UK diagnostics criteria for eczema?

**Definition of environmental exposures**

Antibiotics in utero

Q: Antibiotics taken during pregnancy

1 - No
2 - Yes, occasionally
3 - Yes, for a specific episode
4 - Yes, regularly, at least several times a week

Cesarean section

Q: Birth mode of delivery –

1 - Unassisted vaginal
2 - Operative vaginal
3 - Prelabour LSCS
4 - LSCS in labour
5 - Unknown
6 - Maternal death antepartum

Cat ownership

Q: Do you have any animals at home during pregnancy?

The no. of animals in the home during pregnancy?

Is the animal allowed in the baby's room?

Is the animal allowed in the living room?

Is the animal allowed in the kitchen?

Is the animal only outside?

Dog ownership

Q: Do you have any animals at home during pregnancy?

The no. of animals in the home during pregnancy?

Is the animal allowed in the baby's room?

Is the animal allowed in the living room?

Is the animal allowed in the kitchen?

Is the animal only outside?

Breastfeeding (duration / ever any / ever exclusive)

Q: Type of infant feeding at present (2, 6 month visit)

1 - Breastfed only
2 - Bottle fed infant formula only
3 - Both breast and body fed
4 - Not asked

Q: Current type of infant feeding (12 month visit)?

1 - Breastfed
2 – Bottle fed infant formula
3 - Both breast and formula fed
4 - Other
5 - Not asked

Presence of older siblings

Q: Relationship to child 1, 2, 3, 4, 5 living in the home (5 year visit)?

1-Full sibling
2-Half sibling
3-Step sibling
4-Foster sibling
5-Uncle/aunt
6-Not related
7-Cousin

Q: Age years and months of child 1, 2, 3, 4, 5 living at home (5 year visit)?

Environmental tobacco smoke exposure (in utero / up to 2 years)

Q: Did you stop smoking or reduce the number of cigarettes when you found out you were pregnant?

1 - Yes, stopped completely
2 - Yes, reduced the number of cigarettes
3 - No, continued to smoke at the same level
4 - I had stopped smoking before I became pregnant

Q: Were you regularly exposed to second-hand/passive cigarette smoke while you were pregnant?

0 - No
1 - Yes

Q: Do you smoke currently (2 month visit)?

1 - Yes
2 – No, ex-smoker
3 - No, never smoked
4 – Not asked

Q: Do you smoke (mother) (6 month visit)?

1 - No
2 – Yes daily
3 – Yes, occasionally

Q: Does anyone else smoke inside your home (6 month visit)?

1 - No
2 – Yes daily
3 – Yes, occasionally

Q: Is your child exposed to tobacco smoke (6 month visit)?

1 - No
2 – Yes daily
3 – Yes, occasionally

<u>Washing practices (up to 6 months / up to 2 years)</u>

Q: How often do you bath your baby? (2 month visit)

1 - Once a week
2 - 2-3 times per week
3 - 3-5 times per week
4 - Daily
5 - Other
6 - Not asked

Q: What do you use in the bath? (2 month visit)

1 - Nothing
2 - Soap
3 - Bath foam
4 - Perfumed bath foam
5 - Emollient
6 - Milton
7 - Essential oils
8 - Other
9 - Not asked

Q: How often do you bath your baby? (6 month visit)

1 - Once a week
2 - 2 to 3 times per week
3 - 3 to 5 times per week
4 - Daily
5 - Other
6 - Not asked

Q: What do you use in the bath? (6 month visit)

1 - Nothing
2 - Soap
3 - Bath foam
4 - Perfumed bath foam
5 - Emoillient
6 - Milton
7 - Essential oils
8 - Other
9 - Not asked

**Definition of parental education (as a measure of socioeconomic status)**

Q: Total years of schooling (primary and secondary)

Q: Maternal tertiary Education University

1 - No
2 - Graduated
3 - Still attending
4 - Dropped out

Q: Maternal tertiary Education Other

1 - No
2 - Graduated
3 - Still attending
4 - Dropped out

**Definition of family history of atopic disease**

Questions asked to both mother and father at 2 month appointment

Q: DO or DID you ever suffer from pollen-related rhinitis ('hay fever')?

Q: DO or DID you ever suffer from eczema (atopic dermatitis)?

Q: DO or DID you ever suffer from asthma?

Questions asked about siblings at 5 year appointment

Q: How many natural siblings have been diagnosed with the following conditions

- Sibling diagnosed with asthma
- Sibling diagnosed with allergic rhinitis/hay fever
- Sibling diagnosed with eczema/atopic dermatitis

**Genotyping**

Filaggrin genotyping for four null mutations

- 2282del4
- R2447X
- R501X
- S3247X

**Acknowledgments and Funding:**

The Cork BASELINE Birth Cohort Study was funded by the National Children's Research Centre.

### *Born in Bradford (BiB)*

**Recruitment**

Baseline recruitment occurred between 2007-2011[1]. Women and their partners were recruited between 2007-2011 when women attended a routine antenatal clinic appointment at the City's main maternity unit. Around 83% of pregnant women attended this appointment, and of these >80% were recruited. The cohort recruited 12 453 women with 13 776 pregnancies (recruited at ~26 weeks gestation) and 3448 partners.

[1] Wright J, Small N, Raynor P, et al. Cohort Profile: The Born in Bradford multi-ethnic family cohort study. Int J Epidemiol . 2013 Aug 1;42(4):978–991.

**Definition of eczema cases and controls**

As per Abuabara et al 2017:

Katrina Abuabara, Alexa M. Magyari, Ole Hoffstad, Zarif K. Jabbar-Lopez, Liam Smeeth, Hywel C. Williams, Joel M. Gelfand, David J. Margolis, Sinead M. Langan. Development and Validation of an Algorithm to Accurately Identify Atopic Eczema Patients in Primary Care Electronic Health Records from the UK. Journal of Investigative Dermatology Volume 137 Issue 8. 2017 pp1655-1662.

https://doi.org/10.1016/j.jid.2017.03.029

**Definition of environmental exposures**

Antibiotics (in utero / up to 6 months / up to 12 months)

Extracted from primary care records.

Cesarean section

Obtained from maternity records

Breastfeeding (duration / ever any):

Data was obtained from BiB1000 (questionnaires administered when child was approximately 6, 12, 18, 24 and 36 months) and ALLIN surveys (12 and 24 months). Where more than one questionnaire was completed, information on duration was used from the questionnaire completed nearest in time to the end of breastfeeding to reduce the likelihood of recall bias.

Presence of older siblings:

Information obtained from BiB1000 and ALLIN surveys. Where survey data was missing, parity was used as an indicator.

Environmental tobacco smoke exposure (in utero / up to 2 years):

The following information on in-utero tobacco exposure obtained from the baseline questionnaire:

(1) the number of cigarettes the mother smoked in the first 3 months of pregnancy and since the beginning of the 4$^{th}$ month (none; 1-5 a day; 6-10 a day; 11-20 a day; over 20 a day).

(2) During pregnancy have you been exposed to other peoples' cigarette smoke at work or at home and if yes, for how many hours per day approx. (yes (number of hours); no; less than 1 hour per day/occasionally)

Information on tobacco smoke exposure to 2 years of age was obtained from BiB1000 (6 and 24 months) and ALLIN (12 and 24 months) surveys:

Is your child exposed to other peoples' smoke (yes (number of hours); no; less than 1 hour per day/occasionally)

Air pollution (NO2, PM10)

House dust mite: Der p1 concentration (at birth / 1 year)

**Definition of parental education (as a measure of socioeconomic status)**

A-level or higher (mother or father), obtained from baseline questionnaire

**Definition of family history of atopic disease**

Data obtained from ALLIN (12 month) and MeDALL questionnaires:

- Have you (the baby's mother) ever had any of the following diseases (asthma; hay fever; eczema)
- Has your baby's father ever had any of the following diseases (asthma; hay fever; eczema)
- Have any of your baby's brothers and sisters ever had any of the following diseases (asthma; hay fever; eczema)

**Genotyping**

BiB children (n=7018) were genotyped on two Illumina chips: CoreExome (n=5620) and GSA (n=1398). Classification of individuals as white Europeans (n=3019) and South Asians (n=3999) was based on a combination of PCA and parental reported ethnicity. Imputation using the HRC r1.1 reference panel was performed separately by chip type and ethnicity: CoreExome white European n=2405; CoreExome South Asian n=3215; GSA white European n=614; GSA South Asian n=784.

**Acknowledgments and Funding**

This work was supported by a joint grant from the UK Medical Research Council (MRC) and UK Economic and Social Science Research Council (ESRC) [MR/N024391/1]; the British Heart Foundation [CS/16/4/32482]; a Wellcome Infrastructure Grant [WT101597MA]; The National Institute for Health Research under its Applied Research Collaboration for Yorkshire and Humber [NIHR200166]. The NIHR Clinical Research Network which provided research delivery support for this study. The views expressed in this publication are those of the authors and not necessarily those of the National Institute for Health Research or the Department of Health and Social Care. Born in Bradford is only possible because of the enthusiasm and commitment of the children and parents in BiB. We are grateful to all the participants, health professionals, schools and researchers who have made Born in Bradford happen.

#### *Generation Scotland*

**Recruitment**

Generation Scotland (GS), is a large family-structured cohort study that consists of 24,000 individuals from across Scotland. Participants were identified via Community Health Index numbers, with the support of Scottish Practices and Professionals Involved in Research. The initial phase of recruitment (2006 to 2010) focussed on the Glasgow and Tayside regions of Scotland and was later extended to Ayrshire, Arran, and the Northeast of Scotland. Initial inclusion criteria were age 35-65, with ≥1 first-degree relative and ≥1 full sibling. The age range was later broadened to 18 to 65 years. Family members of probands were also invited to partake in the study. In total, 23,960 individuals were recruited, which encompassed 6,665 probands, 16,007 family members, and 1,288 individuals who self-volunteered without invitation. There were 5,573 families with a mean size of 4 members and 1,400 participants without relatives. The median age at baseline was 47 years and the sample was 59% female [12,13]. Detailed health and lifestyle information were collected via questionnaires at the study baseline alongside venepuncture to obtain whole blood samples from which DNA methylation and genotypes were assayed.

**Definition of eczema cases and controls**

Eczema case status was ascertained from a combination of at least one diagnostic Read code and at least two records for skin disease therapies recorded on separate days (as published in Abuabara et al. J Invest Dermatol 2017).

**Definition of environmental exposures**

**Antibiotics in utero**

Maternal linked prescription records were queried for BNF chapters 5.1, 11.3.1 and 13.10.1 during gestation (derived from 280 days prior to offspring date of birth).

**Presence of older siblings**

Self-reported number of siblings was available in the study questionnaire. Sibling sets were ascertained based on the study pedigree file, considering full sets only. Older sibling status was ascertained using participant date of birth.

**Environmental tobacco smoke exposure (in utero / up to 2 years)**

In-utero exposure to tobacco smoke was ascertained where mother-offspring pairs were present in the data. For ever-smoker mothers, age at which they started smoking and age at which they stopped was used to derive a window within which exposure could be derived based on date of birth of the offspring.

**Definition of parental education (as a measure of socioeconomic status)**

Parental education was available for parent-offspring sets in the study. A ‘higher education’ binary variable was derived from participants’ highest qualification. Higher education was defined as college/university degree, other professional or technical qualification, NVQ, HND, HNC or equivalent,  higher grade, and A-levels, AS levels or equivalent. Non-higher education was defined as standard grade, O levels, GCSEs or equivalent, CSEs or equivalent, school leavers certificate, and no qualification.

**Definition of family history of atopic disease**

Family history of atopic disease was derived from linked healthcare data of parents and siblings, considering eczema, asthma, and hayfever.

**Genotyping**

Samples were genotyped using the Illumina HumanOmniExpressExome-8v1 chip and the Beadstudio-Gencall v3 genotype calling algorithm. Quality control measures were implemented, filtering out samples with a call rate of <98% and SNPs with a call rate of <98%, HWE of $<1x10^{-6}$ and MAF of ≤1%, leaving 20,032 samples and 602,450 SNPs. Phasing of the genotyped SNPs was carried out using SHAPEIT2.

**Acknowledgments and Funding**

All components of GS received ethical approval from the NHS Tayside Committee on Medical Research Ethics (REC Reference Number: 05/S1401/89). GS has also been granted Research Tissue Bank status by the East of Scotland Research Ethics Service (REC Reference Number: 20-ES-0021), providing generic ethical approval for a wide range of uses within medical research. GS received core support from the Chief Scientist Office of the Scottish Government Health Directorates (CZD/16/6) and the Scottish Funding Council (HR03006). Genotyping and DNA methylation profiling of the GS samples was carried out by the Genetics Core Laboratory at the Edinburgh Clinical Research Facility, Edinburgh, Scotland, and was funded by the Medical Research Council UK and the Wellcome Trust (Wellcome Trust Strategic Award STratifying Resilience and Depression Longitudinally (STRADL; Reference 104036/Z/14/Z). We are grateful to all the families who took part, the general practitioners and the Scottish School of Primary Care for their help in recruiting them, and the whole Generation Scotland team, which includes interviewers, computer and laboratory technicians, clerical workers, research scientists, volunteers, managers, receptionists, healthcare assistants and nurses.

#### *Human Early Life Exposome (HELIX)*

**Recruitment**

HELIX project is an ongoing population-based birth control study in six birth cohorts from different European countries: (1) EDEN - Étude des Déterminants pré et postnatals du développement et de la santé de l'enfant, France; (2) Rhea - the Rhea Mother-Child Study in Crete, Greece; (3) KANC - Kaunus Cohort, Lithuania; (4) MoBa – the Norwegian Mother, Father and Child Cohort Study, Norway; (5) INMA - - INfancia y Medio Ambiente, Spain; (6) BiB - Born in Bradford, United Kingdom (UK) (Maitre et al., 2018). The HELIX project aims to implement novel exposure assessment and biomarker methods to measure the early-life exposure to multiple environmental factors and associate these with omics biomarkers and child health outcomes, thus characterizing the "early-life exposome". The entire study population is 31,472 mother-child pairs. The subcohort study includes 1,304 children with exposure, phenotypes, and molecular data measured at age of 6-12 years. The current study selected 748 children of European ancestry with phenotypic and genome-wide genetic from EDEN, MoBA, KAUNAS and RHEA subcohorts. All studies were approved by the national research ethics committees and informed consent to participate was obtained for all participants.

**Definition of eczema cases and controls**

Information on eczema was reported by the mothers at the HELIX visit, around 8 years of age, and included doctor diagnosis of eczema or atopic dermatitis or neurodermatitis.

**Definition of environmental exposures**

<u>Cat ownership</u>

Cat ownership was reported by the mothers and defined as "yes/no" having a cat mainly living in their house in postnatal life.

<u>Dog ownership</u>

Dog ownership was reported by the mothers and defined as "yes/no" having a dog mainly living in their house in postnatal life.

<u>Breastfeeding (duration / ever any / ever exclusive)</u>

Any breastfeeding duration was reported by the mothers using different questionnaires in the different cohorts, and expressed in months.

<u>Presence of older siblings</u>

Number of siblings was reported by the mothers and coded as "0", if none, or "1", with 1 or more than 1 sibling were present.

<u>Environmental tobacco smoke exposure (in utero / up to 2 years)</u>

Exposure to tobacco smoke during pregnancy was self-reported by the mothers and classified in "yes" (any exposure to tobacco smoke, either active or passive smoking), and "no" (no exposure, neither active or passive).

Exposure to tobacco smoke during childhood was also reported by the mothers and classified in "yes" (any exposure to second-hand smoke at home or other places) and "no" (no exposure to second-hand smoke).

**Definition of parental education (as a measure of socioeconomic status)**

Maternal education was used a proxy of socioeconomic status and coded as "0" (primary or secondary) and "1" (university).

**Definition of family history of atopic disease**

Family history of atopic disease was obtained from questionnaire and coded as "0", when none of both parents had atopic disease, and "1", when at least one of the parents had a history of atopic disease.

**Genotyping**

DNA was obtained from buffy coats collected in EDTA tubes at the mean age of 8 years. DNA was extracted by cohort using a Chemagen kit in batches of 12 samples. DNA concentration and quality was evaluated with the NanoDrop 1000 UV-Vis Spectrophotometer (ThermoScientific) and the Quant-iT™ PicoGreen® dsDNA Assay Kit (Life Technologies).

Infinium Global Screening Array (GSA) MD version 1 (Illumina) was used for genome-wide genotyping at the Human Genomics Facility (HuGe-F), Erasmus MC (www.glimdna.org). Genotype calling and annotation were done using the GenTrain2.8 algorithm based on a custom cluster file implemented in the GenomeStudio software and the GSAMD-24v1-0_20011747-A4 manifest. SNP coordinates were reported on human reference GRCh37 and on the source strand.

PLINK program was used for the quality control of the genetic data (Purcell et al., 2007). Briefly, samples were filtered out if they had a call rate <97%, had sex inconsistencies, the heterozygosity was >3 standard deviations, if they were related (sharing >18.5% of alleles) or duplicated. For ancestry prediction from GWAS data, we used the Peddy program (Pedersen and Quinlan, 2017). Then genetic ancestry was contrasted with self-reported ethnicity and discordant samples were excluded. Genetic variants were filtered out if they had a call rate <95%, if they were in the non-canonical pseudo-autosomal region (PAR), if they had a minor allele frequency (MAF) <1%, and if they were not in Hardy-Weinberg equilibrium (HWE) at a p-value <1E -06.

Genome-wide imputation was performed with the Imputation Michigan server using the Haplotype Reference Consortium (HRC) panel, Version r1.1 2016. Before the imputation PLINK data was converted into VCF format and the variants were aligned with the reference genome. Eagle v2.4 was used for the phasing of the haplotypes and minimac4 for the imputation. In the end, we retrieved 40,405,505 variants after imputation. Then we filtered out the genetic variants according to the following parameters: imputation accuracy (R2) <0.9, MAF<5%, and HWE p-value <1E-06. After this the post-imputation dataset had 4,614,947 variants.

**Acknowledgments and Funding**

The study has received funding from the European Community's Seventh Framework Programme (FP7/2007-206) under grant agreement no 308333 (HELIX project) and the H2020-EU.3.1.2. - Preventing Disease Programme under grant agreement no 874583 (ATHLETE project). The genotyping was supported by the project PI17/01225 and PI17/01935, funded by the Instituto de Salud Carlos III and co-funded by European Union (ERDF, "A way to make Europe") and the Centro Nacional de Genotipado-CEGEN (PRB2-ISCIII).

BiB received core infrastructure funding from the Wellcome Trust (WT101597MA) and a joint grant from the UK Medical Research Council (MRC) and Economic and Social Science Research Council (ESRC) (MR/N024397/1), and National Institute for Health Research Applied Research Collaboration Yorkshire and Humber (NIHR200166). The views expressed are those of the author(s), and not necessarily those of the NHS, the NIHR or the Department of Health and Social Care. INMA data collections were supported by grants from the Instituto de Salud Carlos III, CIBERESP, and the Generalitat de Catalunya-CIRIT. KANC was funded by the grant of the Lithuanian Agency for Science Innovation and Technology (6-04-2014_31V-66). The Norwegian Mother, Father and Child Cohort Study is supported by the Norwegian Ministry of Health and Care Services and the Ministry of Education and Research. The Rhea project was financially supported by European projects (EU FP6-2003-Food-3-NewGeneris, EU FP6. STREP Hiwate, EU FP7 ENV.2007.1.2.2.2. Project No 211250

Escape, EU FP7-2008-ENV-1.2.1.4 Envirogenomarkers, EU FP7-HEALTH-2009- single stage CHICOS, EU FP7 ENV.2008.1.2.1.6. Proposal No 226285 ENRIECO, EU- FP7- HEALTH-2012 Proposal No 308333 HELIX), and the Greek Ministry of Health (Program of Prevention of obesity and neurodevelopmental disorders in preschool children, in Heraklion district, Crete, Greece: 2011-2014; "Rhea Plus": Primary Prevention Program of Environmental Risk Factors for Reproductive Health, and Child Health: 2012-15). ISGlobal acknowledges support from the Spanish Ministry of Science and Innovation through the "Centro de Excelencia Severo Ochoa 2019-2023" Program (CEX2018-000806-S), and support from the Generalitat de Catalunya through the CERCA Program. CRG acknowledge the support of the Spanish Ministry of Science, Innovation, and Universities to the EMBL partnership, the Centro de Excelencia Severo Ochoa, and the CERCA Programme/Generalitat de Catalunya.

### *The HUNT Study*

#### Recruitment

The Trøndelag Health Study (HUNT) is a longitudinal population-based health study carried out at four time points over approximately 40 years (HUNT1 [1984-1986], HUNT2 [1995-1997] and HUNT3 [2006-2008] and HUNT4 [2017-2019]) (1, 2). All inhabitants aged 20 years and over residing in Trøndelag County in Norway were invited to participate. The surveys included clinical measurements, blood sampling and questionnaires on general health measures and a broad range of self-reported diseases and symptoms. Participants were linked to regional- and national health registries through their unique national identification number.

#### Definition of eczema cases and controls

- Self-reported eczema was defined by answering `yes' to the `Eczema ever' question in HUNT3: `Have you had, or do you have any of the following diseases: Eczema on hands?'

- ICD-codes, ICD9 691.8 or ICD10 L20, available from regional hospitals and private specialist practitioners (including dermatologists) from the Control and Payment of Health Reimbursement Register (KUHR) or ICPC-2 code S87 from general practitioners from KUHR.

- There were 7000 genotyped individuals in HUNT with either self-reported eczema and/or at least one of the specified codes, with the remaining 42,205 genotyped individuals defined as controls.

#### Definition of environmental exposures

- Cat ownership: Self-reported in HUNT3: Did you grow up with pets? Yes, cat.

- Dog ownership: Self-reported in HUNT3: Did you grow up with pets? Yes, dog.

- Environmental tobacco smoke exposure: Self-reported in HUNT2 and HUNT3: Did any of the adults where you grew up smoke indoors? Yes.

#### Definition of family history of atopic disease

- Self-reported in HUNT3 and HUNT4: Do your parents, siblings or children have, or have they ever had, some of the following diseases? Allergies/hay fever/nasal allergies. Yes to any.

#### Genotyping and imputation

Participants from HUNT2 and HUNT3 were genotyped using one of three different Illumina HumanCoreExome arrays (HumanCoreExome12 v1.0, HumanCoreExome12 v1.1 and UM HUNT Biobank v1.0) (2). Genotype calling was performed with GenTrain v.2.0 in GenomeStudio v.2011.1 (Illumina). Samples with <99% genotype calls, with large chromosomal copy number variants, contamination >2.5% as estimated with BAF Regress (3), with genotypic and phenotypic sex discordance, and not of European ancestry were excluded, leaving 69,422 genotyped subjects. Genetic variants out of Hardy-Weinberg equilibrium (p-value <0.0001) were excluded.

Imputation was conducted on samples of recent European ancestry using a panel from the Haplotype Reference Consortium (HRC) (release v1.1). Phasing was done using EAGLE v 2.3, followed by imputation using Minimac3 (v2.0.1, http://genome.sph.umich.edu/wiki/Minimac3) from a merged reference panel constructed from HRC panel (release v1.1) and a local reference panel based on 2,202 whole-genome sequenced HUNT participants, resulting in 24.9 million SNPs (R2≥0.3).

The analysis sample was restricted to individuals with available information on eczema, at least one environmental exposure and one genetic variant, and confounders, resulting in a dataset comprising 46,439 individuals.

**Ethics**

Participation in HUNT is based on informed consent, and the study has been approved by the Norwegian Data Protection Authority and the Regional Committee for Medical and Health Research Ethics in Central Norway (REK Reference number 27420).

**Acknowledgements and Funding**

The Trøndelag Health Study (The HUNT Study) is a collaboration between HUNT Research Centre (Faculty of Medicine and Health Sciences, NTNU, Norwegian University of Science and Technology), Trøndelag County Council, Central Norway Regional Health Authority, and the Norwegian Institute of Public Health. The genotyping in HUNT was financed by the National Institutes of Health; University of Michigan; the Research Council of Norway; the Liaison Committee for Education, Research and Innovation in Central Norway; and the Joint Research Committee between St Olavs hospital and the Faculty of Medicine and Health Sciences, NTNU. The genetic investigations of the HUNT Study are a collaboration between researchers from the HUNT Center for Molecular and Clinical Epidemiology (formerly known as the K.G. Jebsen Center for Genetic Epidemiology as of August 1[st] 2023), NTNU, and the University of Michigan Medical School and the University of Michigan School of Public Health. We thank HUNT participants for donating their time, samples, and information to help others; clinicians and other employees at Nord-Trøndelag Hospital Trust for their support and for contributing to data collection. ML was supported by grants from the Liaison Committee for Education, Research and Innovation in Central Norway and the Joint Research Committee between St Olavs hospital and the Faculty of Medicine and Health Sciences, NTNU.

### *UK Biobank (UKBB)*

**Recruitment**

UK Biobank is a population-based health research resource consisting of approximately 500,000 people, aged between 38 years and 73 years, who were recruited between the years 2006 and 2010 from across the UK[1]. Particularly focused on identifying determinants of human diseases in middle-aged and older individuals, participants provided a range of information (such as demographics, health status, lifestyle measures, cognitive testing, personality self-report, and physical and mental health measures) via questionnaires and interviews; anthropometric measures, BP readings and samples of blood, urine and saliva were also taken (data available at www.ukbiobank.ac.uk). A full description of the study design, participants and quality control (QC) methods have been described in detail previously[2]. UK Biobank received ethical approval from the Research Ethics Committee (REC reference for UK Biobank is 11/NW/0382; UK Biobank application number 10074).

**Definition of eczema cases and controls**

AD cases were defined based on their response during a verbal interview with a trained staff member at the assessment centre. Participants were asked to tell the interviewer which serious illnesses or disabilities they had been diagnosed with by a doctor, and were defined as AD if this disease was mentioned. Disease information was also obtained from the Hospital Episode Statistics (HES) data extract service where health-related outcomes had been defined by International Classification of Diseases (ICD)- 10 code L20. Additionally, individuals were excluded from the AD controls if they had answered “yes” to “Has a doctor ever told you that you have hay fever, allergic rhinitis or eczema”.

**Definition of environmental exposures**

<u>Breastfeeding (ever any)</u>

Participants were asked via a touchscreen questionnaire “were you breastfed when you were a baby?” where they responded either “yes” or “no”.

<u>Presence of older siblings</u>

Via a touchscreen questionnaire, participants were asked “How many OLDER brothers/sisters do you have? (Please include those who have died, and twins. Do not include half-, step- or adopted brothers and sisters)”.

<u>Environmental tobacco smoke exposure (up to 2 years)</u>

Via a touchscreen questionnaire, participants were asked “Did your mother smoke regularly around the time when you were born?”. Participants answered either “yes”, “no”, “do not know” or “prefer not to answer”.

**Definition of parental education (as a measure of socioeconomic status)**

Participants' highest qualification was used as a proxy for parental education. Participants were asked via a touchscreen questionnaire “Which of the following qualifications do you have?” where they could select “College or University degree”, “A levels/AS levels or equivalent”, “O levels/GCSEs or equivalent”, “CSEs or equivalent”, “NVQ or HND or HNC or equivalent”, “Other professional qualifications eg: nursing, teaching”, “None of the above” or “Prefer not to answer”.

**Genotyping**

The full data release contains the cohort of successfully genotyped samples (n=488,377). 49,979 individuals were genotyped using the UK BiLEVE array and 438,398 using the UK Biobank axiom array. Pre-imputation QC, phasing and imputation are described elsewhere[3]. In brief, prior to phasing, multiallelic SNPs or those with MAF ≤1% were removed. Phasing of genotype data was performed using a modified version of the SHAPEIT2 algorithm[4]. Genotype imputation to a reference set combining the UK10K haplotype and HRC reference panels[5] was performed using IMPUTE2 algorithms[6]. The analyses presented here were restricted to autosomal variants using a graded filtering with varying imputation quality for different allele frequency ranges. Therefore, rarer genetic variants are required to have a higher imputation INFO score (Info>0.3 for MAF >3%; Info>0.6 for MAF 1-3%; Info>0.8 for MAF 0.5-1%; Info>0.9 for MAF 0.1-0.5%) with MAF and Info scores having been recalculated on an in-house derived 'European' subset.

Individuals with sex-mismatch (derived by comparing genetic sex and reported sex) or individuals with sex-chromosome aneuploidy were excluded from the analysis (n=814). We restricted the sample to individuals of white British ancestry who self-report as "White British" and who have very similar ancestral backgrounds according to the PCA (n=409,703), as described by Bycroft[3]. Estimated kinship coefficients using the KING toolset[7] identified 107,162 pairs of related individuals[3]. An inhouse algorithm was then applied to this list and preferentially removed the individuals related to the greatest number of other individuals until no related pairs remain. These individuals were excluded (n=79,448). Additionally 2 individuals were removed due to them relating to a very large number (>200) of individuals.

**Acknowledgments and Funding**

This research has been conducted using the UK Biobank Resource under Application Number 10074. Quality Control filtering of the UK Biobank data was conducted within the MRC-IEU by R.Mitchell, G.Hemani, T.Dudding, L.Corbin, S.Harrison, L.Paternoster as described in the published protocol (doi: 10.5523/bris.1ovaau5sxunp2cv8rcy88688v). The MRC IEU UK Biobank GWAS pipeline was developed by B.Elsworth, R.Mitchell, C.Raistrick, L.Paternoster, G.Hemani, T.Gaunt (doi: 10.5523/bris.pnoat8cxo0u52p6ynfaekeigi).

**Additional file 7: Definition and coding of environmental exposures**

| Environmental exposure | Variable name | Definition | Coding |
|---|---|---|---|
| Eczema (outcome) | eczema | Ever had eczema (restricted to Dr diagnosed if available) | Dichotomous (yes=1; no=0) |
| Cat ownership | cat | Reported ownership *in utero* or at birth | Dichotomous (yes=1; no=0) |
| Dog ownership | dog | Reported ownership *in utero* or at birth | Dichotomous (yes=1; no=0) |
| House dust mite exposure at birth | hdm_birth | Amount of Der p1 in dust samples from baby's sleeping environment (mcg/g of dust) | Continuous |
| House dust mite exposure at 1 year | hdm_1y | | |
| Washing practices up to 6 months | wash_6m | Bath/shower frequency in early childhood comparing daily or more frequent versus less than daily: up to and including 6 months of age | Dichotomous (more frequent=1; less frequent=0) |
| Washing practices up to 2 years | wash_2y | Bath/shower frequency in early childhood comparing Daily or more frequent versus less than daily: up to and including 2 years of age | Dichotomous (more frequent=1; less frequent=0) |
| Smoking in household in utero | smoke_utero | Parental report:<br>in utero | Dichotomous (yes=1; no=0) |
| Smoking in household up to 2 years | smoke_child | Parental report:<br>in early childhood – up to 2 years | Dichotomous (yes=1; no=0) |

| | | | |
|---|---|---|---|
| Antibiotic use in utero | antibio_utero | Antibiotics prescribed:<br>in utero | Dichotomous (yes=1; no=0) |
| Antibiotic use up to 6 months | antibio_6m | Antibiotics prescribed:<br>within first 6 months of life | Dichotomous (yes=1; no=0) |
| Antibiotic use up to 12 months | antibio_12m | Antibiotics prescribed:<br>within first 12 months of life | Dichotomous (yes=1; no=0) |
| Environmental pollution $PM_{10}$ at birth | pm10 | Exposure at birth | Continuous |
| Environmental pollution $NO_2$ at birth | no2 | Exposure at birth | Continuous |
| Breastfeeding ever | ever_brfed | Parental report of 'ever breastfed' yes/no | Dichotomous (yes=1; no=0) |
| Exclusive breastfeeding ever | excl_brfed | Exclusive breastfeeding for >1 week's duration | Dichotomous (yes=1; no=0) |
| Duration of any breastfeeding | dura_brfed | Duration of any breastfeeding in months | Continuous |
| Mode of delivery | Caes | Caesarean section vs vaginal delivery | Dichotomous (yes=1; no=0) |
| Elder siblings | siblings | Presence of elder sibling(s) | Dichotomous (yes=1; no=0) |
| Parental education (PE) (confounder) | pe | Parental education level as appropriate in cohort | Dichotomous |
| Family history (FH) (confounder) | fh | Any vs. none in first degree relatives (ideally measured at time of birth/recruitment/pre-onset) | Dichotomous |
| Sex (confounder) | sex | | Dichotomous (Female=1, Male=0) |

**Additional file 8: Table of selected SNVs, risk alleles and risk allele frequencies**

| SNV or gene | Risk allele | Chromosome | Risk allele frequency* |
|---|---|---|---|
| *FLG* | Any of 4 loss of function mutations | 1 | |
| rs10214237 | T | 5 | 0.72 |
| rs1057258 | C | 2 | 0.82 |
| rs10995251 | C | 10 | 0.62 |
| rs112111458 | A | 2 | 0.87 |
| rs11657987 | T | 17 | 0.51 |
| rs12153855 | T | 6 | 0.88 |
| rs12295535 | T | 11 | 0.03 |
| rs16948048 | G | 17 | 0.37 |
| rs17389644 | A | 4 | 0.24 |
| rs17881320 | T | 17 | 0.08 |
| rs2041733 | T | 16 | 0.47 |
| rs2143950 | T | 14 | 0.17 |
| rs2164983 | A | 19 | 0.19 |
| rs2227483 | T | 12 | 0.55 |
| rs2228145 | C | 1 | 0.29 |
| rs2897442 | C | 5 | 0.30 |
| rs479844 | G | 11 | 0.56 |
| rs6010620 | G | 20 | 0.77 |
| rs6473227 | C | 8 | 0.38 |
| rs7127307 | T | 11 | 0.53 |
| rs7146581 | C | 14 | 0.74 |
| rs7927894 | T | 11 | 0.35 |
| rs13015714 | G | 2 | 0.21 |

* Allele frequencies derived from
http://snipa.helmholtz-muenchen.de/snipa/index.php?task=snp_annotation,
except rs2228145 which is derived from
https://www.ncbi.nlm.nih.gov/snp/rs2228145#frequency_tab

**Additional file 9: Logistic regression models on ever having atopic eczema**

| Model | Model description | Model formula | Reported estimate |
|---|---|---|---|
| **M1** | Main effect of exposure (E), adjusted for sex (sex) | Eczema ever = $\beta_0$ + $\boldsymbol{\beta_1}$**(E)** + $\beta_2$(sex) | $\beta_1$ |
| **M2** | M1, additionally adjusted for parental education (PE) | Eczema ever = $\beta_0$ + $\boldsymbol{\beta_1}$**(E)** + $\beta_2$(sex) + $\beta_3$(PE) | $\beta_1$ |
| **M3** | M1, additionally adjusted for family history of allergies (FH) | Eczema ever = $\beta_0$ + $\boldsymbol{\beta_1}$**(E)** + $\beta_2$(sex) + $\beta_3$(FH) | $\beta_1$ |
| **M4** | M1, additionally adjusted for parental education (PE) and family history of allergies (FH) | Eczema ever = $\beta_0$ + $\boldsymbol{\beta_1}$**(E)** + $\beta_2$(sex) + $\beta_3$(PE) + $\beta_4$(FH) | $\beta_1$ |
| **I1** | Gene-environment interaction model (G*E) | Eczema ever = $\beta_0$ + $\beta_1$(G) + $\beta_2$(E) + $\boldsymbol{\beta_3}$**(G*E)** + $\beta_4$(sex) | $\beta_3$ |
| **I2** | Genetic effect (G) in unexposed stratum | Eczema ever = $\beta_0$ + $\boldsymbol{\beta_1}$**(G)** + $\beta_2$(sex) | $\beta_1$ |
| **I3** | Genetic effect (G) in exposed stratum | Eczema ever = $\beta_0$ + $\boldsymbol{\beta_1}$**(G)** + $\beta_2$(sex) | $\beta_1$ |
| **S1** | Sensitivity analysis:<br>I1, additionally adjusted for interactions of gene with sex (G*sex) and parental education (G*PE) | Eczema ever = $\beta_0$ + $\beta_1$(G) + $\beta_2$(E) + $\boldsymbol{\beta_3}$**(G*E)** + $\beta_4$(sex) + $\beta_5$(G*sex) + $\beta_6$(PE) + $\beta_7$(G*PE) | $\beta_3$ |
| **S2** | Sensitivity analysis:<br>I1, additionally adjusted for interactions of gene with sex (G*sex) and family history (G*FH) | Eczema ever = $\beta_0$ + $\beta_1$(G) + $\beta_2$(E) + $\boldsymbol{\beta_3}$**(G*E)** + $\beta_4$(sex) + $\beta_5$(G*sex) + $\beta_6$(FH) + $\beta_7$(G*FH) | $\beta_3$ |
| **S3** | Sensitivity analysis:<br>I1, additionally adjusted for interactions of gene with sex (G*sex), parental education (G*PE) and family history (G*FH) | Eczema ever = $\beta_0$ + $\beta_1$(G) + $\beta_2$(E) + $\boldsymbol{\beta_3}$**(G*E)** + $\beta_4$(sex) + $\beta_5$(G*sex) + $\beta_6$(PE) + $\beta_7$(G*PE) + $\beta_8$(FH) + $\beta_9$(G*FH) | $\beta_3$ |

Additional file 10: Full results of the replication analysis

| exposure | SNP | model | N | N_studies | OR_fixed | CI_fixed | p_fixed | OR_random | CI_random | p_random | p_heterogeneity |
|---|---|---|---|---|---|---|---|---|---|---|---|
| dura_brfed | flg | M1 | 3060 | 5 | 1 | [0.98-1.03] | 0.69 | 1 | [0.98-1.03] | 0.82 | 0.35 |
| dura_brfed | flg | M2 | 2850 | 5 | 1.01 | [0.98-1.03] | 0.64 | 1.01 | [0.98-1.04] | 0.73 | 0.34 |
| dura_brfed | flg | M3 | 3060 | 5 | 1 | [0.98-1.02] | 0.95 | 1 | [0.97-1.03] | 0.98 | 0.41 |
| dura_brfed | flg | M4 | 2850 | 5 | 1 | [0.98-1.03] | 0.83 | 1 | [0.97-1.03] | 0.89 | 0.36 |
| dura_brfed | flg | I1 | 3056 | 5 | 1.06 | [0.97-1.16] | 0.22 | 1.06 | [0.97-1.16] | 0.22 | 0.34 |
| dura_brfed | flg | S1 | 2846 | 5 | 1.04 | [0.94-1.15] | 0.47 | 1.03 | [0.90-1.19] | 0.71 | 0.14 |
| dura_brfed | flg | S2 | 3056 | 5 | 1.02 | [0.93-1.12] | 0.66 | 1.02 | [0.93-1.12] | 0.66 | 0.23 |
| dura_brfed | flg | S3 | 2846 | 5 | 1.01 | [0.91-1.12] | 0.87 | 1 | [0.87-1.16] | 0.98 | 0.12 |
| siblings | flg | M1 | 6189 | 6 | 0.93 | [0.80-1.09] | 0.39 | 0.93 | [0.80-1.09] | 0.39 | 0.62 |
| siblings | flg | M2 | 5928 | 6 | 0.96 | [0.81-1.13] | 0.63 | 0.96 | [0.81-1.13] | 0.63 | 0.76 |
| siblings | flg | M3 | 6189 | 6 | 0.89 | [0.76-1.04] | 0.14 | 0.89 | [0.76-1.04] | 0.14 | 0.6 |
| siblings | flg | M4 | 5928 | 6 | 0.91 | [0.77-1.08] | 0.28 | 0.91 | [0.77-1.08] | 0.28 | 0.78 |
| siblings | flg | I1 | 6185 | 6 | 1.12 | [0.65-1.94] | 0.68 | 1.12 | [0.65-1.94] | 0.68 | 0.21 |
| siblings | flg | I2 | 2897 | 6 | 2.26 | [1.64-3.13] | 7.20E-07 | 2.35 | [1.61-3.30] | 9.30E-06 | 0.3 |
| siblings | flg | I3 | 3288 | 6 | 2.49 | [1.76-3.53] | 2.90E-07 | 2.56 | [1.70-3.75] | 6.70E-06 | 0.14 |
| siblings | flg | S1 | 5924 | 6 | 1.24 | [0.70-2.21] | 0.46 | 1.24 | [0.70-2.21] | 0.46 | 0.34 |
| siblings | flg | S2 | 6185 | 6 | 1.14 | [0.64-2.02] | 0.67 | 1.14 | [0.64-2.02] | 0.67 | 0.27 |
| siblings | flg | S3 | 5924 | 6 | 1.27 | [0.69-2.33] | 0.44 | 1.27 | [0.69-2.33] | 0.44 | 0.31 |
| dura_brfed | rs13015714 | M1 | 4252 | 5 | 1 | [0.98-1.03] | 0.71 | 1 | [0.97-1.04] | 0.87 | 0.12 |
| dura_brfed | rs13015714 | M2 | 4252 | 5 | 1 | [0.98-1.03] | 0.73 | 1 | [0.97-1.03] | 0.98 | 0.2 |
| dura_brfed | rs13015714 | M3 | 4252 | 5 | 1 | [0.98-1.02] | 0.85 | 1 | [0.97-1.03] | 0.84 | 0.19 |
| dura_brfed | rs13015714 | M4 | 4252 | 5 | 1 | [0.98-1.02] | 0.82 | 1 | [0.97-1.03] | 0.99 | 0.26 |
| dura_brfed | rs13015714 | I1 | 4252 | 5 | 0.99 | [0.95-1.02] | 0.47 | 0.99 | [0.95-1.02] | 0.47 | 0.96 |
| dura_brfed | rs13015714 | S1 | 4252 | 5 | 0.99 | [0.95-1.02] | 0.47 | 0.99 | [0.95-1.02] | 0.47 | 0.85 |
| dura_brfed | rs13015714 | S2 | 4252 | 5 | 0.99 | [0.95-1.02] | 0.45 | 0.99 | [0.95-1.02] | 0.45 | 0.98 |
| dura_brfed | rs13015714 | S3 | 4252 | 5 | 0.99 | [0.95-1.02] | 0.44 | 0.99 | [0.95-1.02] | 0.44 | 0.89 |
| siblings | rs10995251 | M1 | 7529 | 6 | 0.96 | [0.85-1.09] | 0.54 | 0.96 | [0.85-1.09] | 0.54 | 0.89 |
| siblings | rs10995251 | M2 | 7529 | 6 | 0.96 | [0.84-1.09] | 0.52 | 0.96 | [0.84-1.09] | 0.52 | 0.9 |
| siblings | rs10995251 | M3 | 7529 | 6 | 0.96 | [0.85-1.09] | 0.55 | 0.96 | [0.85-1.09] | 0.55 | 0.8 |
| siblings | rs10995251 | M4 | 7529 | 6 | 0.96 | [0.84-1.09] | 0.52 | 0.96 | [0.84-1.09] | 0.52 | 0.81 |
| siblings | rs10995251 | I1 | 7529 | 6 | 1.03 | [0.85-1.25] | 0.74 | 1.03 | [0.85-1.25] | 0.74 | 0.53 |
| siblings | rs10995251 | I2 | 3337 | 6 | 0.97 | [0.85-1.11] | 0.67 | 0.97 | [0.85-1.11] | 0.67 | 0.65 |
| siblings | rs10995251 | I3 | 4192 | 6 | 1 | [0.88-1.14] | 1 | 1 | [0.88-1.14] | 1 | 0.61 |
| siblings | rs10995251 | S1 | 7529 | 6 | 1.04 | [0.86-1.25] | 0.71 | 1.04 | [0.86-1.25] | 0.71 | 0.61 |
| siblings | rs10995251 | S2 | 7529 | 6 | 1.06 | [0.87-1.28] | 0.57 | 1.06 | [0.87-1.28] | 0.57 | 0.68 |
| siblings | rs10995251 | S3 | 7529 | 6 | 1.06 | [0.87-1.28] | 0.58 | 1.06 | [0.87-1.28] | 0.58 | 0.7 |
| siblings | rs7146581 | M1 | 7529 | 6 | 0.96 | [0.85-1.09] | 0.54 | 0.96 | [0.85-1.09] | 0.54 | 0.89 |
| siblings | rs7146581 | M2 | 7529 | 6 | 0.96 | [0.84-1.09] | 0.52 | 0.96 | [0.84-1.09] | 0.52 | 0.9 |
| siblings | rs7146581 | M3 | 7529 | 6 | 0.96 | [0.85-1.09] | 0.55 | 0.96 | [0.85-1.09] | 0.55 | 0.8 |
| siblings | rs7146581 | M4 | 7529 | 6 | 0.96 | [0.84-1.09] | 0.52 | 0.96 | [0.84-1.09] | 0.52 | 0.81 |
| siblings | rs7146581 | I1 | 7529 | 6 | 1.04 | [0.84-1.29] | 0.73 | 1.04 | [0.84-1.29] | 0.73 | 0.58 |
| siblings | rs7146581 | I2 | 3337 | 6 | 1 | [0.86-1.17] | 0.96 | 1 | [0.86-1.17] | 0.96 | 0.54 |
| siblings | rs7146581 | I3 | 4192 | 6 | 1.07 | [0.92-1.24] | 0.38 | 1.07 | [0.92-1.24] | 0.38 | 0.59 |
| siblings | rs7146581 | S1 | 7529 | 6 | 1.04 | [0.83-1.29] | 0.75 | 1.04 | [0.83-1.29] | 0.75 | 0.54 |
| siblings | rs7146581 | S2 | 7529 | 6 | 1.01 | [0.81-1.26] | 0.94 | 1.01 | [0.81-1.26] | 0.94 | 0.49 |
| siblings | rs7146581 | S3 | 7529 | 6 | 1.01 | [0.81-1.26] | 0.93 | 1.01 | [0.81-1.26] | 0.93 | 0.47 |
| siblings | rs7927894 | M1 | 7529 | 6 | 0.96 | [0.85-1.09] | 0.54 | 0.96 | [0.85-1.09] | 0.54 | 0.89 |
| siblings | rs7927894 | M2 | 7529 | 6 | 0.96 | [0.84-1.09] | 0.52 | 0.96 | [0.84-1.09] | 0.52 | 0.9 |
| siblings | rs7927894 | M3 | 7529 | 6 | 0.96 | [0.85-1.09] | 0.55 | 0.96 | [0.85-1.09] | 0.55 | 0.8 |
| siblings | rs7927894 | M4 | 7529 | 6 | 0.96 | [0.84-1.09] | 0.52 | 0.96 | [0.84-1.09] | 0.52 | 0.81 |
| siblings | rs7927894 | I1 | 7529 | 6 | 1 | [0.83-1.21] | 0.97 | 0.97 | [0.74-1.32] | 0.84 | 0.1 |
| siblings | rs7927894 | I2 | 3337 | 6 | 1.08 | [0.95-1.24] | 0.24 | 1.09 | [0.93-1.27] | 0.28 | 0.065 |
| siblings | rs7927894 | I3 | 4192 | 6 | 1.04 | [0.92-1.18] | 0.49 | 1.03 | [0.86-1.25] | 0.75 | 0.097 |
| siblings | rs7927894 | S1 | 7529 | 6 | 1.01 | [0.84-1.22] | 0.92 | 0.98 | [0.75-1.32] | 0.89 | 0.12 |
| siblings | rs7927894 | S2 | 7529 | 6 | 1 | [0.83-1.21] | 0.98 | 0.97 | [0.73-1.32] | 0.8 | 0.096 |
| siblings | rs7927894 | S3 | 7529 | 6 | 1.01 | [0.83-1.22] | 0.91 | 0.98 | [0.74-1.33] | 0.87 | 0.11 |
| smoke_utero | rs16948048 | M1 | 8078 | 5 | 1.02 | [0.89-1.17] | 0.78 | 1.03 | [0.83-1.27] | 0.79 | 0.057 |
| smoke_utero | rs16948048 | M2 | 8078 | 5 | 1.04 | [0.90-1.19] | 0.63 | 1.04 | [0.86-1.25] | 0.68 | 0.12 |
| smoke_utero | rs16948048 | M3 | 8078 | 5 | 1.04 | [0.90-1.20] | 0.59 | 1.05 | [0.84-1.31] | 0.66 | 0.043 |
| smoke_utero | rs16948048 | M4 | 8078 | 5 | 1.05 | [0.91-1.21] | 0.48 | 1.06 | [0.86-1.29] | 0.57 | 0.084 |
| smoke_utero | rs16948048 | I1 | 8078 | 5 | 1.04 | [0.85-1.27] | 0.72 | 1.04 | [0.85-1.27] | 0.72 | 0.75 |
| smoke_utero | rs16948048 | I2 | 5074 | 5 | 0.97 | [0.86-1.09] | 0.6 | 0.98 | [0.82-1.16] | 0.8 | 0.069 |
| smoke_utero | rs16948048 | I3 | 3004 | 5 | 0.97 | [0.82-1.15] | 0.73 | 0.99 | [0.81-1.20] | 0.95 | 0.2 |
| smoke_utero | rs16948048 | S1 | 8078 | 5 | 1.06 | [0.86-1.31] | 0.57 | 1.06 | [0.86-1.31] | 0.57 | 0.5 |
| smoke_utero | rs16948048 | S2 | 8078 | 5 | 1.03 | [0.84-1.27] | 0.78 | 1.03 | [0.84-1.27] | 0.78 | 0.72 |
| smoke_utero | rs16948048 | S3 | 8078 | 5 | 1.06 | [0.86-1.31] | 0.61 | 1.06 | [0.86-1.31] | 0.61 | 0.48 |
| cat | rs16948048 | M1 | 49212 | 3 | 1.01 | [0.96-1.07] | 0.69 | 0.9 | [0.63-1.45] | 0.58 | 0.061 |
| cat | rs16948048 | M2 | 2773 | 3 | 0.93 | [0.70-1.22] | 0.58 | 0.77 | [0.35-2.02] | 0.5 | 0.018 |
| cat | rs16948048 | M3 | 49212 | 3 | 1.02 | [0.97-1.08] | 0.43 | 0.9 | [0.59-1.56] | 0.64 | 0.038 |
| cat | rs16948048 | M4 | 2773 | 3 | 0.97 | [0.73-1.27] | 0.81 | 0.78 | [0.33-2.30] | 0.57 | 0.0094 |
| cat | rs16948048 | I1 | 49212 | 3 | 1 | [0.92-1.08] | 0.98 | 1 | [0.92-1.08] | 0.98 | 0.64 |
| cat | rs16948048 | I2 | 21207 | 3 | 1.05 | [0.99-1.11] | 0.096 | 1.05 | [0.99-1.11] | 0.096 | 0.7 |
| cat | rs16948048 | I3 | 28005 | 3 | 1.06 | [1.01-1.11] | 0.029 | 1.06 | [1.01-1.11] | 0.029 | 0.49 |
| cat | rs16948048 | S1 | 2773 | 3 | 1.04 | [0.70-1.54] | 0.84 | 1.04 | [0.70-1.54] | 0.84 | 0.34 |
| cat | rs16948048 | S2 | 49212 | 3 | 1 | [0.93-1.08] | 1 | 1 | [0.93-1.08] | 1 | 0.67 |
| cat | rs16948048 | S3 | 2773 | 3 | 1.03 | [0.69-1.54] | 0.88 | 1.03 | [0.69-1.54] | 0.88 | 0.36 |
| cat | rs2227483 | M1 | 49212 | 3 | 1.01 | [0.96-1.07] | 0.69 | 0.9 | [0.63-1.45] | 0.58 | 0.061 |
| cat | rs2227483 | M2 | 2773 | 3 | 0.93 | [0.70-1.22] | 0.58 | 0.77 | [0.35-2.02] | 0.5 | 0.018 |
| cat | rs2227483 | M3 | 49212 | 3 | 1.02 | [0.97-1.08] | 0.43 | 0.9 | [0.59-1.56] | 0.64 | 0.038 |
| cat | rs2227483 | M4 | 2773 | 3 | 0.97 | [0.73-1.27] | 0.81 | 0.78 | [0.33-2.30] | 0.57 | 0.0094 |
| cat | rs2227483 | I1 | 49212 | 3 | 1.05 | [0.97-1.13] | 0.25 | 1.05 | [0.97-1.13] | 0.25 | 0.22 |
| cat | rs2227483 | I2 | 21207 | 3 | 1.04 | [0.99-1.10] | 0.14 | 1.04 | [0.99-1.10] | 0.14 | 0.65 |
| cat | rs2227483 | I3 | 28005 | 3 | 1.08 | [1.03-1.14] | 0.0016 | 1.08 | [1.03-1.14] | 0.0016 | 0.16 |
| cat | rs2227483 | S1 | 2773 | 3 | 0.99 | [0.67-1.46] | 0.97 | 1.15 | [0.52-2.19] | 0.72 | 0.084 |
| cat | rs2227483 | S2 | 49212 | 3 | 1.05 | [0.97-1.13] | 0.25 | 1.05 | [0.97-1.13] | 0.25 | 0.17 |
| cat | rs2227483 | S3 | 2773 | 3 | 0.96 | [0.65-1.42] | 0.84 | 1.13 | [0.49-2.19] | 0.78 | 0.074 |

| | | | | | | | | | | | |
|---|---|---|---|---|---|---|---|---|---|---|---|
| cat | rs2228145 | M1 | 49212 | 3 | 1.01 | [0.96-1.07] | 0.69 | 0.9 | [0.63-1.45] | 0.58 | 0.061 |
| cat | rs2228145 | M2 | 2773 | 3 | 0.93 | [0.70-1.22] | 0.58 | 0.77 | [0.35-2.02] | 0.5 | 0.018 |
| cat | rs2228145 | M3 | 49212 | 3 | 1.02 | [0.97-1.08] | 0.43 | 0.9 | [0.59-1.56] | 0.64 | 0.038 |
| cat | rs2228145 | M4 | 2773 | 3 | 0.97 | [0.73-1.27] | 0.81 | 0.78 | [0.33-2.30] | 0.57 | 0.0094 |
| cat | rs2228145 | I1 | 49212 | 3 | 1 | [0.92-1.08] | 0.93 | 1 | [0.92-1.08] | 0.93 | 0.4 |
| cat | rs2228145 | I2 | 21207 | 3 | 1.06 | [1.00-1.12] | 0.04 | 1.06 | [1.00-1.12] | 0.04 | 0.95 |
| cat | rs2228145 | I3 | 28005 | 3 | 1.05 | [1.00-1.11] | 0.037 | 1.05 | [1.00-1.11] | 0.037 | 0.45 |
| cat | rs2228145 | S1 | 2773 | 3 | 1.1 | [0.74-1.65] | 0.62 | 1.02 | [0.56-2.01] | 0.95 | 0.21 |
| cat | rs2228145 | S2 | 49212 | 3 | 1 | [0.93-1.08] | 0.99 | 1 | [0.93-1.08] | 0.99 | 0.43 |
| cat | rs2228145 | S3 | 2773 | 3 | 1.14 | [0.76-1.70] | 0.53 | 1.08 | [0.64-1.94] | 0.77 | 0.25 |
| cat | rs2897442 | M1 | 49212 | 3 | 1.01 | [0.96-1.07] | 0.69 | 0.9 | [0.63-1.45] | 0.58 | 0.061 |
| cat | rs2897442 | M2 | 2773 | 3 | 0.93 | [0.70-1.22] | 0.58 | 0.77 | [0.35-2.02] | 0.5 | 0.018 |
| cat | rs2897442 | M3 | 49212 | 3 | 1.02 | [0.97-1.08] | 0.43 | 0.9 | [0.59-1.56] | 0.64 | 0.038 |
| cat | rs2897442 | M4 | 2773 | 3 | 0.97 | [0.73-1.27] | 0.81 | 0.78 | [0.33-2.30] | 0.57 | 0.0094 |
| cat | rs2897442 | I1 | 49212 | 3 | 1.02 | [0.94-1.11] | 0.59 | 1.02 | [0.94-1.11] | 0.59 | 0.97 |
| cat | rs2897442 | I2 | 21207 | 3 | 1.04 | [0.98-1.10] | 0.18 | 1.04 | [0.98-1.10] | 0.18 | 0.73 |
| cat | rs2897442 | I3 | 28005 | 3 | 1.07 | [1.01-1.12] | 0.016 | 1.07 | [1.01-1.12] | 0.016 | 0.88 |
| cat | rs2897442 | S1 | 2773 | 3 | 1.05 | [0.69-1.59] | 0.83 | 1.05 | [0.69-1.59] | 0.83 | 0.75 |
| cat | rs2897442 | S2 | 49212 | 3 | 1.02 | [0.94-1.11] | 0.62 | 1.02 | [0.94-1.11] | 0.62 | 0.94 |
| cat | rs2897442 | S3 | 2773 | 3 | 1.02 | [0.67-1.56] | 0.91 | 1.02 | [0.67-1.56] | 0.91 | 0.68 |
| cat | flg | M1 | 48246 | 3 | 1.01 | [0.95-1.06] | 0.83 | 0.84 | [0.59-1.44] | 0.32 | 0.055 |
| cat | flg | M2 | 1807 | 3 | 0.71 | [0.51-1.01] | 0.056 | 0.68 | [0.39-1.23] | 0.16 | 0.13 |
| cat | flg | M3 | 48246 | 3 | 1.02 | [0.96-1.07] | 0.59 | 0.83 | [0.57-1.48] | 0.34 | 0.045 |
| cat | flg | M4 | 1807 | 3 | 0.71 | [0.50-1.01] | 0.06 | 0.67 | [0.37-1.28] | 0.19 | 0.11 |
| cat | flg | I1 | 48246 | 3 | 0.86 | [0.68-1.08] | 0.19 | 0.68 | [0.24-2.42] | 0.47 | 0.47 |
| cat | flg | I2 | 20342 | 3 | 1.78 | [1.52-2.09] | 1.80E-12 | 2.4 | [1.26-3.39] | 0.0077 | 0.025 |
| cat | flg | I3 | 27904 | 3 | 1.44 | [1.24-1.67] | 2.40E-06 | 1.42 | [1.10-1.85] | 0.0067 | 0.6 |
| cat | flg | S1 | 1807 | 3 | 0.22 | [0.02-2.05] | 0.18 | 0.22 | [0.02-2.05] | 0.18 | 0.98 |
| cat | flg | S2 | 48246 | 3 | 0.84 | [0.66-1.06] | 0.15 | 0.49 | [0.10-4.32] | 0.4 | 0.29 |
| cat | flg | S3 | 1807 | 3 | 0.14 | [0.01-1.32] | 0.085 | 0.14 | [0.01-1.32] | 0.085 | 0.98 |
| ever_brfed | flg | M1 | 200351 | 4 | 0.65 | [0.54-0.78] | 2.90E-06 | 0.7 | [0.37-1.22] | 0.26 | 1.60E-07 |
| ever_brfed | flg | M2 | 200351 | 4 | 0.66 | [0.55-0.79] | 4.60E-06 | 0.71 | [0.37-1.25] | 0.31 | 9.90E-08 |
| ever_brfed | flg | M3 | 2430 | 4 | 0.9 | [0.73-1.12] | 0.35 | 0.9 | [0.73-1.12] | 0.35 | 0.7 |
| ever_brfed | flg | M4 | 2430 | 4 | 0.92 | [0.74-1.14] | 0.45 | 0.92 | [0.74-1.14] | 0.45 | 0.61 |
| ever_brfed | flg | I1 | 195879 | 4 | 1.18 | [0.65-2.12] | 0.59 | 1.18 | [0.65-2.12] | 0.59 | 0.71 |
| ever_brfed | flg | I2 | 1183 | 4 | 1.81 | [1.17-2.80] | 0.0076 | 1.81 | [1.17-2.80] | 0.0076 | 0.85 |
| ever_brfed | flg | I3 | 194696 | 4 | 1.89 | [1.74-2.05] | 8.60E-52 | 1.89 | [1.74-2.05] | 8.60E-52 | 0.61 |
| ever_brfed | flg | S1 | 195879 | 4 | 1.16 | [0.64-2.13] | 0.62 | 1.16 | [0.64-2.13] | 0.62 | 0.66 |
| ever_brfed | flg | S2 | 2426 | 4 | 1.01 | [0.49-2.07] | 0.98 | 1.01 | [0.49-2.07] | 0.98 | 0.31 |
| ever_brfed | flg | S3 | 2426 | 4 | 1.01 | [0.49-2.08] | 0.99 | 1 | [0.47-2.12] | 0.99 | 0.27 |
| smoke_child | flg | M1 | 150213 | 7 | 0.91 | [0.87-0.96] | 0.00067 | 0.74 | [0.51-1.33] | 0.11 | 0 |
| smoke_child | flg | M2 | 103894 | 7 | 0.51 | [0.45-0.57] | 2.80E-32 | 0.69 | [0.45-0.77] | 0.072 | 0 |
| smoke_child | flg | M3 | 51887 | 7 | 1.03 | [0.97-1.08] | 0.35 | 0.9 | [0.73-1.26] | 0.33 | 0.011 |
| smoke_child | flg | M4 | 5568 | 7 | 0.86 | [0.73-1.00] | 0.057 | 0.85 | [0.68-1.08] | 0.17 | 0.12 |
| smoke_child | flg | I1 | 147880 | 7 | 1.01 | [0.81-1.25] | 0.93 | 1.01 | [0.81-1.25] | 0.93 | 0.86 |
| smoke_child | flg | I2 | 19038 | 7 | 1.75 | [1.48-2.07] | 4.00E-11 | 2 | [1.53-2.29] | 3.60E-07 | 0.15 |
| smoke_child | flg | I3 | 128842 | 7 | 1.82 | [1.67-1.99] | 1.40E-41 | 1.82 | [1.51-2.20] | 6.00E-10 | 0.054 |
| smoke_child | flg | S1 | 101561 | 7 | 0.89 | [0.59-1.33] | 0.56 | 0.89 | [0.59-1.33] | 0.56 | 0.85 |
| smoke_child | flg | S2 | 51883 | 7 | 1.01 | [0.80-1.28] | 0.91 | 1.01 | [0.80-1.28] | 0.91 | 0.7 |
| smoke_child | flg | S3 | 5564 | 7 | 0.79 | [0.46-1.35] | 0.39 | 0.79 | [0.46-1.35] | 0.39 | 0.77 |
| wash_2y | flg | M1 | 1061 | 1 | 0.86 | [0.52-1.43] | 0.57 | 0.86 | [0.52-1.43] | 0.57 | 1 |
| wash_2y | flg | M2 | 1061 | 1 | 0.86 | [0.52-1.43] | 0.57 | 0.86 | [0.52-1.43] | 0.57 | 1 |
| wash_2y | flg | M3 | 1061 | 1 | 0.91 | [0.54-1.53] | 0.71 | 0.91 | [0.54-1.53] | 0.71 | 1 |
| wash_2y | flg | M4 | 1061 | 1 | 0.9 | [0.54-1.52] | 0.7 | 0.9 | [0.54-1.52] | 0.7 | 1 |
| wash_2y | flg | I1 | 1061 | 1 | 1.93 | [0.29-12.99] | 0.5 | 1.93 | [0.29-12.99] | 0.5 | 1 |
| wash_2y | flg | I2 | 986 | 1 | 1.9 | [1.27-2.83] | 0.0017 | 1.9 | [1.27-2.83] | 0.0017 | 1 |
| wash_2y | flg | I3 | 75 | 1 | 3.53 | [0.54-22.99] | 0.19 | 3.53 | [0.54-22.99] | 0.19 | 1 |
| wash_2y | flg | S1 | 1061 | 1 | 1.64 | [0.24-11.33] | 0.62 | 1.64 | [0.24-11.33] | 0.62 | 1 |
| wash_2y | flg | S2 | 1061 | 1 | 1.05 | [0.15-7.39] | 0.96 | 1.05 | [0.15-7.39] | 0.96 | 1 |
| wash_2y | flg | S3 | 1061 | 1 | 1.05 | [0.15-7.41] | 0.96 | 1.05 | [0.15-7.41] | 0.96 | 1 |
| antibio_utero | rs10995251 | M1 | 2666 | 1 | 1.05 | [0.76-1.46] | 0.75 | 1.05 | [0.76-1.46] | 0.75 | 1 |
| antibio_utero | rs10995251 | M2 | 2666 | 1 | 1.06 | [0.76-1.47] | 0.74 | 1.06 | [0.76-1.47] | 0.74 | 1 |
| antibio_utero | rs10995251 | M3 | 2666 | 1 | 1.05 | [0.76-1.45] | 0.77 | 1.05 | [0.76-1.45] | 0.77 | 1 |
| antibio_utero | rs10995251 | M4 | 2666 | 1 | 1.05 | [0.76-1.46] | 0.76 | 1.05 | [0.76-1.46] | 0.76 | 1 |
| antibio_utero | rs10995251 | I1 | 2666 | 1 | 0.88 | [0.55-1.41] | 0.59 | 0.88 | [0.55-1.41] | 0.59 | 1 |
| antibio_utero | rs10995251 | I2 | 1895 | 1 | 0.95 | [0.74-1.23] | 0.7 | 0.95 | [0.74-1.23] | 0.7 | 1 |
| antibio_utero | rs10995251 | I3 | 771 | 1 | 0.84 | [0.56-1.24] | 0.38 | 0.84 | [0.56-1.24] | 0.38 | 1 |
| antibio_utero | rs10995251 | S1 | 2666 | 1 | 0.89 | [0.55-1.43] | 0.62 | 0.89 | [0.55-1.43] | 0.62 | 1 |
| antibio_utero | rs10995251 | S2 | 2666 | 1 | 0.86 | [0.54-1.39] | 0.54 | 0.86 | [0.54-1.39] | 0.54 | 1 |
| antibio_utero | rs10995251 | S3 | 2666 | 1 | 0.87 | [0.54-1.40] | 0.56 | 0.87 | [0.54-1.40] | 0.56 | 1 |
| ever_brfed | rs13015714 | M1 | 199300 | 3 | 0.5 | [0.39-0.64] | 5.20E-08 | 0.65 | [0.27-1.19] | 0.33 | 2.90E-06 |
| ever_brfed | rs13015714 | M2 | 199300 | 3 | 0.5 | [0.39-0.65] | 9.50E-08 | 0.67 | [0.27-1.24] | 0.39 | 1.60E-06 |
| ever_brfed | rs13015714 | M3 | 1379 | 3 | 1.01 | [0.69-1.46] | 0.96 | 1.01 | [0.69-1.46] | 0.96 | 0.66 |
| ever_brfed | rs13015714 | M4 | 1379 | 3 | 1.07 | [0.73-1.57] | 0.73 | 1.07 | [0.73-1.57] | 0.73 | 0.73 |
| ever_brfed | rs13015714 | I1 | 199300 | 3 | 0.74 | [0.46-1.18] | 0.2 | 0.74 | [0.46-1.18] | 0.2 | 0.85 |
| ever_brfed | rs13015714 | I2 | 644 | 3 | 1.42 | [0.92-2.20] | 0.11 | 1.42 | [0.92-2.20] | 0.11 | 0.99 |
| ever_brfed | rs13015714 | I3 | 198656 | 3 | 1.12 | [1.08-1.17] | 6.10E-10 | 1.12 | [1.08-1.17] | 6.10E-10 | 0.55 |
| ever_brfed | rs13015714 | S1 | 199300 | 3 | 0.76 | [0.47-1.22] | 0.25 | 0.76 | [0.47-1.22] | 0.25 | 0.93 |
| ever_brfed | rs13015714 | S2 | 1379 | 3 | 0.63 | [0.28-1.41] | 0.26 | 0.63 | [0.28-1.41] | 0.26 | 0.63 |
| ever_brfed | rs13015714 | S3 | 1379 | 3 | 0.71 | [0.30-1.66] | 0.43 | 0.71 | [0.30-1.66] | 0.43 | 0.88 |
| siblings | rs10214237 | M1 | 5049 | 4 | 0.93 | [0.78-1.12] | 0.47 | 0.93 | [0.78-1.12] | 0.47 | 0.7 |
| siblings | rs10214237 | M2 | 5049 | 4 | 0.93 | [0.77-1.12] | 0.45 | 0.93 | [0.77-1.12] | 0.45 | 0.71 |
| siblings | rs10214237 | M3 | 5049 | 4 | 0.92 | [0.76-1.11] | 0.37 | 0.92 | [0.76-1.11] | 0.37 | 0.6 |
| siblings | rs10214237 | M4 | 5049 | 4 | 0.91 | [0.76-1.10] | 0.34 | 0.91 | [0.76-1.10] | 0.34 | 0.62 |
| siblings | rs10214237 | I1 | 5049 | 4 | 1.09 | [0.82-1.46] | 0.55 | 1.09 | [0.82-1.46] | 0.55 | 0.6 |
| siblings | rs10214237 | I2 | 2001 | 4 | 0.92 | [0.75-1.15] | 0.47 | 0.92 | [0.75-1.15] | 0.47 | 0.96 |
| siblings | rs10214237 | I3 | 3048 | 4 | 1.03 | [0.86-1.24] | 0.75 | 1.05 | [0.81-1.33] | 0.72 | 0.13 |

| | | | | | | | | | | | |
|---|---|---|---|---|---|---|---|---|---|---|---|
| siblings | rs10214237 | S1 | 5049 | 4 | 1.1 | [0.82-1.47] | 0.53 | 1.1 | [0.82-1.47] | 0.53 | 0.63 |
| siblings | rs10214237 | S2 | 5049 | 4 | 1.11 | [0.83-1.50] | 0.47 | 1.11 | [0.83-1.50] | 0.47 | 0.67 |
| siblings | rs10214237 | S3 | 5049 | 4 | 1.12 | [0.83-1.51] | 0.46 | 1.12 | [0.83-1.51] | 0.46 | 0.68 |
| dog | rs10214237 | M1 | 47185 | 2 | 0.98 | [0.93-1.04] | 0.56 | 0.98 | [0.93-1.04] | 0.56 | 0.68 |
| dog | rs10214237 | M2 | 746 | 2 | 0.9 | [0.51-1.59] | 0.71 | 0.9 | [0.51-1.59] | 0.71 | 1 |
| dog | rs10214237 | M3 | 47185 | 2 | 0.98 | [0.92-1.03] | 0.38 | 0.98 | [0.92-1.03] | 0.38 | 0.82 |
| dog | rs10214237 | M4 | 746 | 2 | 0.93 | [0.52-1.66] | 0.81 | 0.93 | [0.52-1.66] | 0.81 | 1 |
| dog | rs10214237 | I1 | 47185 | 2 | 0.91 | [0.83-0.99] | 0.025 | 0.91 | [0.83-0.99] | 0.025 | 0.62 |
| dog | rs10214237 | I2 | 28848 | 2 | 1.09 | [1.04-1.15] | 0.0011 | 1.09 | [1.04-1.15] | 0.0011 | 0.55 |
| dog | rs10214237 | I3 | 18337 | 2 | 0.99 | [0.93-1.06] | 0.75 | 0.99 | [0.93-1.06] | 0.75 | 0.45 |
| dog | rs10214237 | S1 | 746 | 2 | 1.16 | [0.40-3.33] | 0.78 | 1.16 | [0.40-3.33] | 0.78 | 1 |
| dog | rs10214237 | S2 | 47185 | 2 | 0.9 | [0.83-0.98] | 0.02 | 0.9 | [0.83-0.98] | 0.02 | 0.66 |
| dog | rs10214237 | S3 | 746 | 2 | 1.13 | [0.39-3.28] | 0.83 | 1.13 | [0.39-3.28] | 0.83 | 1 |
